# Precision Prognostics for Cardiovascular Disease in Type 2 Diabetes: A Systematic Review and Meta-analysis

**DOI:** 10.1101/2023.04.26.23289177

**Authors:** Abrar Ahmad, Lee-Ling Lim, Mario Luca Morieri, Claudia Ha-ting Tam, Feifei Cheng, Tinashe Chikowore, Monika Dudenhöffer-Pfeifer, Hugo Fitipaldi, Chuiguo Huang, Sarah Kanbour, Sudipa Sarkar, Robert Wilhelm Koivula, Ayesha A. Motala, Sok Cin Tye, Gechang Yu, Yingchai Zhang, Michele Provenzano, Diana Sherifali, Russell J. de Souza, Deirdre Kay Tobias, ADA/EASD PMDI, Maria F. Gomez, Ronald C.W. Ma, Nestoras Mathioudakis

**Author notes:** Corresponding authors: Maria F. Gomez, PhD Professor Department of Clinical Sciences, Lund University Diabetes Centre, Lund University, Malmö, Sweden Ronald C. W. Ma, FRCP Professor Department of Medicine and Therapeutics, The Chinese University of Hong Kong, Prince of Wales Hospital, Hong Kong Nestoras Mathioudakis, MD MHS Associate Professor of Medicine Division of Endocrinology, Diabetes & Metabolism, Department of Medicine, Johns Hopkins University School of Medicine, Baltimore, Maryland USA. co-first authors (contributed equally to this work) (Abrar Ahmad, Lee-Ling Lim, Mario Luca Morieri, Claudia Ha-ting Tam). senior authors (supervised all aspects of this work) (Maria F. Gomez, Ronald C.W. Ma, Nestoras Mathioudakis).

## Abstract

**Background:** Precision medicine has the potential to improve cardiovascular disease (CVD) risk prediction in individuals with type 2 diabetes (T2D).

**Methods:** We conducted a systematic review and meta-analysis of longitudinal studies to identify potentially novel prognostic factors that may improve CVD risk prediction in T2D. Out of 9380 studies identified, 416 studies met inclusion criteria. Outcomes were reported for 321 biomarker studies, 48 genetic marker studies, and 47 risk score/model studies.

**Results:** Out of all evaluated biomarkers, only 13 showed improvement in prediction performance. Results of pooled meta-analyses, non-pooled analyses, and assessments of improvement in prediction performance and risk of bias, yielded the *highest predictive utility* for N-terminal pro b-type natriuretic peptide (NT-proBNP) (high-evidence), troponin-T (TnT) (moderate-evidence), triglyceride-glucose (TyG) index (moderate-evidence), Genetic Risk Score for Coronary Heart Disease (GRS-CHD) (moderate-evidence); *moderate predictive utility* for coronary computed tomography angiography (low-evidence), single-photon emission computed tomography (low-evidence), pulse wave velocity (moderate-evidence); and *low predictive utility* for C-reactive protein (moderate-evidence), coronary artery calcium score (low-evidence), galectin-3 (low-evidence), troponin-I (low-evidence), carotid plaque (low-evidence), and growth differentiation factor-15 (low-evidence). Risk scores showed modest discrimination, with lower performance in populations different from the original development cohort.

**Conclusions:** Despite high interest in this topic, very few studies conducted rigorous analyses to demonstrate incremental predictive utility beyond established CVD risk factors for T2D. The most promising markers identified were NT-proBNP, TnT, TyG and GRS-CHD, with the highest strength of evidence for NT-proBNP. Further research is needed to determine their clinical utility in risk stratification and management of CVD in T2D.

**Plain Language Summary:** Patients with T2D are at high risk for CVD but predicting who will experience a cardiac event is challenging. Current risk tools and prognostic factors, such as laboratory tests, may not accurately predict risk in all patient populations. There is a need for personalized risk prediction tools to classify patients more accurately so that CVD prevention can be targeted to those who need it most. This study summarizes the best available evidence for novel biomarkers, genetic markers, and risk scores that predict CVD in individuals with T2D. We found that four laboratory markers and a genetic risk score for CHD had high predictive utility beyond traditional CVD risk factors. Risk scores had modest predictive utility when tested in diverse populations. More studies are needed to determine their usefulness in clinical practice. The highest strength of evidence was observed for NT-proBNP, a biomarker currently measured to monitor patients with heart failure in clinical practice, but not for CVD prediction in T2D.

## Introduction

Individuals with type 2 diabetes (T2D) have a 1.5 to 2-fold higher risk of developing cardiovascular disease (CVD) compared to those without T2D^1,2^. This is particularly concerning given the high global prevalence of diabetes and the aging population. More than 500 million individuals worldwide are affected by this chronic disease, resulting in significant human and economic costs^3,4^. However, predicting CVD risk in T2D remains a challenge, and existing risk algorithms, such as the UK Prospective Diabetes Study (UKPDS) Risk Engine and Framingham Risk Score (FRS), have shown only modest predictive value in external validation studies^5–7^. Thus, it is essential to identify or develop readily available and cost-effective measures that can accurately identify individuals with a higher absolute risk of developing CVD beyond the risk estimated from established risk factors.

Precision medicine provides a promising approach to optimize risk prediction by integrating multidimensional data (i.e., genetic, clinical, sociodemographic), accounting for individual differences^8^. Recognizing the potential value of precision medicine in improving diabetes prevention and care, the Precision Medicine in Diabetes Initiative (PMDI) was established in 2018 by the American Diabetes Association (ADA) in partnership with the European Association for the Study of Diabetes (EASD) and is led by global leaders in precision diabetes medicine^9^. This systematic review is written on behalf of the ADA/EASD PMDI as part of a comprehensive evidence evaluation in support of the 2^nd^ International Consensus Report on Precision Diabetes Medicine^10^. As part of this broader initiative, we conducted a systematic review and meta-analyses addressing precision prognosis for CVD outcomes.

While previous systematic reviews of biomarkers for prediction of CVD have been conducted in the general population^11–25^, this review focused on patients with T2D. We sought to answer two questions: (1) Which novel markers predict CVD in people with T2D? (2) Is there any evidence that these markers enhance risk prediction beyond current practice? Addressing these questions may inform the development of more effective strategies for detecting and predicting CVD in individuals with T2D, ultimately leading to improved management and prevention of this complication.

Therefore, to identify those biomarkers with most promising clinical utility for CV risk assessment, we followed a rigorous stepwise approach, including evaluation of the incremental value of each biomarker beyond traditional risk factors (i.e. with evaluation of improvement in different metrics such as c-statistic and net reclassification improvement – NRI), as recommended by the statement from the American Heart Association for identification of novel markers for CV disease ^26^.

In summary, employing a stringent study selection process, this systematic review and meta-analysis identified four prognostic factors with high predictive utility, supported by moderate to high-strength evidence. Furthermore, three prognostic factors demonstrated moderate predictive utility, backed by low to moderate-strength evidence, and six prognostic factors showed low predictive utility, with evidence levels ranging from low to moderate. Risk scores demonstrated modest discrimination on internal validation, with diminished performance in external validation, particularly in cohorts diverging from the original population.

## Methods

As a reporting guidance, we followed the Preferred Reporting Items for Systematic reviews and Meta-Analyses (PRISMA) statement^27^. Figure 1 presents the PRISMA flow diagram, illustrating the process that led to the final selection of studies for review. Prior to data collection, the proposed systematic review and meta-analysis was registered on PROSPERO (Registration number: CRD42021262843).

**Figure 1.**
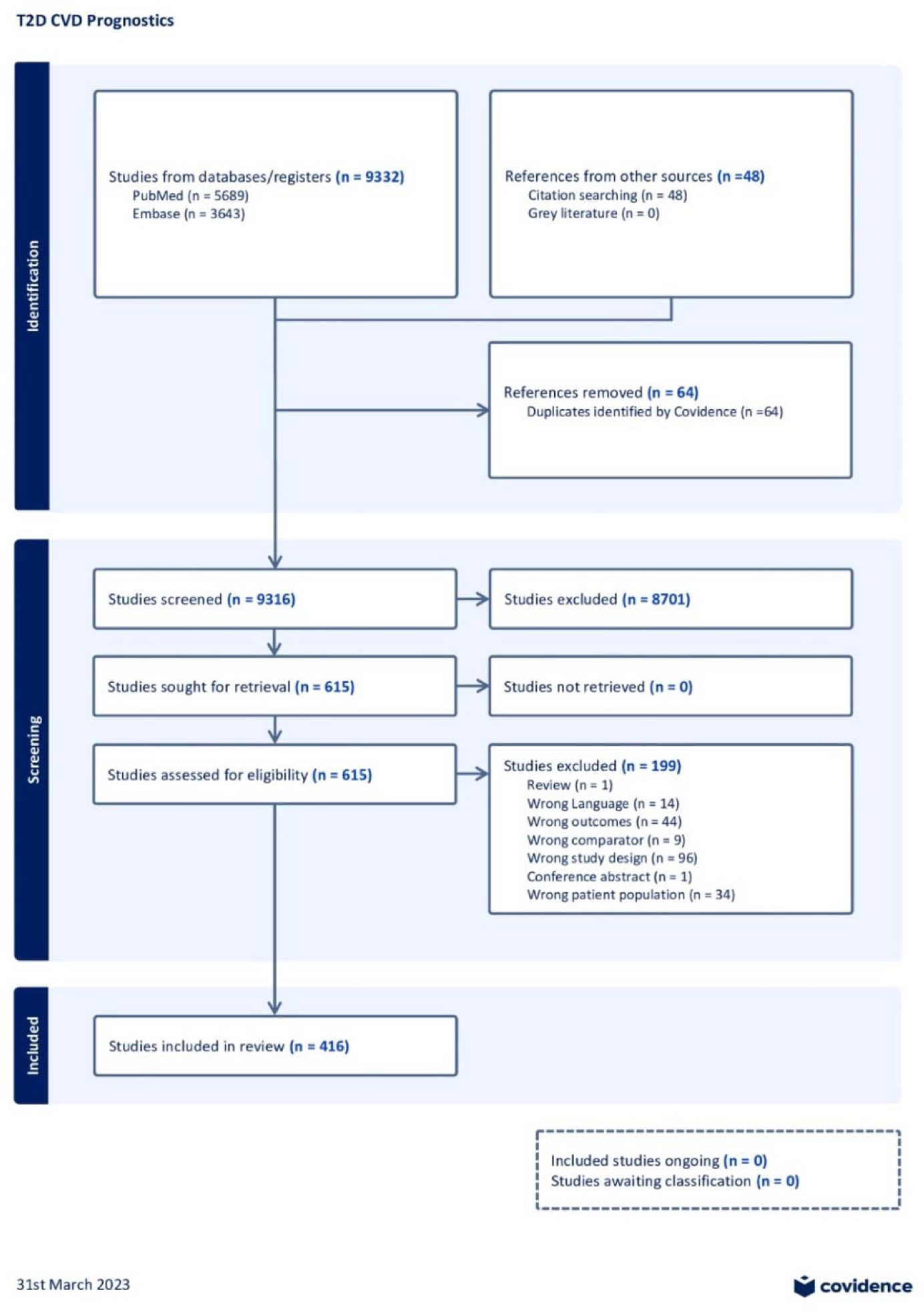
PRISMA flow diagram detailing the process that led to final study inclusion for review.

### Inclusion and Exclusion Criteria

This review included longitudinal studies (prospective or retrospective cohorts, including secondary analyses of cohorts from randomized controlled trials) of participants with T2D (youth-onset and adult-onset). Inclusion criteria included observational studies published from 1990-2021 that reported on the association between a prognostic factor or risk score and one or more CVD outcomes among participants with T2D. During the period of our review, the diagnostic criteria for T2D underwent some modifications (e.g. change in fasting glucose threshold, addition of hemoglobin A1C). We accepted studies that reported the inclusion of participants with T2D as defined in each individual study. Exclusion criteria included cross-sectional studies, studies utilizing surrogate endpoints for cardiovascular (CVD) outcomes such as carotid intima-media thickness, endothelial dysfunction, and arterial stiffness, and studies including only participants with pre-diabetes or only participants with type 1 diabetes. Studies with mixed populations of diabetes were included only if results were reported separately for participants with T2D. Supplemental Table 1 summarizes the Participant Intervention Comparison Outcomes and Studies (PICOS) framework.

### Outcomes

Only studies reporting outcomes on fatal or non-fatal coronary heart disease (CHD) and/or cardiovascular mortality (either alone or as individual component of composite outcomes) were included. A broad definition of CHD, including any outcomes defined by terms such as myocardial infarction, ischemic heart events, cardiac events, coronary artery disease, and major cardiovascular events was used.

### Search Strategy

We conducted a comprehensive search on Medline and Embase of studies published from January 1990 to March 2021 using keywords and MeSH (Medical Subject Headings) terms relevant to T2D and CVD (see Supplemental Text 1). In addition, we searched the reference lists of eligible studies and systematic reviews to identify any further relevant studies. The search strategy was designed by a multi-professional team of researchers with expertise in precision medicine, clinical diabetes, cardiovascular disease, biomarker development and evaluation, genetic markers, and predictive analytics, supported by two librarians with expertise in conducting systematic reviews and meta-analyses. References identified were exported to EndNote (Clarivate Analytics) and imported to Covidence, where studies were assessed for eligibility. After the removal of duplicates, 14 authors participated in screening each title/abstract, and full-text articles were obtained if abstracts were considered eligible by at least one author. Each full-text article was assessed for inclusion independently by two authors (among 12 total authors), and disagreements were resolved by consensus.

### Data extraction

All data were extracted and coded by one author and reviewed by a second author to ensure data accuracy. After undergoing training to ensure consistency in the process, thirteen authors participated in the data extraction process (A.A., C.T., L.L., M.F.G., M.L.M., N.M., R.C.W.M., S.K., C.H., G.Y., Y.Z., M.D.P., S.C.T.). To minimize inter-reviewer variability and ensure consistency in data extraction, all authors underwent training sessions via video conferences and participated in mock assessments.

During data extraction, studies were classified into three categories based on the primary type of prognostic factors reported, namely biomarkers, genetic markers, and risk scores. Biomarkers were broadly defined as non-genetic laboratory tests, clinical conditions, socio-demographics, vital signs, diagnostic procedures, and imaging tests. Genetic markers included specific DNA sequences or variations, such as single nucleotide polymorphisms (SNPs), restriction fragment length polymorphisms (RFLP), or short tandem repeats (STR). Risk scores were defined as predictive models, algorithms, or risk calculation tools that estimated the overall likelihood or category of cardiovascular disease (CVD) based on a set of risk factors. When multiple genetic variants were combined to predict risk (using SNPs), the study was classified as a genetic marker (i.e., genetic risk score) rather than a risk score. Additional details about the included studies can be found in Supplemental Text 2.

The following data were extracted from each article using a standardized data form in Covidence and Excel data tables: *study characteristics* (country or countries of the study population, study start and end year, study design, inclusion/exclusion criteria, study setting, data sources), *participant characteristics* (years of follow-up, follow-up duration, total number of participants, race/ethnicity/ancestry, and baseline characteristics), *prognostic factor(s) characteristics* (name, prognostic factor type, units of measurement, units and cut-offs in regression analyses, transformation methods, effect measures [hazard ratio, odds ratio, c-statistic, net reclassification improvement (NRI), integrated discrimination index (IDI), etc.] and 95% confidence intervals, adjusted covariates), and *outcomes* (CVD outcome definition, number of events and non-events), and validation methods. For genetic markers, we collected risk variants, risk alleles, and closest gene (locus).

For continuous variables, we collected mean and standard deviation or median and interquartile range as reported in the study. We collected fully adjusted effect measures (HR, RR, OR, c-statistic) and their corresponding 95% CIs reported in the original articles. When studies reported multiple multivariate-adjusted effect measures, we collected the estimate from the most fully adjusted model. We did not contact primary authors to obtain data that were not reported. Furthermore, data were collected to evaluate the *risk of biases* in each study as summarized in Supplemental Table 2 and described in the quality assessment paragraph.

### Quality Assessment

We used a modified Newcastle-Ottawa Scale (NOS) to assess quality and risk of biases (Supplemental table 2). The scale assesses studies based on six common domains, including representativeness of the exposed and non-exposed cohorts, ascertainment of exposure and outcome, and adequacy of study follow-up for primary and secondary CVD events, as well as the adequacy of cohort follow-up^28^. For biomarker studies, we added two additional domains to the NOS to address bias due to confounding by evaluating the number of covariates and established CVD risk factors included in the adjusted models. Each study was given a score for each domain and an overall quality evaluation was determined by adding up these scores. The possible range of scores for non-genetic biomarkers, based on 8 domains, was 2 to 28, while for genetic biomarkers and risk scores, scores ranged from 2 to 18 based on 6 domains. Two authors assessed study quality independently, and a third author resolved any disagreements.

We reported the overall risk of bias based on the distribution of scores in each prognostic factor category, with higher scores representing lower risk of bias. Studies in the top, second, and lowest tertiles (according to the distribution specific for each type of study, i.e. non-genetic biomarkers, genetic biomarkers and risk scores) were considered to have low, medium, and high risk of bias, respectively. The score of each domain was also classified as low, medium, or high risk of bias for graphical purposes, as clarified in Supplemental Table 2.

### Statistical Analysis

A random-effects model was used to pool the overall effect estimates in all meta-analyses, only if the heterogeneity test was statistically significant. For studies reporting the same effect measure (e.g. HR), we calculated the pooled effect estimate with 95% CIs for each biomarker or genetic marker and assessed heterogeneity between studies using the Cochran’s *Q* statistic (*p*lJ<0.1),the *I*^2^ index >75%, and/or τ^2^. Due to the limited number of studies per prognostic factor, subgroup analyses by population characteristics or outcomes were not performed. We performed sensitivity analyses by excluding studies with high risk of bias. As the number of studies per prognostic factor was always less than 10, we were unable to assess publication bias using funnel plots. We used R, version 4.2.3 (R Project for Statistical Computing), with the “meta”, “metafor”, and “forestplot” packages for all analyses^29^. Two-sided statistical tests were used with a significance threshold of <0.05.

### Strength of the Evidence

We considered aspects of the GRADE approach^30^ and the JBI critical appraisal tools^31^ in grading the strength of evidence for individual biomarkers and genetic markers/risk scores. We applied relevant GRADE criteria, including indirectness, inconsistency, and imprecision, throughout the study. Since we only included studies that involved patients with T2D and a “hard” clinical CVD outcome, the evidence is considered direct by definition. We analyzed the results from T2D patients with and without baseline CVD and specified all relevant CVD outcomes to assess the applicability of individual biomarkers in specific populations and outcomes. To ensure robustness and validity of our findings, we established strict eligibility criteria, excluding studies that did not adjust for established CVD risk factors (listed in Supplemental Table 4). Furthermore, we scored studies based on the adequacy of adjustment for covariates, including the total number of covariates and established CVD risk factors, in accordance with the JBI criterion for statistical adjustment of confounders.

We used the American Heart Association scientific consensus report for stepwise evaluation of novel markers for CVD risk^26^ to identify promising biomarkers and genetic markers based on their strength of evidence progressing from measures of association, discrimination, improvement in discrimination, net reclassification index (NRI) or integrated discrimination index (IDI). This approach is summarized in Supplemental Table 3 (modified from^26^). For biomarkers and genetic markers, we progressed from those with significant adjusted association in at least one study to those with net positive number of studies showing significant association in a consistent direction. The net positive number of studies was calculated by summing up all studies with positive association and subtracting studies with no association (e.g., three studies showing positive association and two studies with no association yielded a net positive number of one). We identified biomarkers that improved prediction performance when added to established models, based on improvement in at least one of c-statistic, NRI (the probability that a person is appropriately classified into either high-or low-risk), or IDI (quantification of predicted probabilities of events and non-events based on inclusion of the biomarker in the model), and further narrowed down the list to those with improvement in all three indicators.

Accordingly, for each of the prognostic factors that passed our evidence-based screening criteria, predictive utility was classified as *high* (3 points), *moderate* (2 points), or *low* (<2 points) based on three criteria: number of studies with all three performance indicators satisfied (1 point if >0 studies, 0 points if 0 studies), number of pooled meta-analyses showing significant association (1 point if >0, 0 points if 0 studies), non-pooled analysis showing ≥75% of studies had a significant association (1 point if yes, 0 points if no). Strength of Evidence was classified as *high* (4 points), *moderate* (2 or 3 points), or *low* (<2 points) based on four criteria: at least one meta-analysis was conducted regardless of outcome (1 point if yes, 0 points if no), exclusion of high risk of bias studies did not alter inferences from meta-analyses (1 point if unaltered, 0 points if altered), exclusion of high risk of bias studies did not alter inferences from non-pooled analyses (1 point if unaltered, 0 points if altered), and consistencies in the definition of the prognostic marker used in analyses (1 point if yes, 0 points if no).

For the risk scores, we provide a complete assessment of risk of bias and pooled c-statistics; however, we decided not to conduct a corresponding stepwise approach to evidence grading as explained above for biomarkers/genetic markers due to the complexity in verifying specifications of each model over time and across comparisons. Inferences from the risk score results are here meant to guide future work that would permit analyses to handle this complexity.

## Results

### Study selection and characteristics

Out of 9380 studies identified from databases/registries (N=9332) and other sources (N=48), there were 9316 unique studies after removing 64 duplicates. Of these, 615 articles were selected for full-text review, and finally, 416 articles were considered appropriate for inclusion in the analysis^32–291,292–447^. Outcomes were reported for 321 biomarker studies, 48 genetic marker studies, and 47 risk score/model studies, as shown in Supplemental Tables 5, 6, and 7. Figures 1 and 2 provide an overview of the screening and selection process.

**Figure 2.**
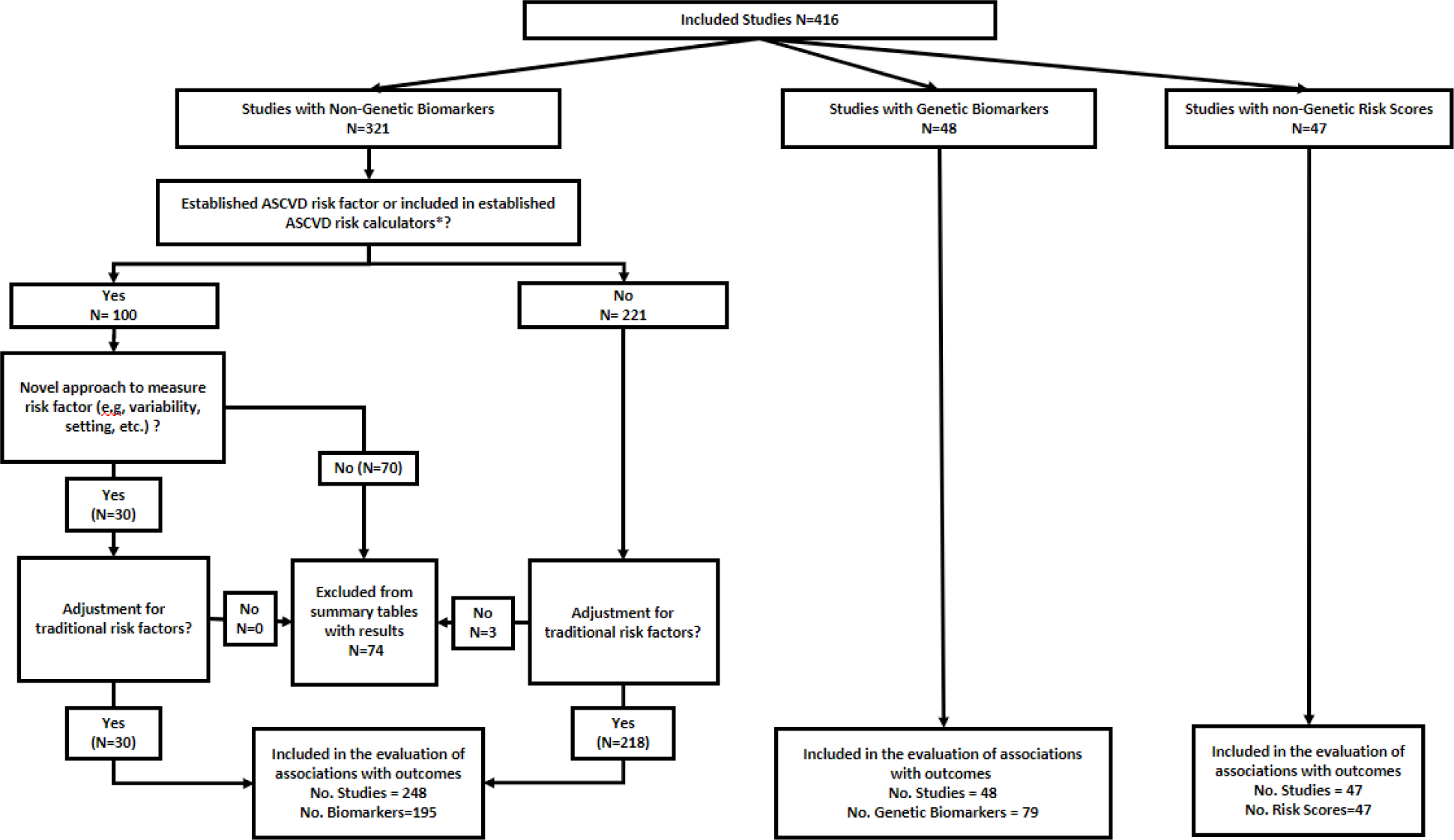
Selection of studies to be included for evaluating the associations of biomarkers, genetic markers and non-genetic risk scores with cardiovascular outcomes

Predominant ancestry in the studied populations were European (57.1%), East Asian (19.7%), South Asian (5.5%) and Hispanic or Latin American (4.2%). Geographically, the United States, United Kingdom, China, Japan, and Italy were the top five represented countries with regards to origin of study participants and author affiliation in the included studies. Figure 3 and online interactive figures (https://hugofitipaldi.shinyapps.io/T2D_prognostic/) offer a detailed breakdown of ethnic and geographic distributions.

**Figure 3.**
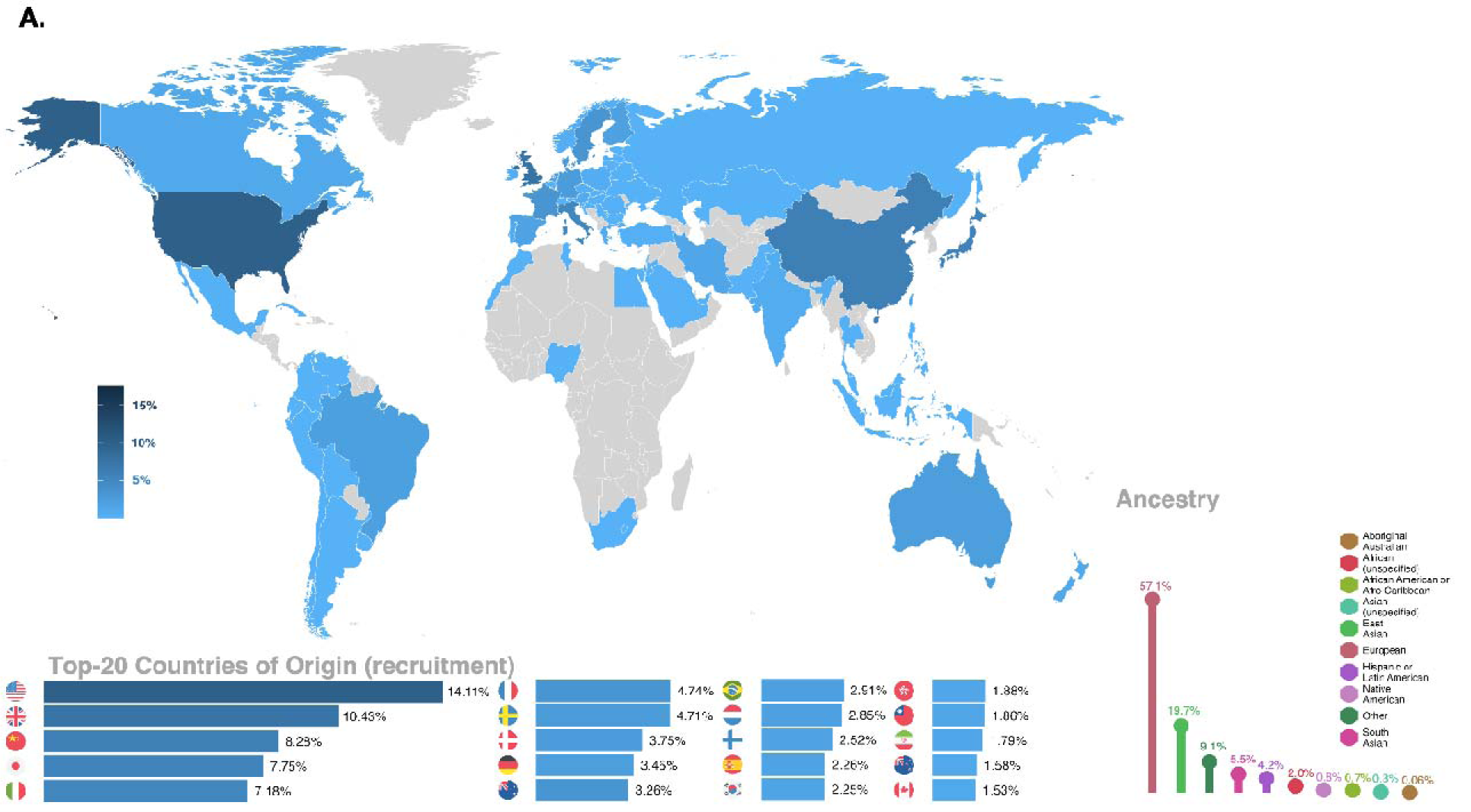
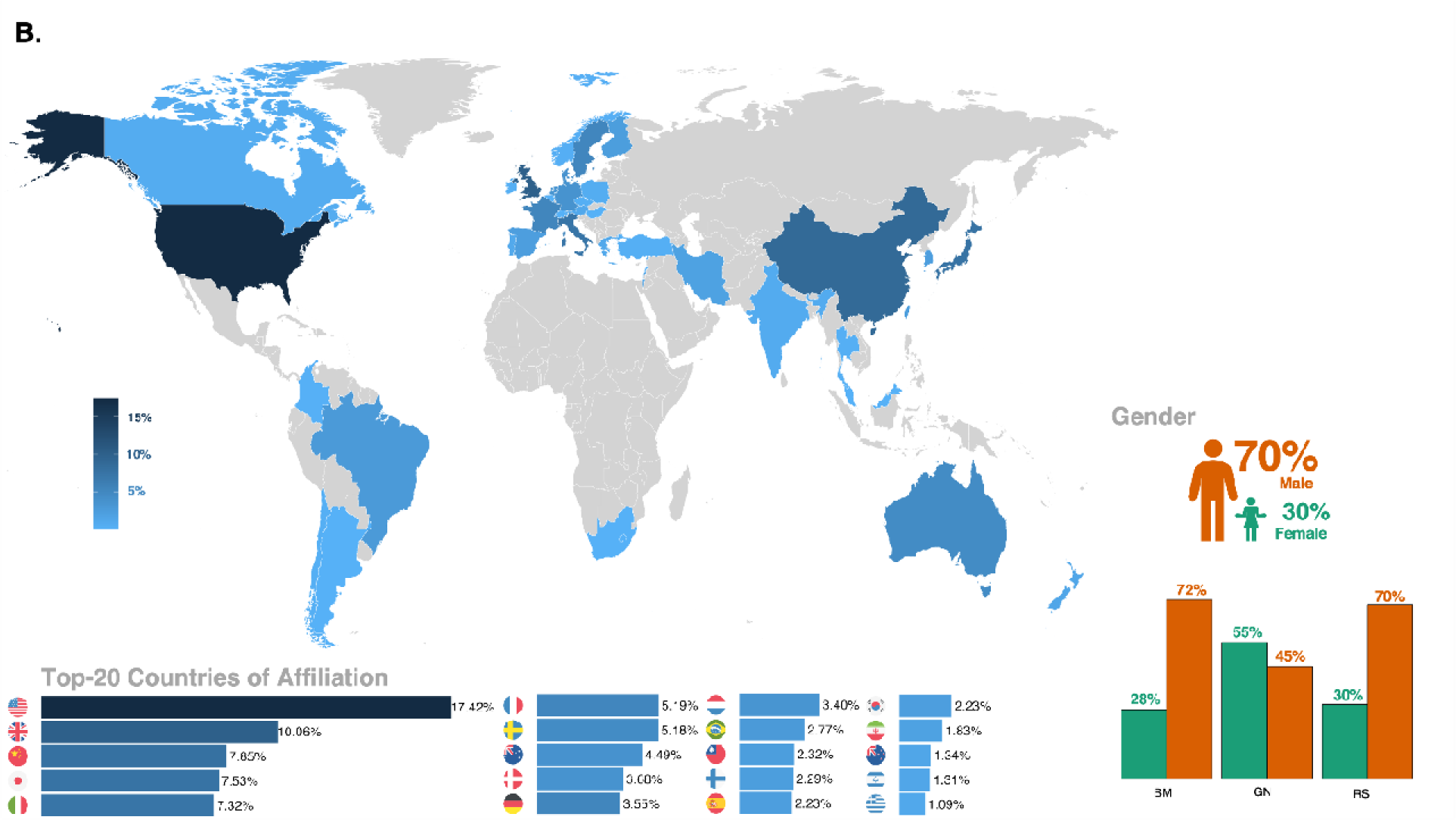
A. Top 20 countries of origin and ancestry of the study populations in the included studies. B. Top 20 countries of affiliation and gender distribution of authors for biomarker (BM), genetics (GN) and risk score studies. The data used for this visualization was obtained from PubMed and PubMed Central through manual curation and by applying text mining functions developed using R software version 4.1.2. The final proportions of ancestries were calculated for each unique study and then aggregated as described in detail here: ^448^

### CVD Outcomes

There was heterogeneity in the CVD outcomes evaluated across the analyzed studies (see Supplemental Figure 1). The median duration of follow-up reported across studies was 5 years (IQR 3.1 to 7.8 years). The most frequently reported outcomes were coronary heart disease, cardiovascular mortality, and stroke, either individually or combined. The vast majority (87%) of studies had a clearly defined outcome based on ICD-10 codes, clinical documentation, or adjudication, with 9% relying on registry or record linkage, and 4% using either patient self-report or having an unclear definition. We classified primary prevention as the prediction of CVD in individuals without a history of the disease, secondary prevention as the prediction of recurrent CVD events or CVD progression in those already diagnosed with the disease, and mixed populations as a combination of both primary and secondary prevention.

### Biomarkers

Among 416 included studies, 321 (77.2%), 48 (11.5%), and 47 (11.2%) were studies of non-genetic biomarkers, genetic biomarkers, and non-genetic risk scores, respectively. Among the 321 studies of non-genetic biomarkers, 70 (21.8%) evaluated established CVD risk factors and were excluded, while 30 studies (9.3%) were included because they used a novel approach (e.g., variability, setting) for an established risk factor (Figure 2). Further, three studies did not adjust for any CVD risk factors and were excluded, leaving 218 studies consisting of 195 unique biomarkers in the analysis.

Among these 195 biomarkers analyzed, 134 (69%) had a significant adjusted association for predicting CVD, based on a net positive number of studies (Figure 4A and Supplemental Table 8). Out of these, 12 (9%) showed improvement in c-statistic, NRI, or IDI in *more than one study*: N-terminal pro b-type natriuretic peptide (NT-proBNP), C-reactive protein (CRP), troponin T (TnT), coronary artery calcium score (CACS), coronary computed tomography angiography (CCTA), single-photon emission computed tomography (SPECT) scintigraphy, pulse wave velocity (PWV), galectin-3 (Gal-3), troponin I (TnI), carotid plaque, growth differentiation factor-15 (GDF-15), and triglyceride-glucose (TyG) index. The following biomarkers showed prediction performance but in *only one study*: SPECT, TnI, TyG, 25-hydroxyvitamin D, poly (ADP-ribose) polymerase (PARP), and interleukin-6 (IL-6).

**Figure 4.**
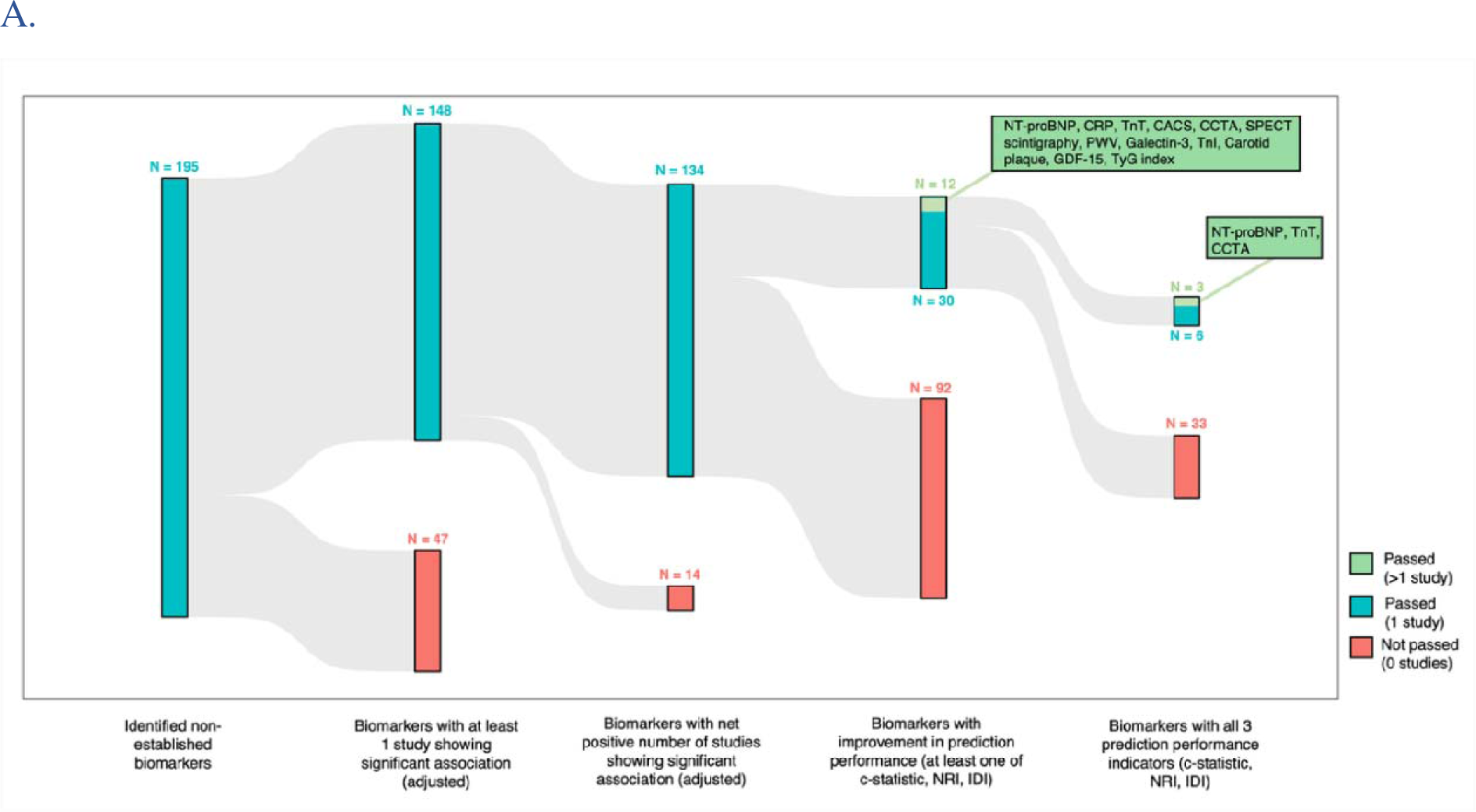
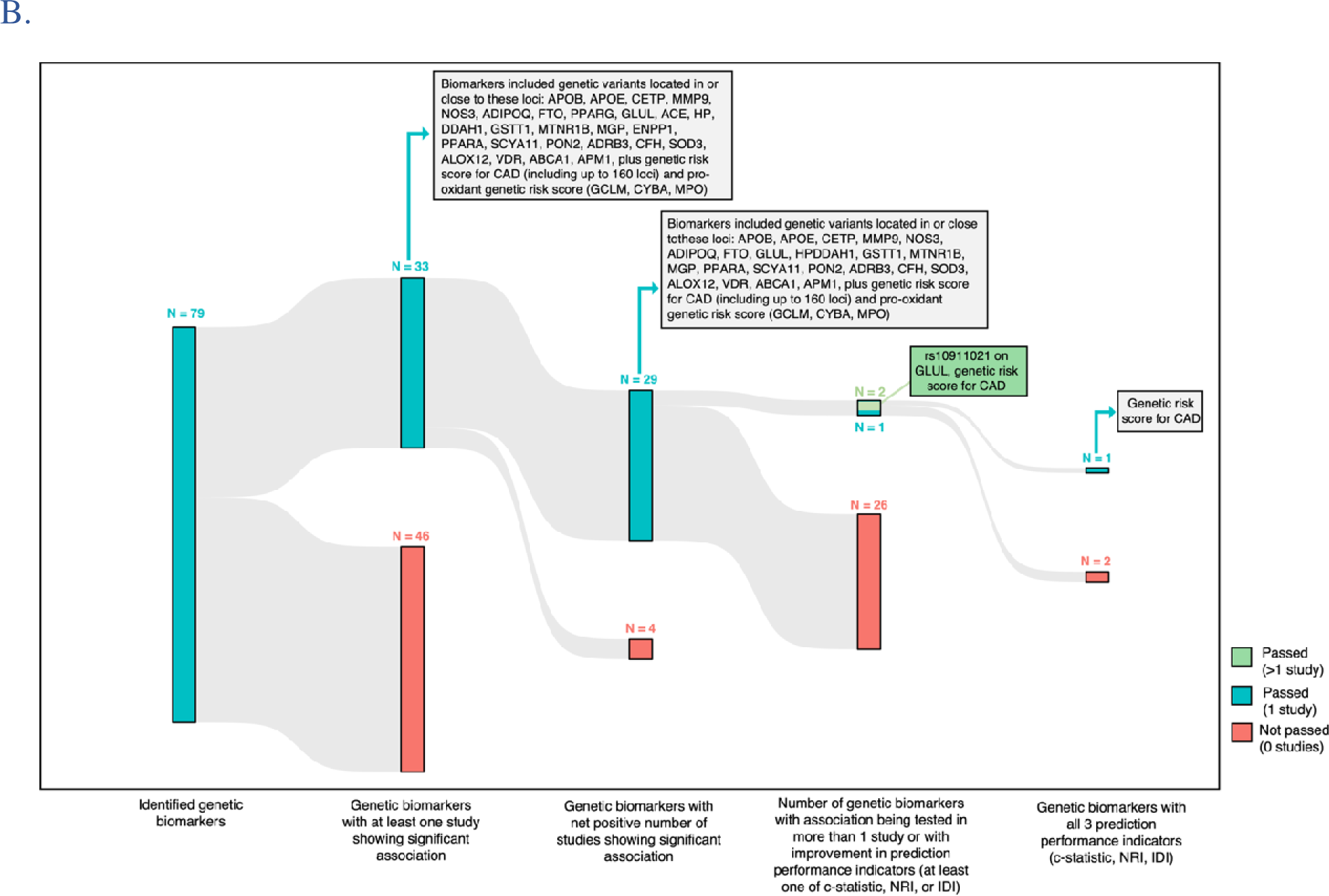
Sankey diagram showing the funneling of identified non-genetic (A) and genetic (B) biomarkers through sequential filtering steps based on the criteria specified at the bottom of the diagram. The number of biomarkers passing or not passing each step is depicted at the top of the colored bars, with biomarkers passing all steps having the strongest predictive performance value.

Biomarkers with *all three* prediction performance indicators satisfied in *more than one study* were NT-proBNP, TnT, and CCTA, with results summarized in Table 1. For NT-proBNP, 5 studies reported improvement in c-statistics ranging from 0.01 to 0.07, significant increase in NRI ranging from 0.04 to 0.50, and significant rIDI ranging from 0.012 to 0.48 (in four studies). For TnT, 3 studies reported improvement in c-statistics ranging from 0.02 to 0.10, significant NRI ranging from 0.150 to 0.44, and rIDI ranging from 0.03 to 0.05. For CCTA, 3 studies reported improvement in c-statistics ranging from 0.08 to 0.35, with one study reporting statistically significant improvements in NRI of 0.55 and rIDI of 0.046. Of these three biomarkers, NT-proBNP showed the strongest incremental predictive value based on the magnitude of these indicators. Supplemental Table 9 shows the degree of variation in measurement methods used for each of these biomarkers.

**Table 1.** Performance of the prediction of 3 biomarkers with the most evidence. Notes: Data on improvement in C-statistics was collected from the study, either as reported or derived by comparing the C-statistic from the reference model with the C-statistic obtained from the combination of the reference model and the novel biomarker. NR, Not Reported; SBP, systolic blood pressure; HF, heart failure; MI, myocardial infarction; BMI, body mass index; AF, atrial fibrillation; PAD, peripheral artery disease; T2D, type 2 diabetes; eGFR, estimated glomerular filtration rate; ACR, albumin-creatinine ratio.

| Biomarker | Study | Clinical factors / biomarkers to be compared | Improvement in C-statistics |  | NRI |  | IDI |  |
| --- | --- | --- | --- | --- | --- | --- | --- | --- |
|  |  |  | Yes /No | Estimate (95% CI) and *P*-value | NRI (95% CI) | *P*-value | IDI (95% CI) | *P*-value |
| NT-proBNP | Sharma 2020 | Age, sex, SBP, history of HF, duration of diabetes, prior MI, hypertension, hyperlipidemia, smoking, eGFR. | Yes | 0.050 (CI and *P*-value NR) | 0.385 (0.304 - 0.472) | NR | 0.088 (0.077 - 0.098) | <0.05 |
|  | Wolsk 2017 | Prior MI, BMI, NSTEMI (index event), heart rate, HbA<sub>1c</sub>, percutaneous coronary intervention at the index event, cerebrovascular disease, AF, prior HF, sodium concentration, macroalbuminuria, PAD, age, and LDL concentration. | Yes | 0.01 (CI NR), *P* <0.05 | 0.106 (5.70 - 16.60) | <0.05 | 0.080 (0.030 - 1.600) | <0.05 |
|  | Wong 2019 | Age ≥ 65 years, Male, T2D, Hypertension | Yes | 0.03 (CI NR), *P* = 0.001 | 0.345 (0.238 - 0.451) | <0.001 | 0.012 (0.007 - 0.017) | <0.001 |
|  | Scirica 2016 | Treatment arms, age, SBP, sex, history of HF-, duration of diabetes, prior MI, history of hypertension, history of hyperlipidemia, smoking, and eGFR | Yes | 0.07 (CI NR), *P* <0.001 | 0.038 (0.032 - 0.044) | <0.05 | 0.480 (0.410 - 0.550) | <0.05 |
|  | Van der Leeuw 2016 | Female sex, age at diabetes diagnosis, duration of diabetes, HbA<sub>1c</sub>, square of HbA<sub>1c</sub>, SBP, square of SBP, TC/HDL ratio, urinary ACR, current smoking status, history of major macrovascular disease | Yes in SMART study | 0.02 (0.00 - 0.04), *P*-value NR | 0.270 (0.100 - 0.440) | <0.05 | NR | NR |
|  | Van der Leeuw 2016 | Female sex, age at diabetes diagnosis, duration of diabetes, HbA<sub>1c</sub>, systolic blood pressure, TC/HDL ratio, eGFR, current smoking status, history of major macrovascular disease | Yes in EPIC-NL study | 0.02 (0.00 - 0.05), *P*-value NR | 0.500 (0.260 - 0.730) | <0.05 | NR | NR |
| TnT | Lepojarvi 2016 | Age, sex, history of acute MI, BMI, Canadian Cardiovascular Society grading of angina pectoris, left ventricular ejection fraction and mass index, HDL cholesterol, ACR, HbA<sub>1c</sub>, and type of glucose metabolism disorder | Yes | 0.101 (CI and *P*-value NR), | 0.231 (0.067 - 0.394) | <0.01 | 0.054 (0.031 - 0.077) | <0.001 |
|  | Scirica 2016 | Treatment arms, age, SBP, sex, history of HF, duration of diabetes, prior MI, history of hypertension, history of hyperlipidemia, smoking, and eGFR | Yes | 0.07 (CI NR), *P* < 0.001 | 0.440 (0.380 - 0.510) | <0.05 | 0.029 (0.023 - 0.034) | <0.05 |
|  | Rørth 2019 | Natural logarithm of NT-proBNP, age, sex, treatment effect, ejection fraction, NYHA class, BMI, heart rate, SBP, creatinine, LDL, prior angina pectoris, AF and pacemaker implantation. | Yes | 0.018 (CI NR), *P* = 0.02 | 0.150 (0.051 - 0.261) | 0.007 | 0.032 (0.012 - 0.064) | <0.001 |
| CCTA | Lee 2017 | Age, male sex, HTN, smokers, hyperlipidemia, eGFR, and HbA<sub>1c</sub> | Yes | 0.072 (CI NR), *P* = 0.0349 | 0.550 (0.343 - 0.757) | <0.0001 | 0.046 (0.020 - 0.072) | 0.0006 |
|  | Halon 2016 | UKPDS and log CAC Score | Yes | 0.35 (CI NR), *P* = 0.021 | 0.632 (CI NR) | NR | 0.647 (CI NR) | NR |

Forest plots in Figure 5A show the HRs for 11 studies evaluating NT-proBNP, conducted in heterogeneous populations (2 primary, 5 mixed, and 4 secondary), outcomes, units in regression analyses (i.e., SD, SD of log), and laboratory units (ng/L, pg/mL). Nonetheless, all studies except one showed a significant association with a CVD outcome. Eight out of 11 (73%) studies were assessed to be at low risk of bias. Figure 6A and Supplemental Figures 2A and 2B show the meta-analysis of NT-proBNP as a continuous variable per logarithmic and per 1 SD unit increase, confirming the highly significant association with CVD (pooled HR 1.53, 95% CI 1.26-1.85 per log increase; pooled HR 1.59, 95% CI 1.27-1.99 per SD increase) after accounting for heterogeneity with the random effects models (I^2^ 90% and I^2^ 83%, respectively). Interestingly, although our review excluded studies focusing exclusively on heart failure patients, among three studies that incorporated EF as a covariate in their models, NT-proBNP was shown to have predictive value for cardiovascular outcomes independent of EF ^241,326,398^ (Supplemental Table 9).

**Figure 5.**
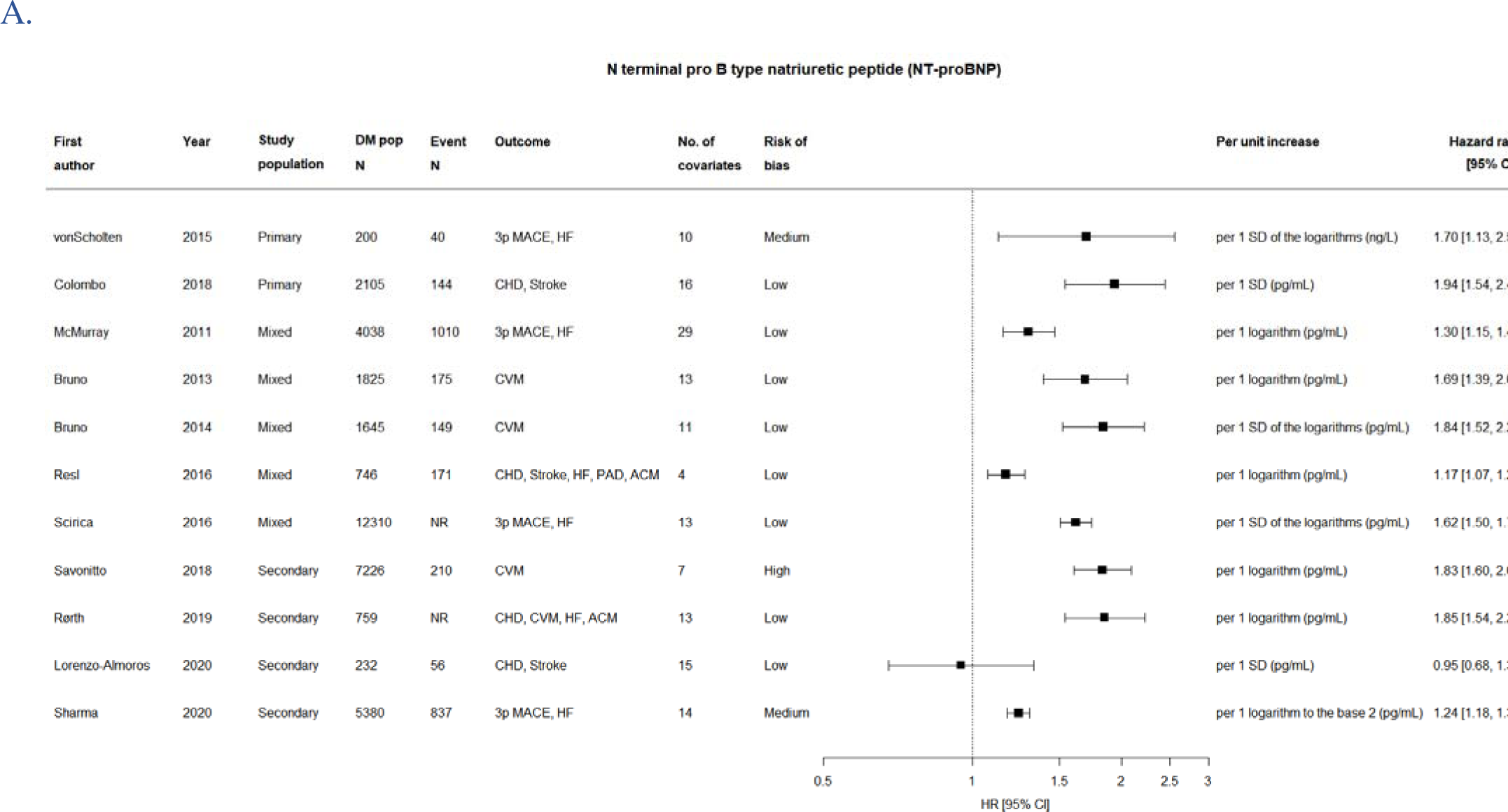
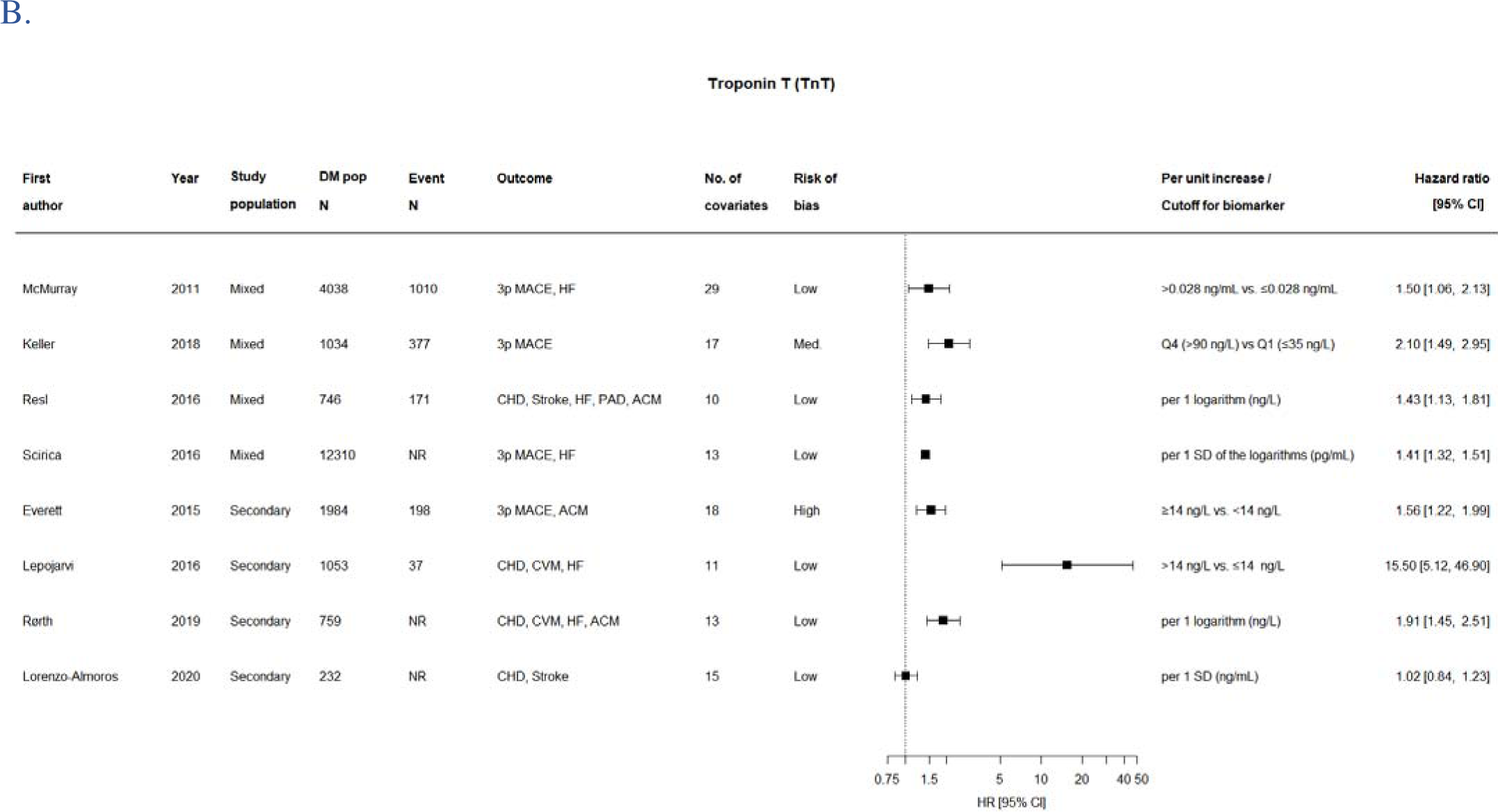
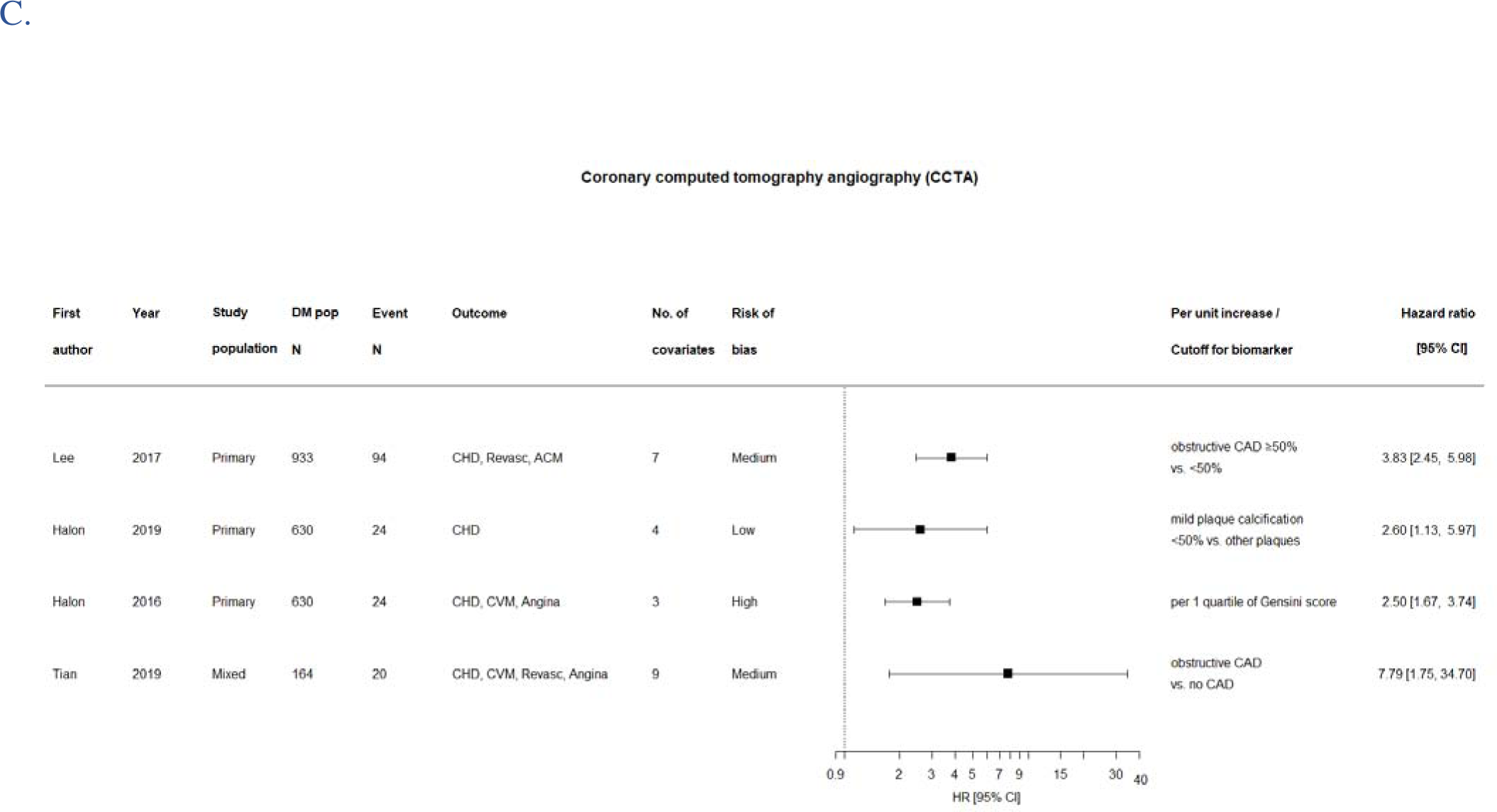
Forest plots for three biomarkers (A. NT-proBNP, B. TnT and C. CCTA) with the most evidence for prediction of CVD outcomes. HR, hazard ratio; CI, confidence interval; DM pop N, sample size for diabetes population; Event N, number of individuals developed CVD outcomes; 3p MACE, 3-point major adverse cardiovascular events; HF, heart failure; CHD, coronary heart disease; CVM, cardiovascular mortality; PAD, peripheral artery disease; ACM, all-cause mortality

**Figure 6.**
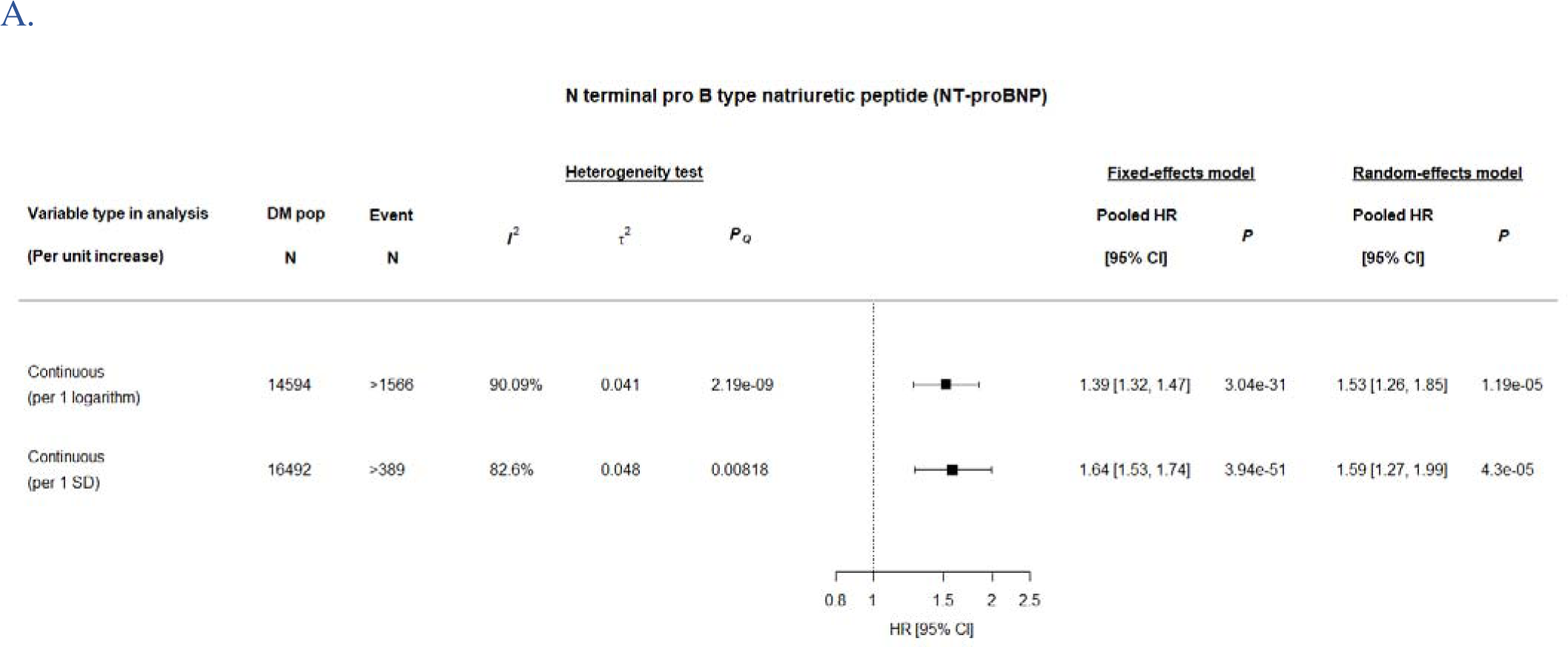
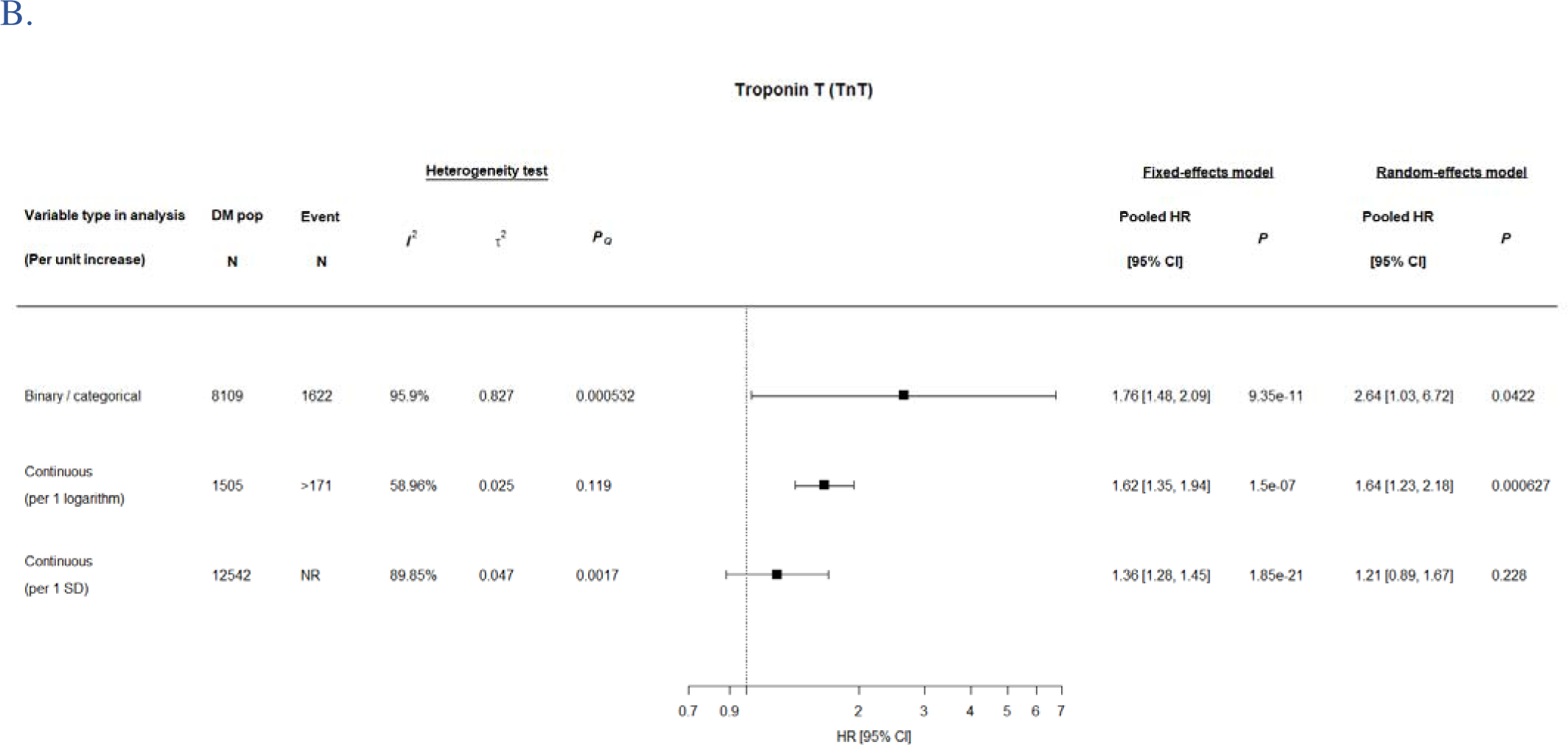
Meta-analysis of A. NT-proBNP and B. TnT for predicting cardiovascular outcomes. Note: *PQ* is the *p*-value obtained from the Cochran’s Q test. HR, hazard ratio; CI, confidence interval; DM pop N, sample size for diabetes population; Event N, number of individuals developed CVD outcomes

Forest plots in Figure 5B show the HRs for 8 studies evaluating TnT, conducted primarily for mixed or secondary populations with variable CVD outcomes. Studies differed with respect to cut-offs and categories for TnT, units of measurement (ng/ml, ng/L) and analysis (per log, per 1 SD log). Among these studies, all but one showed a positive association. Notably, the study by Lepojarvi 2016 was an outlier in its magnitude of effect and confidence intervals. Overall, for TnT, study quality was good with 6 out of 8 (75%) assessed to be at low risk of bias ^227^. A significant association for TnT was observed in studies where the biomarker was evaluated as a continuous variable per 1 log increase with pooled HR 1.64 (95% CI 1.23, 2.18) and I^2^ 59% (Figure 6B and Supplemental Figure 2C); similarly, when treated as a binary or categorical variable, the pooled HR was 2.64 (95% CI 1.03, 6.72) with I^2^=95.9% (Figure 6B and Supplemental Figure 2D). However, when treated as a continuous variable per 1 SD, there was no longer a significant association in a random effects model (Figure 6B and Supplemental Figure 2E).

Forest plots in Figure 5C show the HRs for 5 studies evaluating CCTA conducted primarily for primary CVD prevention with variable CVD outcomes. Studies differed significantly with respect to CCTA definition of subclinical or clinical CHD. All 5 studies showed a significant association; however, 2 of the 5 studies (40%) were assessed to be at a high risk of bias.

Apart from these three biomarkers, SPECT, TnI, TyG, 25-hydroxyvitamin D, poly(ADP-ribose) polymerase (PARP), and interleukin-6 (IL-6) showed prediction performance in *all three* performance indicators but in *only one study*. Forest plots for the remaining 9 biomarkers that showed improvement in *at least one* performance indicator in *more than one study* (CACS, carotid plaque, CRP, gal-3, GDF-15, PWV, SPECT scintigraphy, TnI, and TyG) are shown in Supplemental Figure 3. Again, there was substantial heterogeneity with respect to study populations, outcomes, and units of analysis for these biomarkers. Biomarkers showing positive association in at least 75% of studies included CACS, carotid plaque, gal-3, PWV, SPECT scintigraphy, TnI, and TyG. While CRP did not meet the threshold of 75% of studies showing an association, when meta-analyzed as a binary or categorical variable, it showed a significant pooled association; PWV and TyG also demonstrated significant association in pooled analysis (Supplemental Figure 4).

### Genetic markers

Among the 48 genetic studies analyzed (Supplemental Table 6), 79 genetic biomarkers were examined for their association with incident CVD events (Supplemental Table 10), mainly in populations of European (65%) or Asian (26%) ancestries, with sparse representation of populations of other ancestries (e.g., African 12% or Hispanic 3%), with 12% of associations being tested in mixed populations. Most of the studies (70 out of 79) used single variants as distinct genetic biomarkers (exposure), while 9 studies used a combination of different SNPs into genetic risk scores (GRS) as the exposure. Remarkably, most of these exposures were tested only in one study, and external validation was performed in only 4 out of 48 studies, with only one study using a longitudinal cohort as a validation set, i.e., GRS for CHD. Overall, among the 79 genetic biomarkers, 33 (41.8%) had *at least one* study showing significant association, out of which 29 had a net positive number of studies showing significant association. Out of these 29 genetic biomarkers, two were tested in *more than one* study (rs10911021 on *GLUL*, GRS for CHD [GRS-CHD]), one had improvement in *any* performance indicator in a single study (isoform e4 in *APOE*), and one had improvement in *all three* performance indicators in a single study (GRS-CHD) (Figure 4B).

Notably, the rs10911021 variant in *GLUL* was the only single variant that showed an association with CVD in several studies. This variant was initially identified in T2D patients using a genome-wide approach and subsequently confirmed for its association with CVD in selected populations from two additional studies. For GRS-CHD, four separate studies investigated the combination of up to 204 CHD variants from 160 distinct loci derived from the general population. These studies had distinct but overlapping and increasing numbers of loci and variants tested in more recent investigations. The most recently performed GRSs were externally validated and demonstrated significant improvements in CVD risk reclassification (cNRI) as well as notable enhancements of 8% in relative IDI (rIDI). However, these findings were identified in subjects of European ancestry and ancestry-specific analyses showed consistency in Asian subjects but not in other ancestral backgrounds. Forest plots for variants located on the GRS-CHD and *GLUL* are shown in Figure 7, while their meta-analyses can be found in Supplemental Figure 5.

**Figure 7.**
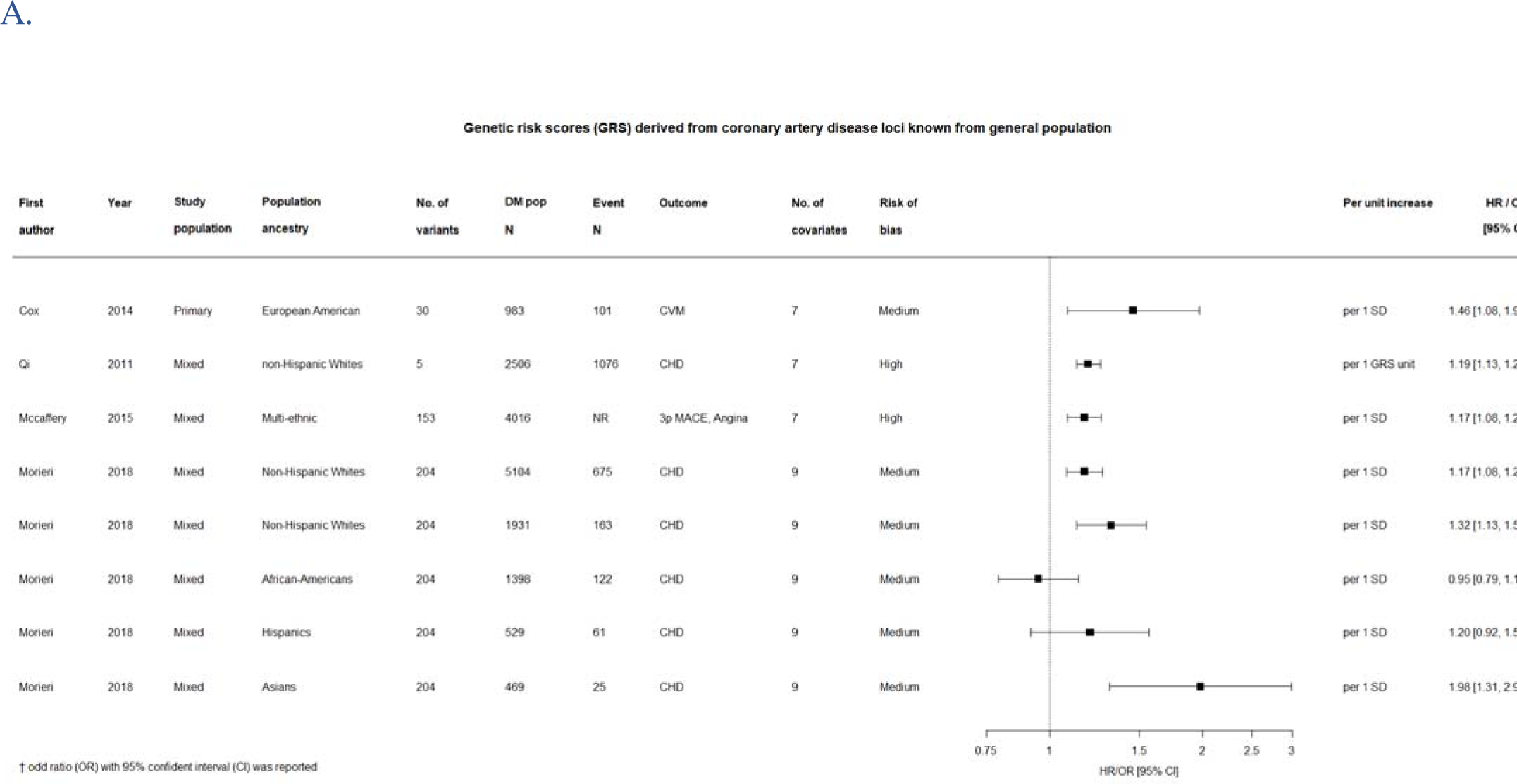
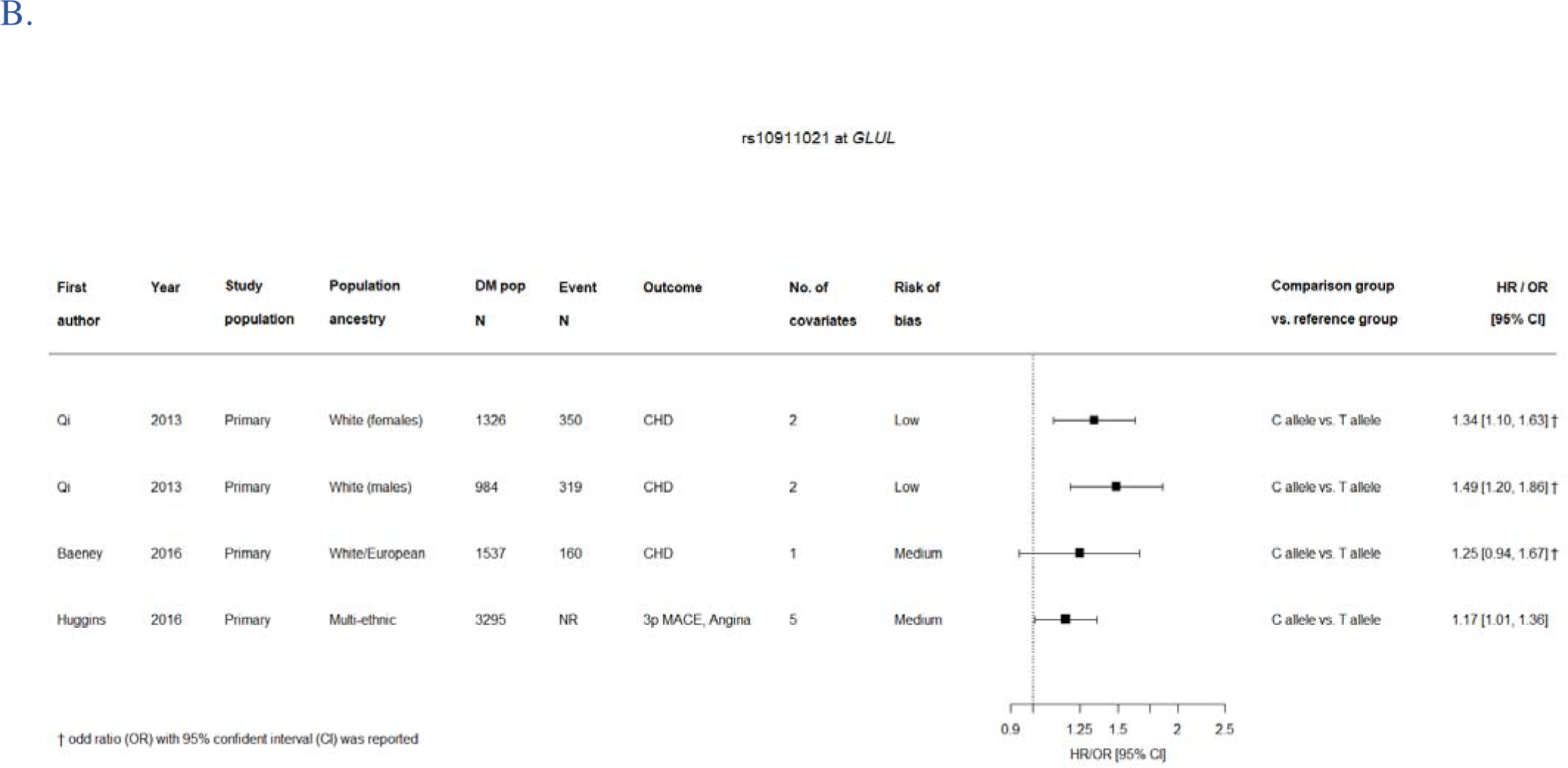
Forest plots of genetic markers for predicting cardiovascular outcomes. A. genetic risk scores, B. *GLUL* variant rs10911021. HR, hazard ratio; CI, confidence interval; DM pop N, sample size for diabetes population; Event N, number of individuals developed CVD outcomes; 3p MACE, 3-point major adverse cardiovascular events

### Risk scores/models

Forty-seven studies reported results of 27 unique CVD risk scores (Supplemental Tables 7 and 11). Figure 8A and 8B provides the c-statistics from internal and external validation analyses, respectively. On both internal and external validation, discrimination was modest. Most risk scores were developed in the United States, Europe, and East Asia and 61.1% of the internal validation studies were assessed to be at a high risk of bias. Model performance tended to decline when validated in countries that differed from the development cohort (Supplemental Figure 6). For example, the FDS study achieved high c-statistics (>0.80) when validated in an Australian cohort, but lower ones (0.58-0.69) when tested in European countries. In line with previous studies^6,7,387^, discrimination for the UKPDS and FRS was generally poor on external validation. Most prediction models focused on baseline characteristics and did not account for time-varying factors that may modify CVD risk (e.g., statin, SGLT-2i, GLP-1 RA). An exception was the BRAVO risk engine, published in 2020 and validated in trials of SGLT-2i patients, showing that this risk engine effectively predicted CV health benefits through improvements in common clinical measures (e.g., A1C, SBP, and BMI)^343^.

**Figure 8.**
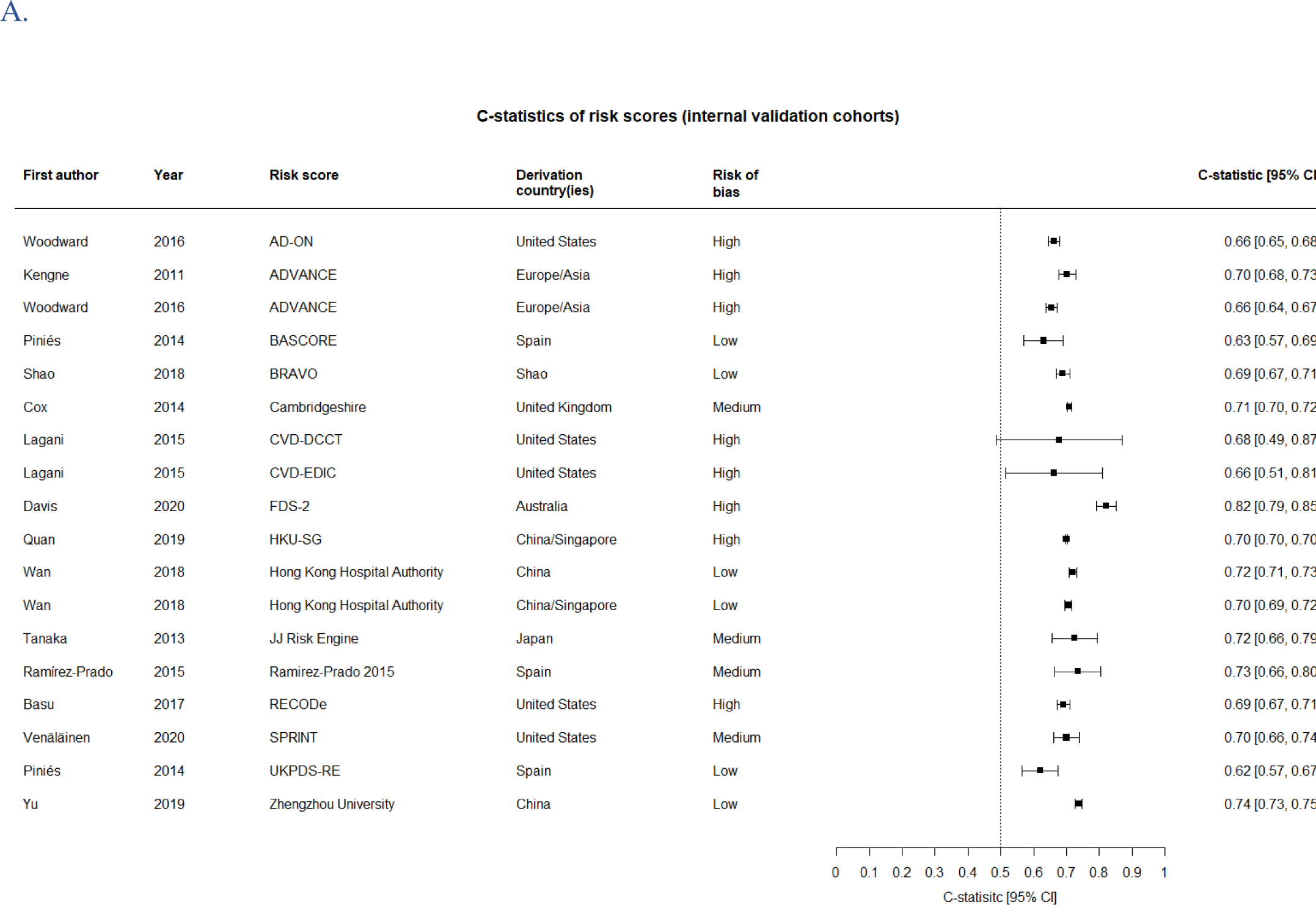
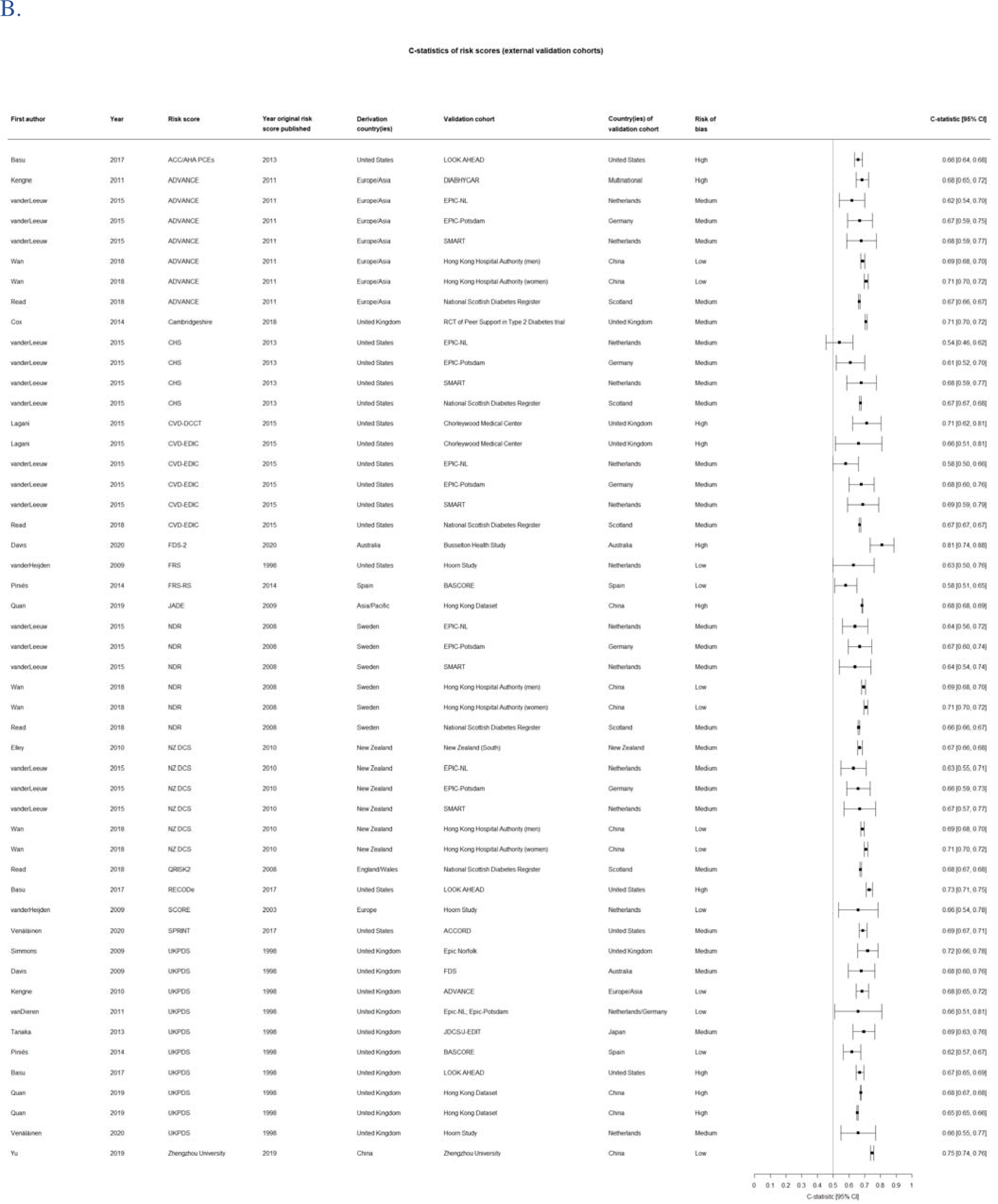
Summary of c-statistics of risk scores for predicting cardiovascular outcomes. Internal validation results are shown in Figure 8A and external validation results in Figure 8B. CI, confidence interval.

Supplemental Figures 7A-F provide the pooled c-statistics from external validation studies on those risk scores for which the analysis was possible: ADVANCE, CHS, CVD-EDIC, NDR, NZ DCS and UKPDS risk scores. All risk scores exhibited modest discrimination (pooled c-statistics ranging from 0.63 to 0.68), with no individual risk score substantially outperforming the others.

Supplemental Figure 8A and 8B provide a histogram of the total number of adjusted covariates and number of adjusted CVD risk factors in each of the studies, respectively. Supplemental Figure 9 is a network figure representing the connections of the adjusted covariates in the 416 included studies.

### Sensitivity Analyses

The results of sensitivity analyses excluding studies with high risk of bias from meta-analyses of biomarkers, genetic risk score, and for risk scores where pooled analyses were possible, respectively, are shown in Supplemental Figures 10, 11, and 12.

### Synthesis

Table 2 provides a summary of findings of studies assessing the most promising biomarkers and genetic markers/scores for precision prognosis of CVD in T2D, along with our conclusions regarding their predictive utility and strength of evidence. In our synthesis of the evidence, we took into account the results from the sensitivity analyses described in the previous paragraph. The highest predictive utility was observed for NT-proBNP (high-evidence), TnT (moderate-evidence), TyG (high-evidence), and GRS-CHD (moderate-evidence). Prognostic factors with moderate predictive utility were CCTA (low-evidence), SPECT scintigraphy (low-evidence), and PWV (moderate-evidence). Prognostic factors with low predictive utility included CRP (moderate-evidence), CACS (low-evidence), Gal-3 (low-evidence), TnI (low-evidence), carotid plaque (low-evidence), and GDF-15 (low-evidence). Supplemental Figures 13A, 13B, and 13C provide the quality assessment for the included biomarker, genetic marker, and risk score studies, respectively.

**Table 2.** Conclusion and strength of the evidence

|  | A | B | C | D | E | F | G | H |
| --- | --- | --- | --- | --- | --- | --- | --- | --- |
| **Prognostic Biomarker** | **No. of studies with all 3 performance indicators satisfied** | **No. of pooled meta-analyses showing significant association** | **No. of pooled meta-analyses showing significant association (excluding high risk of bias)** | **Non-pooled analyses showed that $\geq 75\%$ of studies had significant association** | **Persistent association on sensitivity analysis for non-pooled analyses (excluding high risk of bias)** | **Consistency in definition of prognostic biomarker used in analysis** | **Conclusion (predictive utility)** | **Strength of Evidence** |
| NT-proBNP | 3 | 2/2 | 2/2 | Yes | Yes | Yes | High | High |
| CRP | 0 | 1/2 | 1/2 | No | No | No | Low | Moderate |
| TnT | 4 | 2/3 | 1/3 | Yes | Yes | No | High | Moderate |
| CACS | 0 | 0 | 0 | Yes | No | No | Low | Low |
| CCTA | 2 | 0 | 0 | Yes | No | No | Moderate | Low |
| SPECT | 1 | 0 | 0 | Yes | No | No | Moderate | Low |
| PWV | 0 | 1/1 | 1/1 | Yes | Yes | No | Moderate | Moderate |
| Gal-3 | 0 | 0 | 0 | Yes | Yes | No | Low | Low |
| TnI | 1 | 0 | 0 | No | No | No | Low | Low |
| Carotid plaque | 0 | 0 | 0 | Yes | Yes | No | Low | Low |
| GDF-15 | 0 | 0 | 0 | No | No | No | Low | Low |
| TyG | 1 | 1/1 | NA | Yes | No | Yes | High | Moderate |
| GRS-CHD | 1 | 1/1 | 1/1 | Yes | No | No | High | Moderate |

## Discussion

Our systematic review of prognostic markers for CVD in individuals with T2D has revealed several novel findings. First, among the numerous studies that investigated the prognostic significance of CVD risk markers, only a few have been consistently found to be significantly associated with cardiovascular risk. Namely, NT-proBNP, TnT, TyG, and GRS-CHD demonstrated the highest predictive utility, with NT-proBNP having the strongest evidence. However, most of the remaining markers have not been adequately tested or compared against established CVD risk factors. Finally, even though some markers have demonstrated the capability of predicting cardiovascular events beyond what current risk factor-based models can offer, their application in clinical practice remains limited, as there is inadequate evidence of their contemporary clinical utility.

During the search process, a considerable number of studies were found ineligible for inclusion in our systematic review. Available studies were primarily cross-sectional in design, and only a limited number of them focused specifically on individuals with T2D and examined the early utility of risk factors and biomarkers in predicting future cardiovascular events. A major limitation in many studies was inadequate adjustment for established CVD risk factors; and even if studies considered adjustments, only a small fraction evaluated clinical utility beyond the use of established risk factors. These findings emphasize the need for better-designed studies to improve our understanding of the prognostic value of markers for CVD in T2D.

Most studies included in the final analysis were conducted in people of European, East or South Asian ancestry, with the top-5 countries of recruitment being the United States, UK, China, Japan and Italy. African ancestry and countries were underrepresented. A skewed geographical distribution was also evident regarding countries of author affiliation, with the same top-5 countries dominating the volume of publications. Although the geographical and ancestral imbalance reported here for biomarker studies is less pronounced than what was recently reported for GWAS studies^448^, it highlights the pressing need to enhance data collection, biomarker discovery and validation, as well as the development of population-specific cardiovascular risk prediction models in underrepresented populations and ancestries to hopefully help reduce healthcare disparities^449^.

In our analyses, the novel biomarker emerging as the best predictor was NT-proBNP; indeed, it fulfilled all criteria of predictive and clinical utility with multiple studies showing improvement in all prediction performance indicators, with consistency of results across studies and meta-analyses. Notably, this biomarker had also been found to be useful as a prognostic marker for incident CVD in the general population^450^. Our findings suggest that NT-proBNP, beyond its established role in the diagnosis and management of patients with heart failure, might also be used as a marker to predict CVD. Another biomarker found in the general population to improve primary CVD risk prediction among asymptomatic middle-aged adults is high-sensitivity CRP (hs-CRP). In our review, CRP was found to have low predictive utility with moderate strength of evidence, which may be due to variability in cut-offs used for this marker, the relatively small numbers of studies, differential effects in diabetes, or less sensitive to detect low-grade vascular inflammation (compared with hs-CRP).

Despite numerous genetic studies probing the link between polymorphisms and cardiovascular outcomes in diabetes, few genetic markers have been consistently examined in longitudinal studies or reliably found to be associated with these outcomes. Only one study from the systematic review utilized a genome-wide association study (GWAS) approach, identifying the rs10911021 variant near *GLUL* to be associated with CV outcome in diabetes, at genome-wide significance. The variant at *GLUL* was subsequently confirmed in two independent studies^172^. A more recent GWAS conducted among Chinese patients with T2D identified a variant at *PDE1A* for CHD in T2D, which was not included in our systematic review as it fell beyond our study inclusion period^451^. Polygenic risk scores also appear to emerge as promising tools, and GRS constructed from variants associated with CHD in the general population seem helpful for cardiovascular risk stratification in diabetes^257^.

Based on these limited findings, it becomes clear that we need a greater number of adequately powered GWAS to identify genetic markers associated with CVD in T2D. Nevertheless, we found several examples of studies that evaluated the utility of applying polygenic risk scores, or genome-wide polygenic risk scores, derived from the general population, for CVD risk stratification in T2D. In general, these have fair performance and a similar ability to stratify as in patients without diabetes. Considering the significantly larger sample sizes in currently published meta-analyses of GWAS for CHD in the general population, this approach will probably be more fruitful for the integration of genetic markers into risk stratification of cardiovascular complications. In the limited studies that have evaluated the added benefit of polygenic risk scores above clinical markers, there is, in general, a modest but significant improvement in prediction. Whether polygenic risk scores will become viable options for future risk stratification would partly depend on the availability of these tools, and the cost-effectiveness of adding these measures into clinical practice.

Beyond individual prognostic markers, our review identified several studies that evaluated CVD risk prediction models. While the UKPDS risk engine (developed among subjects with newly diagnosed T2D the UK) and the Framingham risk equation (developed from the general population in the US) were the most widely studied, they do not perform well in contemporary studies of people with T2D. This suggests difficulties in applying certain risk models to current healthcare settings. Nevertheless, our literature review shows that clinical risk models are perhaps the “readiest” for implementation in clinical practice to improve risk stratification in diabetes. On external validation, newer risk scores generally achieved higher discrimination compared to UKPDS and FRS, with Fremantle Diabetes Study 2 (FDS-2) having the highest c-statistic of 0.81 (developed and validated in different populations in Australia). We found that risk models performed better when validated in cohorts similar to the derivation cohort, with c-statistics of 0.699 + 0.015 and 0.668 + 0.006 (95% CI) (P=0.018) for concordant and discordant studies, respectively.

In an era when electronic medical record (EMR)-based prediction models are being increasingly used, our results suggest that researchers should focus on the development of population-specific risk models that are intended to be deployed in the same population from which they were developed since the goal should be to achieve the highest predictive accuracy rather than to find a generic model that performs modestly well in all settings. Despite their potential utility and low implementation costs, we found a paucity of evidence showing integration of risk engine calculators into clinical practice. We are aware of several notable exceptions. For example, the Joint Asia Diabetes Evaluation (JADE) program has incorporated several risk prediction algorithms derived from Asian patients with diabetes into a web-based e-health portal, together with a graphical interface and decision support^452^, and has been evaluated in different clinical settings, including in randomized clinical trials^453–456^. Many EMR systems offer quick calculations of CVD risk using the American College of Cardiology/American Heart Association (ACC/AHA) Pooled Cohort Equations based on inputs available in the patient’s record, and we recommend that future risk scores found to have high predictive accuracy be made easily accessible to clinicians within their EMR workflow.

Given the limitations and gaps that emerged from this review, we recommend that future studies follow several guidelines to improve the quality and impact of studies on precision prognostics in diabetes. First, studies attempting to identify a risk marker should be conducted in prospective or longitudinal cohorts or trials, to provide more robust and reliable data. Second, studies should have sufficient sample size and duration of follow-up (at least 3 years for primary CVD events and at least 1 year for secondary CVD events) to ensure adequate statistical power. Third, studies must adjust for a minimal set of established clinical cardiovascular risk factors, to ensure that known risk factors do not confound any observed associations. Finally, studies must attempt to explore the added utility of biomarkers by comparing against prediction using established risk factors or models, or available risk engines for cardiovascular events. This would include evaluation of the change in c-statistics after adding risk markers/biomarkers of interest but also consider including additional metrics such as NRI and IDI. We believe that if journals make these requirements mandatory when evaluating such studies, it will help ensure that research funders are made aware and future studies are best suited for informing advances in this area especially in resource-limited countries. As in any other research field, harmonization of protocols, methods, and analysis pipelines should be encouraged to allow comparisons across studies and for clinical translation.

There are several unique strengths of this work. To our knowledge, this represents one of the most comprehensive overviews of the current status of knowledge about risk stratification of cardiovascular outcomes in T2D. We included studies from 1990 onwards, to capture some of the older studies, as well as more contemporary studies. Our inclusion of “biomarkers” in the broadest term allowed us to provide an objective overview of the different approaches currently being explored for better risk stratification. Limiting the analyses to studies using longitudinal cohorts allowed us to focus on studies that would inform prognostication. Limiting analyses to “hard” cardiovascular endpoints, rather than also including surrogate endpoints such as carotid intima-medial thickness, allowed us to focus on endpoints that would be of greatest clinical relevance. However, while this approach allows us to maximize the translational approach of our analyses, future studies focused on identification of biomarkers associated with early disease-informative endpoints (i.e. subclinical markers of atherosclerosis or minor cardiovascular disease) might identify different novel biomarkers for early-stage cardiovascular complications.

Our study does have limitations. We had to omit a considerable number of cross-sectional studies due to the extensive scope of the systematic review and the explained focus on longitudinal studies. We included only English language publications. Our search terms, potentially more sensitive towards detecting studies on clinical risk factors and biomarkers than genetic factors, may have led to fewer genetic studies being identified. However, we managed to supplement this by reintegrating some missing articles using the identified literature and the investigators’ expertise.

In conclusion, our systematic review on prognostic markers for cardiovascular endpoints in T2D identified several novel findings and some important knowledge gaps. We found that NT-proBNP, TnT, TyG, and GRS-CHD had high predictive utility beyond traditional CVD risk factors, with the highest strength of evidence for NT-proBNP. Among genetic markers, there was only sufficient evidence for the polygenic risk score for CHD, and among risk scores, predictive utility was modest on external validation. Given the relatively low number of studies analyzing these novel prognostic factors using a rigorous approach, these findings support the need for future studies testing these markers with convincing demonstration of incremental predictive utility. NT-proBNP appears to be the only biomarker ready to be tested prospectively to evaluate its utility in modifying clinical practice for prediction of CVD risk.

## Data Availability

All data produced in the present work are contained in the manuscript

https://hugofitipaldi.shinyapps.io/T2D_prognostic/

## Acknowledgements

A.A., M.D-P., H.F., M.F.G. acknowledge support from the Swedish Heart-Lung Foundation (20190470), Swedish Research Council (EXODIAB, 2009-1039; 2018-02837), Swedish Foundation for Strategic Research (LUDC-IRC, 15-0067), EU H2020-JTI-lMl2-2015-05 (Grant agreement number 115974 – BEAt-DKD) to M.F.G. L-L.L. acknowledge UK Medical Research Council Population and Systems Medicine Board (IF048-2022). M.L.M. is supported by Italian Ministry of Health Grant “Ricerca Finalizzata 2019” – GR-2019-12369702. C.H.T.T., C.H, and R.C.W.M. acknowledge support from the Research Grants Council of the Hong Kong Special Administrative Region (CU R4012-18), the Croucher Foundation Senior Medical Research Fellowship, University Grants Committee Research Grants Matching Scheme and Research Committee Postdoctoral Fellowship Scheme of the Chinese University of Hong Kong. F.F.C. acknowledge the Second Affiliated Hospital of Chongqing Medical University (No. 2022cffkyqdj). T.C. is an international training fellow supported by the Wellcome Trust grant (214205/Z/18/Z). R.W.K. was funded by a Novo Nordisk Foundation (NNF18OC0031650) postdoctoral fellowship. G.Y. and R.C.W.M. acknowledges support from the Provost’s Scheme for PhD scholarship from the Chinese University of Hong Kong. Y.Z. acknowledges a Postgraduate Studentship and Vice-Chancellor’s PhD scholarship from the Chinese University of Hong Kong. The authors wish to acknowledge the support of librarians from Lund University, Maria Björklund and Krister Aronsson for their expert support with the literature search. N.M. is supported by grants from the National Institute of Diabetes and Digestive and Kidney Diseases (R01DK125780, R01DK134955).

## Contributions of authors

Drs. Gomez, Ma, and Mathioudakis had full access to all the data in the study and take full responsibility for the integrity of the data and the accuracy of the data analysis.

*Concept and design:* R.C.W.M., N.M., M.F.G., A.A.M., A.A., L-L.L., R.W.K., S.C.T., C.H.T., M.D-P., F.C., S.S.

*Acquisition, analysis, or interpretation of data:* R.C.W.M., N.M., A.A., M.F.G., M.D-P., C.H.T., R.W.K., F.C., S.C.T., L-L.L., S.S., A.A.M., T.C., M.P., M.L.M., H.F., S.K., G.Y., Y.Z., C.H.

*Drafting of the manuscript:* R.C.W.M., N.M., M.F.G., A.A., M.L.M., L-L.L., C.H.T., H.F., S.K.

*Critical revision of the manuscript for important intellectual content:* R.C.W.M., N.M., M.F.G., A.A., L-L.L., M.L.M., C.H.T., F.C., T.C., M.D-P., H.F., C.H., S.K., S.S., R.W.K., A.A.M., S.C.T., G.Y., Y.Z., M.P., D.S., R.J.dS., D.K.T.

*Statistical analysis:* C.H.T., M.L.M., N.M.

A.A., L-L.L., M.L.M., C.H.T. contributed equally to this work and are considered co-first authors.

M.F.G., R.C.W.M., and N.M. supervised all aspects of this work and are considered co-senior authors.

## References

1 Rawshani, A. et al. Mortality and Cardiovascular Disease in Type 1 and Type 2 Diabetes. New England Journal of Medicine 376, 1407–1418 (2017). https://doi.org/10.1056/NEJMoa1608664

2 Pearson-Stuttard, J. et al. Trends in predominant causes of death in individuals with and without diabetes in England from 2001 to 2018: an epidemiological analysis of linked primary care records. Lancet Diabetes Endocrinol 9, 165–173 (2021). https://doi.org/10.1016/s2213-8587(20)30431-9

3 Nathan, D. M. et al. Glycemia Reduction in Type 2 Diabetes - Glycemic Outcomes. N Engl J Med 387, 1063–1074 (2022). https://doi.org/10.1056/NEJMoa2200433

4 IDF Diabetes Atlas. Diabetes around the world in 2021, https://diabetesatlas.org/ (2021).

5 van Dieren, S. et al. External validation of the UK Prospective Diabetes Study (UKPDS) risk engine in patients with type 2 diabetes. Diabetologia 54, 264–270 (2011). https://doi.org/10.1007/s00125-010-1960-0

6 Szymonifka, J. et al. Cardiovascular disease risk prediction for people with type 2 diabetes in a population-based cohort and in electronic health record data. JAMIA Open 3, 583–592 (2020). https://doi.org/10.1093/jamiaopen/ooaa059

7 Ho, J. C. et al. Evaluation of available risk scores to predict multiple cardiovascular complications for patients with type 2 diabetes mellitus using electronic health records. Computer Methods and Programs in Biomedicine Update 3, 100087 (2023). https://doi.org/https://doi.org/10.1016/j.cmpbup.2022.100087

8 Chung, W. K. et al. Precision Medicine in Diabetes: A Consensus Report From the American Diabetes Association (ADA) and the European Association for the Study of Diabetes (EASD). Diabetes Care 43, 1617–1635 (2020). https://doi.org/10.2337/dci20-0022

9 Nolan, J. J. et al. ADA/EASD Precision Medicine in Diabetes Initiative: An International Perspective and Future Vision for Precision Medicine in Diabetes. Diabetes Care 45, 261–266 (2022). https://doi.org/10.2337/dc21-2216

10 Tobias, e. a. Precision Medicine in Diabetes: Second international consensus report. Nat Med **Under review** (2023).

11 Damen, J. A. et al. Prediction models for cardiovascular disease risk in the general population: systematic review. BMJ 353, i2416 (2016). https://doi.org/10.1136/bmj.i2416

12 Zhiting, G. et al. Cardiovascular disease risk prediction models in the Chinese population-a systematic review and meta-analysis. BMC Public Health 22, 1608 (2022). https://doi.org/10.1186/s12889-022-13995-z

13 Damen, J. A. et al. Performance of the Framingham risk models and pooled cohort equations for predicting 10-year risk of cardiovascular disease: a systematic review and meta-analysis. BMC Med 17, 109 (2019). https://doi.org/10.1186/s12916-019-1340-7

14 Romero-Cabrera, J. L., Ankeny, J., Fernández-Montero, A., Kales, S. N. & Smith, D. L. A Systematic Review and Meta-Analysis of Advanced Biomarkers for Predicting Incident Cardiovascular Disease among Asymptomatic Middle-Aged Adults. Int J Mol Sci 23 (2022). https://doi.org/10.3390/ijms232113540

15 van Holten, T. C. et al. Circulating biomarkers for predicting cardiovascular disease risk; a systematic review and comprehensive overview of meta-analyses. PLoS One 8, e62080 (2013). https://doi.org/10.1371/journal.pone.0062080

16 Guasti, L. et al. TMAO as a biomarker of cardiovascular events: a systematic review and meta-analysis. Intern Emerg Med 16, 201–207 (2021). https://doi.org/10.1007/s11739-020-02470-5

17 Rienks, J., Barbaresko, J. & Nöthlings, U. Association of Polyphenol Biomarkers with Cardiovascular Disease and Mortality Risk: A Systematic Review and Meta-Analysis of Observational Studies. Nutrients 9 (2017). https://doi.org/10.3390/nu9040415

18 Gohel, V., Jones, J. A. & Wehler, C. J. Salivary biomarkers and cardiovascular disease: a systematic review. Clin Chem Lab Med 56, 1432–1442 (2018). https://doi.org/10.1515/cclm-2017-1018

19 Kanbay, M. et al. Sclerostin, cardiovascular disease and mortality: a systematic review and meta-analysis. Int Urol Nephrol 48, 2029–2042 (2016). https://doi.org/10.1007/s11255-016-1387-8

20 Heianza, Y., Ma, W., Manson, J. E., Rexrode, K. M. & Qi, L. Gut Microbiota Metabolites and Risk of Major Adverse Cardiovascular Disease Events and Death: A Systematic Review and Meta-Analysis of Prospective Studies. J Am Heart Assoc 6 (2017). https://doi.org/10.1161/JAHA.116.004947

21 Wallace, T. C., Slavin, M. & Frankenfeld, C. L. Systematic Review of Anthocyanins and Markers of Cardiovascular Disease. Nutrients 8 (2016). https://doi.org/10.3390/nu8010032

22 Emadian, A., Andrews, R. C., England, C. Y., Wallace, V. & Thompson, J. L. The effect of macronutrients on glycaemic control: a systematic review of dietary randomised controlled trials in overweight and obese adults with type 2 diabetes in which there was no difference in weight loss between treatment groups. Br J Nutr 114, 1656–1666 (2015). https://doi.org/10.1017/S0007114515003475

23 Yun, H., Noh, N. I. & Lee, E. Y. Genetic risk scores used in cardiovascular disease prediction models: a systematic review. Rev Cardiovasc Med 23, 8 (2022). https://doi.org/10.31083/j.rcm2301008

24 Jeong, E. G. et al. Depth and combined infection is important predictor of lower extremity amputations in hospitalized diabetic foot ulcer patients. Korean J Intern Med 33, 952–960 (2018). https://doi.org/10.3904/kjim.2016.165

25 Lin, J. S. et al. Nontraditional Risk Factors in Cardiovascular Disease Risk Assessment: Updated Evidence Report and Systematic Review for the US Preventive Services Task Force. Jama 320, 281–297 (2018). https://doi.org/10.1001/jama.2018.4242

26 Hlatky, M. A. et al. Criteria for evaluation of novel markers of cardiovascular risk: a scientific statement from the American Heart Association. Circulation 119, 2408–2416 (2009). https://doi.org/10.1161/circulationaha.109.192278

27 Page, M. J. et al. The PRISMA 2020 statement: an updated guideline for reporting systematic reviews. BMJ 372, n71 (2021). https://doi.org/10.1136/bmj.n71

28 Wells GA, S. B., O’Connell D, Peterson J, Welch V, Losos M, & Tugwell P. The Newcastle-Ottawa Scale (NOS) for assessing the quality of nonrandomised studies in meta-analyses, https://www.ohri.ca/programs/clinical_epidemiology/oxford.asp (

29 Viechtbauer, W. Conducting Meta-Analyses in R with the metafor Package. Journal of Statistical Software 36, 1–48 (2010). https://doi.org/10.18637/jss.v036.i03

30 Owens, D. K. et al. AHRQ series paper 5: grading the strength of a body of evidence when comparing medical interventions--agency for healthcare research and quality and the effective health-care program. J Clin Epidemiol 63, 513–523 (2010). https://doi.org/10.1016/j.jclinepi.2009.03.009

31 Aromataris E, M. Z. E. JBI Manual for Evidence Synthesis. JBI, https://doi.org/10.46658/JBIMES-20-01 (2020).

32 Aboyans, V. et al. The prognosis of diabetic patients with high ankle-brachial index depends on the coexistence of occlusive peripheral artery disease. J Vasc Surg 53, 984–991 (2011). https://doi.org/10.1016/j.jvs.2010.10.054

33 Abu-Lebdeh, H. S., Hodge, D. O. & Nguyen, T. T. Predictors of macrovascular disease in patients with type 2 diabetes mellitus. Mayo Clin Proc 76, 707–712 (2001). https://doi.org/10.4065/76.7.707

34 Afarideh, M. et al. Complex association of serum alanine aminotransferase with the risk of future cardiovascular disease in type 2 diabetes. Atherosclerosis 254, 42–51 (2016). https://doi.org/10.1016/j.atherosclerosis.2016.09.009

35 Afsharian, S. et al. Risk factors for cardiovascular disease and mortality events in adults with type 2 diabeteslJ-lJa 10-year follow-up: Tehran Lipid and Glucose Study. Diabetes Metab Res Rev 32, 596–606 (2016). https://doi.org/10.1002/dmrr.2776

36 Alele, J. D., Luttrell, L. M., Hollis, B. W., Luttrell, D. K. & Hunt, K. J. Relationship between vitamin D status and incidence of vascular events in the Veterans Affairs Diabetes Trial. Atherosclerosis 228, 502–507 (2013). https://doi.org/10.1016/j.atherosclerosis.2013.03.024

37 Alkhalaf, A. et al. Sex specific association between carnosinase gene CNDP1 and cardiovascular mortality in patients with type 2 diabetes (ZODIAC-22). J Nephrol 28, 201–207 (2015). https://doi.org/10.1007/s40620-014-0096-6

38 Anand, D. V., Lahiri, A., Lim, E., Hopkins, D. & Corder, R. The relationship between plasma osteoprotegerin levels and coronary artery calcification in uncomplicated type 2 diabetic subjects. J Am Coll Cardiol 47, 1850–1857 (2006). https://doi.org/10.1016/j.jacc.2005.12.054

39 Anand, D. V. et al. Risk stratification in uncomplicated type 2 diabetes: prospective evaluation of the combined use of coronary artery calcium imaging and selective myocardial perfusion scintigraphy. Eur Heart J 27, 713–721 (2006). https://doi.org/10.1093/eurheartj/ehi808

40 Anavekar, N. S. et al. Predictors of cardiovascular events in patients with type 2 diabetic nephropathy and hypertension: a case for albuminuria. Kidney Int Suppl, S50–55 (2004). https://doi.org/10.1111/j.1523-1755.2004.09213.x

41 Angiolillo, D. J. et al. Impact of platelet reactivity on cardiovascular outcomes in patients with type 2 diabetes mellitus and coronary artery disease. J Am Coll Cardiol 50, 1541–1547 (2007). https://doi.org/10.1016/j.jacc.2007.05.049

42 Anyanwagu, U., Donnelly, R. & Idris, I. Albuminuria Regression and All-Cause Mortality among Insulin-Treated Patients with Type 2 Diabetes: Analysis of a Large UK Primary Care Cohort. Am J Nephrol 49, 146–155 (2019). https://doi.org/10.1159/000496276

43 Apperloo, E. M., Pena, M. J., de Zeeuw, D., Denig, P. & Heerspink, H. J. L. Individual variability in response to renin angiotensin aldosterone system inhibition predicts cardiovascular outcome in patients with type 2 diabetes: A primary care cohort study. Diabetes Obes Metab 20, 1377–1383 (2018). https://doi.org/10.1111/dom.13226

44 Araki, S. et al. Predictive effects of urinary liver-type fatty acid-binding protein for deteriorating renal function and incidence of cardiovascular disease in type 2 diabetic patients without advanced nephropathy. Diabetes Care 36, 1248–1253 (2013). https://doi.org/10.2337/dc12-1298

45 Avogaro, A. et al. Incidence of coronary heart disease in type 2 diabetic men and women: impact of microvascular complications, treatment, and geographic location. Diabetes Care 30, 1241–1247 (2007). https://doi.org/10.2337/dc06-2558

46 Azab, B., Chainani, V., Shah, N. & McGinn, J. T. Neutrophil-lymphocyte ratio as a predictor of major adverse cardiac events among diabetic population: a 4-year follow-up study. Angiology 64, 456–465 (2013). https://doi.org/10.1177/0003319712455216

47 Azevedo, M. J. et al. Value of diagnostic tools for myocardial ischemia used in routine clinical practice to predict cardiac events in patients with type 2 diabetes mellitus: a prospective study. Arq Bras Endocrinol Metabol 50, 46–52 (2006). https://doi.org/10.1590/s0004-27302006000100007

48 Bacci, S. et al. The ENPP1 Q121 variant predicts major cardiovascular events in high-risk individuals: evidence for interaction with obesity in diabetic patients. Diabetes 60, 1000–1007 (2011). https://doi.org/10.2337/db10-1300

49 Backhaus, S. J. et al. Cardiac Magnetic Resonance Myocardial Feature Tracking for Optimized Risk Assessment After Acute Myocardial Infarction in Patients With Type 2 Diabetes. Diabetes 69, 1540–1548 (2020). https://doi.org/10.2337/db20-0001

50 Basu, S. et al. Validation of Risk Equations for Complications of Type 2 Diabetes (RECODe) Using Individual Participant Data From Diverse Longitudinal Cohorts in the U.S. Diabetes Care 41, 586–595 (2018). https://doi.org/10.2337/dc17-2002

51 Basu, S., Sussman, J. B., Berkowitz, S. A., Hayward, R. A. & Yudkin, J. S. Development and validation of Risk Equations for Complications Of type 2 Diabetes (RECODe) using individual participant data from randomised trials. Lancet Diabetes Endocrinol 5, 788–798 (2017). https://doi.org/10.1016/s2213-8587(17)30221-8

52 Bates, R. E. et al. Impact of Stress Testing for Coronary Artery Disease Screening in Asymptomatic Patients With Diabetes Mellitus: A Community-Based Study in Olmsted County, Minnesota. Mayo Clin Proc 91, 1535–1544 (2016). https://doi.org/10.1016/j.mayocp.2016.07.013

53 Beaney, K. E. et al. Variant rs10911021 that associates with coronary heart disease in type 2 diabetes, is associated with lower concentrations of circulating HDL cholesterol and large HDL particles but not with amino acids. Cardiovasc Diabetol 15, 115 (2016). https://doi.org/10.1186/s12933-016-0435-0

54 Beilin, J., Stanton, K. G., McCann, V. J., Knuiman, M. W. & Divitini, M. L. Microalbuminuria in type 2 diabetes: an independent predictor of cardiovascular mortality. Aust N Z J Med 26, 519–525 (1996). https://doi.org/10.1111/j.1445-5994.1996.tb00598.x

55 Bell, K. J. L. et al. Prognostic impact of systolic blood pressure variability in people with diabetes. PLoS One 13, e0194084 (2018). https://doi.org/10.1371/journal.pone.0194084

56 Berkelmans, G. F. N. et al. Prediction of individual life-years gained without cardiovascular events from lipid, blood pressure, glucose, and aspirin treatment based on data of more than 500 000 patients with Type 2 diabetes mellitus. Eur Heart J 40, 2899–2906 (2019). https://doi.org/10.1093/eurheartj/ehy839

57 Bernard, S. et al. Relation between XbA1 apolipoprotein B gene polymorphism and cardiovascular risk in a type 2 diabetic cohort. Atherosclerosis 175, 177–181 (2004). https://doi.org/10.1016/j.atherosclerosis.2004.03.017

58 Bernard, S. et al. Incremental predictive value of carotid ultrasonography in the assessment of coronary risk in a cohort of asymptomatic type 2 diabetic subjects. Diabetes Care 28, 1158–1162 (2005). https://doi.org/10.2337/diacare.28.5.1158

59 Bianco, H. T. et al. Relevance of target-organ lesions as predictors of mortality in patients with diabetes mellitus. Arq Bras Cardiol 103, 272–281 (2014). https://doi.org/10.5935/abc.20140112

60 Biscetti, F. et al. Association between omentin-1 and major cardiovascular events after lower extremity endovascular revascularization in diabetic patients: a prospective cohort study. Cardiovasc Diabetol 19, 170 (2020). https://doi.org/10.1186/s12933-020-01151-z

61 Bonito, B., Silva, A. P., Rato, F., Santos, N. & Neves, P. L. Resistin as a predictor of cardiovascular hospital admissions and renal deterioration in diabetic patients with chronic kidney disease. J Diabetes Complications 33, 107422 (2019). https://doi.org/10.1016/j.jdiacomp.2019.107422

62 Bosevski, M., Borozanov, V., Tosev, S. & Georgievska-Ismail, L. Is assessment of peripheral endothelial dysfunction useful tool for risk stratification of type 2 diabetic patients with manifested coronary artery disease? Int J Cardiol 131, 290–292 (2009). https://doi.org/10.1016/j.ijcard.2007.08.011

63 Bouchi, R. et al. Fluctuations in HbA1c are associated with a higher incidence of cardiovascular disease in Japanese patients with type 2 diabetes. Journal of Diabetes Investigation 3, 148–155 (2012). https://doi.org/10.1111/j.2040-1124.2011.00155.x

64 Brownrigg, J. R. et al. Microvascular disease and risk of cardiovascular events among individuals with type 2 diabetes: a population-level cohort study. Lancet Diabetes Endocrinol 4, 588–597 (2016). https://doi.org/10.1016/s2213-8587(16)30057-2

65 Bruce, D. G., Davis, W. A., Starkstein, S. E. & Davis, T. M. A prospective study of depression and mortality in patients with type 2 diabetes: the Fremantle Diabetes Study. Diabetologia 48, 2532–2539 (2005). https://doi.org/10.1007/s00125-005-0024-3

66 Bruno, G., Barutta, F., Landi, A., Cavallo Perin, P. & Gruden, G. NT-proBNP linking low-moderately impaired renal function and cardiovascular mortality in diabetic patients: the population-based Casale Monferrato Study. PLoS One 9, e114855 (2014). https://doi.org/10.1371/journal.pone.0114855

67 Bruno, G. et al. N-terminal probrain natriuretic peptide is a stronger predictor of cardiovascular mortality than C-reactive protein and albumin excretion rate in elderly patients with type 2 diabetes: the Casale Monferrato population-based study. Diabetes Care 36, 2677–2682 (2013). https://doi.org/10.2337/dc13-0353

68 Bruno, G. et al. Fibrinogen and AER are major independent predictors of 11-year cardiovascular mortality in type 2 diabetes: the Casale Monferrato Study. Diabetologia 48, 427–434 (2005). https://doi.org/10.1007/s00125-004-1667-1

69 Bruno, G. et al. Metabolic syndrome as a predictor of all-cause and cardiovascular mortality in type 2 diabetes: the Casale Monferrato Study. Diabetes Care 27, 2689–2694 (2004). https://doi.org/10.2337/diacare.27.11.2689

70 Burgess, D. C. et al. Incidence and predictors of silent myocardial infarction in type 2 diabetes and the effect of fenofibrate: an analysis from the Fenofibrate Intervention and Event Lowering in Diabetes (FIELD) study. Eur Heart J 31, 92–99 (2010). https://doi.org/10.1093/eurheartj/ehp377

71 Busch, M. et al. The advanced glycation end product N(epsilon)-carboxymethyllysine is not a predictor of cardiovascular events and renal outcomes in patients with type 2 diabetic kidney disease and hypertension. Am J Kidney Dis 48, 571–579 (2006). https://doi.org/10.1053/j.ajkd.2006.07.009

72 Böger, C. A. et al. RANTES gene polymorphisms predict all-cause and cardiac mortality in type 2 diabetes mellitus hemodialysis patients. Atherosclerosis 183, 121–129 (2005). https://doi.org/10.1016/j.atherosclerosis.2005.03.006

73 Cardona, A. et al. Trimethylamine N-oxide and incident atherosclerotic events in high-risk individuals with diabetes: an ACCORD trial post hoc analysis. BMJ Open Diabetes Res Care 7, e000718 (2019). https://doi.org/10.1136/bmjdrc-2019-000718

74 Cardoso, C. R., Ferreira, M. T., Leite, N. C. & Salles, G. F. Prognostic impact of aortic stiffness in high-risk type 2 diabetic patients: the Rio deJaneiro Type 2 Diabetes Cohort Study. Diabetes Care 36, 3772–3778 (2013). https://doi.org/10.2337/dc13-0506

75 Cardoso, C. R., Leite, N. C. & Salles, G. F. Prognostic Importance of C-Reactive Protein in High Cardiovascular Risk Patients With Type 2 Diabetes Mellitus: The Rio de Janeiro Type 2 Diabetes Cohort Study. J Am Heart Assoc 5 (2016). https://doi.org/10.1161/jaha.116.004554

76 Cardoso, C. R., Salles, G. F. & Deccache, W. Prognostic value of QT interval parameters in type 2 diabetes mellitus: results of a long-term follow-up prospective study. J Diabetes Complications 17, 169–178 (2003). https://doi.org/10.1016/s1056-8727(02)00206-4

77 Cardoso, C. R. L., Leite, N. C., Moram, C. B. M. & Salles, G. F. Long-term visit-to-visit glycemic variability as predictor of micro-and macrovascular complications in patients with type 2 diabetes: The Rio de Janeiro Type 2 Diabetes Cohort Study. Cardiovasc Diabetol 17, 33 (2018). https://doi.org/10.1186/s12933-018-0677-0

78 Cardoso, C. R. L., Leite, N. C. & Salles, G. F. Prognostic importance of visit-to-visit blood pressure variability for micro-and macrovascular outcomes in patients with type 2 diabetes: The Rio de Janeiro Type 2 Diabetes Cohort Study. Cardiovasc Diabetol 19, 50 (2020). https://doi.org/10.1186/s12933-020-01030-7

79 Cardoso, C. R. L., Salles, G. C., Leite, N. C. & Salles, G. F. Prognostic impact of carotid intima-media thickness and carotid plaques on the development of micro-and macrovascular complications in individuals with type 2 diabetes: the Rio de Janeiro type 2 diabetes cohort study. Cardiovasc Diabetol 18, 2 (2019). https://doi.org/10.1186/s12933-019-0809-1

80 Carlsson, A. C. et al. Growth differentiation factor 15 (GDF-15) is a potential biomarker of both diabetic kidney disease and future cardiovascular events in cohorts of individuals with type 2 diabetes: a proteomics approach. Ups J Med Sci 125, 37–43 (2020). https://doi.org/10.1080/03009734.2019.1696430

81 Carlsson, A. C. et al. Association of soluble tumor necrosis factor receptors 1 and 2 with nephropathy, cardiovascular events, and total mortality in type 2 diabetes. Cardiovasc Diabetol 15, 40 (2016). https://doi.org/10.1186/s12933-016-0359-8

82 Carnethon, M. R. et al. Association of weight status with mortality in adults with incident diabetes. Jama 308, 581–590 (2012). https://doi.org/10.1001/jama.2012.9282

83 Casiglia, E. et al. Cardiovascular mortality in non-insulin-dependent diabetes mellitus. A controlled study among 683 diabetics and 683 age- and sex-matched normal subjects. Eur J Epidemiol 16, 677–684 (2000). https://doi.org/10.1023/a:1007673123716

84 Cavalot, F. et al. Postprandial blood glucose predicts cardiovascular events and all-cause mortality in type 2 diabetes in a 14-year follow-up: lessons from the San Luigi Gonzaga Diabetes Study. Diabetes Care 34, 2237–2243 (2011). https://doi.org/10.2337/dc10-2414

85 Cavalot, F. et al. Postprandial blood glucose is a stronger predictor of cardiovascular events than fasting blood glucose in type 2 diabetes mellitus, particularly in women: lessons from the San Luigi Gonzaga Diabetes Study. J Clin Endocrinol Metab 91, 813–819 (2006). https://doi.org/10.1210/jc.2005-1005

86 Cea Soriano, L., Johansson, S., Stefansson, B. & Rodríguez, L. A. Cardiovascular events and all-cause mortality in a cohort of 57,946 patients with type 2 diabetes: associations with renal function and cardiovascular risk factors. Cardiovasc Diabetol 14, 38 (2015). https://doi.org/10.1186/s12933-015-0204-5

87 Cederholm, J. et al. Risk prediction of cardiovascular disease in type 2 diabetes: a risk equation from the Swedish National Diabetes Register. Diabetes Care 31, 2038–2043 (2008). https://doi.org/10.2337/dc08-0662

88 Celis-Morales, C. A. et al. Associations Between Diabetes and Both Cardiovascular Disease and All-Cause Mortality Are Modified by Grip Strength: Evidence From UK Biobank, a Prospective Population-Based Cohort Study. Diabetes Care 40, 1710–1718 (2017). https://doi.org/10.2337/dc17-0921

89 Ceriello, A. et al. Empagliflozin reduced long-term HbA1c variability and cardiovascular death: insights from the EMPA-REG OUTCOME trial. Cardiovasc Diabetol 19, 176 (2020). https://doi.org/10.1186/s12933-020-01147-9

90 Cha, S. A. et al. Time- and frequency-domain measures of heart rate variability predict cardiovascular outcome in patients with type 2 diabetes. Diabetes Res Clin Pract 143, 159–169 (2018). https://doi.org/10.1016/j.diabres.2018.07.001

91 Cha, S. A. et al. Diabetic Cardiovascular Autonomic Neuropathy Predicts Recurrent Cardiovascular Diseases in Patients with Type 2 Diabetes. PLoS One 11, e0164807 (2016). https://doi.org/10.1371/journal.pone.0164807

92 Chacko, K. M. et al. Heart rate recovery predicts mortality and cardiovascular events in patients with type 2 diabetes. Med Sci Sports Exerc 40, 288–295 (2008). https://doi.org/10.1249/mss.0b013e31815c4844

93 Chan, J. C. et al. Premature mortality and comorbidities in young-onset diabetes: a 7-year prospective analysis. Am J Med 127, 616–624 (2014). https://doi.org/10.1016/j.amjmed.2014.03.018

94 Chang, C. W. et al. The First Harmonic of Radial Pulse as an Early Predictor of Silent Coronary Artery Disease and Adverse Cardiac Events in Type 2 Diabetic Patients. Cardiology Research and Practice 2018 (2018). https://doi.org/10.1155/2018/5128626

95 Chang, L. H. et al. The Ankle Brachial Index Exhibits Better Association of Cardiovascular Prognosis Than Non-High-Density Lipoprotein Cholesterol in Type 2 Diabetes. Am J Med Sci 351, 492–498 (2016). https://doi.org/10.1016/j.amjms.2016.02.035

96 Chang, L. H. et al. UPSTROKE TIME PER CARDIAC CYCLE IS ASSOCIATED WITH CARDIOVASCULAR PROGNOSIS IN TYPE 2 DIABETES. Endocr Pract 25, 1109–1116 (2019). https://doi.org/10.4158/ep-2019-0078

97 Charlton-Menys, V. et al. Apolipoproteins, cardiovascular risk and statin response in type 2 diabetes: the Collaborative Atorvastatin Diabetes Study (CARDS). Diabetologia 52, 218–225 (2009). https://doi.org/10.1007/s00125-008-1176-8

98 Chen, H. S. et al. Subclinical hypothyroidism is a risk factor for nephropathy and cardiovascular diseases in Type 2 diabetic patients. Diabet Med 24, 1336–1344 (2007). https://doi.org/10.1111/j.1464-5491.2007.02270.x

99 Chen, S. et al. The long-term effectiveness of metabolic control on cardiovascular disease in patients with diabetes in a real-world health care setting - A prospective diabetes management study. Prim Care Diabetes 14, 274–281 (2020). https://doi.org/10.1016/j.pcd.2019.09.006

100 Cheng, F. et al. Shortened relative leukocyte telomere length is associated with prevalent and incident cardiovascular complications in type 2 diabetes: Analysis from the Hong Kong Diabetes Register. Diabetes Care 43, 2257–2265 (2020). https://doi.org/10.2337/dc20-0028

101 Christensen, P. K. et al. QTc interval length and QT dispersion as predictors of mortality in patients with non-insulin-dependent diabetes. Scand J Clin Lab Invest 60, 323–332 (2000). https://doi.org/10.1080/003655100750046486

102 Christiansen, M. S., Hommel, E., Magid, E. & Feldt-Rasmussen, B. Orosomucoid in urine predicts cardiovascular and over-all mortality in patients with Type II diabetes. Diabetologia 45, 115–120 (2002). https://doi.org/10.1007/s125-002-8251-3

103 Christiansen, M. S., Hommel, E., Magid, E. & Feldt-Rasmussen, B. Orosomucoid in urine is a powerful predictor of cardiovascular mortality in normoalbuminuric patients with type 2 diabetes at five years of follow-up. Diabetologia 48, 386–393 (2005). https://doi.org/10.1007/s00125-004-1630-1

104 Church, E. et al. Relationship between estimated glomerular filtration rate and incident cardiovascular disease in an ethnically diverse primary care cohort. N Z Med J 132, 11–26 (2019).

105 Chyun, D. A. et al. Autonomic dysfunction independently predicts poor cardiovascular outcomes in asymptomatic individuals with type 2 diabetes in the DIAD study. SAGE Open Medicine 3 (2015). https://doi.org/10.1177/2050312114568476

106 Cioffi, G. et al. Usefulness of subclinical left ventricular midwall dysfunction to predict cardiovascular mortality in patients with type 2 diabetes mellitus. Am J Cardiol 113, 1409–1414 (2014). https://doi.org/10.1016/j.amjcard.2014.01.415

107 Clarke, P. M. et al. A model to estimate the lifetime health outcomes of patients with type 2 diabetes: the United Kingdom Prospective Diabetes Study (UKPDS) Outcomes Model (UKPDS no. 68). Diabetologia 47, 1747-1759 (2004). https://doi.org/10.1007/s00125-004-1527-z

108 Clarke, P. M. et al. Using the EQ-5D index score as a predictor of outcomes in patients with type 2 diabetes. Med Care 47, 61–68 (2009). https://doi.org/10.1097/MLR.0b013e3181844855

109 Cockcroft, J. R. et al. Pulse pressure predicts cardiovascular risk in patients with type 2 diabetes mellitus. Am J Hypertens 18, 1463–1467; discussion 1468 (2005). https://doi.org/10.1016/j.amjhyper.2005.05.009

110 Colombo, M. et al. Apolipoprotein CIII and N-terminal prohormone b-type natriuretic peptide as independent predictors for cardiovascular disease in type 2 diabetes. Atherosclerosis 274, 182–190 (2018). https://doi.org/10.1016/j.atherosclerosis.2018.05.014

111 Cortigiani, L. et al. Prognostic meaning of coronary microvascular disease in type 2 diabetes mellitus: a transthoracic Doppler echocardiographic study. J Am Soc Echocardiogr 27, 742–748 (2014). https://doi.org/10.1016/j.echo.2014.02.010

112 Cosson, E. et al. Cardiovascular risk prediction is improved by adding asymptomatic coronary status to routine risk assessment in type 2 diabetic patients. Diabetes Care 34, 2101–2107 (2011). https://doi.org/10.2337/dc11-0480

113 Cournot, M. et al. Circulating Concentrations of Redox Biomarkers Do Not Improve the Prediction of Adverse Cardiovascular Events in Patients With Type 2 Diabetes Mellitus. J Am Heart Assoc 7 (2018). https://doi.org/10.1161/jaha.117.007397

114 Cox, A. J. et al. Prediction of mortality using a multi-bed vascular calcification score in the Diabetes Heart Study. Cardiovasc Diabetol 13, 160 (2014). https://doi.org/10.1186/s12933-014-0160-5

115 Cox, A. J. et al. Genetic risk score associations with cardiovascular disease and mortality in the Diabetes Heart Study. Diabetes Care 37, 1157–1164 (2014). https://doi.org/10.2337/dc13-1514

116 Cox, A. J. et al. Usefulness of biventricular volume as a predictor of mortality in patients with diabetes mellitus (from the Diabetes Heart Study). Am J Cardiol 111, 1152–1158 (2013). https://doi.org/10.1016/j.amjcard.2012.12.044

117 Cui, N. H., Yang, J. M., Liu, X. & Wang, X. B. Poly(ADP-Ribose) Polymerase Activity and Coronary Artery Disease in Type 2 Diabetes Mellitus: An Observational and Bidirectional Mendelian Randomization Study. Arterioscler Thromb Vasc Biol 40, 2516–2526 (2020). https://doi.org/10.1161/ATVBAHA.120.314712

118 Daka, B. et al. Low concentrations of serum testosterone predict acute myocardial infarction in men with type 2 diabetes mellitus. BMC Endocr Disord 15, 35 (2015). https://doi.org/10.1186/s12902-015-0034-1

119 Davis, T. M., Coleman, R. L. & Holman, R. R. Ethnicity and long-term vascular outcomes in Type 2 diabetes: a prospective observational study (UKPDS 83). Diabet Med 31, 200–207 (2014). https://doi.org/10.1111/dme.12353

120 Davis, W. A., Colagiuri, S. & Davis, T. M. Comparison of the Framingham and United Kingdom Prospective Diabetes Study cardiovascular risk equations in Australian patients with type 2 diabetes from the Fremantle Diabetes Study. Med J Aust 190, 180–184 (2009). https://doi.org/10.5694/j.1326-5377.2009.tb02684.x

121 Davis, W. A., Hellbusch, V., Hunter, M. L., Bruce, D. G. & Davis, T. M. E. Contemporary cardiovascular risk assessment for type 2 diabetes including heart failure as an outcome: The fremantle diabetes study phase ii. Journal of Clinical Medicine 9 (2020). https://doi.org/10.3390/jcm9051428

122 Davis, W. A., Knuiman, M. W. & Davis, T. M. An Australian cardiovascular risk equation for type 2 diabetes: the Fremantle Diabetes Study. Intern Med J 40, 286–292 (2010). https://doi.org/10.1111/j.1445-5994.2009.01958.x

123 Dayan, A. et al. Coronary calcium score, albuminuria and inflammatory markers in type 2 diabetic patients: associations and prognostic implications. Diabetes Res Clin Pract 98, 98–103 (2012). https://doi.org/10.1016/j.diabres.2012.04.012

124 de Galan, B. E. et al. Cognitive function and risks of cardiovascular disease and hypoglycaemia in patients with type 2 diabetes: the Action in Diabetes and Vascular Disease: Preterax and Diamicron Modified Release Controlled Evaluation (ADVANCE) trial. Diabetologia 52, 2328–2336 (2009). https://doi.org/10.1007/s00125-009-1484-7

125 De Lorenzo, A., Lima, R. S., Siqueira-Filho, A. G. & Pantoja, M. R. Prevalence and prognostic value of perfusion defects detected by stress technetium-99m sestamibi myocardial perfusion single-photon emission computed tomography in asymptomatic patients with diabetes mellitus and no known coronary artery disease. Am J Cardiol 90, 827–832 (2002). https://doi.org/10.1016/s0002-9149(02)02702-9

126 de Santiago, A., García-Lledó, A., Ramos, E. & Santiago, C. [Prognostic value of ECGs in patients with type-2 diabetes mellitus without known cardiovascular disease]. Rev Esp Cardiol 60, 1035–1041 (2007). https://doi.org/10.1157/13111235

127 de Vries, T. I. et al. Normal-range thyroid-stimulating hormone levels and cardiovascular events and mortality in type 2 diabetes. Diabetes Res Clin Pract 157, 107880 (2019). https://doi.org/10.1016/j.diabres.2019.107880

128 Djaileb, L. et al. Prognostic value of SPECT myocardial perfusion entropy in high-risk type 2 diabetic patients. Eur J Nucl Med Mol Imaging 48, 1813–1821 (2021). https://doi.org/10.1007/s00259-020-05110-4

129 Doney, A. S. et al. The FTO gene is associated with an atherogenic lipid profile and myocardial infarction in patients with type 2 diabetes: a Genetics of Diabetes Audit and Research Study in Tayside Scotland (Go-DARTS) study. Circ Cardiovasc Genet 2, 255–259 (2009). https://doi.org/10.1161/circgenetics.108.822320

130 Doney, A. S., Lee, S., Leese, G. P., Morris, A. D. & Palmer, C. N. Increased cardiovascular morbidity and mortality in type 2 diabetes is associated with the glutathione S transferase theta-null genotype: a Go-DARTS study. Circulation 111, 2927–2934 (2005). https://doi.org/10.1161/circulationaha.104.509224

131 Doney, A. S. F. et al. Association of common variation in the PPARA gene with incident myocardial infarction in individuals with type 2 diabetes: A Go-DARTS study. Nuclear Receptor 3 (2005). https://doi.org/10.1186/1478-1336-3-4

132 Donnan, P. T., Donnelly, L., New, J. P. & Morris, A. D. Derivation and validation of a prediction score for major coronary heart disease events in a U.K. type 2 diabetic population. Diabetes Care 29, 1231–1236 (2006). https://doi.org/10.2337/dc05-1911

133 Drinkwater, J. J. et al. Retinopathy predicts stroke but not myocardial infarction in type 2 diabetes: the Fremantle Diabetes Study Phase II. Cardiovasc Diabetol 19, 43 (2020). https://doi.org/10.1186/s12933-020-01018-3

134 Drury, P. L. et al. Estimated glomerular filtration rate and albuminuria are independent predictors of cardiovascular events and death in type 2 diabetes mellitus: the Fenofibrate Intervention and Event Lowering in Diabetes (FIELD) study. Diabetologia 54, 32–43 (2011). https://doi.org/10.1007/s00125-010-1854-1

135 Duan, J. G. et al. Long-term risk of cardiovascular disease among type 2 diabetic patients with asymptomatic intracranial atherosclerosis: a prospective cohort study. PLoS One 9, e106623 (2014). https://doi.org/10.1371/journal.pone.0106623

136 Duan, J. G. et al. Sex differences in epidemiology and risk factors of acute coronary syndrome in Chinese patients with type 2 diabetes: a long-term prospective cohort study. PLoS One 10, e0122031 (2015). https://doi.org/10.1371/journal.pone.0122031

137 Eguchi, K., Hoshide, S. & Kario, K. Target home morning SBP be below 125lJmmHg in type 2 diabetes patients. J Hypertens 36, 1284–1290 (2018). https://doi.org/10.1097/hjh.0000000000001683

138 Eguchi, K. et al. Differential impact of left ventricular mass and relative wall thickness on cardiovascular prognosis in diabetic and nondiabetic hypertensive subjects. Am Heart J 154, 79.e79–15 (2007). https://doi.org/10.1016/j.ahj.2007.04.021

139 Eguchi, K. et al. Night time blood pressure variability is a strong predictor for cardiovascular events in patients with type 2 diabetes. Am J Hypertens 22, 46–51 (2009). https://doi.org/10.1038/ajh.2008.294

140 Eguchi, K. et al. Ambulatory blood pressure is a better marker than clinic blood pressure in predicting cardiovascular events in patients with/without type 2 diabetes. Am J Hypertens 21, 443–450 (2008). https://doi.org/10.1038/ajh.2008.4

141 Eguchi, K. et al. Increased heart rate variability during sleep is a predictor for future cardiovascular events in patients with type 2 diabetes. Hypertens Res 33, 737–742 (2010). https://doi.org/10.1038/hr.2010.61

142 Eijkelkamp, W. B. et al. Renal function and risk for cardiovascular events in type 2 diabetic patients with hypertension: the RENAAL and LIFE studies. J Hypertens 25, 871–876 (2007). https://doi.org/10.1097/HJH.0b013e328014953c

143 Eliasson, B. et al. Clinical usefulness of different lipid measures for prediction of coronary heart disease in type 2 diabetes: a report from the Swedish National Diabetes Register. Diabetes Care 34, 2095–2100 (2011). https://doi.org/10.2337/dc11-0209

144 Elkeles, R. S. et al. Coronary calcium measurement improves prediction of cardiovascular events in asymptomatic patients with type 2 diabetes: the PREDICT study. Eur Heart J 29, 2244–2251 (2008). https://doi.org/10.1093/eurheartj/ehn279

145 Elley, C. R., Kenealy, T., Robinson, E. & Drury, P. L. Glycated haemoglobin and cardiovascular outcomes in people with Type 2 diabetes: a large prospective cohort study. Diabet Med 25, 1295–1301 (2008). https://doi.org/10.1111/j.1464-5491.2008.02581.x

146 Elley, C. R., Robinson, E., Kenealy, T., Bramley, D. & Drury, P. L. Derivation and validation of a new cardiovascular risk score for people with type 2 diabetes: the new zealand diabetes cohort study. Diabetes Care 33, 1347–1352 (2010). https://doi.org/10.2337/dc09-1444

147 Estacio, R. O., Dale, R. A., Schrier, R. & Krantz, M. J. Relation of reduction in urinary albumin excretion to ten-year cardiovascular mortality in patients with type 2 diabetes and systemic hypertension. Am J Cardiol 109, 1743–1748 (2012). https://doi.org/10.1016/j.amjcard.2012.02.020

148 Everett, B. M. et al. Troponin and Cardiac Events in Stable Ischemic Heart Disease and Diabetes. N Engl J Med 373, 610–620 (2015). https://doi.org/10.1056/NEJMoa1415921

149 Fadini, G. P. et al. p66Shc gene expression in peripheral blood mononuclear cells and progression of diabetic complications. Cardiovasc Diabetol 17, 16 (2018). https://doi.org/10.1186/s12933-018-0660-9

150 Fadini, G. P., Rigato, M., Cappellari, R., Bonora, B. M. & Avogaro, A. Long-term Prediction of Cardiovascular Outcomes by Circulating CD34+ and CD34+CD133+ Stem Cells in Patients With Type 2 Diabetes. Diabetes Care 40, 125–131 (2017). https://doi.org/10.2337/dc16-1755

151 Faghihi-Kashani, S. et al. Fasting hyperinsulinaemia and 2-h glycaemia predict coronary heart disease in patients with type 2 diabetes. Diabetes Metab 42, 55–61 (2016). https://doi.org/10.1016/j.diabet.2015.10.001

152 Faglia, E. et al. Cardiac events in 735 type 2 diabetic patients who underwent screening for unknown asymptomatic coronary heart disease: 5-year follow-up report from the Milan Study on Atherosclerosis and Diabetes (MiSAD). Diabetes Care 25, 2032–2036 (2002). https://doi.org/10.2337/diacare.25.11.2032

153 Ferrarezi, D. A. et al. Allelic variations of the vitamin D receptor (VDR) gene are associated with increased risk of coronary artery disease in type 2 diabetics: the DIABHYCAR prospective study. Diabetes Metab 39, 263–270 (2013). https://doi.org/10.1016/j.diabet.2012.11.004

154 Filippella, M. et al. Ankle brachial pressure index usefulness as predictor factor for coronary heart disease in diabetic patients. J Endocrinol Invest 30, 721–725 (2007). https://doi.org/10.1007/bf03350808

155 Florkowski, C. M., Scott, R. S., Coope, P. A. & Moir, C. L. Predictors of mortality from type 2 diabetes mellitus in Canterbury, New Zealand; a ten-year cohort study. Diabetes Res Clin Pract 53, 113–120 (2001). https://doi.org/10.1016/s0168-8227(01)00246-7

156 Folsom, A. R., Chambless, L. E., Duncan, B. B., Gilbert, A. C. & Pankow, J. S. Prediction of coronary heart disease in middle-aged adults with diabetes. Diabetes Care 26, 2777–2784 (2003). https://doi.org/10.2337/diacare.26.10.2777

157 Fox, C. S., Sullivan, L., D’Agostino, R. B., Sr. & Wilson, P. W. The significant effect of diabetes duration on coronary heart disease mortality: the Framingham Heart Study. Diabetes Care 27, 704–708 (2004). https://doi.org/10.2337/diacare.27.3.704

158 Fragoso, A., Mendes, F., Silva, A. P. & Neves, P. L. Insulin resistance as a predictor of cardiovascular morbidity and end-stage renal disease. J Diabetes Complications 29, 1098–1104 (2015). https://doi.org/10.1016/j.jdiacomp.2015.05.010

159 Freemantle, N., Danchin, N., Calvi-Gries, F., Vincent, M. & Home, P. D. Relationship of glycaemic control and hypoglycaemic episodes to 4-year cardiovascular outcomes in people with type 2 diabetes starting insulin. Diabetes Obes Metab 18, 152–158 (2016). https://doi.org/10.1111/dom.12598

160 Friedman, A. N., Hunsicker, L. G., Selhub, J. & Bostom, A. G. Total plasma homocysteine and arteriosclerotic outcomes in type 2 diabetes with nephropathy. J Am Soc Nephrol 16, 3397–3402 (2005). https://doi.org/10.1681/asn.2004100846

161 Fukushima, H. et al. Prognostic value of remnant-like lipoprotein particle levels in patients with coronary artery disease and type II diabetes mellitus. J Am Coll Cardiol 43, 2219–2224 (2004). https://doi.org/10.1016/j.jacc.2003.09.074

162 Fuller, J. H., Stevens, L. K. & Wang, S. L. Risk factors for cardiovascular mortality and morbidity: the WHO Mutinational Study of Vascular Disease in Diabetes. Diabetologia 44 **Suppl 2**, S54–64 (2001). https://doi.org/10.1007/pl00002940

163 Fumisawa, Y. et al. Systematic analysis of risk factors for coronary heart disease in Japanese patients with type 2 diabetes: a matched case-control study. J Atheroscler Thromb 19, 918–923 (2012). https://doi.org/10.5551/jat.13334

164 Gasior, M. et al. Effect of blood glucose levels on prognosis in acute myocardial infarction in patients with and without diabetes, undergoing percutaneous coronary intervention. Cardiol J 15, 422–430 (2008).

165 Gazzaruso, C. et al. Transcutaneous oxygen tension as a potential predictor of cardiovascular events in type 2 diabetes: comparison with ankle-brachial index. Diabetes Care 36, 1720–1725 (2013). https://doi.org/10.2337/dc12-1401

166 Gazzaruso, C. et al. Lipoprotein(a), apolipoprotein(a) polymorphism and restenosis after intracoronary stent placement in Type 2 diabetic patients. J Diabetes Complications 17, 135–140 (2003). https://doi.org/10.1016/s1056-8727(02)00192-7

167 Gazzaruso, C. et al. Erectile dysfunction as a predictor of cardiovascular events and death in diabetic patients with angiographically proven asymptomatic coronary artery disease: a potential protective role for statins and 5-phosphodiesterase inhibitors. J Am Coll Cardiol 51, 2040–2044 (2008). https://doi.org/10.1016/j.jacc.2007.10.069

168 Georgoulias, P. et al. Long-term prognostic value of heart-rate recovery after treadmill testing in patients with diabetes mellitus. Int J Cardiol 134, 67–74 (2009). https://doi.org/10.1016/j.ijcard.2008.01.036

169 Gimeno-Orna, J. A., Lou-Arnal, L. M., Boned-Juliani, B. & Molinero-Herguedas, E. Mild renal insufficiency as a cardiovascular risk factor in non-proteinuric type II diabetes. Diabetes Res Clin Pract 64, 191–199 (2004). https://doi.org/10.1016/j.diabres.2003.10.018

170 Giorda, C. B. et al. Recurrence of cardiovascular events in patients with type 2 diabetes: epidemiology and risk factors. Diabetes Care 31, 2154–2159 (2008). https://doi.org/10.2337/dc08-1013

171 Giovacchini, G. et al. Microalbuminuria predicts silent myocardial ischaemia in type 2 diabetes patients. Eur J Nucl Med Mol Imaging 40, 548–557 (2013). https://doi.org/10.1007/s00259-012-2323-5

172 Group, T. L. A. R. Prospective Association of GLUL rs10911021 With Cardiovascular Morbidity and Mortality Among Individuals With Type 2 Diabetes: The Look AHEAD Study. Diabetes 65, 297–302 (2015). https://doi.org/10.2337/db15-0890

173 Guzder, R. N., Gatling, W., Mullee, M. A. & Byrne, C. D. Impact of metabolic syndrome criteria on cardiovascular disease risk in people with newly diagnosed type 2 diabetes. Diabetologia 49, 49–55 (2006). https://doi.org/10.1007/s00125-005-0063-9

174 Guzder, R. N., Gatling, W., Mullee, M. A., Mehta, R. L. & Byrne, C. D. Prognostic value of the Framingham cardiovascular risk equation and the UKPDS risk engine for coronary heart disease in newly diagnosed Type 2 diabetes: results from a United Kingdom study. Diabet Med 22, 554–562 (2005). https://doi.org/10.1111/j.1464-5491.2005.01494.x

175 Hadaegh, F. et al. Electrocardiography-defined silent CHD and risk of cardiovascular events among diabetic patients in a Middle Eastern population. Eur J Prev Cardiol 19, 1227–1233 (2012). https://doi.org/10.1177/1741826711428065

176 Hadjadj, S. et al. Prognostic value of the insertion/deletion polymorphism of the ACE gene in type 2 diabetic subjects: results from the Non-insulin-dependent Diabetes, Hypertension, Microalbuminuria or Proteinuria, Cardiovascular Events, and Ramipril (DIABHYCAR), Diabete d. Diabetes Care 31, 1847–1852 (2008). https://doi.org/10.2337/dc07-2079

177 Hage, F. G. et al. The heart rate response to adenosine: a simple predictor of adverse cardiac outcomes in asymptomatic patients with type 2 diabetes. Int J Cardiol 167, 2952–2957 (2013). https://doi.org/10.1016/j.ijcard.2012.08.011

178 Halon, D. A. et al. Coronary Computed Tomography (CT) Angiography as a Predictor of Cardiac and Noncardiac Vascular Events in Asymptomatic Type 2 Diabetics: A 7-Year Population-Based Cohort Study. J Am Heart Assoc 5 (2016). https://doi.org/10.1161/jaha.116.003226

179 Halon, D. A. et al. Plaque Morphology as Predictor of Late Plaque Events in Patients With Asymptomatic Type 2 Diabetes: A Long-Term Observational Study. JACC Cardiovasc Imaging 12, 1353–1363 (2019). https://doi.org/10.1016/j.jcmg.2018.02.025

180 Hamada, S. & Gulliford, M. C. Multiple risk factor control, mortality and cardiovascular events in type 2 diabetes and chronic kidney disease: a population-based cohort study. BMJ Open 8, e019950 (2018). https://doi.org/10.1136/bmjopen-2017-019950

181 Hanefeld, M. et al. Risk factors for myocardial infarction and death in newly detected NIDDM: the Diabetes Intervention Study, 11-year follow-up. Diabetologia 39, 1577-1583 (1996). https://doi.org/10.1007/s001250050617

182 Hata, J. et al. Effects of visit-to-visit variability in systolic blood pressure on macrovascular and microvascular complications in patients with type 2 diabetes mellitus: the ADVANCE trial. Circulation 128, 1325–1334 (2013). https://doi.org/10.1161/circulationaha.113.002717

183 Hayashi, T. et al. Metabolic predictors of ischemic heart disease and cerebrovascular attack in elderly diabetic individuals: difference in risk by age. Cardiovasc Diabetol 12, 10 (2013). https://doi.org/10.1186/1475-2840-12-10

184 Hayes, A. J., Leal, J., Gray, A. M., Holman, R. R. & Clarke, P. M. UKPDS outcomes model 2: a new version of a model to simulate lifetime health outcomes of patients with type 2 diabetes mellitus using data from the 30 year United Kingdom Prospective Diabetes Study: UKPDS 82. Diabetologia 56, 1925–1933 (2013). https://doi.org/10.1007/s00125-013-2940-y

185 He, Q., Pan, J., Wang, L., Fang, Y. & Hu, R. Prospective study: Aldehyde dehydrogenase 2 gene is associated with cardio-cerebrovascular complications in type 2 diabetes patients. J Diabetes Investig 12, 1845–1854 (2021). https://doi.org/10.1111/jdi.13538

186 Heidari, B. et al. Assessment of serum 25-hydroxy vitamin D improves coronary heart disease risk stratification in patients with type 2 diabetes. Am Heart J 170, 573–579.e575 (2015). https://doi.org/10.1016/j.ahj.2015.06.017

187 Heijmans, B. T. et al. Common paraoxonase gene variants, mortality risk and fatal cardiovascular events in elderly subjects. Atherosclerosis 149, 91–97 (2000). https://doi.org/10.1016/s0021-9150(99)00311-1

188 Ho, J. S. et al. Association of the PPARG Pro12Ala polymorphism with type 2 diabetes and incident coronary heart disease in a Hong Kong Chinese population. Diabetes Res Clin Pract 97, 483–491 (2012). https://doi.org/10.1016/j.diabres.2012.03.012

189 Hoffmann, M. M., März, W., Genser, B., Drechsler, C. & Wanner, C. Lack of association between the Trp719Arg polymorphism in kinesin-like protein-6 and cardiovascular risk and efficacy of atorvastatin among subjects with diabetes on dialysis: the 4D study. Atherosclerosis 219, 659–662 (2011). https://doi.org/10.1016/j.atherosclerosis.2011.07.126

190 Hong, L. F. et al. Predictive value of non-fasting remnant cholesterol for short-term outcome of diabetics with new-onset stable coronary artery disease. Lipids Health Dis 16, 7 (2017). https://doi.org/10.1186/s12944-017-0410-0

191 Howard, B. V. et al. LDL cholesterol as a strong predictor of coronary heart disease in diabetic individuals with insulin resistance and low LDL: The Strong Heart Study. Arterioscler Thromb Vasc Biol 20, 830–835 (2000). https://doi.org/10.1161/01.atv.20.3.830

192 Hsieh, Y. T. et al. Subnormal Estimated Glomerular Filtration Rate Strongly Predict Incident Cardiovascular Events in Type 2 Diabetic Chinese Population With Normoalbuminuria. Medicine (Baltimore) 95, e2200 (2016). https://doi.org/10.1097/md.0000000000002200

193 Hu, C. P. et al. Platelet Distribution Width on Admission Predicts In-Stent Restenosis in Patients with Coronary Artery Disease and Type 2 Diabetes Mellitus Treated with Percutaneous Coronary Intervention. Chin Med J (Engl) 131, 757–763 (2018). https://doi.org/10.4103/0366-6999.228247

194 Huang, X. H. et al. Angiotensin-converting enzyme gene polymorphism is associated with coronary heart disease in non-insulin-dependent diabetic patients evaluated for 9 years. Metabolism 47, 1258–1262 (1998). https://doi.org/10.1016/s0026-0495(98)90333-x

195 Hunt, K. J. et al. Plasma Connective Tissue Growth Factor (CTGF/CCN2) Levels Predict Myocardial Infarction in the Veterans Affairs Diabetes Trial (VADT) Cohort. Diabetes Care 41, 840–846 (2018). https://doi.org/10.2337/dc17-2083

196 Iijima, K. et al. Lower physical activity is a strong predictor of cardiovascular events in elderly patients with type 2 diabetes mellitus beyond traditional risk factors: the Japanese Elderly Diabetes Intervention Trial. Geriatr Gerontol Int 12 **Suppl 1**, 77–87 (2012). https://doi.org/10.1111/j.1447-0594.2011.00815.x

197 Ikeda, Y. et al. Low human paraoxonase predicts cardiovascular events in Japanese patients with type 2 diabetes. Acta Diabetol 46, 239–242 (2009). https://doi.org/10.1007/s00592-008-0066-3

198 Irie, Y. et al. The utility of ultrasonic tissue characterization of carotid plaque in the prediction of cardiovascular events in diabetic patients. Atherosclerosis 230, 399–405 (2013). https://doi.org/10.1016/j.atherosclerosis.2013.08.015

199 Jeevarethinam, A. et al. Usefulness of Carotid Plaques as Predictors of Obstructive Coronary Artery Disease and Cardiovascular Events in Asymptomatic Individuals With Diabetes Mellitus. Am J Cardiol 121, 910–916 (2018). https://doi.org/10.1016/j.amjcard.2018.01.001

200 Jha, D. et al. Prognostic role of soluble ST2 in acute coronary syndrome with diabetes. Eur J Clin Invest 48, e12994 (2018). https://doi.org/10.1111/eci.12994

201 Jiang, R. et al. Non-HDL cholesterol and apolipoprotein B predict cardiovascular disease events among men with type 2 diabetes. Diabetes Care 27, 1991–1997 (2004). https://doi.org/10.2337/diacare.27.8.1991

202 Jimenez-Corona, A. et al. Electrocardiographic abnormalities predict deaths from cardiovascular disease and ischemic heart disease in Pima Indians with type 2 diabetes. Am Heart J 151, 1080–1086 (2006). https://doi.org/10.1016/j.ahj.2005.06.033

203 Jin, J. L. et al. Triglyceride glucose and haemoglobin glycation index for predicting outcomes in diabetes patients with new-onset, stable coronary artery disease: a nested case-control study. Ann Med 50, 576–586 (2018). https://doi.org/10.1080/07853890.2018.1523549

204 Johnston, S. S. et al. Evidence linking hypoglycemic events to an increased risk of acute cardiovascular events in patients with type 2 diabetes. Diabetes Care 34, 1164–1170 (2011). https://doi.org/10.2337/dc10-1915

205 Juutilainen, A., Lehto, S., Suhonen, M., Rönnemaa, T. & Laakso, M. Thoracoabdominal calcifications predict cardiovascular disease mortality in type 2 diabetic and nondiabetic subjects: 18-year follow-up study. Diabetes Care 33, 583–585 (2010). https://doi.org/10.2337/dc09-1813

206 Kamoi, K., Ito, T., Miyakoshi, M. & Minagawa, S. Usefulness of home blood pressure measurement in the morning in patients with type 2 diabetes: long-term results of a prospective longitudinal study. Clin Exp Hypertens 32, 184–192 (2010). https://doi.org/10.3109/10641960903254513

207 Katakami, N. et al. Accumulation of oxidative stress-related gene polymorphisms and the risk of coronary heart disease events in patients with type 2 diabetes--an 8-year prospective study. Atherosclerosis 235, 408–414 (2014). https://doi.org/10.1016/j.atherosclerosis.2014.05.936

208 Katakami, N. et al. Clinical utility of brachial-ankle pulse wave velocity in the prediction of cardiovascular events in diabetic patients. Cardiovasc Diabetol 13, 128 (2014). https://doi.org/10.1186/s12933-014-0128-5

209 Katakami, N. et al. Ultrasonic tissue characterization of carotid plaque improves the prediction of cardiovascular events in diabetic patients: a pilot study. Diabetes Care 35, 2640–2646 (2012). https://doi.org/10.2337/dc12-0331

210 Kawasaki, R. et al. Risk of cardiovascular diseases is increased even with mild diabetic retinopathy: the Japan Diabetes Complications Study. Ophthalmology 120, 574–582 (2013). https://doi.org/10.1016/j.ophtha.2012.08.029

211 Keavney, B. D. et al. UK prospective diabetes study (UKPDS) 14: association of angiotensin-converting enzyme insertion/deletion polymorphism with myocardial infarction in NIDDM. Diabetologia 38, 948–952 (1995). https://doi.org/10.1007/BF00400584

212 Keller, T. et al. Prognostic Value of High-Sensitivity Versus Conventional Cardiac Troponin T Assays Among Patients With Type 2 Diabetes Mellitus Undergoing Maintenance Hemodialysis. Am J Kidney Dis 71, 822–830 (2018). https://doi.org/10.1053/j.ajkd.2017.10.016

213 Kenealy, T. et al. An association between ethnicity and cardiovascular outcomes for people with Type 2 diabetes in New Zealand. Diabet Med 25, 1302–1308 (2008). https://doi.org/10.1111/j.1464-5491.2008.02593.x

214 Kengne, A. P. et al. The Framingham and UK Prospective Diabetes Study (UKPDS) risk equations do not reliably estimate the probability of cardiovascular events in a large ethnically diverse sample of patients with diabetes: the Action in Diabetes and Vascular Disease: Preterax. Diabetologia 53, 821–831 (2010). https://doi.org/10.1007/s00125-010-1681-4

215 Kengne, A. P. et al. Contemporary model for cardiovascular risk prediction in people with type 2 diabetes. Eur J Cardiovasc Prev Rehabil 18, 393–398 (2011). https://doi.org/10.1177/1741826710394270

216 Khalili, S., Hatami, M., Hadaegh, F., Sheikholeslami, F. & Azizi, F. Prediction of cardiovascular events with consideration of general and central obesity measures in diabetic adults: results of the 8.4-year follow-up. Metab Syndr Relat Disord 10, 218–224 (2012). https://doi.org/10.1089/met.2011.0070

217 Kim, M. K. et al. Hemoglobin glycation index predicts cardiovascular disease in people with type 2 diabetes mellitus: A 10-year longitudinal cohort study. J Diabetes Complications 32, 906–910 (2018). https://doi.org/10.1016/j.jdiacomp.2018.08.007

218 Koch, M., Kutkuhn, B., Grabensee, B. & Ritz, E. Apolipoprotein A, fibrinogen, age, and history of stroke are predictors of death in dialysed diabetic patients: a prospective study in 412 subjects. Nephrol Dial Transplant 12, 2603–2611 (1997). https://doi.org/10.1093/ndt/12.12.2603

219 Koo, B. K., Chung, W. Y. & Moon, M. K. Peripheral arterial endothelial dysfunction predicts future cardiovascular events in diabetic patients with albuminuria: a prospective cohort study. Cardiovasc Diabetol 19, 82 (2020). https://doi.org/10.1186/s12933-020-01062-z

220. Kuricová, K., et al. NOS3 894G>T polymorphism is associated with progression of kidney disease and cardiovascular morbidity in type 2 diabetic patients: NOS3 as a modifier gene for diabetic nephropathy? Kidney Blood Press Res 38, 92–98 (2013). https://doi.org/10.1159/000355757

221 Lagani, V. et al. Development and validation of risk assessment models for diabetes-related complications based on the DCCT/EDIC data. J Diabetes Complications 29, 479–487 (2015). https://doi.org/10.1016/j.jdiacomp.2015.03.001

222 Lapin, B. R. et al. Pain in Patients With Type 2 Diabetes-Related Polyneuropathy Is Associated With Vascular Events and Mortality. J Clin Endocrinol Metab 105 (2020). https://doi.org/10.1210/clinem/dgaa394

223 Lau, K. K. et al. Prognostic implications of surrogate markers of atherosclerosis in low to intermediate risk patients with type 2 diabetes. Cardiovasc Diabetol 11, 101 (2012). https://doi.org/10.1186/1475-2840-11-101

224 Le Feuvre, C. L. et al. Stress myocardial scintigraphy and dobutamine echocardiography in the detection of coronary disease in asymptomatic patients with type 2 diabetes. Diabetes Metab 31, 135–142 (2005). https://doi.org/10.1016/s1262-3636(07)70179-9

225 Lee, K. Y. et al. Computed Tomography Angiography Images of Coronary Artery Stenosis Provide a Better Prediction of Risk Than Traditional Risk Factors in Asymptomatic Individuals With Type 2 Diabetes: A Long-term Study of Clinical Outcomes. Diabetes Care 40, 1241–1248 (2017). https://doi.org/10.2337/dc16-1844

226 Lehto, S., Niskanen, L., Suhonen, M., Rönnemaa, T. & Laakso, M. Medial artery calcification. A neglected harbinger of cardiovascular complications in non-insulin-dependent diabetes mellitus. Arterioscler Thromb Vasc Biol 16, 978–983 (1996). https://doi.org/10.1161/01.atv.16.8.978

227 Lepojärvi, E. S. et al. Usefulness of Highly Sensitive Troponin as a Predictor of Short-Term Outcome in Patients With Diabetes Mellitus and Stable Coronary Artery Disease (from the ARTEMIS Study). Am J Cardiol 117, 515–521 (2016). https://doi.org/10.1016/j.amjcard.2015.11.038

228 Levy, A. P. et al. Haptoglobin phenotype is an independent risk factor for cardiovascular disease in individuals with diabetes: The Strong Heart Study. J Am Coll Cardiol 40, 1984–1990 (2002). https://doi.org/10.1016/s0735-1097(02)02534-2

229 Li, P. I., Wang, J. N. & Guo, H. R. A long-term quality-of-care score for predicting the occurrence of macrovascular diseases in patients with type 2 diabetes mellitus. Diabetes Res Clin Pract 139, 72–80 (2018). https://doi.org/10.1016/j.diabres.2018.02.027

230 Li, W. P. et al. Pregnancy-associated plasma protein-A is a stronger predictor for adverse cardiovascular outcomes after acute coronary syndrome in type-2 diabetes mellitus. Cardiovasc Diabetol 16, 45 (2017). https://doi.org/10.1186/s12933-017-0526-6

231 Liao, K. M. et al. Risk assessment of macrovascular and microvascular events in patients with type 2 diabetes by analyzing the amplitude variation of the fourth harmonic component of radial pulse wave. Physiol Rep 7, e14252 (2019). https://doi.org/10.14814/phy2.14252

232 Lièvre, M. M. et al. Detection of silent myocardial ischemia in asymptomatic patients with diabetes: results of a randomized trial and meta-analysis assessing the effectiveness of systematic screening. Trials 12, 23 (2011). https://doi.org/10.1186/1745-6215-12-23

233 Lim, L. L. et al. Sudomotor dysfunction independently predicts incident cardiovascular-renal events and all-cause death in type 2 diabetes: the Joint Asia Diabetes Evaluation register. Nephrol Dial Transplant 34, 1320–1328 (2019). https://doi.org/10.1093/ndt/gfy154

234 Lim, S. et al. Association of adiponectin and resistin with cardiovascular events in Korean patients with type 2 diabetes: the Korean atherosclerosis study (KAS): a 42-month prospective study. Atherosclerosis 196, 398–404 (2008). https://doi.org/10.1016/j.atherosclerosis.2006.11.017

235 Lin, C. H., Li, H. Y., Jiang, Y. D., Chang, T. J. & Chuang, L. M. Plasma YKL-40 predicts 10-year cardiovascular and all-cause mortality in individuals with type 2 diabetes. Clin Endocrinol (Oxf) 79, 185–191 (2013). https://doi.org/10.1111/cen.12015

236 Lin, E. H. et al. Depression and advanced complications of diabetes: a prospective cohort study. Diabetes Care 33, 264–269 (2010). https://doi.org/10.2337/dc09-1068

237 Lin, L. Y. et al. The ankle brachial index exhibits better association with cardiovascular outcomes than interarm systolic blood pressure difference in patients with type 2 diabetes. Medicine (Baltimore) 98, e15556 (2019). https://doi.org/10.1097/md.0000000000015556

238 Linnemann, B. & Janka, H. U. Prolonged QTc interval and elevated heart rate identify the type 2 diabetic patient at high risk for cardiovascular death. The Bremen Diabetes Study. Exp Clin Endocrinol Diabetes 111, 215–222 (2003). https://doi.org/10.1055/s-2003-40466

239 Linnemann, B., Voigt, W., Nobel, W. & Janka, H. U. C-reactive protein is a strong independent predictor of death in type 2 diabetes: association with multiple facets of the metabolic syndrome. Exp Clin Endocrinol Diabetes 114, 127–134 (2006). https://doi.org/10.1055/s-2006-924012

240 Lopes-Virella, M. F., Hunt, K. J., Baker, N. L., Virella, G. & Moritz, T. The levels of MDA-LDL in circulating immune complexes predict myocardial infarction in the VADT study. Atherosclerosis 224, 526–531 (2012). https://doi.org/10.1016/j.atherosclerosis.2012.08.006

241 Lorenzo-Almorós, A. et al. Galectin-3 is associated with cardiovascular events in post-acute coronary syndrome patients with type-2 diabetes. Journal of Clinical Medicine 9 (2020). https://doi.org/10.3390/jcm9041105

242 Lowe, G. et al. Circulating inflammatory markers and the risk of vascular complications and mortality in people with type 2 diabetes and cardiovascular disease or risk factors: the ADVANCE study. Diabetes 63, 1115–1123 (2014). https://doi.org/10.2337/db12-1625

243 Lu, T. M., Lin, S. J., Lin, M. W., Hsu, C. P. & Chung, M. Y. The association of dimethylarginine dimethylaminohydrolase 1 gene polymorphism with type 2 diabetes: a cohort study. Cardiovasc Diabetol 10, 16 (2011). https://doi.org/10.1186/1475-2840-10-16

244 Lu, W. et al. Non-HDL cholesterol as a predictor of cardiovascular disease in type 2 diabetes: the strong heart study. Diabetes Care 26, 16–23 (2003). https://doi.org/10.2337/diacare.26.1.16

245 Lutgers, H. L. et al. Skin autofluorescence provides additional information to the UK Prospective Diabetes Study (UKPDS) risk score for the estimation of cardiovascular prognosis in type 2 diabetes mellitus. Diabetologia 52, 789–797 (2009). https://doi.org/10.1007/s00125-009-1308-9

246 Masi, S. et al. Telomere length, antioxidant status and incidence of ischaemic heart disease in type 2 diabetes. Int J Cardiol 216, 159–164 (2016). https://doi.org/10.1016/j.ijcard.2016.04.130

247 Massardo, T. et al. Factors associated with silent myocardial ischemia, autonomic or peripheral neuropathies, and survival in diabetes mellitus type 2 patients without cardiovascular symptoms. International Journal of Diabetes in Developing Countries 40, 80–86 (2020). https://doi.org/10.1007/s13410-019-00758-7

248 McEwan, P., Bennett, H., Ward, T. & Bergenheim, K. Refitting of the UKPDS 68 risk equations to contemporary routine clinical practice data in the UK. Pharmacoeconomics 33, 149–161 (2015). https://doi.org/10.1007/s40273-014-0225-z

249 McMurray, J. J. et al. Predictors of fatal and nonfatal cardiovascular events in patients with type 2 diabetes mellitus, chronic kidney disease, and anemia: an analysis of the Trial to Reduce cardiovascular Events with Aranesp (darbepoetin-alfa) Therapy (TREAT). Am Heart J 162, 748–755.e743 (2011). https://doi.org/10.1016/j.ahj.2011.07.016

250 Meerwaldt, R. et al. Skin autofluorescence is a strong predictor of cardiac mortality in diabetes. Diabetes Care 30, 107–112 (2007). https://doi.org/10.2337/dc06-1391

251 Mellbin, L. G. et al. Copeptin, IGFBP-1, and cardiovascular prognosis in patients with type 2 diabetes and acute myocardial infarction: a report from the DIGAMI 2 trial. Diabetes Care 33, 1604–1606 (2010). https://doi.org/10.2337/dc10-0088

252 Mentz, R. J. et al. Effect of Once-Weekly Exenatide on Clinical Outcomes According to Baseline Risk in Patients With Type 2 Diabetes Mellitus: Insights From the EXSCEL Trial. J Am Heart Assoc 7, e009304 (2018). https://doi.org/10.1161/jaha.118.009304

253 Mohammedi, K. et al. Plasma extracellular superoxide dismutase concentration, allelic variations in the SOD3 gene and risk of myocardial infarction and all-cause mortality in people with type 1 and type 2 diabetes. Cardiovasc Diabetol 14, 845 (2015). https://doi.org/10.1186/s12933-014-0163-2

254 Mohammedi, K. et al. Absence of Peripheral Pulses and Risk of Major Vascular Outcomes in Patients With Type 2 Diabetes. Diabetes Care 39, 2270–2277 (2016). https://doi.org/10.2337/dc16-1594

255 Monseu, M. et al. Acute Kidney Injury Predicts Major Adverse Outcomes in Diabetes: Synergic Impact With Low Glomerular Filtration Rate and Albuminuria. Diabetes Care 38, 2333–2340 (2015). https://doi.org/10.2337/dc15-1222

256 Moosaie, F. et al. Lp(a) and Apo-lipoproteins as predictors for micro- and macrovascular complications of diabetes: A case-cohort study. Nutr Metab Cardiovasc Dis 30, 1723–1731 (2020). https://doi.org/10.1016/j.numecd.2020.05.011

257 Morieri, M. L. et al. Genetic Tools for Coronary Risk Assessment in Type 2 Diabetes: A Cohort Study From the ACCORD Clinical Trial. Diabetes Care 41, 2404–2413 (2018). https://doi.org/10.2337/dc18-0709

258 Morrish, N. J., Stevens, L. K., Fuller, J. H., Jarrett, R. J. & Keen, H. Risk factors for macrovascular disease in diabetes mellitus: the London follow-up to the WHO Multinational Study of Vascular Disease in Diabetics. Diabetologia 34, 590–594 (1991). https://doi.org/10.1007/bf00400279

259 Mukamal, K. J. et al. Prediction and classification of cardiovascular disease risk in older adults with diabetes. Diabetologia 56, 275–283 (2013). https://doi.org/10.1007/s00125-012-2772-1

260 Nag, S. et al. All-cause and cardiovascular mortality in diabetic subjects increases significantly with reduced estimated glomerular filtration rate (eGFR): 10 years’ data from the South Tees Diabetes Mortality study. Diabet Med 24, 10–17 (2007). https://doi.org/10.1111/j.1464-5491.2007.02023.x

261 Nagamachi, S. et al. Prognostic value of cardiac I-123 metaiodobenzylguanidine imaging in patients with non-insulin-dependent diabetes mellitus. J Nucl Cardiol 13, 34–42 (2006). https://doi.org/10.1016/j.nuclcard.2005.11.009

262 Nakamura, M. et al. Brachial-ankle pulse wave velocity as a risk stratification index for the short-term prognosis of type 2 diabetic patients with coronary artery disease. Hypertens Res 33, 1018–1024 (2010). https://doi.org/10.1038/hr.2010.126

263 Nam, G. E. et al. Body Weight Variability and the Risk of Cardiovascular Outcomes and Mortality in Patients With Type 2 Diabetes: A Nationwide Cohort Study. Diabetes Care 43, 2234–2241 (2020). https://doi.org/10.2337/dc19-2552

264 Nargesi, A. A. et al. Nonlinear relation between pulse pressure and coronary heart disease in patients with type 2 diabetes or hypertension. J Hypertens 34, 974–980 (2016). https://doi.org/10.1097/hjh.0000000000000866

265 Nazimek-Siewniak, B., Moczulski, D. & Grzeszczak, W. Risk of macrovascular and microvascular complications in Type 2 diabetes: results of longitudinal study design. J Diabetes Complications 16, 271–276 (2002). https://doi.org/10.1016/s1056-8727(01)00184-2

266 Ndrepepa, G. et al. Prognostic value of uric acid in patients with Type 2 diabetes mellitus and coronary artery disease. Clin Sci (Lond) 124, 259–268 (2013). https://doi.org/10.1042/cs20120336

267 Nelson, R. G. et al. Low incidence of fatal coronary heart disease in Pima Indians despite high prevalence of non-insulin-dependent diabetes. Circulation 81, 987–995 (1990). https://doi.org/10.1161/01.cir.81.3.987

268 Neves, A. L. et al. Allelic variations in superoxide dismutase-1 (SOD1) gene and renal and cardiovascular morbidity and mortality in type 2 diabetic subjects. Mol Genet Metab 106, 359–365 (2012). https://doi.org/10.1016/j.ymgme.2012.04.023

269 Nichols, G. A., Joshua-Gotlib, S. & Parasuraman, S. Independent contribution of A1C, systolic blood pressure, and LDL cholesterol control to risk of cardiovascular disease hospitalizations in type 2 diabetes: an observational cohort study. J Gen Intern Med 28, 691–697 (2013). https://doi.org/10.1007/s11606-012-2320-1

270 Nilsson, P. M. et al. Smoking as an independent risk factor for myocardial infarction or stroke in type 2 diabetes: a report from the Swedish National Diabetes Register. Eur J Cardiovasc Prev Rehabil 16, 506–512 (2009). https://doi.org/10.1097/HJR.0b013e32832ccc50

271 Ningshen, R., Moathung Odyuo, Z., Arvind, G., Deba Singh, T. S. & Devi, B. A study of QTc interval prolongation as an independent predictor of cardiac mortality in type 2 diabetes mellitus. JMS - Journal of Medical Society 26, 21–25 (2012).

272 Niskanen, L., Turpeinen, A., Penttilä, I. & Uusitupa, M. I. Hyperglycemia and compositional lipoprotein abnormalities as predictors of cardiovascular mortality in type 2 diabetes: a 15-year follow-up from the time of diagnosis. Diabetes Care 21, 1861–1869 (1998). https://doi.org/10.2337/diacare.21.11.1861

273 Niskanen, L. K., Penttilã, I., Parviainen, M. & Uusitupa, M. I. Evolution, risk factors, and prognostic implications of albuminuria in NIDDM. Diabetes Care 19, 486–493 (1996). https://doi.org/10.2337/diacare.19.5.486

274 Nitenberg, A. et al. Cardiovascular outcome of patients with abnormal coronary vasomotion and normal coronary arteriography is worse in type 2 diabetes mellitus than in arterial hypertension: a 10 year follow-up study. Atherosclerosis 183, 113–120 (2005). https://doi.org/10.1016/j.atherosclerosis.2005.02.030

275 Novo-Rodríguez, C. et al. Circulating levels of sclerostin are associated with cardiovascular mortality. PLoS One 13, e0199504 (2018). https://doi.org/10.1371/journal.pone.0199504

276 Odeberg, J., Larsson, C. A., Råstam, L. & Lindblad, U. The Asp298 allele of endothelial nitric oxide synthase is a risk factor for myocardial infarction among patients with type 2 diabetes mellitus. BMC Cardiovasc Disord 8, 36 (2008). https://doi.org/10.1186/1471-2261-8-36

277 Oellgaard, J. et al. Application of urinary proteomics as possible risk predictor of renal and cardiovascular complications in patients with type 2-diabetes and microalbuminuria. J Diabetes Complications 32, 1133–1140 (2018). https://doi.org/10.1016/j.jdiacomp.2018.09.012

278 Oliveira, J. L. et al. Prognostic value of exercise echocardiography in diabetic patients. Cardiovasc Ultrasound 7, 24 (2009). https://doi.org/10.1186/1476-7120-7-24

279 Ong, K. L. et al. The relationship of fibroblast growth factor 21 with cardiovascular outcome events in the Fenofibrate Intervention and Event Lowering in Diabetes study. Diabetologia 58, 464–473 (2015). https://doi.org/10.1007/s00125-014-3458-7

280 Ong, K. L. et al. Relationships of adipocyte-fatty acid binding protein and lipocalin 2 with risk factors and chronic complications in type 2 diabetes and effects of fenofibrate: A fenofibrate Intervention and event lowering in diabetes sub-study. Diabetes Res Clin Pract 169, 108450 (2020). https://doi.org/10.1016/j.diabres.2020.108450

281 Ortega Moreno, L., et al. Evidence of a causal relationship between high serum adiponectin levels and increased cardiovascular mortality rate in patients with type 2 diabetes. Cardiovasc Diabetol 15, 17 (2016). https://doi.org/10.1186/s12933-016-0339-z

282 Oshima, M. et al. Early Change in Albuminuria with Canagliflozin Predicts Kidney and Cardiovascular Outcomes: A Post Hoc Analysis from the CREDENCE Trial. J Am Soc Nephrol 31, 2925–2936 (2020). https://doi.org/10.1681/asn.2020050723

283 Otto, S. et al. Microembolization and myonecrosis during elective percutaneous coronary interventions in diabetic patients: an intracoronary Doppler ultrasound study with 2-year clinical follow-up. Basic Res Cardiol 107, 289 (2012). https://doi.org/10.1007/s00395-012-0289-x

284 Pagidipati, N. J. et al. Association of obesity with cardiovascular outcomes in patients with type 2 diabetes and cardiovascular disease: Insights from TECOS. Am Heart J 219, 47–57 (2020). https://doi.org/10.1016/j.ahj.2019.09.016

285 Panero, F. et al. Uric acid is not an independent predictor of cardiovascular mortality in type 2 diabetes: a population-based study. Atherosclerosis 221, 183–188 (2012). https://doi.org/10.1016/j.atherosclerosis.2011.11.042

286 Park, G. M. et al. Coronary computed tomographic angiographic findings in asymptomatic patients with type 2 diabetes mellitus. Am J Cardiol 113, 765–771 (2014). https://doi.org/10.1016/j.amjcard.2013.11.028

287 Peng, W. H. et al. Decreased serum esRAGE level is associated with angiographically determined coronary plaque progression in diabetic patients. Clin Biochem 42, 1252–1259 (2009). https://doi.org/10.1016/j.clinbiochem.2009.04.017

288 Peters, K. E., Chubb, S. A., Davis, W. A. & Davis, T. M. The relationship between hypomagnesemia, metformin therapy and cardiovascular disease complicating type 2 diabetes: the Fremantle Diabetes Study. PLoS One 8, e74355 (2013). https://doi.org/10.1371/journal.pone.0074355

289 Petretta, M. et al. Transient ischemic dilation in patients with diabetes mellitus: prognostic value and effect on clinical outcome after coronary revascularization. Circ Cardiovasc Imaging 6, 908–915 (2013). https://doi.org/10.1161/circimaging.113.000497

290 Pfister, R., Cairns, R., Erdmann, E. & Schneider, C. A. Prognostic impact of electrocardiographic signs in patients with Type 2 diabetes and cardiovascular disease: results from the PROactive study. Diabet Med 28, 1206–1212 (2011). https://doi.org/10.1111/j.1464-5491.2011.03281.x

291 Pickup, J. C. & Mattock, M. B. Activation of the innate immune system as a predictor of cardiovascular mortality in Type 2 diabetes mellitus. Diabet Med 20, 723–726 (2003). https://doi.org/10.1046/j.1464-5491.2003.00990.x

292 Piniés, J. A. et al. Development of a prediction model for fatal and non-fatal coronary heart disease and cardiovascular disease in patients with newly diagnosed type 2 diabetes mellitus: the Basque Country Prospective Complications and Mortality Study risk engine (BASCORE). Diabetologia 57, 2324–2333 (2014). https://doi.org/10.1007/s00125-014-3370-1

293 Pintó, X. et al. [Factors predictive of cardiovascular disease in patients with type-2 diabetes and hypercholesterolemia. ESODIAH study]. Rev Esp Cardiol 60, 251–258 (2007).

294 Poon, P. Y., Szeto, C. C., Kwan, B. C., Chow, K. M. & Li, P. K. Relationship between beta1-adrenergic receptor polymorphisms and cardiovascular disease in patients with diabetic nephropathy. Nephrology (Carlton) 15, 242–247 (2010). https://doi.org/10.1111/j.1440-1797.2009.01182.x

295 Poon, P. Y., Szeto, C. C., Kwan, B. C., Chow, K. M. & Li, P. K. Relationship between HSP70-2 A+1267G Polymorphism and Cardiovascular Events of Chinese Peritoneal Dialysis Patients. Nephron Clin Pract 128, 153–158 (2014). https://doi.org/10.1159/000368237

296 Porchay-Baldérelli, I. et al. The CETP TaqIB polymorphism is associated with the risk of sudden death in type 2 diabetic patients. Diabetes Care 30, 2863–2867 (2007). https://doi.org/10.2337/dc07-0869

297 Porchay-Baldérelli, I. et al. Relationships between common polymorphisms of adenosine triphosphate-binding cassette transporter A1 and high-density lipoprotein cholesterol and coronary heart disease in a population with type 2 diabetes mellitus. Metabolism 58, 74–79 (2009). https://doi.org/10.1016/j.metabol.2008.08.009

298 Prentice, J. C., Pizer, S. D. & Conlin, P. R. Identifying the independent effect of HbA1c variability on adverse health outcomes in patients with Type 2 diabetes. Diabetic Medicine 33, 1640–1648 (2016). https://doi.org/10.1111/dme.13166

299 Qi, L. et al. The +276 polymorphism of the APM1 gene, plasma adiponectin concentration, and cardiovascular risk in diabetic men. Diabetes 54, 1607–1610 (2005). https://doi.org/10.2337/diabetes.54.5.1607

300 Qi, L. et al. Genetic susceptibility to coronary heart disease in type 2 diabetes: 3 independent studies. J Am Coll Cardiol 58, 2675–2682 (2011). https://doi.org/10.1016/j.jacc.2011.08.054

301 Qi, L. et al. Association between a genetic variant related to glutamic acid metabolism and coronary heart disease in individuals with type 2 diabetes. JAMA 310, 821–828 (2013). https://doi.org/10.1001/jama.2013.276305

302 Qi, Q., Workalemahu, T., Zhang, C., Hu, F. B. & Qi, L. Genetic variants, plasma lipoprotein(a) levels, and risk of cardiovascular morbidity and mortality among two prospective cohorts of type 2 diabetes. Eur Heart J 33, 325–334 (2012). https://doi.org/10.1093/eurheartj/ehr350

303 Qin, Z. et al. The atherogenic index of plasma plays an important role in predicting the prognosis of type 2 diabetic subjects undergoing percutaneous coronary intervention: results from an observational cohort study in China. Cardiovasc Diabetol 19, 23 (2020). https://doi.org/10.1186/s12933-020-0989-8

304 Quan, J. et al. Risk Prediction Scores for Mortality, Cerebrovascular, and Heart Disease Among Chinese People With Type 2 Diabetes. J Clin Endocrinol Metab 104, 5823–5830 (2019). https://doi.org/10.1210/jc.2019-00731

305 Raghavan, S. et al. Diabetes Mellitus-Related All-Cause and Cardiovascular Mortality in a National Cohort of Adults. J Am Heart Assoc 8, e011295 (2019). https://doi.org/10.1161/jaha.118.011295

306 Ramírez-Prado, D. et al. A four-year cardiovascular risk score for type 2 diabetic inpatients. PeerJ 2015 (2015). https://doi.org/10.7717/peerj.984

307 Rana, B. S. et al. QT interval abnormalities are often present at diagnosis in diabetes and are better predictors of cardiac death than ankle brachial pressure index and autonomic function tests. Heart 91, 44–50 (2005). https://doi.org/10.1136/hrt.2003.017632

308 Rasmussen, D. G. K. et al. Higher Collagen VI Formation Is Associated With All-Cause Mortality in Patients With Type 2 Diabetes and Microalbuminuria. Diabetes Care 41, 1493–1500 (2018). https://doi.org/10.2337/dc17-2392

309 Ravassa, S. et al. Association of low GLP-1 with oxidative stress is related to cardiac disease and outcome in patients with type 2 diabetes mellitus: a pilot study. Free Radic Biol Med 81, 1–12 (2015). https://doi.org/10.1016/j.freeradbiomed.2015.01.002

310 Rawshani, A. et al. Risk Factors, Mortality, and Cardiovascular Outcomes in Patients with Type 2 Diabetes. N Engl J Med 379, 633–644 (2018). https://doi.org/10.1056/NEJMoa1800256

311 Rawshani, A. et al. Association Between Socioeconomic Status and Mortality, Cardiovascular Disease, and Cancer in Patients With Type 2 Diabetes. JAMA Intern Med 176, 1146–1154 (2016). https://doi.org/10.1001/jamainternmed.2016.2940

312 Read, S. H. et al. Performance of Cardiovascular Disease Risk Scores in People Diagnosed With Type 2 Diabetes: External Validation Using Data From the National Scottish Diabetes Register. Diabetes Care 41, 2010–2018 (2018). https://doi.org/10.2337/dc18-0578

313 Reinhard, H. et al. Osteoprotegerin and mortality in type 2 diabetic patients. Diabetes Care 33, 2561–2566 (2010). https://doi.org/10.2337/dc10-0858

314 Resl, M. et al. Serum uric acid is related to cardiovascular events and correlates with N-terminal pro-B-type natriuretic peptide and albuminuria in patients with diabetes mellitus. Diabet Med 29, 721–725 (2012). https://doi.org/10.1111/j.1464-5491.2011.03515.x

315 Resl, M. et al. Targeted multiple biomarker approach in predicting cardiovascular events in patients with diabetes. Heart 102, 1963–1968 (2016). https://doi.org/10.1136/heartjnl-2015-308949

316 Rossi, M. C. et al. Quality of diabetes care predicts the development of cardiovascular events: results of the AMD-QUASAR study. Diabetes Care 34, 347–352 (2011). https://doi.org/10.2337/dc10-1709

317 Rotbain Curovic, V., et al. Urinary tubular biomarkers as predictors of kidney function decline, cardiovascular events and mortality in microalbuminuric type 2 diabetic patients. Acta Diabetol 55, 1143–1150 (2018). https://doi.org/10.1007/s00592-018-1205-0

318 Roumeliotis, A. et al. Carotid intima-media thickness is an independent predictor of all-cause mortality and cardiovascular morbidity in patients with diabetes mellitus type 2 and chronic kidney disease. Ren Fail 41, 131–138 (2019). https://doi.org/10.1080/0886022x.2019.1585372

319 Roumeliotis, A. K. et al. Association of ALOX12 gene polymorphism with all-cause and cardiovascular mortality in diabetic nephropathy. Int Urol Nephrol 50, 321–329 (2018). https://doi.org/10.1007/s11255-017-1755-z

320 Roumeliotis, S. et al. Matrix Gla protein T-138C polymorphism is associated with carotid intima media thickness and predicts mortality in patients with diabetic nephropathy. J Diabetes Complications 31, 1527–1532 (2017). https://doi.org/10.1016/j.jdiacomp.2017.06.012

321 Rozing, M. P. et al. Changes in HbA1c during the first six years after the diagnosis of Type 2 diabetes mellitus predict long-term microvascular outcomes. PLoS One 14, e0225230 (2019). https://doi.org/10.1371/journal.pone.0225230

322 Ruggenenti, P. et al. Measurable urinary albumin predicts cardiovascular risk among normoalbuminuric patients with type 2 diabetes. J Am Soc Nephrol 23, 1717–1724 (2012). https://doi.org/10.1681/asn.2012030252

323 Russo, G. T. et al. Mild hyperhomocysteinemia, C677T polymorphism on methylenetetrahydrofolate reductase gene and the risk of macroangiopathy in type 2 diabetes: a prospective study. Acta Diabetol 48, 95–101 (2011). https://doi.org/10.1007/s00592-009-0169-5

324 Rutter, M. K., Wahid, S. T., McComb, J. M. & Marshall, S. M. Significance of silent ischemia and microalbuminuria in predicting coronary events in asymptomatic patients with type 2 diabetes. J Am Coll Cardiol 40, 56–61 (2002). https://doi.org/10.1016/s0735-1097(02)01910-1

325 Rådholm, K. et al. The impact of using sagittal abdominal diameter to predict major cardiovascular events in European patients with type 2 diabetes. Nutr Metab Cardiovasc Dis 27, 418–422 (2017). https://doi.org/10.1016/j.numecd.2017.02.001

326 Rørth, R. et al. The prognostic value of troponin T and N-terminal pro B-type natriuretic peptide, alone and in combination, in heart failure patients with and without diabetes. Eur J Heart Fail 21, 40–49 (2019). https://doi.org/10.1002/ejhf.1359

327 Saely, C. H. et al. The metabolic syndrome, insulin resistance, and cardiovascular risk in diabetic and nondiabetic patients. J Clin Endocrinol Metab 90, 5698–5703 (2005). https://doi.org/10.1210/jc.2005-0799

328 Saely, C. H. et al. Lipoprotein(a), type 2 diabetes and vascular risk in coronary patients. Eur J Clin Invest 36, 91–97 (2006). https://doi.org/10.1111/j.1365-2362.2006.01604.x

329 Saito, I. et al. Nontraditional risk factors for coronary heart disease incidence among persons with diabetes: the Atherosclerosis Risk in Communities (ARIC) Study. Ann Intern Med 133, 81–91 (2000). https://doi.org/10.7326/0003-4819-133-2-200007180-00007

330 Sakai, K. et al. Small dense low-density lipoprotein cholesterol is a promising biomarker for secondary prevention in older men with stable coronary artery disease. Geriatr Gerontol Int 18, 965–972 (2018). https://doi.org/10.1111/ggi.13287

331 Salles, G. F., Leite, N. C., Pereira, B. B., Nascimento, E. M. & Cardoso, C. R. Prognostic impact of clinic and ambulatory blood pressure components in high-risk type 2 diabetic patients: the Rio de Janeiro Type 2 Diabetes Cohort Study. J Hypertens 31, 2176–2186 (2013). https://doi.org/10.1097/HJH.0b013e328364103f

332 Satirapoj, B., Tasanavipas, P. & Supasyndh, O. Role of TCF7L2 and PPARG2 Gene Polymorphisms in Renal and Cardiovascular Complications among Patients with Type 2 Diabetes: A Cohort Study. Kidney Diseases 5, 220–227 (2019). https://doi.org/10.1159/000497100

333 Saulnier, P. J. et al. Urinary Sodium Concentration Is an Independent Predictor of All-Cause and Cardiovascular Mortality in a Type 2 Diabetes Cohort Population. J Diabetes Res 2017, 5327352 (2017). https://doi.org/10.1155/2017/5327352

334 Savonitto, S. et al. Predictors of mortality in hospital survivors with type 2 diabetes mellitus and acute coronary syndromes. Diab Vasc Dis Res 15, 14–23 (2018). https://doi.org/10.1177/1479164117735493

335 Schimke, K., Chubb, S. A. P., Davis, W. A. & Davis, T. M. E. Helicobacter pylori cytotoxin-associated gene-A antibodies do not predict complications or death in type 2 diabetes: The Fremantle Diabetes Study. Atherosclerosis 212, 321–326 (2010). https://doi.org/10.1016/j.atherosclerosis.2010.05.021

336 Schulze, M. B. et al. C-reactive protein and incident cardiovascular events among men with diabetes. Diabetes Care 27, 889–894 (2004). https://doi.org/10.2337/diacare.27.4.889

337 Schulze, M. B. et al. Joint role of non-HDL cholesterol and glycated haemoglobin in predicting future coronary heart disease events among women with type 2 diabetes. Diabetologia 47, 2129–2136 (2004). https://doi.org/10.1007/s00125-004-1593-2

338 Scirica, B. M. et al. Prognostic implications of biomarker assessments in patients with type 2 diabetes at high cardiovascular risk: A secondary analysis of a randomized clinical trial. JAMA Cardiology 1, 989–998 (2016). https://doi.org/10.1001/jamacardio.2016.3030

339 Scirica, B. M. et al. Cardiovascular Outcomes According to Urinary Albumin and Kidney Disease in Patients With Type 2 Diabetes at High Cardiovascular Risk: Observations From the SAVOR-TIMI 53 Trial. JAMA Cardiol 3, 155–163 (2018). https://doi.org/10.1001/jamacardio.2017.4228

340 Seferovic, J. P. et al. Retinopathy, Neuropathy, and Subsequent Cardiovascular Events in Patients with Type 2 Diabetes and Acute Coronary Syndrome in the ELIXA: The Importance of Disease Duration. J Diabetes Res 2018, 1631263 (2018). https://doi.org/10.1155/2018/1631263

341 Seyoum, B., Estacio, R. O., Berhanu, P. & Schrier, R. W. Exercise capacity is a predictor of cardiovascular events in patients with type 2 diabetes mellitus. Diab Vasc Dis Res 3, 197–201 (2006). https://doi.org/10.3132/dvdr.2006.030

342 Shao, H., Fonseca, V., Stoecker, C., Liu, S. & Shi, L. Novel Risk Engine for Diabetes Progression and Mortality in USA: Building, Relating, Assessing, and Validating Outcomes (BRAVO). Pharmacoeconomics 36, 1125–1134 (2018). https://doi.org/10.1007/s40273-018-0662-1

343 Shao, H., Shi, L. & Fonseca, V. A. Using the BRAVO Risk Engine to Predict Cardiovascular Outcomes in Clinical Trials With Sodium-Glucose Transporter 2 Inhibitors. Diabetes Care 43, 1530–1536 (2020). https://doi.org/10.2337/dc20-0227

344 Sharma, A. et al. Causes of Death in a Contemporary Cohort of Patients With Type 2 Diabetes and Atherosclerotic Cardiovascular Disease: Insights From the TECOS Trial. Diabetes Care 40, 1763–1770 (2017). https://doi.org/10.2337/dc17-1091

345 Sharma, A., et al. Clinical and Biomarker Predictors of Expanded Heart Failure Outcomes in Patients With Type 2 Diabetes Mellitus After a Recent Acute Coronary Syndrome: Insights From the EXAMINE Trial. J Am Heart Assoc 9, e012797 (2020). https://doi.org/10.1161/jaha.119.012797

346 Shin, S. H. et al. Hyperglycaemia, ejection fraction and the risk of heart failure or cardiovascular death in patients with type 2 diabetes and a recent acute coronary syndrome. Eur J Heart Fail 22, 1133–1143 (2020). https://doi.org/10.1002/ejhf.1790

347 Siddique, A. et al. Relationship of mildly increased albuminuria and coronary artery revascularization outcomes in patients with diabetes. Catheter Cardiovasc Interv 93, E217–e224 (2019). https://doi.org/10.1002/ccd.27890

348 Sidorenkov, G., Voorham, J., de Zeeuw, D., Haaijer-Ruskamp, F. M. & Denig, P. Do treatment quality indicators predict cardiovascular outcomes in patients with diabetes? PLoS One 8, e78821 (2013). https://doi.org/10.1371/journal.pone.0078821

349 Silva, A. P. et al. Phosphorus as an early marker of morbidity and mortality in type 2 chronic kidney disease diabetic patients. J Diabetes Complications 27, 328–332 (2013). https://doi.org/10.1016/j.jdiacomp.2013.02.007

350 Silva, A. P. et al. What is the role of apelin regarding cardiovascular risk and progression of renal disease in type 2 diabetic patients with diabetic nephropathy? Biomed Res Int 2013, 247649 (2013). https://doi.org/10.1155/2013/247649

351 Simmons, R. K. et al. Performance of the UK Prospective Diabetes Study Risk Engine and the Framingham Risk Equations in Estimating Cardiovascular Disease in the EPIC-Norfolk Cohort. Diabetes Care 32, 708–713 (2009). https://doi.org/10.2337/dc08-1918

352 Smáradóttir, M. I. et al. Copeptin and insulin-like growth factor binding protein-1 during follow-up after an acute myocardial infarction in patients with type 2 diabetes: A report from the Diabetes Mellitus Insulin-Glucose Infusion in Acute Myocardial Infarction 2 cohort. Diab Vasc Dis Res 16, 22–27 (2019). https://doi.org/10.1177/1479164118804451

353 So, W. Y. et al. Aldose reductase genotypes and cardiorenal complications: an 8-year prospective analysis of 1,074 type 2 diabetic patients. Diabetes Care 31, 2148–2153 (2008). https://doi.org/10.2337/dc08-0712

354 Soejima, H. et al. Proteinuria is independently associated with the incidence of primary cardiovascular events in diabetic patients. J Cardiol 75, 387–393 (2020). https://doi.org/10.1016/j.jjcc.2019.08.021

355 Soinio, M., Marniemi, J., Laakso, M., Lehto, S. & Rönnemaa, T. Elevated plasma homocysteine level is an independent predictor of coronary heart disease events in patients with type 2 diabetes mellitus. Ann Intern Med 140, 94–100 (2004). https://doi.org/10.7326/0003-4819-140-2-200401200-00009

356 Sone, H. et al. Waist circumference as a cardiovascular and metabolic risk in Japanese patients with type 2 diabetes. Obesity (Silver Spring) 17, 585–592 (2009). https://doi.org/10.1038/oby.2008.481

357 Sone, H. et al. Comparison of various lipid variables as predictors of coronary heart disease in Japanese men and women with type 2 diabetes: subanalysis of the Japan Diabetes Complications Study. Diabetes Care 35, 1150–1157 (2012). https://doi.org/10.2337/dc11-1412

358 Sone, H. et al. Serum level of triglycerides is a potent risk factor comparable to LDL cholesterol for coronary heart disease in Japanese patients with type 2 diabetes: subanalysis of the Japan Diabetes Complications Study (JDCS). J Clin Endocrinol Metab 96, 3448–3456 (2011). https://doi.org/10.1210/jc.2011-0622

359 Sone, H. et al. Leisure-time physical activity is a significant predictor of stroke and total mortality in Japanese patients with type 2 diabetes: analysis from the Japan Diabetes Complications Study (JDCS). Diabetologia 56, 1021–1030 (2013). https://doi.org/10.1007/s00125-012-2810-z

360 Spoelstra-de Man, A. M., Brouwer, C. B., Stehouwer, C. D. & Smulders, Y. M. Rapid progression of albumin excretion is an independent predictor of cardiovascular mortality in patients with type 2 diabetes and microalbuminuria. Diabetes Care 24, 2097–2101 (2001). https://doi.org/10.2337/diacare.24.12.2097

361 Standl, E. et al. Predictors of 10-year macrovascular and overall mortality in patients with NIDDM: the Munich General Practitioner Project. Diabetologia 39, 1540–1545 (1996). https://doi.org/10.1007/s001250050612

362 Stehouwer, C. D., Gall, M. A., Hougaard, P., Jakobs, C. & Parving, H. H. Plasma homocysteine concentration predicts mortality in non-insulin-dependent diabetic patients with and without albuminuria. Kidney Int 55, 308–314 (1999). https://doi.org/10.1046/j.1523-1755.1999.00256.x

363 Stevens, R. J., Kothari, V., Adler, A. I. & Stratton, I. M. The UKPDS risk engine: a model for the risk of coronary heart disease in Type II diabetes (UKPDS 56). Clin Sci (Lond) 101, 671–679 (2001).

364 Strojek, K. et al. Factors Associated With Cardiovascular Events in Patients With Type 2 Diabetes and Acute Myocardial Infarction. J Clin Endocrinol Metab 101, 243–253 (2016). https://doi.org/10.1210/jc.2015-1962

365 Sultan, A. et al. Myocardial perfusion imaging and cardiac events in a cohort of asymptomatic patients with diabetes living in southern France. Diabet Med 23, 410–418 (2006). https://doi.org/10.1111/j.1464-5491.2006.01818.x

366 Svendstrup, M., Christiansen, M. S., Magid, E., Hommel, E. & Feldt-Rasmussen, B. Increased orosomucoid in urine is an independent predictor of cardiovascular and all-cause mortality in patients with type 2 diabetes at 10 years of follow-up. J Diabetes Complications 27, 570–575 (2013). https://doi.org/10.1016/j.jdiacomp.2013.05.008

367 Svensson, E. et al. Early Glycemic Control and Magnitude of HbA(1c) Reduction Predict Cardiovascular Events and Mortality: Population-Based Cohort Study of 24,752 Metformin Initiators. Diabetes Care 40, 800–807 (2017). https://doi.org/10.2337/dc16-2271

368 Svensson, M. K., Cederholm, J., Eliasson, B., Zethelius, B. & Gudbjörnsdottir, S. Albuminuria and renal function as predictors of cardiovascular events and mortality in a general population of patients with type 2 diabetes: a nationwide observational study from the Swedish National Diabetes Register. Diab Vasc Dis Res 10, 520–529 (2013). https://doi.org/10.1177/1479164113500798

369 Takao, T., Suka, M., Yanagisawa, H. & Iwamoto, Y. Impact of postprandial hyperglycemia at clinic visits on the incidence of cardiovascular events and all-cause mortality in patients with type 2 diabetes. J Diabetes Investig 8, 600–608 (2017). https://doi.org/10.1111/jdi.12610

370 Tan, X. & Benedict, C. Increased risk of myocardial infarction among patients with type 2 diabetes who carry the common rs10830963 variant in the MTNR1B gene. Diabetes Care 43, 2289–2292 (2020). https://doi.org/10.2337/dc20-0507

371 Tanaka, S. et al. Predicting macro- and microvascular complications in type 2 diabetes: the Japan Diabetes Complications Study/the Japanese Elderly Diabetes Intervention Trial risk engine. Diabetes Care 36, 1193–1199 (2013). https://doi.org/10.2337/dc12-0958

372 Thanyasiri, P., Celermajer, D. S. & Adams, M. R. Predictors of long-term outcome following percutaneous coronary intervention in patients with type 2 diabetes mellitus. Coron Artery Dis 17, 131–138 (2006). https://doi.org/10.1097/00019501-200603000-00006

373 The Look, A. R. G. Prospective association of a genetic risk score and lifestyle intervention with cardiovascular morbidity and mortality among individuals with type 2 diabetes: the Look AHEAD randomised controlled trial. Diabetologia 58, 1803–1813 (2015). https://doi.org/10.1007/s00125-015-3610-z

374 Theilade, S. et al. Pulse pressure is not an independent predictor of outcome in type 2 diabetes patients with chronic kidney disease and anemia--the Trial to Reduce Cardiovascular Events with Aranesp Therapy (TREAT). J Hum Hypertens 30, 46–52 (2016). https://doi.org/10.1038/jhh.2015.22

375 Thomas, M. C. et al. Relationship Between Plasma 8-OH-Deoxyguanosine and Cardiovascular Disease and Survival in Type 2 Diabetes Mellitus: Results From the ADVANCE Trial. J Am Heart Assoc 7 (2018). https://doi.org/10.1161/jaha.117.008226

376 Tian, P., Zheng, X., Li, M., Li, W. & Niu, Q. Long-term prognostic value of coronary computed tomography angiography for asymptomatic patients with cad in type 2 diabetes mellitus. Experimental and Therapeutic Medicine 18, 747–754 (2019). https://doi.org/10.3892/etm.2019.7593

377 Ting, R. Z. et al. Lipid control and use of lipid-regulating drugs for prevention of cardiovascular events in Chinese type 2 diabetic patients: a prospective cohort study. Cardiovasc Diabetol 9, 77 (2010). https://doi.org/10.1186/1475-2840-9-77

378 Tobias, D. K. et al. Circulating Branched-Chain Amino Acids and Incident Cardiovascular Disease in a Prospective Cohort of US Women. Circ Genom Precis Med 11, e002157 (2018). https://doi.org/10.1161/circgen.118.002157

379 Turner, R. C. et al. Risk factors for coronary artery disease in non-insulin dependent diabetes mellitus: United Kingdom Prospective Diabetes Study (UKPDS: 23). Bmj 316, 823–828 (1998). https://doi.org/10.1136/bmj.316.7134.823

380 Twito, O. et al. New-onset diabetes in elderly subjects: association between HbA1c levels, mortality, and coronary revascularization. Diabetes Care 36, 3425–3429 (2013). https://doi.org/10.2337/dc12-2503

381 Umamahesh, K., Vigneswari, A., Surya Thejaswi, G., Satyavani, K. & Viswanathan, V. Incidence of cardiovascular diseases and associated risk factors among subjects with type 2 diabetes - an 11-year follow up study. Indian Heart J 66, 5–10 (2014). https://doi.org/10.1016/j.ihj.2013.12.009

382 Valmadrid, C. T., Klein, R., Moss, S. E. & Klein, B. E. The risk of cardiovascular disease mortality associated with microalbuminuria and gross proteinuria in persons with older-onset diabetes mellitus. Arch Intern Med 160, 1093–1100 (2000). https://doi.org/10.1001/archinte.160.8.1093

383 Valoti, E. et al. Impact of a Complement Factor H Gene Variant on Renal Dysfunction, Cardiovascular Events, and Response to ACE Inhibitor Therapy in Type 2 Diabetes. Front Genet 10, 681 (2019). https://doi.org/10.3389/fgene.2019.00681

384 van der Heijden, A. A., Ortegon, M. M., Niessen, L. W., Nijpels, G. & Dekker, J. M. Prediction of coronary heart disease risk in a general, pre-diabetic, and diabetic population during 10 years of follow-up: accuracy of the Framingham, SCORE, and UKPDS risk functions: The Hoorn Study. Diabetes Care 32, 2094–2098 (2009). https://doi.org/10.2337/dc09-0745

385 van der Leeuw, J. et al. Novel Biomarkers to Improve the Prediction of Cardiovascular Event Risk in Type 2 Diabetes Mellitus. J Am Heart Assoc 5 (2016). https://doi.org/10.1161/JAHA.115.003048

386 van der Leeuw, J. et al. The validation of cardiovascular risk scores for patients with type 2 diabetes mellitus. Heart 101, 222–229 (2015). https://doi.org/10.1136/heartjnl-2014-306068

387 van Dieren, S. et al. External validation of the UK Prospective Diabetes Study (UKPDS) risk engine in patients with type 2 diabetes. Diabetologia 54, 264–270 (2011). https://doi.org/10.1007/s00125-010-1960-0

388 Vanzetto, G. et al. Prediction of cardiovascular events in clinically selected high-risk NIDDM patients. Prognostic value of exercise stress test and thallium-201 single-photon emission computed tomography. Diabetes Care 22, 19–26 (1999). https://doi.org/10.2337/diacare.22.1.19

389 Vavruch, C. et al. Using proximity extension proteomics assay to discover novel biomarkers associated with circulating leptin levels in patients with type 2 diabetes. Sci Rep 10, 13097 (2020). https://doi.org/10.1038/s41598-020-69473-2

390 Velho, G. et al. Plasma copeptin, kidney disease, and risk for cardiovascular morbidity and mortality in two cohorts of type 2 diabetes. Cardiovasc Diabetol 17, 110 (2018). https://doi.org/10.1186/s12933-018-0753-5

391 Vengen, I. T., Dale, A. C., Wiseth, R., Midthjell, K. & Videm, V. Lactoferrin is a novel predictor of fatal ischemic heart disease in diabetes mellitus type 2: long-term follow-up of the HUNT 1 study. Atherosclerosis 212, 614–620 (2010). https://doi.org/10.1016/j.atherosclerosis.2010.06.008

392 Venskutonyte, L., Brismar, K., Öhrvik, J., Rydén, L. & Kjellström, B. Self-rated health predicts outcome in patients with type 2 diabetes and myocardial infarction: a DIGAMI 2 quality of life sub-study. Diab Vasc Dis Res 10, 361–367 (2013). https://doi.org/10.1177/1479164113482694

393 Venskutonyte, L., Malmberg, K., Norhammar, A., Wedel, H. & Rydén, L. Effect of gender on prognosis in patients with myocardial infarction and type 2 diabetes. J Intern Med 268, 75–82 (2010). https://doi.org/10.1111/j.1365-2796.2010.02215.x

394 Venuraju, S. M. et al. Duration of type 2 diabetes mellitus and systolic blood pressure as determinants of severity of coronary stenosis and adverse events in an asymptomatic diabetic population: PROCEED study. Cardiovasc Diabetol 18, 51 (2019). https://doi.org/10.1186/s12933-019-0855-8

395 Venuraju, S. M. et al. Association of Epicardial Fat Volume With the Extent of Coronary Atherosclerosis and Cardiovascular Adverse Events in Asymptomatic Patients With Diabetes. Angiology 72, 442–450 (2021). https://doi.org/10.1177/0003319720984607

396 Venäläinen, M. S., Klén, R., Mahmoudian, M., Raitakari, O. T. & Elo, L. L. Easy-to-use tool for evaluating the elevated acute kidney injury risk against reduced cardiovascular disease risk during intensive blood pressure control. J Hypertens 38, 511–518 (2020). https://doi.org/10.1097/hjh.0000000000002282

397 Vepsäläinen, T. et al. Proteinuria modifies the effect of systolic blood pressure on total and cardiovascular disease mortality in patients with type 2 diabetes. J Intern Med 272, 611–619 (2012). https://doi.org/10.1111/j.1365-2796.2012.02581.x

398 von Scholten, B. J. et al. Additive prognostic value of plasma N-terminal pro-brain natriuretic peptide and coronary artery calcification for cardiovascular events and mortality in asymptomatic patients with type 2 diabetes. Cardiovasc Diabetol 14, 59 (2015). https://doi.org/10.1186/s12933-015-0225-0

399 von Scholten, B. J. et al. Urinary biomarkers are associated with incident cardiovascular disease, all-cause mortality and deterioration of kidney function in type 2 diabetic patients with microalbuminuria. Diabetologia 59, 1549–1557 (2016). https://doi.org/10.1007/s00125-016-3937-0

400 von Scholten, B. J. et al. Markers of inflammation and endothelial dysfunction are associated with incident cardiovascular disease, all-cause mortality, and progression of coronary calcification in type 2 diabetic patients with microalbuminuria. J Diabetes Complications 30, 248–255 (2016). https://doi.org/10.1016/j.jdiacomp.2015.11.005

401 Wallander, M. et al. IGF binding protein 1 predicts cardiovascular morbidity and mortality in patients with acute myocardial infarction and type 2 diabetes. Diabetes Care 30, 2343–2348 (2007). https://doi.org/10.2337/dc07-0825

402 Wan, E. Y., Fong, D. Y., Fung, C. S. & Lam, C. L. Incidence and predictors for cardiovascular disease in Chinese patients with type 2 diabetes mellitus - a population-based retrospective cohort study. J Diabetes Complications 30, 444–450 (2016). https://doi.org/10.1016/j.jdiacomp.2015.12.010

403 Wan, E. Y. F. et al. Development of a cardiovascular diseases risk prediction model and tools for Chinese patients with type 2 diabetes mellitus: A population-based retrospective cohort study. Diabetes Obes Metab 20, 309–318 (2018). https://doi.org/10.1111/dom.13066

404 Wang, Y. et al. Independent predictive roles of eotaxin Ala23Thr, paraoxonase 2 Ser311Cys and beta-adrenergic receptor Trp64Arg polymorphisms on cardiac disease in Type 2 Diabetes--an 8-year prospective cohort analysis of 1297 patients. Diabet Med 27, 376–383 (2010). https://doi.org/10.1111/j.1464-5491.2010.02980.x

405 Wang, Y. et al. Prognostic effect of insertion/deletion polymorphism of the ace gene on renal and cardiovascular clinical outcomes in Chinese patients with type 2 diabetes. Diabetes Care 28, 348–354 (2005). https://doi.org/10.2337/diacare.28.2.348

406 Watson, C. et al. Investigation of association of genetic variant rs3918242 of matrix metalloproteinase-9 with hypertension, myocardial infarction and progression of ventricular dysfunction in Irish Caucasian patients with diabetes: a report from the STOP-HF follow-up prog. BMC Cardiovasc Disord 21, 87 (2021). https://doi.org/10.1186/s12872-021-01860-7

407 Wei, M., Gaskill, S. P., Haffner, S. M. & Stern, M. P. Effects of diabetes and level of glycemia on all-cause and cardiovascular mortality. The San Antonio Heart Study. Diabetes Care 21, 1167–1172 (1998). https://doi.org/10.2337/diacare.21.7.1167

408 Wells, B. J. et al. Prediction of morbidity and mortality in patients with type 2 diabetes. PeerJ 2013 (2013). https://doi.org/10.7717/peerj.87

409 Wijkman, M., Länne, T., Östgren, C. J. & Nystrom, F. H. Aortic pulse wave velocity predicts incident cardiovascular events in patients with type 2 diabetes treated in primary care. J Diabetes Complications 30, 1223–1228 (2016). https://doi.org/10.1016/j.jdiacomp.2016.06.008

410 Wijkman, M., Länne, T., Östgren, C. J. & Nystrom, F. H. Diastolic orthostatic hypertension and cardiovascular prognosis in type 2 diabetes: a prospective cohort study. Cardiovasc Diabetol 15, 83 (2016). https://doi.org/10.1186/s12933-016-0399-0

411 Winkler, K. et al. Apolipoprotein E genotype predicts cardiovascular endpoints in dialysis patients with type 2 diabetes mellitus. Atherosclerosis 208, 197–202 (2010). https://doi.org/10.1016/j.atherosclerosis.2009.06.036

412 Wolsk, E. et al. Role of B-Type Natriuretic Peptide and N-Terminal Prohormone BNP as Predictors of Cardiovascular Morbidity and Mortality in Patients With a Recent Coronary Event and Type 2 Diabetes Mellitus. J Am Heart Assoc 6 (2017). https://doi.org/10.1161/jaha.116.004743

413 Wong, Y. K. et al. High-sensitivity troponin I and B-type natriuretic peptide biomarkers for prediction of cardiovascular events in patients with coronary artery disease with and without diabetes mellitus. Cardiovasc Diabetol 18, 171 (2019). https://doi.org/10.1186/s12933-019-0974-2

414 Woodward, M. et al. Prediction of 10-year vascular risk in patients with diabetes: the AD-ON risk score. Diabetes Obes Metab 18, 289–294 (2016). https://doi.org/10.1111/dom.12614

415 Yamada, T., Itoi, T., Kiuchi, Y., Nemoto, M. & Yamashita, S. Proliferative diabetic retinopathy is a predictor of coronary artery disease in Japanese patients with type 2 diabetes. Diabetes Res Clin Pract 96, e4–6 (2012). https://doi.org/10.1016/j.diabres.2011.12.007

416 Yamasaki, Y. et al. Carotid intima-media thickness in Japanese type 2 diabetic subjects: predictors of progression and relationship with incident coronary heart disease. Diabetes Care 23, 1310–1315 (2000). https://doi.org/10.2337/diacare.23.9.1310

417 Yang, F., Ye, J., Pomerantz, K. & Stewart, M. Potential modification of the UKPDS risk engine and evaluation of macrovascular event rates in controlled clinical trials. *Diabetes*, Metabolic Syndrome and Obesity: Targets and Therapy 6, 247–256 (2013). https://doi.org/10.2147/DMSO.S43724

418 Yang, G. R. et al. Association between Neck Circumference and the Occurrence of Cardiovascular Events in Type 2 Diabetes: Beijing Community Diabetes Study 20 (BCDS-20). Biomed Res Int 2019, 4242304 (2019). https://doi.org/10.1155/2019/4242304

419 Yang, S. H. et al. Triglyceride to High-Density Lipoprotein Cholesterol Ratio and Cardiovascular Events in Diabetics With Coronary Artery Disease. Am J Med Sci 354, 117–124 (2017). https://doi.org/10.1016/j.amjms.2017.03.032

420 Yang, S. H. et al. Serum fibrinogen and cardiovascular events in Chinese patients with type 2 diabetes and stable coronary artery disease: a prospective observational study. BMJ Open 7, e015041 (2017). https://doi.org/10.1136/bmjopen-2016-015041

421 Yang, X. et al. Impacts of chronic kidney disease and albuminuria on associations between coronary heart disease and its traditional risk factors in type 2 diabetic patients - the Hong Kong diabetes registry. Cardiovasc Diabetol 6, 37 (2007). https://doi.org/10.1186/1475-2840-6-37

422 Yang, X. et al. Development and validation of a total coronary heart disease risk score in type 2 diabetes mellitus. Am J Cardiol 101, 596–601 (2008). https://doi.org/10.1016/j.amjcard.2007.10.019

423 Yang, Z. K. et al. Elevated glycated albumin and reduced endogenous secretory receptor for advanced glycation endproducts levels in serum predict major adverse cardio-cerebral events in patients with type 2 diabetes and stable coronary artery disease. Int J Cardiol 197, 241–247 (2015). https://doi.org/10.1016/j.ijcard.2015.06.003

424 Yeboah, J. et al. Development of a new diabetes risk prediction tool for incident coronary heart disease events: the Multi-Ethnic Study of Atherosclerosis and the Heinz Nixdorf Recall Study. Atherosclerosis 236, 411–417 (2014). https://doi.org/10.1016/j.atherosclerosis.2014.07.035

425 Yeboah, P., Hsu, F. C., Bertoni, A. G. & Yeboah, J. Body Mass Index, Change in Weight, Body Weight Variability and Outcomes in Type 2 Diabetes Mellitus (from the ACCORD Trial). Am J Cardiol 123, 576–581 (2019). https://doi.org/10.1016/j.amjcard.2018.11.016

426 Yiu, K. H. et al. Predictive value of high-sensitivity troponin-I for future adverse cardiovascular outcome in stable patients with type 2 diabetes mellitus. Cardiovasc Diabetol 13, 63 (2014). https://doi.org/10.1186/1475-2840-13-63

427 Yoshida, M. et al. Combination of the Framingham risk score and carotid intima-media thickness improves the prediction of cardiovascular events in patients with type 2 diabetes. Diabetes Care 35, 178–180 (2012). https://doi.org/10.2337/dc11-1333

428 Yoshimura, T. et al. Low blood flow estimates in lower-leg arteries predict cardiovascular events in Japanese patients with type 2 diabetes with normal ankle-brachial indexes. Diabetes Care 29, 1884–1890 (2006). https://doi.org/10.2337/dc06-0142

429 Young, J. B. et al. Development of predictive risk models for major adverse cardiovascular events among patients with type 2 diabetes mellitus using health insurance claims data. Cardiovasc Diabetol 17, 118 (2018). https://doi.org/10.1186/s12933-018-0759-z

430 Yu, D. et al. Development and External Validation of Risk Scores for Cardiovascular Hospitalization and Rehospitalization in Patients With Diabetes. J Clin Endocrinol Metab 103, 1122–1129 (2018). https://doi.org/10.1210/jc.2017-02293

431 Yu, D. et al. Total/high density lipoprotein cholesterol and cardiovascular disease (re)hospitalization nadir in type 2 diabetes. J Lipid Res 59, 1745–1750 (2018). https://doi.org/10.1194/jlr.P084269

432 Yu, D. et al. Derivation and external validation of a risk prediction algorithm to estimate future risk of cardiovascular death among patients with type 2 diabetes and incident diabetic nephropathy: prospective cohort study. BMJ Open Diabetes Res Care 7, e000735 (2019). https://doi.org/10.1136/bmjdrc-2019-000735

433 Yu, D. & Simmons, D. Relationship between HbA1c and risk of all-cause hospital admissions among people with Type 2 diabetes. Diabetic Medicine 30, 1407–1411 (2013). https://doi.org/10.1111/dme.12235

434 Yu, D. & Simmons, D. Association between blood pressure and risk of cardiovascular hospital admissions among people with type 2 diabetes. Heart 100, 1444–1449 (2014). https://doi.org/10.1136/heartjnl-2013-304799

435 Yun, J. S., Park, Y. M., Cha, S. A., Ahn, Y. B. & Ko, S. H. Progression of cardiovascular autonomic neuropathy and cardiovascular disease in type 2 diabetes. Cardiovasc Diabetol 17, 109 (2018). https://doi.org/10.1186/s12933-018-0752-6

436 Zafrir, B. et al. Low cardiorespiratory fitness and coronary artery calcification: Complementary cardiovascular risk predictors in asymptomatic type 2 diabetics. Atherosclerosis 241, 634–640 (2015). https://doi.org/10.1016/j.atherosclerosis.2015.06.020

437 Zafrir, B. et al. Resting heart rate and measures of effort-related cardiac autonomic dysfunction predict cardiovascular events in asymptomatic type 2 diabetes. Eur J Prev Cardiol 23, 1298–1306 (2016). https://doi.org/10.1177/2047487315624747

438 Zethelius, B. et al. A new model for 5-year risk of cardiovascular disease in type 2 diabetes, from the Swedish National Diabetes Register (NDR). Diabetes Res Clin Pract 93, 276–284 (2011). https://doi.org/10.1016/j.diabres.2011.05.037

439 Zhang, C. et al. Genetic variation in the hepatic lipase gene and the risk of coronary heart disease among US diabetic men: potential interaction with obesity. Diabetologia 49, 1552–1559 (2006). https://doi.org/10.1007/s00125-006-0235-2

440 Zhang, X. L. et al. Improved Framingham Risk Scores of Patients with Type 2 Diabetes Mellitus in the Beijing Community: A 10-Year Prospective Study of the Effects of Multifactorial Interventions on Cardiovascular Risk Factors (The Beijing Communities Diabetes Study 22). Diabetes Therapy 11, 885–903 (2020). https://doi.org/10.1007/s13300-020-00782-5

441 Zhao, Q. et al. Impacts of triglyceride-glucose index on prognosis of patients with type 2 diabetes mellitus and non-ST-segment elevation acute coronary syndrome: results from an observational cohort study in China. Cardiovasc Diabetol 19, 108 (2020). https://doi.org/10.1186/s12933-020-01086-5

442 Zhou, J. B. et al. Prediction of Proliferative Diabetic Retinopathy to Asymptomatic Obstructive Coronary Artery Disease in Chinese Type 2 Diabetes Individuals: An Exploratory Study. Metab Syndr Relat Disord 17, 367–373 (2019). https://doi.org/10.1089/met.2018.0140

443 Zimering, M. B., Anderson, R. J., Ge, L. & Moritz, T. E. Increased plasma basic fibroblast growth factor is associated with coronary heart disease in adult type 2 diabetes mellitus. Metabolism 60, 284–291 (2011). https://doi.org/10.1016/j.metabol.2010.02.003

444 Zimering, M. B., Anderson, R. J., Ge, L., Moritz, T. E. & Duckworth, W. C. Basic fibroblast growth factor predicts cardiovascular disease occurrence in participants from the veterans affairs diabetes trial. Frontiers in Endocrinology 4 (2013). https://doi.org/10.3389/fendo.2013.00183

445 Zobel, E. H. et al. Toe-brachial index as a predictor of cardiovascular disease and all-cause mortality in people with type 2 diabetes and microalbuminuria. Diabetologia 60, 1883–1891 (2017). https://doi.org/10.1007/s00125-017-4344-x

446 Zoppini, G. et al. Usefulness of the triglyceride to high-density lipoprotein cholesterol ratio for predicting mortality risk in type 2 diabetes: role of kidney dysfunction. Atherosclerosis 212, 287–291 (2010). https://doi.org/10.1016/j.atherosclerosis.2010.04.035

447 Østergaard, H. B. et al. Limited benefit of haemoglobin glycation index as risk factor for cardiovascular disease in type 2 diabetes patients. Diabetes Metab 45, 254–260 (2019). https://doi.org/10.1016/j.diabet.2018.04.006

448 Fitipaldi, H. & Franks, P. W. Ethnic, gender and other sociodemographic biases in genome-wide association studies for the most burdensome non-communicable diseases: 2005-2022. Hum Mol Genet 32, 520–532 (2023). https://doi.org/10.1093/hmg/ddac245

449 Misra, S. et al. Temporal trends in emergency admissions for diabetic ketoacidosis in people with diabetes in England before and during the COVID-19 pandemic: a population-based study. The Lancet Diabetes & Endocrinology 9, 671–680 (2021). https://doi.org/https://doi.org/10.1016/S2213-8587(21)00208-4

450 Di Angelantonio, E. et al. B-type natriuretic peptides and cardiovascular risk: systematic review and meta-analysis of 40 prospective studies. Circulation 120, 2177–2187 (2009). https://doi.org/10.1161/circulationaha.109.884866

451 Tam, C. H. T. et al. Identification of a Common Variant for Coronary Heart Disease at PDE1A Contributes to Individualized Treatment Goals and Risk Stratification of Cardiovascular Complications in Chinese Patients With Type 2 Diabetes. Diabetes Care (2023). https://doi.org/10.2337/dc22-2331

452 Chan, J. et al. The Joint Asia Diabetes Evaluation (JADE) Program: a web-based program to translate evidence to clinical practice in Type 2 diabetes. Diabet Med 26, 693–699 (2009). https://doi.org/10.1111/j.1464-5491.2009.02751.x

453 Chan, J. C. N. et al. Effect of a Web-Based Management Guide on Risk Factors in Patients With Type 2 Diabetes and Diabetic Kidney Disease: A JADE Randomized Clinical Trial. JAMA Netw Open 5, e223862 (2022). https://doi.org/10.1001/jamanetworkopen.2022.3862

454 Lim, L. L. et al. Association of technologically assisted integrated care with clinical outcomes in type 2 diabetes in Hong Kong using the prospective JADE Program: A retrospective cohort analysis. PLoS Med 17, e1003367 (2020). https://doi.org/10.1371/journal.pmed.1003367

455 Chan, J. C. et al. Effects of telephone-based peer support in patients with type 2 diabetes mellitus receiving integrated care: a randomized clinical trial. JAMA Intern Med 174, 972–981 (2014). https://doi.org/10.1001/jamainternmed.2014.655

456 Lim, L. L. et al. Effects of a Technology-Assisted Integrated Diabetes Care Program on Cardiometabolic Risk Factors Among Patients With Type 2 Diabetes in the Asia-Pacific Region: The JADE Program Randomized Clinical Trial. JAMA Netw Open 4, e217557 (2021). https://doi.org/10.1001/jamanetworkopen.2021.7557

