## Supplemental Materials for "Precision Prognostics for Cardiovascular Disease in Type 2 Diabetes: A Systematic Review and Meta-analysis"

**Supplemental Text 1.** Search strategy for the systematic review and meta-analysis

Literature search strategy

Pubmed <1990 to 2021 Week 25>

#1. "Diabetes Mellitus, Type 2"[Mesh] OR NIDDM[Title/Abstract] OR T2DM[Title/Abstract] OR T2D[Title/Abstract] OR "non insulin depend*"[Title/Abstract] OR "noninsulin depend*"[Title/Abstract] OR noninsulin-depend*[Title/Abstract] OR non-insulindepend*[Title/Abstract] OR ((type 2[Title/Abstract] OR type II[Title/Abstract] OR

type2[Title/Abstract] OR type II[Title/Abstract] OR Ketosis-Resistant[Title/Abstract]) AND

diabet*[Title/Abstract]) OR ((late[Title/Abstract] OR adult[Title/Abstract] OR stable[Title/Abstract]) AND onset[Title/Abstract] AND diabet*[Title/Abstract])

=215774

#2. prognoses[Title/Abstract] OR prognosis[Title/Abstract] OR predict*[Title/Abstract] OR

nomogram*[Title/Abstract] OR model*[Title/Abstract] OR prognosti*,[Title/Abstract] OR "survival analys*"[Title/Abstract] OR prospective[Title/Abstract] outcome[Title/Abstract] OR progression[Title/Abstract] OR efficacy[Title/Abstract] OR effectiveness[Title/Abstract] OR longitudinal[Title/Abstract] OR "time series"[Title/Abstract]

=2441175

#3. "risk score*"[Title/Abstract] OR "risk assessment*"[Title/Abstract] OR "risk categor*"[Title/Abstract] OR "risk factor*"[Title/Abstract] OR test*[Title/Abstract] OR biomarker*[Title/Abstract] OR assay*[Title/Abstract] OR tool* [Title/Abstract] OR genetic[Title/Abstract] OR genomic[Title/Abstract] OR polymorphism*[Title/Abstract] OR variant*[Title/Abstract] OR algorithm*[Title/Abstract] OR equation*[Title/Abstract] OR precision[Title/Abstract] OR personali*[Title/Abstract]

=7187775

#4. #2 AND #3

=843991

#5. #2 OR #3

=8773559

#6. ("Atherosclerosis"[Mesh]) OR (((((((("Cardiovascular Diseases"[Mesh]) OR "Myocardial Infarction"[Mesh]) OR "Myocardial Ischemia"[Mesh]) OR "Acute Coronary Syndrome"[Mesh]) OR "Coronary Artery Disease"[Mesh]) OR "Coronary Artery Bypass"[Mesh]) OR "Angioplasty"[Mesh]) OR "Coronary Occlusion"[Mesh]) OR "Percutaneous Coronary Intervention"[Mesh])

=2521345

#7. "cardiovascular disease*"[Title/Abstract] OR "myocardial infarct*"[Title/Abstract] OR "heart infarct*"[Title/Abstract] OR "heart attack*"[Title/Abstract] OR "heart disease*"[Title/Abstract] OR "atheroscleros*"[Title/Abstract] OR "atherosclerotic"[Title/Abstract] OR "myocardial ischemia"[Title/Abstract] OR "myocardial ischaemia"[Title/Abstract] OR "myocardial revascularisation"[Title/Abstract] OR "acute coronary syndrome*"[Title/Abstract] OR "coronary heart disease*"[Title/Abstract] OR "coronary artery disease*"[Title/Abstract] OR "coronary disease*"[Title/Abstract] OR "ischemic heart disease*"[Title/Abstract] OR "ischaemic heart disease*"[Title/Abstract] OR ASCVD[Title/Abstract] OR IHD[Title/Abstract] OR AMI[Title/Abstract] OR

ACS[Title/Abstract] OR CHD[Title/Abstract] OR CAD[Title/Abstract] OR angioplast*[Title/Abstract] OR "coronary artery bypass*"[Title/Abstract] OR "chronic total occlusion*"[Title/Abstract] OR "chronic total coronary occlusion*"[Title/Abstract] OR CTO[Title/Abstract] OR "Percutaneous Coronary Intervention*"[Title/Abstract] OR "Percutaneous Coronary Revascularization"[Title/Abstract] OR "Percutaneous Coronary Revascularisation"[Title/Abstract] OR "Percutaneous coronary angioplasty"[Title/Abstract] OR "PCI"[Title/Abstract] OR "major adverse cardiovascular event"[Title/Abstract] OR "major adverse cardiac events"[Title/Abstract] OR MACE[Title/Abstract]

=851592

#8. #6 OR #7

=2767282

#9. #1 AND #4 AND #8

=4851

#10. #9 Filters, Humans, 1990-2021

=4192

#11. #1 AND #5 AND #8

=32616

#12. #11 Filters, Humans, 1990-2021

=28170

References article: 12502653

Retrieved in #12, not in #10

#13. prognoses[Title/Abstract] OR prognosis[Title/Abstract] OR predict*[Title/Abstract] OR

nomogram*[Title/Abstract] OR model*[Title/Abstract] OR prognosti*,[Title/Abstract] OR "survival analys*"[Title/Abstract] OR prospective[Title/Abstract] outcome[Title/Abstract] OR progression[Title/Abstract] OR efficacy[Title/Abstract] OR effectiveness[Title/Abstract] OR longitudinal[Title/Abstract] OR "time series"[Title/Abstract] OR predictor[Title/Abstract] OR predictors[Title/Abstract]

=2722451

#14. #3 AND #13

=972873

#15. #1 AND #8 AND #14 Filters, Humans, From 1990-2021

=5689 references

#### Embase <1990 to 2021 Week 26>

#1. 'non insulin dependent diabetes mellitus'/exp OR niddm:ab,ti OR t2dm:ab,ti OR t2d:ab,ti OR 'noninsulin dependent':ab,ti OR 'non insulin dependent':ab,ti OR 'type 2':ab,ti OR 'type ii':ab,ti OR type2:ab,ti OR typeii:ab,ti OR 'ketosis resistant':ab,ti OR (diabet*:ab,ti AND onset:ab,ti AND (late:ab,ti OR adult:ab,ti OR stabl*:ab,ti))

526627

#2. (biomarker*:ab,ti OR test*:ab,ti OR assay*:ab,ti OR tool*:ab,ti OR genetic:ab,ti OR genomic:ab,ti OR polymorphism*:ab,ti OR variant*:ab,ti OR 'genetic risk score*':ab,ti OR 'polygenic risk score*':ab,ti) AND (prognos*:ab,ti OR predict*:ab,ti OR model*:ab,ti OR nomogram*:ab,ti OR 'survival analysis':ab,ti OR efficacy:ab,ti OR effectiveness:ab,ti OR prospective:ab,ti OR longitudinal:ab,ti OR 'time series':ab,ti OR statistical:ab,ti OR score*:ab,ti OR risk*:ab,ti OR 'risk score':ab,ti OR 'risk categor*':ab,ti OR 'risk factor*':ab,ti OR 'risk assessment':ab,ti OR algorithm*:ab,ti OR equation*:ab,ti OR precision:ab,ti OR personali*:ab,ti OR outcome*:ab,ti OR progression:ab,ti)

=4,101,858

#3. 'cardiovascular disease'/exp OR 'heart infarction'/exp OR 'heart disease'/exp OR

'atherosclerosis'/exp OR 'heart muscle ischemia'/exp OR 'heart muscle revascularization'/exp OR 'acute coronary syndrome'/exp OR 'ischemic heart disease'/exp OR 'angioplasty'/exp OR 'coronary artery bypass graft'/exp OR 'chronic total occlusion'/exp OR 'percutaneous coronary intervention'/exp OR 'percutaneous coronary revascularization'/exp

=4704886

#4. 'cardiovascular disease':ab,ti OR 'myocardial infarction':ab,ti OR 'heart infarction':ab,ti OR 'heart attack':ab,ti OR 'heart disease':ab,ti OR atherosclerosis:ab,ti OR atherosclerotic:ab,ti OR 'myocardial ischemia':ab,ti OR 'myocardial revasculari*':ab,ti OR 'acute coronary syndrome':ab,ti OR 'coronary heart disease':ab,ti OR 'coronary artery disease':ab,ti OR 'coronary disease':ab,ti OR 'ischemic heart disease':ab,ti OR ascvd:ab,ti OR ihd:ab,ti OR ami:ab,ti OR acs:ab,ti OR chd:ab,ti OR cad:ab,ti OR angioplasty:ab,ti OR 'coronary artery bypass':ab,ti OR 'chronic total occlusion':ab,ti OR 'chronic total coronary occlusion':ab,ti OR cto:ab,ti OR 'percutaneous coronary intervention':ab,ti OR 'percutaneous coronary revasculari*':ab,ti OR 'percutaneous coronary angioplasty':ab,ti OR pci:ab,ti OR 'major adverse cardiovascular events':ab,ti OR 'major adverse cardiac events':ab,ti OR mace:ab,ti

=1111566

#5. #3 OR #4

=4,841,054

#6. #1 AND #2 AND #5

=29,072

AND [embase]/lim NOT ([embase]/lim AND [medline]/lim)

=14567

NOT 'conference abstract':it

=3783

NOT (animal* NOT human*)

=3650

Publication date limitation 1990-present

3643 references

**Supplemental Text 2.** Criteria used for data analyses from the included studies.

The objective of this study was to identify prognostic factors that may refine CVD risk prediction beyond already known risk factors. Therefore, we excluded from the analysis those studies that evaluated biomarkers already established as CVD risk factors (Supplemental Table 3), such as smoking, hypertension, microalbuminuria, BMI, and dyslipidemia, as defined in the 2021 European Society of Cardiology (ESC) guidelines on cardiovascular disease prevention in clinical practice [1]. However, we considered including biomarkers that evaluated a novel variation of an established CVD risk factor, such as HbA1c variability compared to a single time-point measurement.

The strength and quality of evidence in observational prognostic studies depend on how well any known or suspected confounding variables between the prognostic factor and the outcome are accounted for. To be considered a novel risk marker, a biomarker must improve risk prediction beyond traditional markers [1]. Thus, we began by excluding all studies that did not adjust for any CVD risk factors.

[1] Frank L J Visseren and others, 2021 ESC Guidelines on cardiovascular disease prevention in clinical practice: Developed by the Task Force for cardiovascular disease prevention in clinical practice with representatives of the European Society of Cardiology and 12 medical societies With the special contribution of the European Association of Preventive Cardiology (EAPC), *European Heart Journal*, Volume 42, Issue 34, 7 September 2021, Pages 3227–3337, https://doi.org/10.1093/eurheartj/ehab484

#### Supplemental text 3: Membership of the EASD/ADA PMID

Deirdre K. Tobias[1,2]*, Jordi Merino[3-5]*, Abrar Ahmad[6]*, Catherine Aiken[7,8]*, Jamie L. Benham[9]*, Dhanasekaran Bodhini[10]*, Amy L. Clark[11], Kevin Colclough[12]*, Rosa Corcoy[13-15], Sara J. Cromer[4,16,17]*, Daisy Duan[18]*, Jamie L. Felton[19]*, Ellen C. Francis[20]*, Pieter Gillard[21]*, Véronique Gingras[22,23]*, Romy Gaillard[24]*, Eram Haider[25]*, Alice Hughes[12]*, Jennifer M. Ikle[26,27]*, Laura M. Jacobsen[28]*, Anna R. Kahkoska[29]*, Jarno L.T. Kettunen[30-32]*, Raymond J. Kreienkamp[4,5,16,33]*, Lee-Ling Lim[34-36]*, Jonna M.E. Männistö[37,38]*, Robert Massey[25]*, Niamh-Maire Mclennan[39]*, Rachel G. Miller[40], Mario Luca Morieri[41,42]*, Jasper Most[43]*, Rochelle N. Naylor[44]*, Bige Ozkan[45,46]*, Kashyap Amratlal Patel[12]*, Scott J. Pilla[47,48]*, Katsiaryna Prystupa[49,50]*, Sridaran Raghaven[51,52]*, Mary R. Rooney[45,53]*, Martin Schön[49,50,54]*, Zhila Semnani-Azad[2], Magdalena Sevilla-Gonzalez[16,17,55]*, Pernille Svalastoga[56,57]*, Wubet Worku Takele[58]*, Claudia Ha-ting Tam[36,59,60]*, Anne C. Baun Thuesen[3]*, Mustafa Tosur[61-63]*, Amelia S. Wallace[45,53]*, Caroline C. Wang[53]*, Jessie J. Wong[64]*, Jennifer M. Yamamoto[65]*, Katherine Young[12]*, Chloé Amouyal[66,67], Mette K. Andersen[3], Maxine P. Bonham[68], Mingling Chen[69], Feifei Cheng[70], Tinashe Chikowore[17,71-73], Sian C Chivers[74], Christoffer Clemmensen[3], Dana Dabelea[75], Adem Y. Dawed[25], Aaron J. Deutsch[5,16,17], Laura T. Dickens[76], Linda A. DiMeglio[77,78], Monika Dudenhöffer-Pfeifer[6], Carmella Evans-Molina[78-80], María Mercè Fernández-Balsells[81,82], Hugo Fitipaldi[6], Stephanie L. Fitzpatrick[83], Stephen E. Gitelman[84], Mark O. Goodarzi[85,86], Jessica A. Grieger[87,88], Marta Guasch-Ferré[2,89], Nahal Habibi[87,88], Torben Hansen[3], Chuiguo Huang[36,59] , Arianna Harris-Kawano[19], Heba M. Ismail[19], Benjamin Hoag[90,91], Randi K. Johnson[92,93], Angus G. Jones[12,94], Robert W. Koivula[95], Aaron Leong[4,17,96], Gloria K.W. Leung[68], Ingrid M. Libman[97], Kai Liu[87], S. Alice Long[98], William L. Lowe, Jr.[99], Robert W. Morton[100-102], Ayesha A. Motala[103], Suna Onengut-Gumuscu[104], James S. Pankow[105], Maleesa Pathirana[87,88], Sofia Pazmino[106], Dianna Perez[19], John R. Petrie[107], Camille E. Powe[4,16,17,108], Alejandra Quinteros[87], Rashmi Jain[109,110], Debashree Ray[53,111], Mathias Ried-Larsen[112,113], Zeb Saeed[114], Vanessa Santhakumar[1], Sarah Kanbour[47,115], Sudipa Sarkar[47], Gabriela S.F. Monaco[78,79], Denise M. Scholtens[116], Elizabeth Selvin[45,53], Wayne Huey-Herng Sheu[117-119], Cate Speake[120], Maggie A. Stanislawski[92], Nele Steenackers[106], Andrea K. Steck[121], Norbert Stefan[50,54,122], Julie Støy[123], Rachael Taylor[124], Sok Cin Tye[125,126], Gebresilasea Gendisha Ukke[58], Marzhan Urazbayeva[62,127], Bart Van der Schueren[106,128], Camille Vatier[129,130], John M. Wentworth[131-133], Wesley Hannah[134,135], Sara L. White[74,136], Gechang Yu[36,59], Yingchai Zhang[36,59], Shao J. Zhou[88,137], Jacques Beltrand[138,139], Michel Polak[138,139], Ingvild Aukrust[56,140], Elisa de Franco[12], Sarah E. Flanagan[12], Kristin A. Maloney[141], Andrew McGovern[12], Janne Molnes[56,140], Mariam Nakabuye[3], Pål Rasmus Njølstad[56,57], Hugo Pomares-Millan[6,142], Michele Provenzano[143], Cécile Saint-Martin[144], Cuilin Zhang[145,146], Yeyi Zhu[147,148], Sungyoung Auh[149], Russell J. de Souza[101,150], Andrea J Fawcett[151,152], Chandra Gruber[153], Eskedar Getie Mekonnen[154,155], Emily Mixter[156], Diana Sherifali[101,157], Robert H. Eckel[158], John J. Nolan[159,160], Louis H. Philipson[156], Rebecca J. Brown[149]†, Liana K. Billings[161,162]†, Kristen Boyle[75]†, Tina Costacou[40], John M. Dennis[12]†, Jose C. Florez[4,5,16,17]†, Anna L. Gloyn[26,27,163]†, Maria F. Gomez[6,164]†, Peter A. Gottlieb[121]†, Siri Atma W. Greeley[165]†, Kurt Griffin[110,166]†, Andrew T. Hattersley[12,94]†, Irl B. Hirsch[167]†, Marie-France Hivert[4,168,169]†, Korey K. Hood[64]†, Jami L. Josefson[151]†, Soo Heon Kwak[170]†, Lori M. Laffel[171]†, Siew S. Lim[58]†, Ruth J.F. Loos[3,172]†, Ronald C.W. Ma[36,59,60]†, Chantal Mathieu[21]†, Nestoras Mathioudakis[47]†, James B. Meigs[17,96,173]†, Shivani Misra[174,175]†, Viswanathan Mohan[176]†, Rinki Murphy[177-179]†, Richard Oram[12,94]†, Katharine R. Owen[95,180]†, Susan E. Ozanne[181]†, Ewan R. Pearson[25]†, Wei Perng[75]†, Toni I. Pollin[141,182]†, Rodica Pop-Busui[183], Richard E. Pratley[184]†, Leanne M. Redman[185]†, Maria J. Redondo[61,62]†, Rebecca M. Reynolds[39]†, Robert K. Semple†[39,186], Jennifer L. Sherr[187]†, Emily K. Sims[188]†, Arianne Sweeting[189,190]†, Tiinamaija Tuomi[30-32]†, Miriam S. Udler[4,5,16,17]†, Kimberly K. Vesco[191]†, Tina Vilsbøll[192,193]†, Robert Wagner[49,50,194]†, Stephen S. Rich[104]†, Paul W. Franks[2,6,95,102]†

**AFFILIATIONS**

[1]Division of Preventative Medicine, Department of Medicine, Brigham and Women's Hospital and Harvard Medical School, Boston, MA, USA.

[2]Department of Nutrition, Harvard T.H. Chan School of Public Health, Boston, MA, USA.

[3]Novo Nordisk Foundation Center for Basic Metabolic Research, Faculty of Health and Medical Sciences, University of Copenhagen, Copenhagen, Denmark.

[4]Diabetes Unit, Endocrine Division, Massachusetts General Hospital, Boston, MA, USA.

[5]Center for Genomic Medicine, Massachusetts General Hospital, Boston, MA, USA.

[6]Department of Clinical Sciences, Lund University Diabetes Centre, Lund University, Malmö, Sweden.

[7]Department of Obstetrics and Gynaecology, the Rosie Hospital, Cambridge, UK.

[8]NIHR Cambridge Biomedical Research Centre, University of Cambrdige, Cambridge, UK.

[9]Departments of Medicine and Community Health Sciences, Cumming School of Medicine, University of Calgary, Calgary, AB, Canada.

[10]Department of Molecular Genetics, Madras Diabetes Research Foundation, Chennai, India.

[11]Division of Pediatric Endocrinology, Department of Pediatrics, Saint Louis University School of Medicine, SSM Health Cardinal Glennon Children's Hospital, St. Louis, MO, USA.

[12]Department of Clinical and Biomedical Sciences, University of Exeter Medical School, Exeter, Devon, UK.

[13]CIBER-BBN, ISCIII, Madrid, Spain.

[14]Institut d'Investigació Biomèdica Sant Pau (IIB SANT PAU), Barcelona, Spain.

[15]Departament de Medicina, Universitat Autònoma de Barcelona, Bellaterra, Spain.

[16]Programs in Metabolism and Medical & Population Genetics, Broad Institute, Cambridge, MA, USA.

[17]Department of Medicine, Harvard Medical School, Boston, MA, USA.

[18]Division of Endocrinology, Diabetes and Metabolism, Johns Hopkins University School of Medicine, Baltimore, MD, USA.

[19]Department of Pediatrics, Indiana University School of Medicine, Indianapolis, IN, USA.

[20]Department of Biostatistics and Epidemiology, Rutgers School of Public Health, Piscataway, NJ, USA.

[21]University Hospital Leuven, Leuven, Belgium.

[22]Department of Nutrition, Université de Montréal, Montreal, Quebec, Canada.

[23]Research Center, Sainte-Justine University Hospital Center, Montreal, Quebec, Canada.

[24]Department of Pediatrics, Erasmus Medical Center, Rotterdam, The Netherlands.

[25]Division of Population Health & Genomics, School of Medicine, University of Dundee, Dundee, UK.

[26]Department of Pediatrics, Stanford School of Medicine, Stanford University, CA, USA.

[27]Stanford Diabetes Research Center, Stanford School of Medicine, Stanford University, CA, USA.

[28]University of Florida, Gainesville, FL, USA.
[29]Department of Nutrition, University of North Carolina at Chapel Hill, Chapel Hill, NC, USA.
[30]Helsinki University Hospital, Abdominal Centre/Endocrinology, Helsinki, Finland.
[31]Folkhalsan Research Center, Helsinki, Finland.
[32]Institute for Molecular Medicine Finland FIMM, University of Helsinki, Helsinki, Finland.
[33]Department of Pediatrics, Division of Endocrinology, Boston Children's Hospital, Boston, MA, USA.
[34]Department of Medicine, Faculty of Medicine, University of Malaya, Kuala Lumpur, Malaysia.
[35]Asia Diabetes Foundation, Hong Kong SAR, China.
[36]Department of Medicine & Therapeutics, Chinese University of Hong Kong, Hong Kong SAR, China.
[37]Departments of Pediatrics and Clinical Genetics, Kuopio University Hospital, Kuopio, Finland.
[38]Department of Medicine, University of Eastern Finland, Kuopio, Finland.
[39]Centre for Cardiovascular Science, Queen's Medical Research Institute, University of Edinburgh, Edinburgh, UK.
[40]Department of Epidemiology, University of Pittsburgh, Pittsburgh, PA, USA.
[41]Metabolic Disease Unit, University Hospital of Padova, Padova, Italy.
[42]Department of Medicine, University of Padova, Padova, Italy.
[43]Department of Orthopedics, Zuyderland Medical Center, Sittard-Geleen, The Netherlands.
[44]Departments of Pediatrics and Medicine, University of Chicago, Chicago, Illinois, USA.
[45]Welch Center for Prevention, Epidemiology, and Clinical Research, Johns Hopkins Bloomberg School of Public Health, Baltimore, Maryland, USA.
[46]Ciccarone Center for the Prevention of Cardiovascular Disease, Johns Hopkins School of Medicine, Baltimore, MD, USA.
[47]Department of Medicine, Johns Hopkins University, Baltimore, MD, USA.
[48]Department of Health Policy and Management, Johns Hopkins University Bloomberg School of Public Health, Baltimore, Maryland, USA.
[49]Institute for Clinical Diabetology, German Diabetes Center, Leibniz Center for Diabetes Research at Heinrich Heine University Düsseldorf, Auf'm Hennekamp 65, 40225 Düsseldorf, Germany.
[50]German Center for Diabetes Research (DZD), Ingolstädter Landstraße 1, 85764, Neuherberg, Germany.
[51]Section of Academic Primary Care, US Department of Veterans Affairs Eastern Colorado Health Care System, Aurora, CO, USA.
[52]Department of Medicine, University of Colorado School of Medicine, Aurora, CO, USA.
[53]Department of Epidemiology, Johns Hopkins Bloomberg School of Public Health, Baltimore, Maryland, USA.
[54]Institute of Diabetes Research and Metabolic Diseases (IDM), Helmholtz Center Munich, Neuherberg, Germany.
[55]Clinical and Translational Epidemiology Unit, Massachusetts General Hospital, Boston MA, USA.
[56]Mohn Center for Diabetes Precision Medicine, Department of Clinical Science, University of Bergen, Bergen, Norway.
[57]Children and Youth Clinic, Haukeland University Hospital, Bergen, Norway.
[58]Eastern Health Clinical School, Monash University, Melbourne, Victoria, Australia.
[59]Laboratory for Molecular Epidemiology in Diabetes, Li Ka Shing Institute of Health Sciences, The Chinese University of Hong Kong, Hong Kong, China.

[60]Hong Kong Institute of Diabetes and Obesity, The Chinese University of Hong Kong, Hong Kong, China.
[61]Department of Pediatrics, Baylor College of Medicine, Houston, TX, USA.
[62]Division of Pediatric Diabetes and Endocrinology, Texas Children's Hospital, Houston, TX, USA.
[63]Children's Nutrition Research Center, USDA/ARS, Houston, TX, USA.
[64]Stanford University School of Medicine, Stanford, CA, USA.
[65]Internal Medicine, University of Manitoba, Winnipeg, MB, Canada.
[66]Department of Diabetology, APHP, Paris, France.
[67]Sorbonne Université, INSERM, NutriOmic team, Paris, France.
[68]Department of Nutrition, Dietetics and Food, Monash University, Melbourne, Victoria, Australia.
[69]Monash Centre for Health Research and Implementation, Monash University, Clayton, VIC, Australia.
[70]Health Management Center, The Second Affiliated Hospital of Chongqing Medical University, Chongqing Medical University, Chongqing, China.
[71]MRC/Wits Developmental Pathways for Health Research Unit, Department of Paediatrics, Faculty of Health Sciences, University of the Witwatersrand, Johannesburg, South Africa.
[72]Channing Division of Network Medicine, Brigham and Women's Hospital, Boston, MA, USA.
[73]Sydney Brenner Institute for Molecular Bioscience, Faculty of Health Sciences, University of the Witwatersrand, Johannesburg, South Africa.
[74]Department of Women and Children's health, King's College London, London, UK.
[75]Lifecourse Epidemiology of Adiposity and Diabetes (LEAD) Center, University of Colorado Anschutz Medical Campus, CO, USA.
[76]Section of Adult and Pediatric Endocrinology, Diabetes and Metabolism, Kovler Diabetes Center, University of Chicago, Chicago, USA.
[77]Department of Pediatrics, Riley Hospital for Children, Indiana University School of Medicine, Indianapolis, IN, USA.
[78]Center for Diabetes and Metabolic Diseases, Indiana University School of Medicine, IN, USA.
[79]Herman B Wells Center for Pediatric Research, Indiana University School of Medicine, IN, USA.
[80]Richard L. Roudebush VAMC, Indianapolis, IN, USA.
[81]Biomedical Research Institute Girona, IdIBGi, Girona, Spain.
[82]Diabetes, Endocrinology and Nutrition Unit Girona, University Hospital Dr Josep Trueta, Girona, Spain.
[83]Institute of Health System Science, Feinstein Institutes for Medical Research, Northwell Health, Manhasset, NY, USA.
[84]University of California at San Francisco, Department of Pediatrics, Diabetes Center; San Francisco, CA, USA.
[85]Division of Endocrinology, Diabetes and Metabolism, Cedars-Sinai Medical Center, Los Angeles, CA, USA.
[86]Department of Medicine, Cedars-Sinai Medical Center, Los Angeles, CA, USA.
[87]Adelaide Medical School, Faculty of Health and Medical Sciences, The University of Adelaide, Adelaide, Australia.
[88]Robinson Research Institute, The University of Adelaide, Adelaide, Australia.
[89]Department of Public Health and Novo Nordisk Foundation Center for Basic Metabolic Research, Faculty of Health and Medical Sciences, University of Copenhagen, 1014 Copenhagen, Denmark.

[90]Division of Endocrinology and Diabetes, Department of Pediatrics, Sanford Children's Hospital, Sioux Falls, SD, USA.
[91]University of South Dakota School of Medicine, E Clark St, Vermillion, SD, USA.
[92]Department of Biomedical Informatics, University of Colorado Anschutz Medical Campus, Aurora, CO, USA.
[93]Department of Epidemiology, Colorado School of Public Health, Aurora, CO, USA.
[94]Royal Devon University Healthcare NHS Foundation Trust, Exeter, UK.
[95]Oxford Centre for Diabetes, Endocrinology and Metabolism, University of Oxford, Oxford, UK.
[96]Division of General Internal Medicine, Massachusetts General Hospital, Boston, MA, USA.
[97]UPMC Children's Hospital of Pittsburgh, Pittsburgh, PA, USA.
[98]Center for Translational Immunology, Benaroya Research Institute, Seattle, WA, USA.
[99]Department of Medicine, Northwestern University Feinberg School of Medicine, Chicago, IL USA.
[100]Department of Pathology & Molecular Medicine, McMaster University, Hamilton, Canada
[101]Population Health Research Institute, Hamilton, Canada.
[102]Department of Translational Medicine, Medical Science, Novo Nordisk Foundation, Tuborg Havnevej 19, 2900 Hellerup, Denmark.
[103]Department of Diabetes and Endocrinology, Nelson R Mandela School of Medicine, University of KwaZulu-Natal, Durban, South Africa.
[104]Center for Public Health Genomics, Department of Public Health Sciences, University of Virginia, Charlottesville, VA, USA.
[105]Division of Epidemiology and Community Health, School of Public Health, University of Minnesota, MN, USA.
[106]Department of Chronic Diseases and Metabolism, Clinical and Experimental Endocrinology, KU Leuven, Leuven, Belgium.
[107]School of Health and Wellbeing, College of Medical, Veterinary and Life Sciences, University of Glasgow, UK.
[108]Department of Obstetrics, Gynecology, and Reproductive Biology, Massachusetts General Hospital and Harvard Medical School, Boston, MA, USA.
[109]Sanford Children's Specialty Clinic, Sioux Falls, SD, USA.
[110]Department of Pediatrics, Sanford School of Medicine, University of South Dakota, Sioux Falls, SD, USA.
[111]Department of Biostatistics, Johns Hopkins Bloomberg School of Public Health, Baltimore, Maryland, USA.
[112]Centre for Physical Activity Research, Rigshospitalet, Copenhagen, Denmark.
[113]Institute for Sports and Clinical Biomechanics, University of Southern Denmark, Denmark.
[114]Indiana University, Department of Endocrinology, Diabetes and Metabolism, Indianapolis, IN, USA.
[115]AMAN Hospital, Doha, Qatar.
[116]Department of Preventive Medicine, Division of Biostatistics, Northwestern University Feinberg School of Medicine, Chicago, IL, USA.
[117]Institute of Molecular and Genomic Medicine, National Health Research Institutes, Taiwan.
[118]Divsion of Endocrinology and Metabolism, Taichung Veterans General Hospital, Taichung, Taiwan.
[119]Division of Endocrinology and Metabolism, Taipei Veterans General Hospital, Taipei, Taiwan.
[120]Center for Interventional Immunology, Benaroya Research Institute, Seattle, WA, USA.

[121]Barbara Davis Center for Diabetes, University of Colorado Anschutz Medical Campus, CO, USA.
[122]University Hospital of Tübingen, Tübingen, Germany.
[123]Steno Diabetes Center Aarhus, Aarhus University Hospital, Aarhus, Denmark.
[124]University of Newcastle, Newcastle upon Tyne, UK.
[125]Sections on Genetics and Epidemiology, Joslin Diabetes Center, Harvard Medical School, Boston, MA, USA.
[126]Department of Clinical Pharmacy and Pharmacology, University Medical Center Groningen, Groningen, The Netherlands.
[127]Gastroenterology, Baylor College of Medicine, Houston, TX, USA.
[128]Department of Endocrinology, University Hospitals Leuven, Belgium.
[129]Sorbonne University, Inserm U938, Saint-Antoine Research Centre, Institute of Cardiometabolism and Nutrition, Paris 75012, France.
[130]Department of Endocrinology, Diabetology and Reproductive Endocrinology, Assistance Publique-Hôpitaux de Paris, Saint-Antoine University Hospital, National Reference Center for Rare Diseases of Insulin Secretion and Insulin Sensitivity (PRISIS), Paris, France.
[131]Royal Melbourne Hospital Department of Diabetes and Endocrinology, Parkville, Vic, Australia.
[132]Walter and Eliza Hall Institute, Parkville, Vic, Australia.
[133]University of Melbourne Department of Medicine, Parkville, Vic, Australia.
[134]Deakin University, Melbourne, Australia.
[135]Department of Epidemiology, Madras Diabetes Research Foundation, Chennai, India.
[136]Department of Diabetes and Endocrinology, Guy's and St Thomas' Hospitals NHS Foundation Trust, London, UK.
[137]School of Agriculture, Food and Wine, University of Adelaide, Adelaide, Australia.
[138]Institut Cochin, Inserm U 10116, Paris, France.
[139]Pediatric endocrinology and diabetes, Hopital Necker Enfants Malades, APHP Centre, université de Paris, Paris, France.
[140]Department of Medical Genetics, Haukeland University Hospital, Bergen, Norway.
[141]Department of Medicine, University of Maryland School of Medicine, Baltimore, MD, USA.
[142]Department of Epidemiology, Geisel School of Medicine at Dartmouth, Hanover, NH, USA.
[143]Nephrology, Dialysis and Renal Transplant Unit, IRCCS—Azienda Ospedaliero-Universitaria di Bologna, Alma Mater Studiorum University of Bologna, Bologna, Italy.
[144]Department of Medical Genetics, AP-HP Pitié-Salpêtrière Hospital, Sorbonne University, Paris, France.
[145]Global Center for Asian Women's Health, Yong Loo Lin School of Medicine, National University of Singapore, Singapore.
[146]Department of Obstetrics and Gynecology, Yong Loo Lin School of Medicine, National University of Singapore, Singapore.
[147]Kaiser Permanente Northern California Division of Research, Oakland, California, USA.
[148]Department of Epidemiology and Biostatistics, University of California San Francisco, California, USA.
[149]National Institute of Diabetes and Digestive and Kidney Diseases, National Institutes of Health, Bethesda, MD, USA.
[150]Department of Health Research Methods, Evidence, and Impact, Faculty of Health Sciences, McMaster University, Hamilton, ON, Canada.
[151]Ann & Robert H. Lurie Children's Hospital of Chicago, Department of Pediatrics, Northwestern University Feinberg School of Medicine, Chicago, IL, USA.

[152]Department of Clinical and Organizational Development, Chicago, IL, USA.
[153]American Diabetes Association, Arlington, Virginia, USA.
[154]College of Medicine and Health Sciences, University of Gondar, Gondar, Ethiopia.
[155]Global Health Institute, Faculty of Medicine and Health Sciences, University of Antwerp, 2160 Antwerp, Belgium.
[156]Department of Medicine and Kovler Diabetes Center, University of Chicago, Chicago, IL, USA.
[157]School of Nursing, Faculty of Health Sciences, McMaster University, Hamilton, Canada.
[158]Division of Endocrinology, Metabolism, Diabetes, University of Colorado, CO, USA.
[159]Department of Clinical Medicine, School of Medicine, Trinity College Dublin, Dublin, Ireland.
[160]Department of Endocrinology, Wexford General Hospital, Wexford, Ireland.
[161]Division of Endocrinology, NorthShore University HealthSystem, Skokie, IL, USA.
[162]Department of Medicine, Prtizker School of Medicine, University of Chicago, Chicago, IL, USA.
[163]Department of Genetics, Stanford School of Medicine, Stanford University, CA, USA.
[164]Faculty of Health, Aarhus University, Denmark.
[165]Departments of Pediatrics and Medicine and Kovler Diabetes Center, University of Chicago, Chicago, USA.
[166]Sanford Research, Sioux Falls, SD, USA.
[167]University of Washington School of Medicine, Seattle, WA, USA.
[168]Department of Population Medicine, Harvard Medical School, Harvard Pilgrim Health Care Institute, Boston, MA, USA.
[169]Department of Medicine, Universite de Sherbrooke, Sherbrooke, QC, Canada.
[170]Department of Internal Medicine, Seoul National University College of Medicine, Seoul National University Hospital, Seoul, Republic of Korea.
[171]Joslin Diabetes Center, Harvard Medical School, Boston, MA, USA.
[172]Charles Bronfman Institute for Personalized Medicine, Icahn School of Medicine at Mount Sinai, New York, NY, USA.
[173]Broad Institute, Cambridge, MA, USA.
[174]Division of Metabolism, Digestion and Reproduction, Imperial College London, London, UK.
[175]Department of Diabetes & Endocrinology, Imperial College Healthcare NHS Trust, London, UK.
[176]Department of Diabetology, Madras Diabetes Research Foundation & Dr. Mohan's Diabetes Specialities Centre, Chennai, India.
[177]Department of Medicine, Faculty of Medicine and Health Sciences, University of Auckland, Auckland, New Zealand.
[178]Auckland Diabetes Centre, Te Whatu Ora Health New Zealand, Auckland, New Zealand.
[179]Medical Bariatric Service, Te Whatu Ora Counties, Health New Zealand, Auckland, New Zealand.
[180]Oxford NIHR Biomedical Research Centre, University of Oxford, Oxford, UK.
[181]University of Cambridge, Metabolic Research Laboratories and MRC Metabolic Diseases Unit, Wellcome-MRC Institute of Metabolic Science, Cambridge, UK.
[182]Department of Epidemiology & Public Health, University of Maryland School of Medicine, Baltimore, MD.
[183]Department of Internal Medicine, Division of Metabolism, Endocrinology and Diabetes, University of Michigan, MI, USA.
[184]AdventHealth Translational Research Institute, Orlando, FL, USA.
[185]Pennington Biomedical Research Center, Baton Rouge, LA, USA.

[186]MRC Human Genetics Unit, Institute of Genetics and Cancer, University of Edinburgh, Edinburgh, UK.
[187]Yale School of Medicine, New Haven, CT, USA.
[188]Pediatric Endocrinology and Diabetology, Center for Diabetes and Metabolic Diseases, Herman B Wells Center for Pediatric Research, Indiana University School of Medicine, IN, USA.
[189]Faculty of Medicine and Health, University of Sydney, Sydney, NSW, Australia.
[190]Department of Endocrinology, Royal Prince Alfred Hospital, Sydney, NSW, Australia
[191]Kaiser Permanente Northwest, Kaiser Permanente Center for Health Research, Portland, OR, USA.
[192]Clinial Research, Steno Diabetes Center Copenhagen, Herlev, Denmark.
[193]Department of Clinical Medicine, Faculty of Health and Medical Sciences, University of Copenhagen, Copenhagen, Denmark.
[194]Department of Endocrinology and Diabetology, University Hospital Düsseldorf, Heinrich Heine University Düsseldorf, Moorenstr. 5, 40225 Düsseldorf, Germany.

#### Supplemental Table 1. Participant Intervention Comparison Outcomes and Setting (PICOS) framework.

| Items | Details |
|---|---|
| ***Participants*** | Patients with Type 2 Diabetes |
| ***Intervention/Exposure*** | Non-Genetic Biomarkers, or Genetic-Biomarkers, or Risk Scores/Engine |
| ***Comparison*** | Not applicable |
| ***Outcomes*** | Coronary heart disease (CHD) and/or cardiovascular (CV) mortality |
| ***Study Design*** | Longitudinal studies |

#### Supplemental Table 2. Risk of Bias Assessment using Modified Newcastle-Ottawa Scale.

| Items | Quality Points | Risk of bias |
|---|---|---|
| ***Representativeness of the exposed cohort*** | | |
| 1. Truly representative | 3 | Low |
| 2. Somewhat representative | 2 | Medium |
| 3. Selected group | 1 | High |
| 4. No description of the derivation of the cohort | 0 | High |
| ***Selection of the non-exposed cohort*** | | |
| 1. Drawn from the same community as the exposed cohort | 2 | Low |
| 2. Drawn from a different source | 1 | Medium |
| 3. No description of the derivation of the non-exposed cohort | 0 | High |
| ***Ascertainment of exposure*** | | |
| 1. Measured by investigators or from electronic medical record review | 2 | Low |
| 2. Patient self-reported | 1 | Medium |
| 3. No description | 0 | High |
| ***Assessment of outcome*** | | |
| 1. Clearly defined outcome (e.g., ICD-10 codes, clinical documentation) or adjudication | 3 | Low |
| 2. Record linkage (registry) | 2 | Medium |
| 3. Other (e.g., reported by patients) | 1 | High |
| 4. No description | 0 | High |
| ***Was follow-up long enough for outcomes to occur?*** | | |
| 1. Yes (3 years for naive CVD event; 1 year for recurrent CVD event after intervention) | 3 | Low |
| 2. No | 1 | Medium |
| ***Adequacy of follow-up of cohorts*** | | |
| 1. Complete follow-up - all subjects accounted for | 3 | Low |
| 2. Subjects' loss to follow up unlikely to introduce bias - number lost less than or equal to 20% or description of those lost suggested no different from those followed. | 2 | Medium |
| 3. Follow up rate less than 80% and no description of those lost | 1 | Medium |
| 4. No statement | 0 | High |
| ***Number of covariates being included in the models*** | | |
| 1. Highest/fourth quartile of distribution | 4 | Stratified by tertiles of distribution |
| 2. Third quartile of distribution | 3 | |
| 3. Second quartile of distribution | 2 | |
| 4. Lowest/first quartile of distribution | 1 | |
| ***Number of established CVD risk factors being accounted for*** | | |
| 1. N > 75 percentile | 6 | Stratified by tertiles of distribution |
| 2. 50 < N ≤ 75 percentile | 4 | |
| 3. 25 < N ≤ 50 percentile | 2 | |
| 4. N ≤ 25 percentile | 0 | |

#### Supplemental Table 3. Evaluation of the clinical utility of new biomarkers included in the present meta-analysis.

| Step | Evaluation | Description |
|---|---|---|
| 1 | Association | Has a statistically significant association been reported with cardiovascular disease for the new biomarker with and without adjustment for traditional risk factors? |
| 2 | Discrimination | Does addition of the new biomarker to a model with traditional risk factors lead to significant improvement in discrimination (assessed by the C-statistic)? |
| | Net reclassification improvement | Does addition of the new biomarker to a model with traditional risk factors lead to appropriate reclassification of people to either high- or low-risk status? |
| | Integrated discrimination index | Quantification of predicted probabilities of cardiovascular events and non-cardiovascular events based on inclusion of the biomarker in a model with traditional risk factors |

Modified from (1, 2). Calibration, which is an additional parameter used for evaluation of clinical utility by Hlatky et al., was not used in the present meta-analysis. Calibration is defined as *"Does addition of the new factor to a traditional risk factor model result in improved calibration, defined as agreement between the predicted and observed rates of end points?"*; very few articles provided sufficient information to allow evaluation of this parameter.

**References:**

1. Hlatky MA, Greenland P, Arnett DK, Ballantyne CM, Criqui MH, Elkind MSV, et al. Criteria for evaluation of novel markers of cardiovascular risk: a scientific statement from the American Heart Association. Circulation. 2009;119(17):2408-16.
2. US National Heart L, and Blood Institute. US National Heart, Lung, and Blood Institute. 2013 Report on the Assessment of Cardiovascular Risk: Full Work Group Report Supplement.

#### Supplemental Table 4. Established CVD risk factors according to 2021 ESC Guidelines[1].

| Established CVD Risk Factors |
|---|
| • Age |
| • Sex/gender |
| • Hypertension (systolic/diastolic blood pressure, treated hypertension) |
| • Established CVD (cardiovascular disease, cerebrovascular, PVD, heart failure) |
| • Antithrombotic treatment (aspirin) |
| • Dyslipidemia or hyperlipidemia (LDL, HDL, total cholesterol) |
| • Lipid-lowering treatment (statin, PCSK-9 inhibitor, ezetimibe) |
| • A1C |
| • Insulin therapy |
| • Duration of diabetes |
| • Atrial fibrillation |
| • Overweight/obesity/adiposity (body weight, BMI) |
| • Cigarette smoking |
| • Nephropathy or chronic kidney disease |
| • eGFR |
| • Albumin/creatinine ratio (albuminuria, both micro + macro) or proteinuria |
| • Retinopathy |
| • Neuropathy |
| • Any Microvascular disease |
| • Geographic region |
| • Race/ethnicity |

CVD, cardiovascular disease; PVD, peripheral vascular disease; LDL, low-density lipoprotein; HDL, high-density lipoprotein; PCSK-9, proprotein convertase subtilisin/kexin-type 9; A1C, hemoglobin A1C; BMI, body mass index; eGFR, estimated glomerular filtration rate.

**Reference:**

1. Visseren, F. L. J. *et al.* 2021 ESC Guidelines on cardiovascular disease prevention in clinical practice. *Eur Heart J* **42**, 3227-3337, doi:10.1093/eurheartj/ehab484 (2021).

#### Supplementary Table 5. Included 321 biomarker studies.

| Study;<br>Design;<br>Country | No. of participant;<br>Enrollment year;<br>Follow-up (years) | Study Population | Exposure(s) | Outcome | Only established risk factors |
|---|---|---|---|---|---|
| Nelson 1990;<br>Prosp. Cohort;<br>United States | 1093;<br>1975 to 1984;<br>FUP mean 5 | No ASCVD | proteinuria; renal insufficiency; an abnormal electrocardiogram; diabetic retinopathy; insulin therapy; medial arterial calcification | CHD | 0 |
| Morrish 1991;<br>Prosp. Cohort;<br>United Kingdom | 497;<br>1985 to 1987;<br>FUP mean 8.33 | No ASCVD | proteinuria; Systolic blood pressure; serum cholesterol; smoking | CHD | 1 |
| Standl 1996;<br>Prosp. Cohort;<br>Germany | 290;<br>1984 to 1985;<br>FUP mean 10 | No ASCVD | HbA1c; age; von Willebrand-factor protein | CVM, Stroke | 0 |
| Niskanen 1996;<br>Prosp. Cohort;<br>Finland | 133;<br>1979 to 1981;<br>FUP mean 10 | Any CVD risk | Albuminuria/Hyperinsulinemia | CVM | 0 |
| Lehto 1996;<br>Prosp. Cohort;<br>Finland | 1059;<br>1982 to 1984;<br>FUP mean 7 | Any CVD risk | Medial artery calcification (MAC) | 3p MACE, PAD | 0 |
| Hanefeld 1996;<br>Prosp. Cohort;<br>Germany | 1139;<br>1986 to 1988;<br>FUP mean 12 | Any CVD risk | blood pressure; smoking | CHD | 1 |
| Beilin 1996;<br>Prosp. Cohort;<br>Australia | 666;<br>1986 to 1993;<br>FUP mean 4.7 | No ASCVD | urine albumin excretion | CHD, CVM | 1 |
| Koch 1997;<br>Prosp. Cohort;<br>Germany | 412;<br>1985 to 1994;<br>FUP mean 3 | Any CVD risk | Age; Stroke; Apo-A-I; Fibrinogen | CHD, CVM | 0 |
| Wei 1998;<br>Prosp. Cohort;<br>United States | 471;<br>1979 to 1988;<br>FUP mean 7.5 | Any CVD risk | Fasting plasma glucose | CVM | 0 |
| Turner 1998;<br>Prosp. Cohort;<br>United Kingdom | 2693;<br>1977 to 1991;<br>FUP median 7.9 | No ASCVD | BP; HDL; LDL; smoking; hyperglycaemia | CHD, Angina | 0 |

| | | | | | |
|---|---|---|---|---|---|
| Niskanen 1998; Prosp. Cohort; Finland | 133; 1979 to 1981; FUP mean 15 | No ASCVD | hyperglycemia | CVM | 0 |
| Vanzetto 1999; Prosp. Cohort; France | 158; 1989 to 1994; FUP mean 1.9 | No ASCVD with High CVD risk | Exercise stress test; Thallium-201 scintigraphy | CHD, CVM | 0 |
| Stehouwer 1999; Prosp. Cohort; Netherlands | 211; 1989 to 1990; FUP median 6.4 | Any CVD risk | Plasma homocysteine concentration (tHcy) | CVM | 0 |
| Yamasaki 2000; Prosp. Cohort; Japan | 287; NR; FUP mean 3.1 | No ASCVD | Carotid intima-media thickness (IMT) | CHD, Angina | 0 |
| Valmadrid 2000; Prosp. Cohort; United States | 840; 1980 to 1982; FUP mean 12 | Any CVD risk | Gross proteinuria | CVM | 1 |
| Saito 2000; Prosp. Cohort; United States | 1676; 1987 to 1989; FUP mean 6.6 | No ASCVD | albumin; von Willebrand factor; leukocyte count; factor VIII; fibrinogen | 3p MACE, Revasc, HF | 0 |
| Howard 2000; Retrosp. Cohort; Multinational | 4378; 1989 to 1992; FUP mean 4.8 | Est. ASCVD | LDL cholesterol | CVM | 1 |
| Casiglia 2000; Retrosp. Case-control; Brazil, Italy | 683; NR; FUP mean 6 | Any CVD risk | macroalbuminuria; age; hypertension; coronary heart disease (CHD) | CVM | 1 |
| Christensen 2000; Prosp. Cohort; Denmark | 324; 1987 to 1987; FUP median 9.4 | Any CVD risk | QT dispersion | CVM | 0 |
| Spoelstra-deMan 2001; Prosp. Cohort; Netherlands | 58; 1989 to 1996; FUP median 7 | Any CVD risk | progression of microalbuminuria | CVM | 1 |
| Fuller 2001; Prosp. Cohort; Multinational | 3483; up to 1978; FUP mean 12 | Any CVD risk | Proteinuria; Systolic blood pressure (SBP); Serum cholesterol; Retinopathy; Smoking; ECG abnormalities; Diabetes duration; Serum triglyceride; Plasma glucose | 3p MACE | 0 |
| Florkowski 2001; | 447; | Est. ASCVD or | HbA1C; Smoking | CVM | 1 |

| | | | | | |
|---|---|---|---|---|---|
| Prosp. Cohort; New Zealand | from 1989; FUP mean 10 | High CVD risk | | | |
| Abu-Lebdeh 2001; Prosp. Cohort; United States | 449; 1968 to 1982; FUP mean 13 | No ASCVD | Age; Smoking; Triglycerides; Fasting glucose | CHD | 0 |
| Rutter 2002; Prosp. Cohort; United Kingdom | 86; NR; FUP mean 2.8 | No ASCVD | Microalbuminuria; Silent MI | CHD, CVM, Angina | 0 |
| Nazimek-Siewniak 2002; Prosp. Cohort; Poland | 2175; 1980 to 1994; NR | Any CVD risk | arterial blood pressure | CHD, Angina | 1 |
| DeLorenzo 2002; Prosp. Cohort; Brazil | 180; 1992 to 1997; FUP mean 3 | No ASCVD with Mod. CVD risk | Stress myocardial perfusion single-photon emission computed tomography (SPECT) | CHD, CVM | 0 |
| Faglia 2002; Prosp. Cohort; Italy | 735; 1993 to 1993; FUP mean 5 | No ASCVD | exercise treadmill test (ETT) | CHD, CVM, Angina | 0 |
| Christiansen 2002; Prosp. Cohort; Denmark | 430; 1995 to 1998; FUP mean 2.4 | No ASCVD | Urinary orosomucoid excretion rate (UOER) | CVM | 0 |
| Pickup 2003; Prosp. Cohort; United Kingdom | 128; 1988 to 1989; FUP mean 12.8 | Any CVD risk | Serum sialic acid (marker of innate immunity) | CVM | 0 |
| Linnemann 2003; Prosp. Cohort; Germany | 475; 1990 to 1991; FUP mean 5.2 | No ASCVD | elevated heart rate; peripheral arterial disease; serum creatinine; smoking; Prolong QTc interval | CVM | 0 |
| Lu 2003; Prosp. Cohort; United States | 2108; 1989 to 1992; FUP mean 9 | No ASCVD | non-HDL cholesterol | 3p MACE, HF | 1 |
| Gazzaruso 2003; Retrosp. Cohort; Italy | 95; NR; FUP median 0.5 | ASCVD | Lipoprotein(a); apolipoprotein(a) phenotypes | CHD, Revasc | 0 |

| | | | | | |
|---|---|---|---|---|---|
| Cardoso 2003; Prosp. Cohort; Brazil | 471;1994 to 1996; FUP median 4.75 | No ASCVD with Mod. CVD risk | QT interval | 3p MACE, Revasc, HF, PAD | 0 |
| Soinio 2004;Prosp. Cohort; Finland | 830;1999 to 2006. NR | No ASCVD | elevated plasma homocysteine levels | CHD, CVM | 0 |
| Schulze 2004; Prosp. Cohort; United States | 921; 1989 to 1990; FUP mean 7.4 | No ASCVD | HbA1C; non-HDL cholesterol | CHD, Revasc | 1 |
| Schulze 2004; Prosp. Cohort; United States | 746; 1993 to 1994; FUP mean 5 | No ASCVD | C-reactive protein (CRP) | CHD, Stroke, Revasc | 0 |
| Jiang 2004; Prosp. Cohort; United States | 746; 1986 to 1994; FUP mean 6 | No ASCVD | non-HDL cholesterol; apolipoprotein B | CHD, Stroke, Revasc | 0 |
| Fukushima 2004; Prosp. Cohort; Japan | 240; NR; FUP median 0.85 | Any CVD risk | Remnant-like lipoprotein particles (RPL) cholesterol | CHD, CVM, Revasc, Angina | 0 |
| Fox 2004; Prosp. Cohort; United States | 588; 1954 to 1995; FUP mean 12 | No ASCVD | Diabetes duration | CHD, CVM | 1 |
| Gimeno-Orna 2004; Prosp. Cohort; Spain | 423; 1994 to 1998; FUP mean 4.7 | Any CVD risk | Mild renal insufficiency | 3p MACE, Revasc, Angina, PAD | 1 |
| Anavekar 2004; Prosp. Cohort; Multinational | 1715; 1996 to 1999; FUP mean 2.6 | Est. ASCVD or High CVD risk | Urinary urine albumin/creatinine ratio; History of CV disease | 3p MACE, Revasc, HF, PAD | 1 |
| Bruno 2004; Prosp. Cohort; Italy | 1565; 1991 to 1992; FUP median 8.21 | No ASCVD | Metabolic syndrome | CVM | 0 |
| Saely 2005; Prosp. Cohort; Austria | 756; 1999 to 2000; FUP mean 2.3 | Est. ASCVD or High CVD risk | Insulin resistance; Metabolic syndrome (MetS) | 3p MACE, Revasc, HF, ACM | 0 |
| Nitenberg 2005; Prosp. Cohort; France | 59; 1994 to 1997; FUP mean 9.3 | No ASCVD | coronary artery constriction with angiographically normal coronary arteries (cold-pressor test) | 3p MACE, Revasc, Angina | 0 |

| | | | | | |
|---|---|---|---|---|---|
| Rana 2005; Prosp. Cohort; United Kingdom | 192; 1982 to 1988; FUP mean 12.7 | Est. ASCVD | QT interval | CVM | 0 |
| LeFeuvre 2005; Prosp. Cohort; France | 100; 1999 to 2001; FUP mean 2 | No ASCVD with Mod. CVD risk | Dobutamine echocardiography (DE); Single photon emission computed tomography (SPECT) | CHD, CVM, Revasc | 0 |
| Friedman 2005; Prosp. Cohort; Multinational | 1575; 1996 to 1999; FUP mean 2.6 | Est. ASCVD or High CVD risk | Total serum homocysteine (tHcy) | 3p MACE, Revasc, PAD | 0 |
| Bruno 2005; Prosp. Cohort; Italy | 1565; 1991 to 1992; FUP mean 11 | No ASCVD | AER (albumin excretion rate); Fibrinogen | CVM | 0 |
| Bruce 2005; Prosp. Cohort; Australia | 1273; 1993 to 1996; FUP mean 7.8 | No ASCVD | Depression | CVM | 0 |
| Bernard 2005; Prosp. Cohort; France | 229; NR; FUP mean 5 | No ASCVD with High CVD risk | Carotid IMT | 3p MACE, Angina | 0 |
| Cockcroft 2005; Retrosp. Cohort; United Kingdom | 2911; from 1996; FUP mean 4 | Any CVD risk | Pulse pressure (PP) | CHD | 0 |
| Christiansen 2005; Prosp. Cohort; Denmark | 430; 1995 to 1998; FUP mean 4.9 | Any CVD risk | Urinary orosomucoid excretion rate (UOER) | CVM | 0 |
| Thanyasiri 2006; Retrosp. Cohort; Australia. | 327; 1996 to 2000; FUP mean 2.1 | Est. ASCVD or High CVD risk | Extent of CAD; Renal complications of diabetes; Medical diabetic treatment | CHD | 0 |
| Yoshimura 2006; Prosp. Cohort; Japan | 129; 1997 to 2003; FUP mean 4.8 | No ASCVD | Ankle-brachial index (ABI) | 3p MACE, Revasc, PAD | 0 |
| Sultan 2006; Prosp. Cohort; France | 447; 1999 to 2002; FUP median 2.1 | No ASCVD | age > median; Low-density lipoprotein cholesterol 3.35 mmol/l; abnormal MPI | CHD, CVM, HF, Angina | 0 |
| Seyoum 2006; Prosp. Cohort; | 468; 1993 to NR; | Any CVD risk | Exercise capacity (peak V02) | 3p MACE, HF, PAD | 0 |

| | | | | | |
|---|---|---|---|---|---|
| United States | FUP mean 5 | | | | |
| Saely 2006;<br>Prosp. Cohort;<br>Austria | 587;<br>1999 to 2000;<br>FUP mean 3.9 | Est. ASCVD or High CVD risk | Lipoprotein(a) | 3p MACE, Revasc, HF | 0 |
| Nagamachi 2006;<br>Retrosp. Cohort;<br>Japan | 144;<br>1995 to 1998;<br>FUP mean 7.2 | Any CVD risk | cardiac I-123metaiodobenzylguanidine (MIBG) imaging | CHD | 0 |
| Linnemann 2006;<br>Prosp. Cohort;<br>Germany | 292;<br>NR;<br>FUP mean 5.3 | Any CVD risk | C-reactive protein (CRP) | 3p MACE, PAD, ACM | 0 |
| Jimenez-Corona 2006;<br>Prosp. Cohort;<br>United States | 1605;<br>1965 to 1992;<br>FUP median 14.1 | Est. ASCVD | electrocardiographic (ECG) abnormalities | CVM, ACM | 0 |
| Guzder 2006;<br>Prosp. Cohort;<br>United Kingdom | 428;<br>1996 to 1998;<br>FUP mean 4.2 | No ASCVD | Metabolic syndrome | CHD, Stroke, HF, Angina, PAD | 0 |
| Cavalot 2006;<br>Prosp. Cohort;<br>Italy | 529;<br>1995 to 2000;<br>FUP mean 5 | No ASCVD | Postprandial blood glucose | 3p MACE, Revasc, Angina, PAD | 0 |
| Anand 2006;<br>Prosp. Cohort;<br>United Kingdom | 510;<br>NR;<br>FUP median 2.2 | No ASCVD | Coronary artery calcium (CAC) score | 3p MACE, Revasc | 0 |
| Anand 2006a;<br>Prosp. Cohort;<br>United Kingdom | 510;<br>NR;<br>FUP mean 1.5 | No ASCVD with Mod. CVD risk | Osteoprotogerin | 3p MACE, Revasc | 0 |
| Busch 2006;<br>Prosp. Cohort;<br>Multinational | 450;<br>1996 to 1998;<br>FUP mean 2.6 | Nephropathy | N(epsilon)-carboxymethyllysine (CML) | 3p MACE, Revasc, HF, PAD | 0 |
| Azevedo 2006;<br>Prosp. Cohort;<br>Brazil | 135;<br>from 1996;<br>FUP mean 3.4 | No ASCVD | WHO cardiovascular questionnaire | CHD, CVM, HF | 0 |
| Wallander 2007;<br>Prosp. Cohort;<br>Sweden | 575;<br>1998 to 2003;<br>FUP median 2.2 | Est. ASCVD | IGF- binding protein (IGFBP)-1 | 3p MACE | 0 |
| Yang 2007;<br>Prosp. Cohort;<br>China | 7067;<br>1995 to 2005;<br>FUP median 5.4 | No ASCVD | Albuminuria | CHD | 1 |

| | | | | | |
|---|---|---|---|---|---|
| Pintó 2007;<br>Prosp. Cohort;<br>Spain | 838;<br>1999 to 2002;<br>FUP mean 2 | No ASCVD | HbA1C (<7.5% vs. >=7.5%); Albuminuria (<30 mg/24 h vs. >=30 mg/24 h); Age (<=65 years vs. >65 years); Obesity (BMI <30 vs. >=30); History of stroke (No vs. Yes); History of angina (No vs. Yes); LDL-C (<135 mg/dL vs. >=135 mg/dL); Retinopathy (No vs. Yes); Non-HDL cholesterol (<165 mg/dL vs. >=165 mg/dL); History of peripheral artery disease (No vs. Yes); Sed; Triglycerides (<200 mg/dL vs. >=200 mg/dL) | CHD, Stroke, Angina, PAD | 0 |
| Nag 2007;<br>Prosp. Cohort;<br>United Kingdom | 3288;<br>from 1994;<br>FUP median 10.5 | No ASCVD | estimated glomerular filtration rate (eGFR) | CVM | 1 |
| Meerwaldt 2007;<br>Prosp. Cohort;<br>Netherlands | 69;<br>NR;<br>FUP mean 5 | Any CVD risk | Skin autofluorescence | CHD, ACM | 0 |
| Filippella 2007;<br>Prosp. Cohort;<br>Italy | 969;<br>2003 to 2003;<br>FUP mean 1.525 | No ASCVD | Ankle brachial index (ABI) | CHD, Angina | 0 |
| Eijkelkamp 2007;<br>Prosp. Cohort;<br>Multinational | 2708;<br>1995 to 1998;<br>FUP mean 4 | Any CVD risk | Serum creatinine | 3p MACE | 1 |
| Eguchi 2007;<br>Prosp. Cohort;<br>Japan | 157;<br>1996 to 2001;<br>FUP mean 4.9 | No ASCVD with Mod. CVD risk | Left ventricular (LV) mass and relative wall thickness (echocardiography) | 3p MACE, HF, Angina, Renal | 0 |
| deSantiago 2007;<br>Prosp. Cohort;<br>Spain | 221;<br>from 1994;<br>FUP mean 5.9 | No ASCVD with Mod. CVD risk | ECG abnormalities | CHD, Stroke, HF, Angina, PAD, ACM | 0 |
| Chen 2007;<br>Prosp. Cohort;<br>Taiwan | 556;<br>2001 to 2002;<br>FUP mean 3.7 | No ASCVD with Mod. CVD risk | Subclinical hypothyroidism | CHD, Stroke, HF, Angina | 0 |
| Angiolillo 2007;<br>Prosp. Cohort;<br>Spain | 173;<br>2003 to 2005;<br>FUP mean 2 | Est. ASCVD | high platelet reactivity (HPR) | 3p MACE | 0 |
| Avogaro 2007;<br>Prosp. Cohort;<br>Italy | 11644;<br>1998 to 1999;<br>FUP mean 4 | No ASCVD | Sex | CHD, Revasc | 1 |
| Lim 2008;<br>Prosp. Cohort; | 343;<br>from 2001; | No ASCVD with High CVD risk | adiponectin; resistin | 3p MACE, Revasc, Angina, Renal | 0 |

| | | | | | |
|---|---|---|---|---|---|
| Korea | FUP mean 3.5 | | | | |
| Kenealy 2008; Prosp. Cohort; New Zealand | 48444; 2000 to 2005; FUP median 2.4 | No ASCVD with Mod. CVD risk | Ethnicity | CHD, Stroke, PAD | 1 |
| Gazzaruso 2008; Prosp. Cohort; Italy | 291; 1998 to 2006; FUP mean 1.97 | ASCVD | Erectile dysfunction (International Index Erectile Function-5 Questionnaire) | 3p MACE, Revasc, HF, Angina, PAD | 0 |
| Gasior 2008; Prosp. Cohort; Poland | 1310; NR; FUP mean 1 | Est. ASCVD | Blood glucose level on admission | CHD, ACM | 0 |
| Eguchi 2008; Prosp. Cohort; Japan | 1268; 1990 to 1998; FUP mean 5.7 | No ASCVD | Ambulatory blood pressure | 3p MACE | 1 |
| Giorda 2008; Retrosp. Cohort; Italy | 3174; 1998 to 1999; FUP median 4 | Any CVD risk | Older age; Male sex; Insulin use; Serum triglycerides | CHD, Stroke, Revasc, Angina, PAD | 0 |
| Chacko 2008; Prosp. Cohort; United States | 871; 1991 to 1993; FUP median 5 | No ASCVD | Heart rate recovery (HRR) | CHD, CVM, HF, PAD | 0 |
| Elley 2008; Prosp. Cohort; New Zealand | 48444; 2001 to 2005; FUP median 2.4 | Any CVD risk | HbA1C | CHD, Stroke, PAD | 1 |
| Elkeles 2008; Prosp. Cohort; United Kingdom | 589; 2000 to 2003; FUP median 4 | No ASCVD | Coronary artery calcium (CAC) score | 3p MACE, Angina | 0 |
| Sone 2009; Prosp. Cohort; Japan | 1424; 1996 to 1996; FUP mean 8 | No ASCVD | Excess waist circumference (WC) | CHD, Stroke | 0 |
| Peng 2009; Prosp. Cohort; China | 265; 2004 to 2005; FUP mean 1 | Est. ASCVD | serum levels of endogenous secretory receptor for advanced glycation endproducts (esRAGE) | 3p MACE, Revasc | 0 |
| Oliveira 2009; Retrosp. Cohort; Brazil | 193; 2001 to 2006; FUP median 2.42 | Any CVD risk | Exercise echocardiography | CHD, CVM, Revasc | 0 |
| Nilsson 2009; | 13087; | No ASCVD | smoking; smoking | CHD, Stroke, | 1 |

| | | | | | |
|---|---|---|---|---|---|
| Prosp. Cohort;<br>Sweden | 1996 to 2003;<br>FUP mean 5.7 | | | ACM | |
| Lutgers 2009;<br>Prosp. Cohort;<br>Netherlands | 973;<br>2001 to 2002;<br>FUP median 3.1 | Any CVD risk | Skin autofluorescence | CVM | 0 |
| Ikeda 2009;<br>Prosp. Cohort;<br>Japan | 88;<br>1996 to 1996;<br>FUP mean 10 | Any CVD risk | Serum paraoxonase (PON1) | CHD, Stroke, HF, PAD | 0 |
| Eguchi 2009;<br>Prosp. Cohort;<br>Japan | 300;<br>1990 to 1998;<br>FUP mean 4.5 | Any CVD risk | Night time blood pressure variability | 3p MACE | 0 |
| Georgoulias 2009;<br>Prosp. Cohort;<br>Greece | 258;<br>2002 to 2004;<br>FUP mean 2.6 | No ASCVD with High CVD risk | Heart rate recovery (HRR) | CHD, CVM,<br>Revasc | 0 |
| Charlton-Menys 2009;<br>Prosp. Cohort;<br>United Kingdom, Ireland | 2627;<br>1997 to 2001;<br>FUP median 3.9 | No ASCVD with High CVD risk | ApoB:A-I ratio | CHD, Stroke | 0 |
| deGalan 2009;<br>Prosp. Cohort;<br>Multinational | 11132;<br>2001 to 2003;<br>FUP median 5 | Est. ASCVD or High CVD risk | Cognitive dysfunction | 3p MACE, ACM | 0 |
| Bosevski 2009;<br>Prosp. Cohort;<br>Macedonia | 82;<br>NR;<br>FUP mean 1 | No ASCVD | Peripheral impaired endothelial-dependent vasodilation of brachial artery<br>(B-mode ultrasound) | 3p MACE, HF,<br>Angina | 0 |
| Clarke 2009;<br>Prosp. Cohort;<br>Multinational | 7348;<br>from 1996;<br>FUP mean 2.4 | Any CVD risk | EQ-5D index score | 3p MACE, Angina | 0 |
| Schimke 2010;<br>Prosp. Cohort;<br>Australia | 1179;<br>1993 to 1996;<br>FUP mean 10.3 | No ASCVD | Helicobacter pylori cytotoxin-associated gene-A antibodies | 3p MACE | 0 |
| Venskutonyte 2010;<br>Prosp. Cohort;<br>Multinational | 1253;<br>1996 to 2003;<br>FUP median 2.1 | Est. ASCVD | age; gender | 3p MACE, ACM | 1 |
| Vengen 2010;<br>Prosp. Cohort; | 200;<br>1984 to 1986; | Any CVD risk | Lactoferrin | CHD | 0 |

| | | | | | |
|---|---|---|---|---|---|
| Norway | NR | | | | |
| Ting 2010;<br>Prosp. Cohort;<br>Hong Kong | 4521;<br>1996 to 2005;<br>FUP median 4.9 | No ASCVD | HDL-C, mmol/L; LDL-C ≥3.0 mmol/L vs. <3.0 mmol/L | CHD, Stroke | 1 |
| Zoppini 2010;<br>Prosp. Cohort;<br>Italy | 3084;<br>2001 to 2002;<br>FUP mean 4.9 | Any CVD risk | triglyceride to high-density lipoprotein cholesterol (TG/HDL-C) ratio | CVM | 0 |
| Reinhard 2010;<br>Prosp. Cohort;<br>Denmark | 283;<br>1987 to 1987;<br>FUP median 16.8 | Any CVD risk | Osteoprotegerin | CVM | 0 |
| Nakamura 2010;<br>Prosp. Cohort;<br>Japan | 191;<br>2004 to 2007;<br>FUP median 2.12 | Any CVD risk | Brachial-ankle pulse wave velocity (baPWV) | CHD, Revasc, HF, ACM | 0 |
| Mellbin 2010;<br>Prosp. Cohort;<br>Sweden | 393;<br>1999 to 2003;<br>FUP median 2.5 | Any CVD risk | insulin growth factor binding protein-1 (IGFBP-1);<br>C-terminal provasopressin (copeptin) | 3p MACE | 0 |
| Lin 2010;<br>Prosp. Cohort;<br>United States | 4623;<br>2000 to 2002;<br>FUP mean 4.5 | No ASCVD | Major depression | CHD, Stroke, ACM | 0 |
| Kamoi 2010;<br>Prosp. Cohort;<br>Japan | 400;<br>1999 to 2005;<br>FUP mean 3.5 | Any CVD risk | Home blood pressure | CHD, Stroke | 1 |
| Juutilainen 2010;<br>Prosp. Cohort;<br>Finland | 833;<br>1982 to 1984;<br>FUP mean 18 | Any CVD risk | Thoracoabdominal calcifications in native radiograms | CVM, ACM | 0 |
| Eguchi 2010;<br>Prosp. Cohort;<br>Japan | 200;<br>1996 to 2002;<br>FUP mean 5.56 | No ASCVD | Heart rate variability (HRV) during sleep | 3p MACE | 0 |
| Burgess 2010;<br>Prosp. Cohort;<br>Multinational | 9795;<br>1998 to 2003;<br>FUP mean 5 | Est. ASCVD or High CVD risk | ECG changes (silent MI) | CHD | 0 |
| Zimering 2011; | 399; | Any CVD risk | baseline plasma bFGF immunoreactivity (IR) | CHD | 0 |

| | | | | | |
|---|---|---|---|---|---|
| Prosp. Cohort;<br>United States | 2000 to 2003;<br>FUP mean 5.6 | | | | |
| Sone 2011;<br>Prosp. Cohort;<br>Japan | 1771;<br>1995 to 1996;<br>FUP median 7.86 | No ASCVD | LDL Cholesterol; serum log-transformed triglyceride level | CHD | 0 |
| Pfister 2011;<br>Prosp. Cohort;<br>Multinational | 5231;<br>2001 to 2002;<br>FUP mean 2.9 | No ASCVD with High CVD risk | Electrocardiographic (ECG) signs | CHD, Stroke, ACM | 0 |
| McMurray 2011;<br>Prosp. Cohort;<br>United States | 3847;<br>2004 to 2007;<br>FUP median 2.4 | No ASCVD with DKD | log urine protein/ creatinine ratio; Age; prior HF; serum N-terminal pro B-type natriuretic peptide; C-reactive protein; abnormal electrocardiogram; troponin T | 3p MACE, HF | 0 |
| Lievre 2011;<br>Prosp. Cohort;<br>France | 631;<br>2000 to 2005;<br>FUP mean 3.5 | No ASCVD | screening for silent ischemia with bicycle exercise test or dipyridamole single photon emission computed tomography | CHD, Stroke, HF, ACM | 0 |
| Johnston 2011;<br>Retrosp. Cohort;<br>United States | 860845;<br>2006 to 2007;<br>FUP mean 3.5 | Any CVD risk | hypoglycemic events (ICD codes) | CHD, Revasc, Angina | 0 |
| Drury 2011;<br>Prosp. Cohort;<br>Multinational | 9795;<br>2000 to 2003;<br>FUP median 5 | No ASCVD | Lower estimated GFR (eGFR) | CHD, Stroke | 1 |
| Cavalot 2011;<br>Prosp. Cohort;<br>Italy | 505;<br>from 1995;<br>FUP mean 14 | No ASCVD | A1C; Postprandial blood glucose | 3p MACE, Revasc, Angina, PAD | 0 |
| Eliasson 2011;<br>Prosp. Cohort;<br>Sweden | 18673;<br>2002 to 2003;<br>FUP mean 4.8 | Any CVD risk | Non-HDL:HDL | CHD | 0 |
| Cosson 2011;<br>Prosp. Cohort;<br>France | 688;<br>1992 to 2006;<br>FUP mean 5.4 | No ASCVD | Silent myocardial ischemia (stress myocardial scintigraphy) | 3p MACE, Revasc, HF, PAD | 0 |

| | | | | | |
|---|---|---|---|---|---|
| Aboyans 2011;<br>Retrosp. Cohort;<br>France | 403;<br>1999 to 2000;<br>FUP mean 6.5 | Est. ASCVD or High CVD risk | Ankle-brachial index (ABI) | CHD, Stroke, ACM | 0 |
| Ningshen 2012;<br>Prosp. Cohort;<br>India | 100;<br>2009 to 2010;<br>FUP mean 2 | No ASCVD | QTc interval prolongation | CVM | 0 |
| Bouchi 2012;<br>Prosp. Cohort;<br>Japan | 689;<br>2003 to 2008;<br>FUP median 3.3 | No ASCVD | HbA1C variability | CHD, Stroke, Revasc, Angina | 0 |
| Vepsäläinen 2012;<br>Prosp. Cohort;<br>Finland | 881;<br>1982 to 1984;<br>FUP mean 18 | Any CVD risk | Proteinuria + systolic blood pressure <130 mmgHg | CVM, ACM | 0 |
| Yamada 2012;<br>Retrosp. Cohort;<br>Japan | 371;<br>2005 to 2006;<br>FUP median 2.8 | No ASCVD | Proliferative diabetic retinopathy | CHD | 1 |
| Sone 2012;<br>Prosp. Cohort;<br>Japan | 1771;<br>1995 to 1996;<br>FUP median 7.86 | No ASCVD | HDL-cholesterol (HDLC); total cholesterol (TC); LDL-cholesterol (LDLC);<br>non-HDLC in Men; LDLC/HDLC ratio; TC/HDLC ratio in Men;<br>TG/HDLC ratio; triglycerides (TGs) in women | CHD, Angina | 0 |
| Ruggenenti 2012;<br>Prosp. Cohort;<br>Italy | 1208;<br>2001 to 2007;<br>FUP median 9.12 | Any CVD risk | Measurable albuminuria | CHD, Stroke, Revasc, HF, PAD | 1 |
| Resl 2012;<br>Prosp. Cohort;<br>Austria | 494;<br>2005 to 2007;<br>FUP mean 12.8 | Any CVD risk | Uric acid | CHD, HF | 0 |
| Panero 2012;<br>Prosp. Cohort;<br>Italy | 1509;<br>1991 to 1992;<br>FUP median 13.7 | Any CVD risk | Uric acid | CVM | 0 |
| Otto 2012;<br>Prosp. Cohort;<br>German | 48;<br>NR;<br>FUP mean 2 | Est. ASCVD | Real-time microembolization during PCI (‘‘HITS’’) detected by an intracoronary Doppler | CHD, Revasc, ACM | 0 |

| | | | | | |
|---|---|---|---|---|---|
| Lopes-Virella 2012;<br>Prosp. Cohort;<br>United States | 907;<br>2000 to 2003;<br>FUP mean 3.7 | Est. ASCVD or High CVD risk | levels of malondialdehyde (MDA) - LDL | 3p MACE, HF, PAD | 0 |
| Khalili 2012;<br>Prosp. Cohort;<br>Iran | 1010;<br>1999 to 2001;<br>FUP median 8.4 | No ASCVD | Waist-to-height ratio (WHtR); Waist-to-hip ratio (WHR) | 3p MACE, Angina | 0 |
| Katakami 2012;<br>Prosp. Cohort;<br>Japan | 85;<br>1997 to 2011;<br>FUP median 7.9 | No ASCVD with High CVD risk | Calibrated integrated backscattered (IBS) ultrasonic tissue characterization of carotid plaque | 3p MACE, Angina | 0 |
| Lau 2012;<br>Prosp. Cohort;<br>China | 151;<br>2005 to 2006;<br>FUP mean 5.08 | No ASCVD with Mod. CVD risk | Coronary artery calcium score (CACS) | 3p MACE, Revasc, PAD | 0 |
| Iijima 2012;<br>Prosp. Cohort;<br>Japan | 938;<br>2000 to 2002;<br>FUP mean 5.43 | Any CVD risk | Total activity score (TAS) | CHD, Stroke, HF, Angina, PAD | 0 |
| Fumisawa 2012;<br>Retrosp. Case-control;<br>Japan | 800;<br>2002 to 2010;<br>NR | Any CVD risk | non-HDL cholesterol | CHD, Revasc | 1 |
| Carnethon 2012;<br>Prosp. Cohort;<br>United States | 2625;<br>1987 to 2011;<br>NR | Any CVD risk | Weight | 3p MACE | 1 |
| Hadaegh 2012;<br>Prosp. Cohort;<br>Iran | 926;<br>1999 to 2001;<br>FUP median 9.2 | Any CVD risk | ECG-determined coronary heart disease | CHD, Stroke, Angina | 0 |
| Estacio 2012;<br>Prosp. Cohort;<br>Multinational | 470;<br>1991 to 2003;<br>FUP mean 10 | No ASCVD | Urinary albumin excretion (UAE) | CVM | 1 |
| Dayan 2012;<br>Prosp. Cohort; | 128;<br>2008 to 2009; | Est. ASCVD or High CVD risk | Coronary artery calcium (CAC) score | CHD, Stroke, Revasc, HF, PAD | 0 |

| | | | | | |
|---|---|---|---|---|---|
| Turkey | FUP mean 3.05 | | | | |
| Zimering 2013;<br>Prosp. Cohort;<br>United States | 399;<br>from 2000;<br>FUP mean 6 | No ASCVD | Age; Prior CV event; Triglycerides; plasma basic fibroblast growth factor<br>(bFGF) levels | CHD, CVM,<br>Revasc | 0 |
| Yu 2013;<br>Prosp. Cohort;<br>United Kingdom | 4704;<br>2008 to 2009;<br>FUP mean 2 | Any CVD risk | HbA1C | CHD | 1 |
| Venskutonyte 2013;<br>Prosp. Cohort;<br>Multinational | 465;<br>2014 to 2017;<br>FUP median 2.1 | Est. ASCVD | Self-rated health | CHD, Stroke | 0 |
| Svensson 2013;<br>Prosp. Cohort;<br>Sweden | 66065;<br>2003 to 2006;<br>FUP mean 5.7 | Any CVD risk | albuminuria; renal impairment | CHD, ACM | 1 |
| Svendstrup 2013;<br>Prosp. Cohort;<br>Denmark | 430;<br>1995 to 1998;<br>FUP mean 10.5 | Any CVD risk | Increased urinary orosomucoid excretion rate (UOER) | CVM | 0 |
| Sone 2013;<br>Prosp. Cohort;<br>Japan | 1702;<br>1995 to 1996;<br>FUP median 8.05 | No ASCVD | leisure-time physical activity (LTPA); leisure-time physical activity (LTPA) | CHD | 0 |
| Twito 2013;<br>Retrosp. Cohort;<br>Israel | 2994;<br>2003 to 2011;<br>FUP mean 5.6 | Any CVD risk | HbA1C | CHD, Revasc,<br>ACM | 1 |
| Sidorenkov 2013;<br>Retrosp. Cohort;<br>Netherlands | 8455;<br>2007 to 2007;<br>FUP median 3.1-3.6 | Any CVD risk | Treated with glucose lowering drugs in patients with HbA1c >7 (%);<br>Treated with lipid lowering drugs | CHD, Stroke,<br>Revasc, PAD,<br>ACM | 1 |
| Silva 2013;<br>Prosp. Cohort;<br>Portugal | 150;<br>2005 to 2011;<br>FUP mean 2.975 | No ASCVD with<br>Mod. CVD risk | Apelin | CHD, CVM | 0 |
| Silva 2013; | 119; | Any CVD risk | Creatinine; Phosphorus | CHD, CVM | 0 |

| | | | | | |
|---|---|---|---|---|---|
| Prosp. Cohort; Portugal | 2006 to 2011; FUP mean 4 | | | | |
| Petretta 2013; Prosp. Cohort; Italy | 692; NR; FUP mean 1 | Est. ASCVD | Transient ischemic dilation (TID) | CHD, CVM, Revasc | 0 |
| Peters 2013; Prosp. Cohort; Australia | 940; 1982 to 2010; FUP mean 12.3 | Est. ASCVD | Hypomagnesemia | CHD, Revasc, Angina | 0 |
| Salles 2013; Prosp. Cohort; Brazil | 565; 2004 to 2008; FUP median 5.75 | Any CVD risk | Ambulatory blood pressure monitoring (ABPM) | 3p MACE, ACM | 0 |
| Nichols 2013; Prosp. Cohort; United States | 26636; 2002 to 2010; FUP mean 5.6 | No ASCVD | glycosylated hemoglobin (A1C); systolic blood pressure (SBP); low-density lipoprotein cholesterol (LDL-C) | CHD, Stroke | 1 |
| Ndrepepa 2013; Prosp. Cohort; Germany | 3705; 2000 to 2009; | Est. ASCVD | UA (uric acid) | CVM | 0 |
| Lin 2013; Prosp. Cohort; Taiwan | 628; 1996 to 2003; FUP median 10.4 | No ASCVD | Plasma YKL-40 concentration | CVM, ACM | 0 |
| Kawasaki 2013; Prosp. Cohort; Japan | 1620; from 1996; NR | No ASCVD | Diabetic retinopathy | CHD, Stroke | 1 |
| Irie 2013; Prosp. Cohort; Japan | 287; 2007 to 2009; FUP mean 4.6 | Est. ASCVD or High CVD risk | Gray-scale median (GSM) ultrasonic tissue characterization of carotid plaque | CHD, Stroke, Revasc, Angina, PAD | 0 |
| Gazzaruso 2013; Prosp. Cohort; Italy | 361; NR; FUP mean 1.9 | No ASCVD | Transcutaneous oxygen tension (TcPO2) | 3p MACE, Revasc, HF, Angina, PAD | 0 |

| | | | | | |
|---|---|---|---|---|---|
| Hayashi 2013;<br>Prosp. Cohort;<br>Japan | 4014;<br>2004 to 2005;<br>FUP mean 5.5 | No ASCVD with<br>Mod. CVD risk | HbA1C; Systolic blood pressure (SBP); LDL;<br>Non-HDL cholesterol; LDL/HDL ratio | CHD, Stroke,<br>Revasc, Angina | 0 |
| Hata 2013;<br>Prosp. Cohort;<br>Multinational | 8811;<br>2001 to 2002;<br>FUP median 2.4 | Est. ASCVD or<br>High CVD risk | Systolic blood pressure (SBP); Visit-to-visit varaibility in systolic blood pressure (SBP) | 3p MACE | 0 |
| Giovacchini 2013;<br>Prosp. Cohort;<br>Italy | 77;<br>2006 to 2008;<br>FUP median 4.1 | No ASCVD | Microalbuminuria | CHD, CVM | 1 |
| Hage 2013;<br>Prosp. Cohort;<br>United States, Canada | 518;<br>2000 to 2002;<br>FUP mean 4.7 | No ASCVD with<br>Mod. CVD risk | Heart rate response to adenosine (marker of cardiac autonomic neuropathy) | CHD, CVM | 0 |
| Cardoso 2013;<br>Prosp. Cohort;<br>Brazil | 565;<br>2004 to 2008;<br>FUP median 5.75 | Est. ASCVD or<br>High CVD risk | Carotid-femoral PWV (pulse wave velocity) | 3p MACE, Revasc,<br>HF, PAD, ACM | 0 |
| Araki 2013;<br>Prosp. Cohort;<br>Japan | 618;<br>1996 to 2000;<br>FUP median 1 | No ASCVD with<br>Mod. CVD risk | Urinary liver-type fatty acid–binding protein(L-FABP) | 3p MACE, Angina,<br>PAD | 0 |
| Alele 2013;<br>Prosp. Cohort;<br>United States | 936;<br>2000 to 2003;<br>FUP mean 3.7 | No ASCVD | 25-hydroxyitamin D | CHD, HF | 0 |
| Bruno 2013;<br>Prosp. Cohort;<br>Italy | 1825;<br>2000 to 2006;<br>FUP median 5.5 | No ASCVD | NT-proBNP | CVM | 0 |
| Azab 2013;<br>Retrosp. Cohort;<br>United States | 338;<br>2007 to 2011;<br>NR | No ASCVD | Neutrophil-lymphocyte ratio (NLR) | 3p MACE | 0 |
| Cox 2013; | 771; | Any CVD risk | Biventricular volume (BiVV) | CVM, ACM | 0 |

| | | | | | |
|---|---|---|---|---|---|
| Prosp. Cohort;<br>United States | 1998 to 2005;<br>FUP mean 8.4 | | | | |
| Yu 2014;<br>Prosp. Cohort;<br>United Kingdom | 4704;<br>2008 to 2009;<br>FUP median 2 | Any CVD risk | Blood pressure | CHD | 1 |
| Yiu 2014;<br>Retrosp. Case-control;<br>Hong Kong | 391;<br>NR;<br>FUP median 4.9 | No ASCVD | high-sensitivity Troponin I (hs-TnI) | 3p MACE, HF | 0 |
| Umamahesh 2014;<br>Retrosp. Cohort;<br>India | 249;<br>2000 to 2000;<br>FUP mean 11 | Any CVD risk | Smoking; Heavy alcohol consumption | CHD, Stroke,<br>Angina, PAD | 0 |
| Park 2014;<br>Prosp. Cohort;<br>South Korea | 557;<br>2008 to 2011;<br>FUP mean 2.8 | No ASCVD | coronary computed tomographic angiography (CCTA) | CHD, CVM,<br>Revasc | 0 |
| Lowe 2014;<br>Prosp. Cohort;<br>United States | 3865;<br>2001 to 2008;<br>FUP median 5 | Est. ASCVD or<br>High CVD risk | C-reactive protein (CRP); interleukin-6; fibrinogen | 3p MACE, Renal | 0 |
| Katakami 2014;<br>Prosp. Cohort;<br>Japan | 1040;<br>2005 to 2005;<br>FUP median 7.5 | No ASCVD | Brachial-ankle pulse wave velocity (PWV) | 3p MACE, Revasc,<br>HF, Angina, PAD | 0 |
| Duan 2014;<br>Prosp. Cohort;<br>China | 2197;<br>1994 to 1996;<br>FUP median 14.51 | NR | Middle cerebral artery (MCA) stenosis using transcranial doppler | 3p MACE | 0 |
| Chan 2014;<br>Prosp. Cohort;<br>China | 9509;<br>from 1995;<br>FUP median 7.5 | No ASCVD with<br>Mod. CVD risk | Age at diabetes diagnosis | CHD, Stroke, HF,<br>PAD | 0 |
| Davis 2014;<br>Prosp. Cohort;<br>United Kingdom | 4273;<br>1977 to 1997;<br>FUP median 18 | No ASCVD | Ethnicity | 3p MACE, HF,<br>Angina, PAD,<br>Renal | 1 |

| | | | | | |
|---|---|---|---|---|---|
| Bruno 2014;<br>Prosp. Cohort;<br>Italy | 1645;<br>from 1988;<br>FUP mean 6 | No ASCVD | NT-proBNP | CVM | 0 |
| Bianco 2014;<br>Prosp. Cohort;<br>Brazil | 323;<br>2001 to 2011;<br>FUP median 9.2 | Est. ASCVD | Prior myocardial infarction; GFR < 60 mL/min; Smoking; Long QTc interval; Left ventricular hypertrophy | CVM | 0 |
| Cortigiani 2014;<br>Prosp. Cohort;<br>Italy | 144;<br>2006 to 2009;<br>FUP mean 2.42 | No ASCVD with High CVD risk | Dopper-derived coronary flow velocity reserve (CFVR) of LAD | CHD, Revasc,<br>ACM | 0 |
| Cioffi 2014;<br>Prosp. Cohort;<br>Italy | 360; | No ASCVD | Subclinical systolic LV dysfunction (stress-corrected midwall shortening) | CVM | 0 |
| Chyun 2015; Prosp. Cohort;<br>United States, Canada | | | Cardiac autonomic neuropathy (CAN) | CHD, CVM,<br>Revasc, HF | 0 |
| vonScholten 2015;<br>Prosp. Cohort;<br>Denmark | 2007 to 2008;<br>FUP median 6.1 | Mod. CVD risk | NT-proBNP; Coronary artery calcium score (CAC) | 3p MACE, HF | 0 |
| Zafrir 2015;<br>Prosp. Cohort;<br>Israel | 600;<br>2006 to 2008;<br>FUP mean 6.7 | Any CVD risk | Cardiorespiratory fitness (quantified by percent predicted METs) | CHD, Stroke,<br>ACM | 0 |
| Yang 2015;<br>Prosp. Cohort;<br>China | 576;<br>2008 to 2010;<br>FUP mean 2 | No ASCVD | Glycated albumin (GA); endogenous secretory receptor for advanced glycation endproducts (esRAGE) | 3p MACE | 0 |
| Ong 2015;<br>Prosp. Cohort;<br>Australia | 9697;<br>1998 to 2000;<br>FUP median 5 | No ASCVD | Fibroblast growth factor 21 (FGF21) | 3p MACE, Revasc | 0 |
| Ravassa 2015;<br>Retrosp. Cohort; | 72;<br>2004 to 2005; | Est. ASCVD | Glucagon-like peptide-1 (GLP-1) | CHD, Stroke,<br>Revasc, HF, PAD | 0 |

| | | | | | |
|---|---|---|---|---|---|
| Spain | FUP median 6 | | | | |
| Monseu 2015;<br>Prosp. Cohort;<br>France | 1371;<br>2002 to 2011;<br>FUP mean 4.8 | Any CVD risk | albuminuria; eGFR; acute kidney injury (AKI) | CHD, Stroke,<br>Revasc, HF, PAD | 0 |
| Fragoso 2015;<br>Not specified;<br>Portugal | 119;<br>NR;<br>FUP mean 4.7 | No ASCVD<br>with<br>DKD | Insulin resistance | CHD, Stroke, HF,<br>PAD | 0 |
| Duan 2015;<br>Prosp. Cohort;<br>China | 2135;<br>1994 to 1996;<br>FUP median 14.53 | Any CVD risk | Sex | CHD, CVM | 1 |
| Heidari 2015;<br>Prosp. Cohort;<br>Iran | 2607;<br>2005 to 2013;<br>FUP median 8.5 | No ASCVD<br>with<br>Mod. CVD risk | 25-hydroxyvitamin D | CHD, CVM,<br>Angina | 0 |
| CeaSoriano 2015;<br>Prosp. Cohort;<br>United Kingdom | 57946;<br>2000 to 2005;<br>FUP mean 6.65 | Any CVD risk | eGFR | CHD, Stroke,<br>ACM | 1 |
| Everett 2015;<br>Prosp. Cohort;<br>United States | 2285;<br>from 2001;<br>FUP median 5 | Est. ASCVD | Cardiac troponin T | 3p MACE, ACM | 0 |
| Daka 2015;<br>Prosp. Cohort;<br>Sweden | 194;<br>1993 to 1994;<br>FUP mean 14.1 | Any CVD risk | Endogenous testosterone concentrations | CHD | 0 |
| Scirica 2016;<br>Prosp. Cohort;<br>Multinational | 12310;<br>2010 to 2013;<br>FUP median 2.1 | Est. ASCVD or<br>High CVD risk | N-terminal pro–B-type natriuretic peptide; high-sensitivity C-reactive<br>protein; high-sensitivity troponin T | 3p MACE, HF | 0 |
| Prentice 2016;<br>Retrosp. Cohort;<br>United States | 50861;<br>2000 to 2002;<br>FUP mean 3.3 | Any CVD risk | HbA1C variability | CHD, Stroke, HF,<br>Angina, PAD,<br>ACM | 0 |
| Wan 2016;<br>Retrosp. Cohort; | 115470;<br>2008 to 2008; | No ASCVD | HbA1C; urine albumin/creatinine ratio (ACR); BMI; DBP;<br>Smoking; TC/HDL-C ratio | 3p MACE, HF | 0 |

| | | | | | |
|---|---|---|---|---|---|
| China | FUP median 5.3 | | | | |
| vonScholten 2016;<br>Prosp. Cohort;<br>Denmark | 200;<br>2007 to 2008;<br>NR | No ASCVD with DKD | hsCRP; sICAM-1; sICAM-3; IL-6; IL-1-beta; IL-8; SAA; TNF-alpha;<br>sVCAM-1; sE-selectin; sP-selectin; thrombomodulin | 3p MACE, HF | 0 |
| vonScholten 2016;<br>Prosp. Cohort;<br>Denmark | 200;<br>2007 to 2008;<br>FUP median 6.1 | No ASCVD with DKD | Urinary hepatocyte growth facotr (HGF); Urinary adiponectin | 3p MACE, HF | 0 |
| Theilade 2016;<br>Prosp. Cohort;<br>Multinational | 4038;<br>2004 to 2007;<br>FUP median 2.4 | Any CVD risk | Pulse pressure (PP) | 3p MACE, HF,<br>Renal | 0 |
| Zafrir 2016;<br>Prosp. Cohort;<br>Israel | 594;<br>2006 to 2008;<br>FUP mean 6.6 | No ASCVD with High CVD risk | Abnromal heart rate recovery at 1 minute (<18 beats per minute);<br>Chronotropic incompetence (<80% heart rate reserve);<br>Resting tachycardia (>100 bpm) | CHD, Stroke,<br>ACM | 0 |
| Wijkman 2016;<br>Prosp. Cohort;<br>Sweden | 627;<br>2005 to 2008;<br>FUP median 7.9 | No ASCVD with Mod. CVD risk | Aortic pulse wave velocity (aPWV) | 3p MACE | 0 |
| Wijkman 2016;<br>Prosp. Cohort;<br>Sweden | 749;<br>2005 to 2008;<br>FUP median 7.8 | Any CVD risk | Diastolic orthostatic hypertension | 3p MACE | 0 |
| Strojek 2016;<br>Prosp. Cohort;<br>Multinational | 1115;<br>2004 to 2006;<br>FUP mean 2.7 | Est. ASCVD | Glucose (Average change across visits in mean 2-hour blood glucose level after meals) | 3p MACE, Revasc | 0 |
| Resl 2016;<br>Prosp. Cohort;<br>Austria | 746;<br>2005 to 2008;<br>FUP median 5 | Est. ASCVD | N-terminal pro B-type natriuretic peptide (NT-proBNP);<br>growth differentiation factor 15 (GDF-15);<br>high-sensitive troponin T (hs-TnT) | CHD, Stroke, HF,<br>PAD, ACM | 0 |
| Rawshani 2016;<br>Prosp. Cohort;<br>Multinational | 217364;<br>2003 to 2010;<br>FUP mean 5.6 | No ASCVD | Socioeconomic status | CVM | 0 |
| Nargesi 2016; | 2607; | No ASCVD | Wide pulse pressure (PP) | CHD | 0 |

| | | | | | |
|---|---|---|---|---|---|
| Prosp. Cohort;<br>Iran | NR;<br>FUP mean 7.8 | | | | |
| Masi 2016;<br>Prosp. Cohort;<br>United Kingdom | 489;<br>2001 to 2002;<br>FUP mean 10 | Any CVD risk | Plasma total anti-oxidant status (TAOS); leukocyte telomere length (LTL) | CHD | 0 |
| Lepojärvi 2016;<br>Prosp. Cohort;<br>Finland | 1243;<br>2011 to 2013;<br>FUP mean 2 | Est. ASCVD | B-type natriuretic peptide (BNP); highly sensitive C-reactive protein<br>(hs-CRP); galectin-3; soluble suppressor of tumorigenicity-2 (sST2);<br>highly sensitivity TroponinT (hs-TnT) | CHD, CVM, HF | 0 |
| Mohammedi 2016;<br>Prosp. Cohort;<br>Australia | 11120;<br>2001 to 2002;<br>FUP median 5 | Est. ASCVD or High CVD risk | Absence of Peripheral Pulses | 3p MACE | 0 |
| Hsieh 2016;<br>Prosp. Cohort;<br>China | 1291;<br>2004 to 2005;<br>FUP mean 0.4 | No ASCVD with<br>Mod. CVD risk | eGFR | CHD, Stroke,<br>ACM | 1 |
| Freemantle 2016;<br>Prosp. Cohort;<br>Multinational | 2999;<br>2010 to 2013;<br>FUP median 4.2 | Any CVD risk | HbA1C | 3p MACE | 1 |
| Chang 2016;<br>Retrosp. Cohort;<br>Taiwan | 452;<br>2005 to 2007;<br>FUP mean 5.8 | Any CVD risk | Ankle-brachial index (ABI) | CHD, Stroke,<br>Revasc, PAD,<br>ACM | 0 |
| Cha 2016;<br>Prosp. Cohort;<br>Korea | 206;<br>2001 to 2009;<br>FUP median 8.9 | Est. ASCVD | Cardiovascular autonomic neuropathy (CAN) | 3p MACE, HF,<br>PAD | 0 |
| Carlsson 2016;<br>Prosp. Cohort;<br>Sweden | 607;<br>2005 to 2008;<br>FUP mean 7.4 | No ASCVD | soluble tumor necrosis factor receptors 1 and 2 (sTNFR1 and sTNFR2) | 3p MACE | 0 |
| Cardoso 2016;<br>Prosp. Cohort; | 616;<br>2004 to 2010; | Est. ASCVD or High CVD risk | CRP (c-reactive protein) | 3p MACE, ACM | 0 |

| | | | | | |
|---|---|---|---|---|---|
| Brazil | FUP median 8.4 | | | | |
| Halon 2016;<br>Prosp. Cohort;<br>Israel | 630;<br>2006 to 2008;<br>FUP mean 6.6 | No ASCVD with High CVD risk | Coronary computed tomography (CT) angiography | CHD, CVM,<br>Angina | 0 |
| Faghihi-Kashani 2016;<br>Prosp. Cohort;<br>Iran | 2607;<br>from 2005;<br>FUP mean 7.2 | No ASCVD | 2 hour post-prandial glucose; Fasting insulin level | CHD, CVM,<br>Angina | 0 |
| Afsharian 2016;<br>Prosp. Cohort;<br>United States, Iran | 1198;<br>1999 to 2005;<br>FUP median 10.28 | No ASCVD | Hypertension; Hypercholesterolemia; High waist to hip ratio;<br>Fasting glucose | CHD, Stroke, HF,<br>Angina | 0 |
| Afarideh 2016;<br>Prosp. Cohort;<br>Iran | 2244;<br>1994 to 2015;<br>FUP mean 7.5 | Any CVD risk | ALT | CHD, Stroke,<br>Revasc | 0 |
| Brownrigg 2016;<br>Prosp. Cohort;<br>United Kingdom | 49027;<br>2008 to 2014;<br>FUP mean 5.5 | No ASCVD | Microvascular disease (retinopathy, peripheral neuropathy, nephropathy) | 3p MACE | 1 |
| Bates 2016;<br>Retrosp. Cohort;<br>United States | 3146;<br>1992 to 2008;<br>FUP median 9.1 | No ASCVD | Cardiac stress test | CHD, ACM | 0 |
| Van der Leeuw 2016;<br>Prosp. Cohort;<br>Netherlands | 1002;<br>1996 to 2006;<br>FUP median 9.2 | Any CVD risk | Adiponectin; basic fibroblast growth factor (bFGF); thrombomodulin;<br>sICAM-1; sICAM-3; sVCAM-1; sE-selectin; sP-selectin; serum amyloid A<br>(SAA); MMP-1; MMP-3; MMP-9; PLGF; sFit-1; VEGF; Osteocalcin;<br>Osteopontin; H-FABP; E-FABP; TIMP-1 | 3p MACE, HF,<br>PAD | 0 |
| Zobel 2017;<br>Prosp. Cohort;<br>Denmark | 200;<br>2007 to 2008;<br>FUP median 6.1 | No ASCVD with DKD | ankle–brachial index (ABI); Toe–brachial index (TBI) | 3p MACE, HF | 0 |
| Yang 2017; | 1466; | Est. ASCVD | Serum fibrinogen | 3p MACE, Revasc, | 0 |

| | | | | | |
|---|---|---|---|---|---|
| Prosp. Cohort; China | 2011 to 2015; FUP mean 1.68 | | | Angina | |
| Yang 2017; Prosp. Cohort; China | 1447; 2011 to 2015; FUP mean 1.7 | Any CVD risk | Triglyceride to HDL cholesterol ratio (TG/HDL-C) | 3p MACE, Revasc, Angina | 0 |
| Wolsk 2017; Prosp. Cohort; Multinational | 5525; 2010 to 2013; FUP median 2.1 | Est. ASCVD | B-type natriuretic peptide (BNP) | 3p MACE, HF, ACM | 0 |
| Takao 2017; Retrosp. Cohort; Japan | 646; 1995 to 1996; FUP median 15.6 | No ASCVD | postprandial hyperglycemia | 3p MACE, Revasc | 0 |
| Svensson 2017; Prosp. Cohort; Denmark | 24752; 2000 to 2012; FUP median 2.6 | Any CVD risk | HbA1c levels | 3p MACE | 1 |
| Sharma 2017; Prosp. Cohort; Multinational | 14671; 2008 to 2012; FUP median 3 | ASCVD | HbA1C; Age; BMI; Systolic blood pressure; History of cerebrovascular disease; History of PCI; NYHA class (heart failure); Sex; eGFR | CVM | 1 |
| Saulnier 2017; Prosp. Cohort; France | 1439; 2002 to 2012; FUP median 5.7 | No ASCVD | Urinary sodium concentration | CVM | 0 |
| Radholm 2017; Prosp. Cohort; Sweden | 635; 2005 to 2010; FUP median 7.1 | No ASCVD with Mod. CVD risk | sagital abdominal diameter (SAD) | 3p MACE | 0 |
| Li 2017; Prosp. Cohort; China | 320; 2012 to 2012; FUP mean 2 | Any CVD risk | pregnancyassociated plasma proteinA (PAPPA) | 3p MACE | 0 |
| Lee 2017; | 933; | No ASCVD | Coronary computed tomography angiography (CCTA) | CHD, Revasc, | 0 |

| | | | | | |
|---|---|---|---|---|---|
| Prosp. Cohort; Korea | 2006 to 2010; FUP mean 5.5 | | | ACM | |
| Hong 2017; Prosp. Cohort; China | 328; 2011 to 2012; FUP mean 1 | No ASCVD with High CVD risk | non-fasting remnant cholesterol | CHD, CVM, Revasc | 0 |
| Celis-Morales 2017; Prosp. Cohort; United Kingdom | 13373; 2007 to 2010; FUP median 4.9 | Any CVD risk | Grip strength | CHD, CVM | 0 |
| Fadini 2017; Retrosp. Cohort; Italy | 187; 2004 to 2014; FUP median 6.1 | No ASCVD | CD34+ and CD34+CD133+ stem cells | 3p MACE | 0 |
| Chang 2018; Prosp. Cohort; Taiwan | 1968; 2017 to 2018; FUP mean 1.225 | No ASCVD with Mod. CVD risk | First Harmonic (C1) of Radial Pulse | CHD, CVM, HF | 0 |
| Velho 2018; Prosp. Cohort; France | 2521; 2002 to 2011; FUP median 5 | Nephropathy | Plasma copeptin | CHD, CVM, Revasc, HF | 0 |
| Tobias 2018; Retrosp. Cohort; United States | 27041; 1993 to 2004; FUP mean 18.6 | No ASCVD | branched-chain amino acids (BCAAs; isoleucine, leucine, and valine) | CHD, Stroke | 0 |
| Thomas 2018; Retrosp. Case-control; Multinational | 3766; 2001 to 2003; FUP median 5 | Any CVD risk | Plasma 8-OH-Deoxyguanosine (8-oxo-20 -dG) | CVM | 0 |
| Yun 2018; Prosp. Cohort; South Korea | 578; 2000 to 2008; FUP mean 2.3 | No ASCVD | Cardiac autonomic neuropathy (CAN) | CHD, Stroke | 0 |
| Yu 2018; | 4704; | Any CVD risk | Total cholesterol to HDL cholesterol ratio (TC/HDL) | CHD, Stroke, PAD | 1 |

| | | | | | |
|---|---|---|---|---|---|
| Prosp. Cohort;<br>United Kingdom | 2008 to 2009;<br>FUP mean 2 | | | | |
| Seferovic 2018;<br>Prosp. Cohort;<br>Multinational | 6068;<br>2010 to 2013;<br>FUP median 2.1 | Any CVD risk | Neuropathy; Retinopathy; Longer diabetes duration | CHD | 1 |
| Scirica 2018;<br>Prosp. Cohort;<br>Multinational | 15760;<br>2010 to 2013;<br>FUP median 2.1 | Est. ASCVD or High CVD risk | Urinary albumin to creatinine ratio (UACR) | 3p MACE, HF | 1 |
| Savonitto 2018;<br>Prosp. Cohort;<br>Multinational | 7226;<br>2010 to 2012;<br>FUP median 2 | No ASCVD | glycated haemoglobin; age; Lack of coronary revascularization;<br>N-terminal pro-type natriuretic peptide (NT-proBNP); haemoglobin;<br>heart rate | CVM | 0 |
| RotbainCurovic 2018;<br>Prosp. Cohort;<br>Denmark | 200;<br>2007 to 2008;<br>FUP mean 6.1 | Est. ASCVD | Urinary levels of kidney injury molecule 1 (u-KIM-1);<br>Urinary levels of neutrophil gelatinase-associated lipocalin (u-NGAL) | 3p MACE, HF,<br>ACM | 0 |
| Sakai 2018;<br>Retrosp. Cohort;<br>Japan | 345;<br>2003 to 2011;<br>FUP mean 5 | Est. ASCVD | small dense LDL (sdLDL) | 3p MACE, Revasc,<br>HF | 0 |
| Oellgaard 2018;<br>Prosp. Cohort;<br>Denmark | 151;<br>from 1993;<br>FUP mean 19 | Nephropathy | urinary proteomics (CAD238, ACSP and ACSP75) | 3p MACE, Revasc,<br>PAD | 0 |
| Novo-RodrÃguez 2018;<br>Retrosp. Case-control;<br>Spain | 130;<br>2006 to 2007;<br>FUP mean 7 | Any CVD risk | Circulating levels of sclerostin | CVM | 0 |
| Rawshani 2018;<br>Prosp. Cohort;<br>Sweden | 271174;<br>1998 to 2012;<br>FUP median 5.7 | No ASCVD | A1c>7, SBP>140 or DBP>80, albuminuria, smoking, LDL >=2.5 | CHD, Stroke | 0 |
| Rasmussen 2018; | 198; | Any CVD risk | serum PRO-C (S-PRO-C6) | 3p MACE, HF | 0 |

| | | | | | |
|---|---|---|---|---|---|
| Prosp. Cohort;<br>Denmark | 2007 to 2008;<br>FUP median 6.5 | | | | |
| Kim 2018;<br>Prosp. Cohort;<br>Korea | 1302;<br>2003 to 2004;<br>FUP mean 11.1 | Any CVD risk | Hemoglobin glycation index (HGI) | CHD, Stroke | 0 |
| Keller 2018;<br>Prosp. Cohort;<br>Germany | 1034;<br>1998 to 2002;<br>FUP median 4.04 | Any CVD risk | Cardiac troponin T (TnT) | 3p MACE | 0 |
| Jin 2018;<br>Retrosp. Case-control;<br>China | 1282;<br>2011 to 2014;<br>FUP mean NR | Any CVD risk | Haemoglobin glycation index (HGI); Triglyceride glucose (TyG) index | 3p MACE, Revasc,<br>Angina | 0 |
| Hunt 2018;<br>Prosp. Cohort;<br>United States | 952;<br>2000 to 2003;<br>FUP median 1.94 | Any CVD risk | Connective tissue growth factor (CTGF/CCN2) | 3p MACE, HF,<br>PAD | 0 |
| Hu 2018;<br>Retrosp. Cohort;<br>China | 438;<br>2012 to 2013;<br>FUP mean 3 | No ASCVD with<br>High CVD risk | Platelet distribution width (PDW) | CHD, Revasc | 0 |
| Jha 2018;<br>Prosp. Cohort;<br>India | 122;<br>2017 to 2017;<br>FUP median 0.5 | Any CVD risk | soluble ST2 (sST2) | CHD, Stroke,<br>Angina | 0 |
| Jeevarethinam 2018;<br>Prosp. Cohort;<br>United Kingdom | 259;<br>2006 to 2010;<br>FUP mean 1.9 | No ASCVD with<br>Mod. CVD risk | Carotid intima-media thickness (IMT) | CHD, Stroke,<br>Revasc, ACM | 0 |
| Eguchi 2018;<br>Prosp. Cohort;<br>Japan | 4308;<br>2005 to 2012;<br>FUP mean 4 | Est. ASCVD or<br>High CVD risk | Home systolic blood pressure (SBP) | 3p MACE, PAD | 1 |
| Cha 2018; | 508; | No ASCVD | Heart rate variability (HRV) | CHD, Stroke | 0 |

| | | | | | |
|---|---|---|---|---|---|
| Prosp. Cohort;<br>South Korea | 2009 to 2010;<br>FUP median 7.8 | | | | |
| Cardoso 2018;<br>Prosp. Cohort;<br>Brazil | 654;<br>2004 to 2008;<br>FUP median 9.3 | Est. ASCVD or<br>High CVD risk | Glycemic variability (fasting glycemia and A1C standard deviation) | 3p MACE, ACM | 0 |
| Fadini 2018;<br>Retrosp. Cohort;<br>Italy | 100;<br>2011 to 2012;<br>FUP median 5.6 | Any CVD risk | p66Shc gene expression in PBMCs | 3p MACE | 0 |
| Apperloo 2018;<br>Retrosp. Cohort;<br>Netherlands | 1600;<br>2007 to 2013;<br>FUP median 4.4 | No ASCVD | Albuminuria (urinary albumin creatinine ratio [UACR]);<br>systolic blood pressure (SBP) | CHD, Stroke,<br>Revasc | 1 |
| Bell 2018;<br>Retrosp. Cohort;<br>Sweden | 9855;<br>1999 to 2009;<br>FUP median 4 | No ASCVD with<br>Mod. CVD risk | Blood pressure variability (BPV) | CVM | 0 |
| Cournot 2018;<br>Prosp. Cohort;<br>France | 1467;<br>2002 to 2011;<br>FUP median 5.33 | Any CVD risk | Redox biomarkers (Advanced oxidation protein products,<br>oxidative hemolysis inhibition assay, ischemia-modified albumin,<br>and total reductive capacity of plasma) | 3p MACE | 0 |
| Colombo 2018;<br>Prosp. Cohort;<br>United Kingdom | 2105;<br>1997 to 2001;<br>FUP median 2.1 | No ASCVD with<br>High CVD risk | Apolipoprotein CIII; NT-proBNP | CHD, Stroke | 0 |
| Tian 2019;<br>Retrosp. Cohort;<br>China | 164;<br>2011 to 2012;<br>FUP median 5 | No ASCVD | coronary computed tomography angiography (CTA) | CHD, CVM,<br>Revasc, Angina | 0 |
| Venuraju 2019;<br>Prosp. Cohort;<br>United Kingdom | 259;<br>2012 to NR;<br>FUP median 1.9 | No ASCVD | Diabetes duration; Coronary artery calcium score (CAC) | CHD, Revasc,<br>ACM | 0 |
| Østergaard 2019; | 1910; | Est. ASCVD | hemoglobin glycation index (HGI) | 3p MACE, HF | 0 |

| | | | | | |
|---|---|---|---|---|---|
| Prosp. Cohort;<br>Netherlands | 1996 to 2015;<br>FUP median 9.6 | | | | |
| Zhou 2019;<br>Prosp. Cohort;<br>China | 1332;<br>2011 to 2012;<br>FUP median 4.3 | Any CVD risk | proliferative diabetic retinopathy (PDR) | CHD | 1 |
| Yang 2019;<br>Prosp. Cohort;<br>China | 3009;<br>2008 to 2009;<br>FUP mean 8 | No ASCVD | Neck circumference | 3p MACE, Revasc, HF, Angina | 0 |
| Wong 2019;<br>Prosp. Cohort;<br>China | 2275;<br>2003 to 2014;<br>FUP median 4.25 | Est. ASCVD | B-type natriuretic peptide (BNP); High-sensitivity troponin I (hs-TnI) | 3p MACE, PAD | 0 |
| Yeboah 2019;<br>Prosp. Cohort;<br>United States | 10251;<br>2001 to 2005;<br>FUP mean 3.7 | Est. ASCVD or High CVD risk | Body weight variability | 3p MACE | 0 |
| Smáradóttir 2019;<br>Prosp. Cohort;<br>Multinational | 575;<br>1996 to 2003;<br>FUP mean 2.5 | Est. ASCVD | insulin-like growth factor binding protein-1 (IGFBP-1); Copeptin | 3p MACE | 0 |
| Siddique 2019;<br>Prosp. Cohort;<br>Multinational | 2176;<br>2001 to 2005;<br>FUP mean 4.3 | ASCVD | urine albumin– creatinine ratio (uACR) | CHD, Stroke, ACM | 1 |
| Rørth 2019;<br>Prosp. Cohort;<br>Multinational | 759;<br>2009 to 2012;<br>FUP median 2.25 | Est. ASCVD | N-terminal pro B-type natriuretic peptide (NT-proBNP); high-sensitive troponin T (hs-TnT) | CHD, CVM, HF, ACM | 0 |
| Rozing 2019;<br>Prosp. Cohort;<br>Denmark | 494;<br>1989 to 1991;<br>FUP mean 13 | Est. ASCVD | Changes in HbA1C during first 6 yrs after diagnosis | 3p MACE, PAD, ACM | 0 |
| Roumeliotis 2019; | 142; | Est. ASCVD | Carotid intima-media thickness (cIMT) | CHD, Stroke, PAD | 0 |

| | | | | | |
|---|---|---|---|---|---|
| Prosp. Cohort;<br>Greece | 2008 to 2015;<br>FUP mean 7 | | | | |
| Raghavan 2019;<br>Retrosp. Cohort;<br>United States | 329624;<br>2002 to 2003;<br>FUP mean 8 | Any CVD risk | HbA1C | CVM | 1 |
| Lin 2019;<br>Retrosp. Cohort;<br>Taiwan | 446;<br>2005 to 2007;<br>FUP mean 5.8 | Any CVD risk | ankle brachial index (ABI); Interarm systolic blood pressure difference<br>(IASBPD) | CHD, Stroke,<br>Revasc, PAD,<br>ACM | 0 |
| Lim 2019;<br>Prosp. Cohort;<br>China | 2008;<br>2012 to 2013;<br>FUP mean 2.3 | Any CVD risk | SUDOSCAN (electrochemical skin conductance test for<br>sudomotor dysfunction) | CHD, Stroke, PAD,<br>Renal | 0 |
| Liao 2019;<br>Prosp. Cohort;<br>Taiwan | 2324;<br>2017 to 2019;<br>FUP mean 1.8 | Any CVD risk | fourth harmonic amplitude of the radial pulse wave (C4CV) | CHD, Stroke,<br>Revasc, Angina | 0 |
| deVries 2019;<br>Prosp. Cohort;<br>Netherlands | 1265;<br>2003 to 2017;<br>FUP median 6.4 | Est. ASCVD or<br>High CVD risk | Plasma thyroid-stimulating hormone (TSH) level | 3p MACE, ACM | 0 |
| Chang 2019;<br>Retrosp. Cohort;<br>Taiwan | 452;<br>2005 to 2007;<br>FUP mean 5.8 | Any CVD risk | Upstroke time per cardiac cycle (UTCC) | CHD, Stroke,<br>Revasc, PAD,<br>ACM | 0 |
| Cardoso 2019;<br>Prosp. Cohort;<br>Brazil | 478;<br>2004 to 2008;<br>FUP median 10.8 | Est. ASCVD or<br>High CVD risk | Ultrasonographic parameters of carotid atherosclerosis,<br>intima-media thickness (CIMT) and plaques | 3p MACE, Revasc,<br>HF, PAD | 0 |
| Halon 2019;<br>Retrosp. Cohort;<br>Canada, Israel | 630;<br>NR;<br>FUP mean 9.2 | Est. ASCVD or<br>High CVD risk | Computed tomography angiography (CTA) plaque morphology | CHD | 0 |
| Cardona 2019; | 330; | No ASCVD with | trimethylamine N-oxide (TMAO) | 3p MACE, Revasc | 0 |

| | | | | | |
|---|---|---|---|---|---|
| Prosp. Cohort;<br>United States, Canada | 1999 to 2005;<br>FUP mean 4.7 | High CVD risk | | | |
| Anyanwagu 2019;<br>Retrosp. Cohort;<br>United Kingdom | 11074;<br>2006 to 2014;<br>NR | Nephropathy | Overt albuminuria (urinary albumin-creatinine ratio [ACR] | 3p MACE | 1 |
| Bonito 2019;<br>Prosp. Cohort;<br>Portugal | 78;<br>2010 to 2018;<br>NR | No ASCVD with<br>DKD | Resistin | CHD | 0 |
| Berkelmans 2019;<br>Prosp. Cohort;<br>Multinational | 389366;<br>2002 to 2012;<br>FUP mean 10 | No ASCVD | HbA1C; Macroalbuminuria; BMI; systolic PB; History of CVD;<br>Male sex; Diabetes duration; Insulin therapy; eGFR (mL/min/1.73 m2)b;<br>non-HDL-c; Current smoking | 3p MACE | 1 |
| Church 2019;<br>Prosp. Cohort;<br>New Zealand | 44416;<br>2002 to 2015;<br>FUP mean 5.4 | No ASCVD | eGFR | CHD, Stroke, HF,<br>Angina, PAD | 1 |
| Massardo 2020;<br>Prosp. Cohort;<br>Chile | 40;<br>2008 to 2010;<br>FUP mean 9.9 | No ASCVD | QT interval (QTc) | CVM | 0 |
| Lorenzo-Almorós 2020;<br>Prosp. Cohort;<br>Spain | 232;<br>2006 to 2014;<br>NR | Est. ASCVD | Plasma levels of Galectin-3 | CHD, Stroke | 0 |
| Cheng 2020;<br>Retrosp. Cohort;<br>Hong Kong | 5349;<br>1995 to 2017;<br>FUP mean 13.4 | Any CVD risk | Relative leukocyte telemore length | CHD, Stroke, HF,<br>PAD | 0 |
| Vavruch 2020;<br>Prosp. Cohort;<br>Sweden | 661;<br>2005 to 2008;<br>FUP mean 7.8 | Any CVD risk | adipocyte fatty acid binding protein (A-FABP); leptin | 3p MACE | 0 |
| Zhao 2020; | 798; | Est. ASCVD | triglycerideglucose index (TyG index) | CHD, Revasc, | 0 |

| | | | | | |
|---|---|---|---|---|---|
| Retrosp. Cohort;<br>China | 2015 to 2015;<br>FUP mean 3 | | | ACM | |
| Soejima 2020;<br>Prosp. Cohort;<br>Japan | 2494;<br>2002 to 2008;<br>FUP median 10.3 | No ASCVD | Proteinuria | 3p MACE, Angina,<br>PAD | 1 |
| Shin 2020;<br>Prosp. Cohort;<br>Multinational | 4091;<br>2010 to 2013;<br>FUP median 2.1 | Est. ASCVD | A1C; Left ventricular ejection fraction (LVEF) | CVM, HF | 0 |
| Sharma 2020;<br>Prosp. Cohort;<br>Multinational | 5154;<br>2009 to 2013;<br>FUP median 1.6 | Est. ASCVD | NT-proBNP; growth-differentiation-factor-15 (GDF-15);<br>galectin-3 (Gal-3); adiponectin; high-sensitivity troponin I | 3p MACE, HF | 0 |
| Pagidipati 2020;<br>Prosp. Cohort;<br>Multinational | 14534;<br>from 2008;<br>FUP median 3 | Clearly defined | Overweight/obesity | 3p MACE, Angina | 1 |
| Oshima 2020;<br>Retrosp. Cohort;<br>Multinational | 4401;<br>2014 to 2017;<br>FUP median 2.6 | Nephropathy | albuminuria | 3p MACE, HF | 1 |
| Ong 2020;<br>Prosp. Cohort;<br>Australia | 2000;<br>1998 to 2000;<br>FUP mean 5 | Est. ASCVD or<br>High CVD risk | plasma adipocyte-fatty acid binding protein lipocalin 2 (A-FABP);<br>lipocalin 2 (LCN2) | 3p MACE, Revasc | 0 |
| Qin 2020;<br>Prosp. Cohort;<br>China | 2356;<br>2014 to 2014;<br>FUP median 3.96 | Any CVD risk | atherogenic index of plasma (AIP) | 3p MACE, Revasc | 0 |
| Nam 2020;<br>Prosp. Cohort;<br>Korea | 624237;<br>2009 to 2010;<br>FUP mean 7.6 | Any CVD risk | body weight variability | CHD, Stroke,<br>ACM | 0 |
| Moosaie 2020; | 1202; | No ASCVD | Apo B; Apo B/Apo A1 ratio; Apo A1; Lipoprotein (Lp (a) | CHD, Revasc, | 0 |

| | | | | | |
|---|---|---|---|---|---|
| Retrosp. Case-control;<br>Iran | 2014 to 2019;<br>FUP median 5 | | | Angina | |
| Koo 2020;<br>Prosp. Cohort;<br>Korea | 149;<br>NR;<br>FUP median 4.14 | No ASCVD | Peripheral arterial endothelial dysfunction assessed using reactive<br>hyperemia-peripheral arterial tonometry (RH-PAT) | 3p MACE | 0 |
| Lapin 2020;<br>Retrosp. Cohort;<br>United States | 43945;<br>2009 to 2016;<br>FUP median 3.1 | Any CVD risk | Painful diabetic peripheral neuropathy (DPN) | CHD, Stroke, PAD | 1 |
| Drinkwater 2020;<br>Prosp. Cohort;<br>Australia | 1473;<br>2008 to 2011;<br>FUP mean 6.6 | Any CVD risk | Diabetic retinopathy (DR) | CHD, Stroke | 1 |
| Chen 2020;<br>Prosp. Cohort;<br>China | 733;<br>2008 to 2017;<br>FUP median 8 | No ASCVD | A1C; Systolic blood pressure (SBP); Triglycerides | 3p MACE | 0 |
| Ceriello 2020;<br>Prosp. Cohort;<br>Multinational | 7020;<br>2010 to 2013;<br>FUP median 3.1 | Est. ASCVD | HbA1C variability | 3p MACE | 0 |
| Carlsson 2020;<br>Prosp. Cohort;<br>Sweden | 813;<br>2005 to 2008;<br>FUP median 7.9 | Nephropathy | Growth differentiation factor 15 (GDF-15) | CHD, Stroke | 0 |
| Cardoso 2020;<br>Prosp. Cohort;<br>Brazil | 632;<br>2004 to 2008;<br>FUP median 11.3 | Est. ASCVD or<br>High CVD risk | Visit-to-visit blood pressure variability (BP-VVV) | 3p MACE, ACM | 0 |
| Biscetti 2020;<br>Prosp. Cohort;<br>Italy | 207;<br>2018 to 2019;<br>FUP mean 1 | Est. ASCVD | Omentin-1 | 3p MACE | 0 |
| Backhaus 2020; | 288; | Est. ASCVD | Killip classification; Global Longitudinal Strain for LV Function | 3p MACE | 0 |

| | | | | | |
|---|---|---|---|---|---|
| Prosp. Cohort; Germany | 2008 to 2011; FUP mean 1 | | | | |
| Cui 2020; Prosp. Cohort; China | 3149; 2016 to 2019; FUP mean 1 | Est. ASCVD or High CVD risk | PARP (poly[ADP-ribose]polymerase) activation in circulating leukocytes | 3p MACE, Revasc, ACM | 0 |
| Venuraju 2021; Retrosp. Cohort; United Kingdom | 221; 2012 to NR; FUP mean 1.9 | Any CVD risk | Epicardial fat volume (computed tomography coronary angiogram) | CHD, Revasc, ACM | 0 |
| Djaileb 2021; Prosp. Cohort; France | 168; 2007 to 2010; FUP median 4.6 | No ASCVD with High CVD risk | Stress myocardial perfusion entropy (MPE) quantified from SPECT myocardial perfusion images | CHD, CVM, Revasc, Angina | 0 |

NR, not reported; Prosp. Cohort, prospective chort; Retrosp. Cohort, retrospective cohort; Retrosp. Case-control, retrospective case-control cohort; Est. ASCVD, established atherosclerotic cardiovascular disease; Est. ASCVD or High CVD risk, established atherosclerotic cardiovascular disease or high cardiovascular disease risk; No ASCVD with Mod. CVD risk, no atherosclerotic cardiovascular disease with moderate cardiovascular disease risk; DKD, diabetic kidney disease; 3p MACE, 3-point major adverse cardiovascular events; Revasc, revascularization; HF, heart failure; CHD, coronary heart disease; PAD, peripheral artery disease; CVM, cardiovascular mortality; ACM, all-cause mortality

#### Supplemental Table 6. Included 48 genetic markers studies.

| Study; Design; Country | No. of participant.; Enrollment year; Follow-up (years) | Study Population | Gene(s) locus(i) | Variant(s) | Outcome |
|---|---|---|---|---|---|
| **Keavney, 1995; Prosp. Cohort; United Kingdo** | 326; from 1989; NR | Any CVD risk | ACE | (I/D) polymorphism | CHD |
| **Hong Huang, 1998; Prosp. Cohort; Finland** | 83; 1985-1988; FUP 9.1 | Any CVD Risk | ACE | (I/D) polymorphism | CHD |
| **Heijmans, 2000; Prosp. Cohort; Netherlands** | 72; up to 1986; FUP 10 | Elderly | PON | Met-55/Leu; Gln-192/Arg | CVM |
| **Levy 2002; Retrosp. Case-control; United States** | 412; 1989 to 1992; FUP median 6 | Any CVD risk | Haptoglobin | Haptoglobin phenotype | CHD |
| **Bernard 2004; Prosp. Cohort; France** | 212; 1994 to 1997; FUP mean 5 | No ASCVD | APOB | XbA1 | 3p MACE, Angina |
| **Doney 2005a; Prosp. Cohort; United Kingdo** | 2015; 1995 to 2004; FUP mean 3.1 | Any CVD risk | GSTT1; GSTP1; GSTM1 | GSTT1 null; Ile105Val; 15-kb deletion | 3p MACE |
| **Bager 2005; Prosp. Cohort; Germany** | 225; 1999 to 2000; FUP mean 4.3 | Enr. High CVD risk | RANTES | In1.1T/C (rs2280789); G-403A (rs2107538) | CVM |
| **Lu Qi, 2005; Prosp. Cohort; United States** | 879; 1993 to 1999; NR | No CVD risk | APM1 | G276T | 3p MACE, Revasc |
| **Zhang, 2005; Prosp. Cohort; United States** | 220; 1986 to 2000; NR | No CVD risk | LIPC | 514 C to T | CHD |
| **Wang 2005; Prosp. Cohort; Hong Kong** | 1281; from 1995; FUP mean 3.4 | Any CVD risk | ACE | (I/D) polymorphism | 3p MACE, Revasc, PAD |
| **Doney 2005; Prosp. Cohort; United Kingdo** | 1810; 2009 to 2011; FUP mean 4.13 | Any CVD risk | PPARA (PPAR-alpha) | L162V variant; C2528G variant | CHD, ACM |

| Study; Design; Country | No. of participant.; Enrollment year; Follow-up (years) | Study Population | Gene(s) locus(i) | Variant(s) | Outcome |
|---|---|---|---|---|---|
| **Porchay-Baldarelli 2007; Prosp. Cohort; France** | 3124; 1987 to 1987; FUP mean 4 | Enr. High CVD risk | CETP | CETP TaqIB Polymorphism | CHD |
| **Odeberg 2008; Prosp. Cohort; Sweden** | 403; 1992 to 1993; NR | Any CVD risk | NOS3 (eNOS) | Asp 298 polymorphism | CHD |
| **Hadjadj, 2008; Prosp. Cohort; France** | 3126; NR; FUP 4 | Enr. high CV risk | ACE | (I/D) polymorphism | 3p MACE, ACM, Renal |
| **So 2008; Prosp. Cohort; Hong Kong** | 1074; 1995 to 1998; FUP median 8.4 | No ASCVD | ALR2 | C-106T; 5'-(CA) microsatellite | 3p MACE |
| **Doney 2009; Prosp. Cohort; United Kingdo** | 4897; 1997 to 2006; FUP mean 3.6 | Any CVD risk | FTO | rs9939609 | CHD |
| **Porchay-Baldérelli , 2009; Prosp. Cohort; Austria, Belg** | 3129; NR; FUP 4 | DKD | ABCA1 | R219K; R1587K; (+)69C> T | CHD |
| **Winkler 2010; Prosp. Cohort; Germany** | 1255; NR; FUP mean 3.71 | No CVD | ApoE | ApoE4 | 3p MACE |
| **Poon 2010; Prosp. Cohort; China** | 219; 1987 to 1987; FUP mean 5.3 | DKD | B1AR | Arg389Gly | 3p MACE, Revasc, Angina, PAD, Renal |
| **Wang 2010; Prosp. Cohort; Hong Kong** | 1297; 1994 to 1998; FUP median 8.3 | No ASCVD | SCYA11 (Eotaxin); PON2; ADRB3 | Ala23Thr; Ser311Cys; Trp64Arg; Ala23Thr, Ser311Cys, Trp64Arg | CHD |
| **Russo 2011; Prosp. Cohort; Italy** | 216; NR; FUP mean 5.4 | Any CVD risk | MTHFR | C677T polymorphism | CHD, Stroke, Revasc, PAD |
| **Qi 2011; Retrosp. Case-control; United States** | 2506; up to 2006; NR | Any CVD risk | GRS for CAD | 5 SNP-based GRS | CHD |
| **Lu 2011; Prosp. Cohort; Taiwan** | 309; 2006 to 2009; FUP median 2.35 | Enr. High CVD risk | DDAH1 gene | rs233112, rs1498373, rs1498374, rs587843, rs1403956, rs1241321 | 3p MACE |

| Study; Design; Country | No. of participant.; Enrollment year; Follow-up (years) | Study Population | Gene(s) locus(i) | Variant(s) | Outcome |
|---|---|---|---|---|---|
| **Hoffmann 2011; Prosp. Cohort; Germany** | 1232; 2011 to 2012; FUP median 4 | on dialysis | KIF6 gene | Trp719Arg genotypes (rs20455) | 3p MACE, ACM |
| **Bacci 2011; Prosp. Cohort; Italy** | 330; 2001 to 2008; FUP mean 3.1 | Est. ASCVD | ENPP1 | K121Q (rs1044498) | 3p MACE |
| **Ho 2012; Prosp. Cohort; Hong Kong** | 1417; 1995 to 2002; FUP mean 10.6 | No ASCVD | HNF4A; ADIPOQ; PPARG | rs1884614; rs1063539; Pro12Ala (rs1801282) | CHD |
| **Qi 2012; Prosp. Cohort; United States** | 2308; 1989 to 1999; FUP 12-16 | Any CVD risk | LPA genotype score; LPA | 5 SNP Genetic Score; rs783147 ; rs6935921 ; rs6919346; rs2048327; ; rs12214416; rs10455872 | 3p MACE |
| **Neves AL 2012; Prosp. Cohort; France** | 3744; NR; FUP 10 | DKD | SOD1 | rs9974610; rs2173962; rs2070424; rs202449; rs17880135; rs10432782; rs1041740/ rs17880196 | 3p MACE |
| **Kuricova¡ 2013; Prosp. Cohort; Czech Republi** | 311; 2002 to 2007; FUP median 3.2 | Any CVD risk | NOS3 (eNOS) | 894G> T (rs1799983) | 3p MACE, Revasc, PAD |
| **Qi 2013; Prosp. Cohort; United States** | 2310; up to 2006; NR | No ASCVD | GLUL | rs10911021 | CHD |
| **Ferrarezi, 2013; Prosp. Cohort; France** | 3137; NR; FUP mean 4.4 | CVD risk | VDR | rs7975232; rs731236; rs1544410 | CHD, Revasc |
| **Cox 2014; Prosp. Cohort; United States** | 699; 1998 to 2005; FUP mean 8.4 | Any CVD risk | GRS for CAD | 30 SNP-based GRS | CVM |
| **Katakami 2014; Prosp. Cohort; Japan** | 1977; 2005 to 2005; FUP median 7.5 | No ASCVD | pro-oxidant gene risk score; SOD2; NOS3 (eNOS); MPO; GCLM; CYBA | Val16Ala, C-588T, G894T, C242T, G-463A; Val16Ala; G894T; G-463A; C-588T; C242T | CHD |
| **PYK Poon · 2014; Prosp. Cohort; China** | 347; NR; FUP mean 3.3 | ESRD | HSPA1B | A+1267G | CHD, HF |
| **Alkhalaf 2015; Prosp. Cohort; Netherlands** | 871; 1998 to 1999; FUP median 9.5 | Any CVD risk | CNDP1 | 5-leucine repeat (5L5L) | CVM |

| Study;<br>Design;<br>Country | No. of participant.;<br>Enrollment year;<br>Follow-up (years) | Study Population | Gene(s) locus(i) | Variant(s) | Outcome |
|---|---|---|---|---|---|
| **Mccaffery 2015;<br>Prosp. Cohort;<br>United States** | 4016;<br>2001 to 2004;<br>FUP median 9.6 | Any CVD risk | GRS for CAD | 153 SNP-based GRS | 3p MACE, Angina |
| **Mohammedi 2015;<br>Prosp. Cohort;<br>Belgium, Fran** | 3137;<br>NR;<br>FUP median 4.7 | DKD | SOD3 | rs758946; rs2284659; rs17552548 | CHD, CVM |
| **Ortega Moreno, 2016;<br>Prosp. Cohort;<br>Italy** | 368;<br>2001 to 2008;<br>FUP mean 5.4 | Est. ASCVD | ADIPOQ | rs822354 | CVM |
| **Huggins 2016;<br>Prosp. Cohort;<br>United States** | 3845;<br>2001-2004;<br>FUP median 9.6 | No ASCVD | GLUL | rs10911021 | 3p MACE, Angina |
| **Baeney 2016;<br>Prosp. Cohort;<br>United Kingdo** | 6531;<br>NR;<br>FUP median 10 | No ASCVD | GLUL | rs10911021 | CHD |
| **Roumeliotis 2017;<br>Prosp. Cohort;<br>Greece** | 142;<br>2008 to 2015;<br>FUP mean 4.2 | DKD | MGP | MGP T-138C polymorphism | CVM, Renal |
| **Morieri 2018;<br>Prosp. Cohort;<br>Canada, Unite** | 7291;<br>2001 to 2005;<br>FUP mean 4.7 | Enr. High CVD risk | GRS for CAD | 204 SNP (160 loci)-based GRS | 3p MACE |
| **Roumeliotis, 2018;<br>Prosp. Cohort;<br>Greece** | 145;<br>2008 to 2008;<br>FUP mean 7 | CVD risk | ALOX12 | rs14309 | 3p MACE, PAD |
| **Valoti 2019;<br>Prosp. Cohort;<br>Italy** | 1158;<br>NR;<br>FUP median 5 | High CVD risk | CFH | p.Glu936Asp (c.2808G> T) | 3p MACE, Revasc,<br>HF, Angina |
| **Satirapoj 2019;<br>Prosp. Cohort;<br>Thailand** | 422;<br>2011 to 2015;<br>FUP median 5 | Any CVD risk | TCFL7; PPARG | rs7903146; Pro12Ala (rs1801282) | CHD |
| **Tan 2020;<br>Prosp. Cohort;<br>United Kingdo** | 13655;<br>2006 to 2010;<br>FUP mean 6.8 | Any CVD risk | MTNR1B | rs10830963 | CHD |
| **He 2021;<br>Prosp. Cohort;<br>China** | 2500;<br>2016 to 2018;<br>NR (<3) | no ASCVD | ALDH2 | rs671 | 3p MACE |

| Study; Design; Country | No. of participant.; Enrollment year; Follow-up (years) | Study Population | Gene(s) locus(i) | Variant(s) | Outcome |
|---|---|---|---|---|---|
| **Watson 2021; Prosp. Cohort; Ireland** | 498; 2005 to 2011; FUP 3.5 | Any CVD risk | MMP-9 | rs3918242 | CHD |

NR, not reported; Prosp. Cohort, prospective chort; Retrosp. Cohort, retrospective cohort; Prosp. from RCT, prospective cohort from randomized clinical trial; Est. ASCVD, established atherosclerotic cardiovascular disease; Enr. High CVD risk, enrolled in high cardiovascular disease risk; DKD, diabetic kidney disease; 3p MACE, 3-point major adverse cardiovascular events; Revasc, revascularization; HF, heart failure; CHD, coronary heart disease; PAD, peripheral artery disease; CVM, cardiovascular mortality; ACM, all-cause mortality.

#### Supplemental Table 7. Included 47 risk score studies.

| Study; Design; Country | No. of participant; Enrollment year; Follow-up (years) | Study Population | Exposure(s) | Outcome |
|---|---|---|---|---|
| **Stevens 2001; Prosp. Cohort; UK** | 4540; 1977 to 1997; FUP mean 10.4 yrs | No ASCVD | Risk score (Age at diabetes diagnosis), duration of diabetes, sex, ethnicity, smoking status, atrial fibrillation, systolic blood pressure, total cholesterol, HDL cholesterol, A1C | CHD |
| **Folsom 2003; Prosp. Cohort; US** | 1,500; 1987 to 1989; FUP median 10.2 | Without ASCVD | Risk score (age, race, total cholesterol, HDL cholesterol, systolic blood pressure, use of antihypertensive medication, smoking status, BMI, waist-to-hip ratio, lipoprotein (a), albumin, creatinine, fibrinogen, factor VIII, WBC, Keys score, sport activity index, LVH, and IMT) | CHD, CVM, Revasc |
| **Clarke 2004; Prosp. Cohort; UK** | 5102; 1977 to 1997; | No ASCVD | Risk score (age, sex, ethnicity, A1C, systolic blood pressure, total:HDL cholesterol, ischemic heart disease, CHF) | CHD |
| **Guzder 2005; Prosp. Cohort; United Kingdom** | 428;C21996 to 1998; FUP median 4.2 | NO CVD | Framingham equation; The United Kingdom Prospective Diabetes Study (UKPDS) risk engine | CHD, Stroke, HF, Angina, PAD |
| **Donnan 2006; Prosp. Cohort; United Kingdom** | 4569; 1995 to 2004; FUP median 4.1 | No ASCVD | Diabetes Audit and Research in Tayside, Scotland (DARTS), database risk score; The Salford Diabetes Information System (DIS) risk score | CHD, CVM |
| **Cederholm 2008; Prosp. Cohort; Sweden** | 11646; 1998 to 2003; FUP mean 5.64 | No CVD | Swedish National Diabetes Register (NDR) CVD risk score; UKPDS risk score | CHD |
| **Yang 2008; Prosp. Cohort; China** | 7067; 1995 to 2005; FUP median 5.4 | Without ASCVD | Risk score (age, sex, smoking, duration of diabetes, eGFR, spot urinary albumin-creatinine ratio, non-HDL cholesterol). | CHD |
| **vanderHeijden 2009; Prosp. Cohort; Netherlands** | 125; 1989 to 2000; FUP mean 10 | No ASCVD | Systematic Coronary Risk Evaluation (SCORE); Framingham risk score (FRS); UK Prospective Diabetes Study (UKPDS) risk function | CHD |
| **Davis 2009; Prosp. Cohort; Multinational** | 815; 1993 to 1996; FUP mean 5 | Est. ASCVD or High CVD risk | FRS; UKPDS CV risk score | CHD |

| | | | | |
|---|---|---|---|---|
| **Simmons 2009; Prosp. Cohort; United States** | 10137; 1993-1998; FUP median 10.1 | without CVD | UKPDS Risk Engine (version 3) and the Framingham risk equations (2008) | 3p MACE, PAD |
| **Elley 2010; Prosp. Cohort; New Zealand** | 36127; 2000 to 2006; FUP median 3.9 | No CVD | Diabetes Cohort Study (DCS) Risk Score; Framingham risk Score; UKPDS risk Score | CHD, Stroke, PAD |
| **Davis 2010; Prosp. Cohort; Australia** | 1240; 1993 to 1996; | No CVD | Busselton Health Study (BHS) risk score; Fremantle Diabetes Study(FDS) risk score | 3p MACE |
| **Kengne 2010; Prosp. Cohort; Multinational** | 11,140; 2001 to 2007; FUP mean 5 | Without ASCVD | Validation of FRS and UKPDS scores in ADVANCE cohort | CHD, Stroke, Revasc, Angina |
| **Rossi 2011; Prosp. Cohort; Italy** | 5181; 2006 to 2007; FUP median 2.3 | Est. ASCVD or High CVD risk | Quality of care scoring: Includes A1c, BP, no of lipid measurements/year, LDL, macroalb measurements/year, tx with ACEI/ARB if MA present | 3p MACE, Revasc, Angina |
| **Kengne 2011; Prosp. Cohort; Multinational** | 7168; 1999 to 2001; FUP mean 4.5 | No ASCVD | ADVANCE risk equation | 3p MACE |
| **Zethelius 2011; Prosp. Cohort; Sweden** | 24288; 2002 to 2007; FUP mean 5 | With or without ASCVD | Risk score (age at diabetes onset, diabetes duration, TC:HDL, A1C, SBP, BMI, sex, smoker, microalbuminuria, macroalbuminuria, atrial fibrillation, previous CVD) | CHD |
| **van Dieren 2011; Prosp. Cohort; Netherlands, Germany** | 1662; 1993 to 1998; FUP mean 8 | Without ASCVD | Validation of UKDPS risk score in EPIC-NL and EPIC-Potsdam cohorts | CHD, Stroke |
| **Yoshida 2012; Retrosp. Cohort; Japan** | 783; 2003 to 2005; FUP mean 5.46 | No ASCVD | Carotid IMT + Framingham Risk Score (FRS) | 3p MACE, Angina |
| **Yang 2013; Prosp. Cohort; Multinational** | 39585; 2004 to 2006; | with or without CVD | modified UKPDS risk engine | 3p MACE |

| | | | | |
|---|---|---|---|---|
| **Wells 2013; Prosp. Cohort; United States** | 23906; 1998 to 2006; FUP median 1.2 | No ASCVD | A risk score for CHD | CHD |
| **Tanaka 2013; Prosp. Cohort; Japan** | 1748; 1996 to 2000; FUP median 7.2 | No CVD | The Japan Diabetes Complications Study/the Japanese Elderly Diabetes Intervention Trial risk engine | CVM, Stroke, Renal |
| **Hayes 2013; Prosp. Cohort; UK** | 5102; 1977 to 2007; FUP median 17.6 yrs | No ASCVD | Risk score (age, duration of diabetes, sex, ethnicity, smoker, systolic blood pressure, A1C, LDL, HDL, BMI, eGFR, heart rate, atrial fibrillation, PVD, albuminuria, hemoglobin, white blood cells, MI, stroke, IHD, CHF, blindness, amputation, renal failure, diabetic ulcer) | CHD |
| **Mukamal 2013; Prosp. Cohort; US** | 782; 1989 to 1993; FUP mean 10 | Without ASCVD | Risk score (age, smoking, systolic blood pressure, total cholesterol, HDL cholesterol, creatinine, use of glucose-lowering medications, ankle-brachial index, ECG signs of left ventricular hypertrophy, carotid IMT) | 3p MACE |
| **Cox 2014a; Prosp. Cohort; United Kingdom** | 4704; 2011 to 2012; FUP median 2 | Any CVD risk | risk score (age .=70), male, A1c +/7.4, BMI, SBP, DBP, total cholesterol, HDL, LDL) | CHD, Stroke, Angina, PAD |
| **Yeboah 2014; Prosp. Cohort; Multinational** | 1343; 2000 to 2003; FUP mean 8.5 | No ASCVD | MESA and Heinz risk score (age, sex, SBP, DM duration, CAC) | CHD |
| **Pinias 2014; Prosp. Cohort; Spain** | 777; 2000 to 2000; FUP mean 8 | No ASCVD | Basque Country Prospective Complications and Mortality Study risk engine (BASCORE) (Age, the ratio of non-HDL- to HDL-cholesterol, HbA1c, systolic blood pressure and smoking); The Framingham Risk Score-Regicor Study (FRS-RS); UK Prospective Diabetes Study risk engine (UKPDS-RE) | CHD, Stroke, Angina, PAD |
| **Cox 2014; Prosp. Cohort; United States** | 699; 1998 to 2005; FUP mean 8.4 | Any CVD risk | Multi-bed vascular calcification score (coronary, carotid, abdominal vascular beds) | 3p MACE |
| **Ramarez-Prado 2015; Prosp. Cohort; Spain** | 112; 2010 to 2012; FUP mean 2.3 | Patients at emergency department, no CVD | A novel four-year cardiovascular risk score | 3p MACE, HF, Renal |
| **McEwan 2015; Retrosp. Cohort; United Kingdom** | 123159; 2000 to 2009; FUP mean 5.82 | low- and intermediate-risk patients. | Refitted UKPDS 68 event equations | CHD |

| | | | | |
|---|---|---|---|---|
| **Lagani 2015; Retrosp. Cohort; United States** | 344; 2004 to 2014; FUP mean 3.26 | No ASCVD | CVD-DCCT (Cox Regression); CVD-EDIC (Accelerated Failure Model) | 3p MACE, Revasc, HF, Angina |
| **van der Leeuw 2015; Prosp. Cohort; Multinational** | 453, 1174 and 584; NR ; NR | Without ASCVD | Validation of 10 risk models: CHS, ADVANCE, Fremantle, DCS, NDR, DCS, HKDR, DARTS, ARIC, UKPDS | CHD |
| **Woodward 2016; Prosp. Cohort; United Kingdom** | 7301; 2005 to 2006; FUP mean 9.9 | High risk for CVD | the AD-ON global vascular risk score | 3p MACE |
| **Basu 2017; Prosp. Cohort; United States** | 15413; 1992 to 2008; | Any CVD risk | Action to Control Cardiovascular Risk in Diabetes study(ACCORD) for Risk Equations for ComplicationsComplications Of type 2 Diabetes (RECODe); Diabetes Prevention Program Outcomes Study (DPPOS) CV risk socre | CHD, Stroke |
| **Wan 2018; Retrosp. Cohort; China** | 137935; 2010 to 2010; FUP median 5 | No ASCVD | Risk engine (age, amoker, DM duration, anti-HT, isulin use, BMI, A1c, SBP, DBP, TC/HDL, LnACR, eGFR, Age interaction | 3p MACE, HF |
| **Young 2018; Retrosp. Cohort; Canada** | 181619; 2006 to 2016; FUP median 1 | Any CVD risk | risk score (age, female, baseline period, insurance type, payer type, other CVD related conditions, DM hospitalization, coagulopathy, time interval) | 3p MACE |
| **Shao 2018; Prosp. Cohort; United Kingdom** | 10251; from 2001; FUP median 3.7 | Any CVD risk | BRAVO risk engine (A1cl, LDL, age at diagnosis, Severe hypo, white race, MI history, stroke history) | CHD |
| **Mentz 2018; Prosp. Cohort; Multinational** | 14752; 2010 to 2015; FUP median 3.2 | WIth and wthout CVD risk | A novel risk score | 3p MACE |
| **Li 2018; Retrosp. Cohort; China, Taiwan** | 4275; 1999 to 2003; FUP mean 8.2 | Any CVD risk | quality-of-care score | CHD, Stroke, HF, PAD |
| **Hamada 2018; Retrosp. Cohort; United Kingdom** | 35196; 2006 to 2010; FUP mean 6 | Any CVD risk | Multiple risk factor control (A1C, blood pressure, total cholesterol, smoking) | 3p MACE, ACM |

| | | | | |
|---|---|---|---|---|
| **Basu 2018; Prosp. Cohort; United States** | 3301; 1992 to 2008; FUP median 8.5 | Any CVD risk | the Jackson Heart Study (JHS) RECODe; the Multi-Ethnic Study of Atherosclerosis (MESA) | CHD, Stroke |
| **Read 2018; Prosp. C; Scotland** | 181,399; 2004 to 2016; FUP median 5 | Without ASCVD | Validation of 6 risk scores: QRISK2, ADVANCE, Cardiovascular Health Study (CHS), New Zealand Diabetes Cohort Study (NZ DCS), Fremantle Diabetes Study, and Swedish National Diabetes Register (NDR) | CHD, Stroke, Angina, PAD |
| **Yu 2019; Prosp. Cohort; China** | 2282; 2015 to 2016; FUP mean 2 | Any CVD risk | CVD in nephropathy score (gender, age, BMI, SBP, DBP, FG, HDL, TG, INR, TSH, Mg, bili, ALT, urine protein, MCV, Hb, HCT, LDL | CVM |
| **Quan 2019; Prosp. Cohort; China** | 678750; 2004 to 2014; FUP mean 6.3 | No ASCVD | HKU-SG risk score; JADE risk score; RECODe risk score; UKPDS risk score | CHD |
| **Zhang 2020; Prosp. Cohort; China** | 3232; 2008 to 2009; FUP mean 10 | Any CVD risk | improved Framingham Risk Score (IFRS) | CHD, Stroke, Angina |
| **Davis 2020; Prosp. Cohort; Australia** | 1551; 2008 to 2011; FUP mean 5 | Any CVD risk | FDS, BHS three-point and four-point conventional major adverse cardiovascular events (MACE) risk score | 3p MACE, HF |
| **Venalainen 2020; Prosp. Cohort; Multinational** | 4733; 2005 to 2010; FUP mean 4.7 | Est. ASCVD or High CVD risk | Risk score (age, CVD history, current smoking, creat, urine ACR, no of anti-HT agens, assignment to intensive tx) | 3p MACE, HF |
| **Shao 2020; Prosp. Cohort; United States** | 34322; 2008 to 2012; | Any CVD risk | the Building, Relating, Assessing, and Validating Outcomes (BRAVO) risk engine | 3p MACE |

NR, not reported; Prosp. Chort, prospective cohort; Retrosp. Cohort, retrospective cohort; Est. ASCVD, established atherosclerotic cardiovascular disease; 3p MACE, 3-point major adverse cardiovascular events; Revasc, revascularization; HF, heart failure; CHD, coronary heart disease; PAD, peripheral artery disease; CVM, cardiovascular mortality; ACM, all-cause mortality.

**Supplemental Table 8. Summary of the results on the associations between biomarkers and CV outcomes (248 studies on overall 195 different biomarkers).**

| Biomarker (N=195) | No of Studies Per biomarker | No. of studies with positive association | Author (Year) | Outcome | Results (Green = Statistically significant association in any direction; red= no association) |
|---|---|---|---|---|---|
| NT-proBNP | 14 | 13 | Colombo 2018 | CHD, Stroke | HR 1.94 (1.54-2.45) |
| | | | Bruno 2014 | CVM | HR 1.84 (1.52-2.22) (log-transformed) |
| | | | Bruno 2013 | CVM | HR 1.690 (1.39-2.05) (log-transformed) |
| | | | McMurray 2011 | 3p MACE, HF | HR 1.30 (1.15-1.46) (log-transformed) |
| | | | Resl 2016 | CHD, Stroke, HF, PAD, ACM | HR 1.17 (1.07-1.27) |
| | | | Savonitto 2018 | CVM | HR 1.83 (1.61-2.1) (log-transformed) |
| | | | Sharma 2020 | 3p MACE, HF | HR 1.24 (1.18-1.31) (log-transformed);<br>Adding NT-proBNP to clinical variable, c-statistic changed from 0.66 to 0.71;<br>NRI 0.3854 (0.3037-0.4721);<br>IDI 0.0877 (0.077-0.0976) |
| | | | Wolsk 2017 | 3p MACE, HF, ACM | CVM: c-statistic 0.83 (0.80-0.86);<br>NRI 0.309 (0.217-0.369);<br>IDI 0.04 (0.024-0.0630);<br><br>MI: c-statistic 0.72 (0.67-0.74);<br>NRI 0.106 (0.0570-0.1660);<br>IDI 0.0080 (0.0030-0.0160) |
| | | | Wong 2019 | 3p MACE, PAD | C-statistic 0.67 (0.64-0.69);<br>difference in AUCs over risk factor model 0.03 (p = 0.001);<br>NRI 0.345 (0.238-0.451)<br>IDI 0.012 (0.007-0.017) |

| | | | | | |
|---|---|---|---|---|---|
| | | | vonScholten 2015 | 3p MACE, HF | HR 1.70 (1.10-2.50) |
| | | | Scirica 2016 | 3p MACE, HF | C-statistic 0.72 (0.71-0.74, *p* <0.001);<br>IDI 0.0379 (0.0321-0.0438);<br>NRI 0.480 (0.410-0.550) |
| | | | Lorenzo-Almorós 2020 | CHD, Stroke | HR 0.95 (0.68-1.33) per 1 SD increase |
| | | | Rørth 2019 | CHD, CVM, HF, ACM | NT-proBNP in highest tertile HR 3.35 (2.19-5.11), or 4.5-fold higher risk when combined with elevated TnT ≥18 ng/ml |
| | | | Van der Leeuw 2016 | 3p MACE, HF, PAD | SMART cohort: change in c-statistic 0.02 (0.00-0.04) on top of base model of 0.70 (0.67-0.74);<br><br>EPIC-NL cohort: change in c-statistic 0.02 (0.00-0.05) on top of base model of 0.69 (0.64-0.74);<br><br>SMART NRI 0.27 (0.10-0.44);<br><br>EPIC NRI 0.50 (0.26-0.73) |
| BNP | 1 | 1 | Lepojarvi 2016 | CVM, HF | HR 5.90 (2.8-12.40) for BNP ≥116 ng/L;<br>Adding BNP to clinical variable, c-index changed from 0.794 to 0.844; NRI 0.829 (0.512-1.146);<br>IDI 0.059 (0.024-0.094, *p* <0.001) |
| NT-proBNP + MMP-3 + osteopontin | 1 | 1 | Van der Leeuw 2016 | 3p MACE, HF, PAD | SMART cohort: change in c-statistic 0.03 (0.01 to 0.05) on top of base model of 0.70 (0.67-0.74);<br><br>EPIC-NL cohort: change in c-statistic 0.03 (0.00 to 0.03) on top of base model of 0.69 (0.64-0.74);<br><br>SMART NRI 0.12 (0.03-0.21);<br><br>EPIC NRI 0.07 (0.04-0.17) |

| | | | | | |
|---|---|---|---|---|---|
| Troponin T (TnT) | 8 | 7 | McMurray 2011 | 3p MACE, HF | HR 1.3 (1.15-1.46) alone or HR 1.5 (1.06-2.13) for TnT >0.028 ng/ml |
| | | | Lepojarvi 2016 | CVM, HF | HR 15.5 (5.1-46.7) for hs-TnT >14 ng/ml;<br>C-index 0.895 (vs 0.794 in basal models);<br>NRI 0.231 (0.067-0.394);<br>IDI 0.054 (0.031-0.077) |
| | | | Keller 2018 | 3p MACE | HR 1.35 (1.01-1.82) for hsTnT >35-55 ng/L;<br>HR 1.73 (1.24-2.42) for >55-90 ng/ml,<br>HR 2.1 (1.49-2.95) for hsTnT >90 ng/ml |
| | | | Everett 2015 | 3p MACE, ACM | HR 1.85 (1.48-2.32) for TnT ≥14 ng/ml |
| | | | Scirica 2016 | 3p MACE, HF | HR 1.41 (1.31-1.51) for 1 SD;<br>C-statistic 0.72 (0.70-0.73) change in c-statistic -0.007 (-0.08 to -0.05); IDI 0.0285 (0.0233-0.0336);<br>NRI 0.44 (0.38-0.51); |
| | | | Resl 2016 | CHD, Stroke, HF, PAD, ACM | HR 1.43 (1.13-1.82, p<0.01) or HR 1.15 (1.0-1.32) with GDF-15 |
| | | | Lorenzo-Almoros 2020 | CHD, Stroke | HR 1.02 (0.84–1.23) |
| | | | Rørth 2019 | CHD, CVM, HF, ACM | Combined elevated TnT ≥18 ng/ml and NT-proBNP in highest quartileile: 4.5-fold higher risk of primary outcome. |
| Troponin I | 3 | 3 | Yiu 2014 | 3p MACE, HF | HR 2.85 (1.15-7.03) for MACE;<br><br>HR 4.88 (1.12-21.31) for HF |
| | | | Sharma 2020 | 3p MACE, HF | HR 1.04 (1-1.09);<br>adding troponin I to clinical variable, c-statistic changed from 0.66 to 0.68;<br>NRI 0.2914 (0.2054, 0.3705);<br>IDI 0.0340 (0.0283, 0.0406) |
| | | | Wong 2019 | 3p MACE, PAD | HR 1.75 (1.41-2.17) for TnI >14.2 pg/ml;<br>NRI 0.349 (0.248-0.45);<br>IDI 0.024 (0.016-0.032) |

| | | | | | |
|---|---|---|---|---|---|
| CRP | 10 | 7 | Schulze 2004 | CHD, Stroke, Revasc | RR 2.62 (1.29-5.32) for Q4 vs, Q1;<br>HR 2.52 (1.31-4.86) for Q3 vs. Q1;<br>HR 1.51 (0.76-2.98) |
| | | | McMurray 2011 | 3p MACE, HF | HR 1.44 (1.23-1.69) for CRP ≥6.6 mg/L;<br>HR 1.2 (1.02-1.41) for CRP >3.0 to <6.6 mg/L |
| | | | Lepojarvi 2016 | CVM, HF | HR 4.6 (2.1-9.9) for hsCRP >1.6 mg/L;<br>C-index 0.842 (vs 0.794);<br>NRI not significant;<br>IDI 0.033 (0.009-0.057, *p* <0.01) |
| | | | Lowe 2014 | 3p MACE, Renal | HR 0.95 (0.85-1.05) for log CRP - 1 SD;<br>C-statistic 0.683 (0.662-0.705);<br>IDI 0.0004 (-0.0017-0.0026);<br>NRI 0.008 (-0.124-0.131) |
| | | | Cardoso 2016 | 3p MACE, ACM | 3p MACE: HR 1.3 (1.09-1.55) for log CRP - 1 SD;<br>HR 1.72 (1.21-2.44) for CRP ≥3.0 mg/L;<br>HR 1.84 (1.19-2.83) for CRP ≥4.8 mg/L;<br><br>CVM: HR 1.13 (0.84-1.52) for log CRP - 1 SD;<br>HR 1.38 (0.77-2.46) for CRP ≥3.0 mg/L;<br>HR 1.24 (0.62-2.48) for CRP ≥4.8 mg/L |
| | | | Scirica 2016 | 3p MACE, HF | HR 1.37 (1.2-1.56) for hscRP >3 mg/L;<br>HR 1.1 (1.03-1.17) for log CRP -1 SD;<br>NRI and IDI <0.20 |
| | | | vonScholten 2016 | 3p MACE, HF | HR 1 (0.7-1.4) for logCRP - 1 SD |
| | | | Lorenzo-Almoros 2020 | CHD, Stroke | HR 0.97 (0.93-1.03) per 1 SD increase |
| | | | Linnemann 2006 | 3p MACE, PAD, ACM | OR 5.4 (2.13-13.76) |
| | | | Van der Leeuw 2016 | 3p MACE, HF, PAD | SMART cohort: change in c-statistic 0.00 (-0.01 to 0.01) on top of base model of 0.70 (0.67-0.74);<br>EPIC-NL cohort: change in c-statistic 0.02 (0.00 to 0.04) on top of base model of 0.69 (0.64-0.74) |

| | | | | | |
|---|---|---|---|---|---|
| Hemoglobin glycation index | 2 | 2 | JIn 2018 | 3p MACE, Revasc, Angina | HR 1.215 (1.046-1.411);<br>C-stat not improved |
| | | | Kim 2018 | CHD, Stroke | HR 0.98 (0.63-1.51) for 2nd vs. 1st quartile;<br>HR 0.90 (0.55-1.46) for 3rd vs. 1st quartile;<br>HR 1.74 (1.08-2.81) for 4th vs. 1st quartile |
| A1C variability | 6 | 4 | Ceriello 2020 | 3p MACE | HR 1.97 (1.36-2.84) |
| | | | Rozing 2019 | 3p MACE, PAD, ACM | HR 0.82 (0.55-1.22) |
| | | | Prentice 2016 | CHD, Stroke, HF, Angina, PAD, ACM | HR 1.25 (1.1-1.41) for 3rd vs. 1st quartile;<br>HR 1.23 (1.07-1.42) for 4th vs. 1st quartile |
| | | | Cardoso 2018 | 3p MACE, ACM | HR 1.11 (0.91-1.34) per 1 SD at 12 months;<br>HR 1.2 (1.0-1.44) per 1 SD at 24 months |
| | | | Bouchi 2012 | CHD, Stroke, Revasc, Angina | HR 1.7 (0.96-3.0) per 1 SD;<br>HR 1.48 (0.59-3.71) for 2nd vs. 1st quartile;<br>HR 2.32 (0.97-5.58) for 3rd vs. 1st quartile;<br>HR 3.38 (1.07-10.63) for 4th vs. 1st quartile |
| | | | Astergaard 2019 | 3p MACE, HF | HR 1.3 (1.07-1.57) |
| Fasting glucose | 4 | 4 | Wei 1998 | CVM | HR 4.9 (2.3-10.3) for 4th vs. 1st/2nd quartile;<br>HR 2.8 (1.3-5.9 for 3rd vs. 1st/2nd quartile |
| | | | Afsharian 2016 | CHD, Stroke, HF, Angina | HR 1.4 (0.94-2.04) for FG 7.22-<10 mmol/L vs. <7.22 mmol/L;<br>HR 2.31 (1.55-3.2) for FG ≥10 mmol/L vs. <7.22 mmol/L |
| | | | Abu-Lebdeh 2001 | CHD | HR 1.63 (1.17-2.25) for log FG |
| | | | Niskanen 1998 | CVM | Baseline variables: OR 1.13 (0.93-1.10; p=0.04) (per mmol/L) *note discordance between CI and *p*-value; 5-year fasting glucose: OR 1.16 (1.13-16.4), *p* = 0.02 |

| | | | | | |
|---|---|---|---|---|---|
| Post-prandial or random glucose | 6 | 5 | Gasior 2008 | CHD, ACM | HR 1.03 (0.98- 1.08) |
| | | | Takao 2017 | 3p MACE, Revasc | HR 1.13 (1.04-1.23) |
| | | | Cavalot 2006 | 3p MACE, Revasc, Angina, PAD | Men: HR 1.33 (0.65-2.73) for BG after breakfast;<br>HR 2.12 (1.04-4.32) BG for after lunch (tertiles);<br><br>Women: HR 0.72 (0.22-2.36) for BG after breakfast;<br>HR 5.54 (1.45-21.2) for BG after lunch |
| | | | Cavalot 2011 | 3p MACE, Revasc, Angina, PAD | Men: HR 1.33 (0.65-2.73) for post-breakfast BG;<br>HR 2.12 (1.04-4.32) for post-lunch BG (tertiles);<br><br>Women: HR 0.72 (0.22-2.36) for post-breakfast BG;<br>HR 5.54 (1.45-21.2) for post-lunch BG (tertile).<br><br>C-statistic improved from 0.656 to 0.682, *p* <0.0001) |
| | | | Faghihi-Kashani 2016 | CHD, CVM, Angina | HR 1.09 (0.96-1.23) (tertiles) |
| | | | Strojek 2016 | 3p MACE, Revasc | mean time to 75% event free survival for average change in mean 2 hr BG levels after meals across visits less than or equal to -0.14 mmol/L was 43.48 weeks vs. average change more than -0.14 was 29.10 weeks. |
| Triglycerides | 7 | 6 | Sone 2011 | CHD | HR 1.54 (1.22-1.94) (log-transformed) |
| | | | Sone 2012 | CHD, Angina | HR 1.72 (1.21-2.43) (log-transformed) |
| | | | Fuller 2001 | 3p MACE | SRR: 1.2 (1.1-2.43) for TG >1.26 mmol/L |
| | | | Giorda 2008 | CHD, Stroke, Revasc, Angina, PAD | HR 1.93 (1.1-3.4) for TG ≥ 1.69 mmol/L |
| | | | Chen 2020 | 3p MACE | HR 1.08 (0.99-1.11) |
| | | | Abu-Ledeh 2001 | CHD | HR 1.49 (1.15-1.92) (log-transformed) |
| | | | Pintó 2007 | CHD, Stroke, Angina, PAD | HR 1.648 (1.029-2.641) for TG ≥200 mg/dl |

| | | | | | |
|---|---|---|---|---|---|
| CCTA | 6 | 5 | Park 2014 | CHD, CVM, Revasc | OR 14.62 (3.24-66.07) *unadjusted |
| | | | Lee 2017 | CHD, Revasc, ACM | HR 1.21 (1.09-1.34) for coronary artery calcium score;<br><br>HR 3.83 (2.45-5.98);<br>C-statistic 0.788 (0.747-0.829);<br>IDI 0.046 (0.02-0.072);<br>NRI 0.55 (0.343-0.757) for CCTA detected obstructive CAD |
| | | | Halon 2019 | CHD | HR 1.03 (1.01-1.04) for CCTA plaque volume;<br><br>HR 1.4 (1.1-1.7) for % low density content;<br><br>HR 2.6 (1.1-5.8) for mild plaque calcification;<br><br>HR 8.7 (2.6-29.3) for all 3 high-risk attributes + stenosis |
| | | | Halon 2016 | CHD, CVM, Angina | C-statistic 0.789 (0.728-0.85) for CCTA plaque burden + clinical risk score + CACS;<br><br>C-statistic 0.824 (0.768-0.881) for CCTA plaque burden + clinical risk score + angiographic score;<br><br>Categorical NRI 0.24;<br><br>Continuous NRI 0.632;<br><br>IDI 0.647 |
| | | | Tian 2019 | CHD, CVM, Revasc, Angina | HR 11.132 (1.857-66.742) for non-obstructive CAD;<br><br>HR 7.792 (1.75-34.698) for obstructive CAD vs no CAD detected on CCTA |
| QT interval | 7 | 6 | Rana 2005 | CVM | HR 1.0151 (1.0072-1.0231);<br>c-statistic 0.71 (for QTc >475 ms) |

| | | | | | |
|---|---|---|---|---|---|
| | | | Linnemann 2003 | CVM | OR 2.7 (1.21-6.01) |
| | | | Cardoso 2003 | 3p MACE, Revasc, HF, PAD | HR 1.29 (1.1-1.52) per 10 ms;<br><br>C-statistic 0.59 (0.51-0.66) for QTc >470 ms,<br><br>C-statistic 0.67 (0.57-0.77) for QTd >60 ms |
| | | | Bianco 2014 | CVM | HR 5.1 (1.7-15.2) for QT >450 for men or >470 ms for women |
| | | | Christensen 2000 | CVM | RR 1.08 (1.01-1.15) per 10 ms |
| | | | Pfister 2011 | CHD, Stroke, ACM | HR 1.06 (1.03-1.09) |
| | | | Massardo 2020 | CVM | OR 7.48 (0.99-56.44) for QTc control (baseline myocardial SPECT);<br><br>OR 5.14 (0.69-38.32) for QTc control (control cholesterol);<br><br>C-statistic 0.79;<br><br>HR 37.58 (0.63-2227.83) |
| ECG abnormalities | 7 | 7 | Pfister 2011 | CHD, Stroke, ACM | HR 1.13 (1.07-1.19) for heart rate (per 10/min);<br><br>HR for RBBB and LBBB non-significant |
| | | | McMurray 2011 | 3p MACE, HF | HR 1.42 (1.21-1.66) for abnormal ECG |

| | | | | | |
|---|---|---|---|---|---|
| | | | Jimenez-Corona 2006 | CVM, ACM | HR 1.22 (0.76-1.97) for CV Death with minor ECG abnl;<br><br>HR 1.32 (0.7-2.5) for ischemic heart disease with minor ECG abnl;<br><br>HR 1.83 (1.21-2.76) for CV Death with major ECG abnormalities;<br><br>HR 2.12 (1.26-3.57) |
| | | | Fuller 2001 | 3p MACE | SRR 3.0 (2.0-4.50) for possible CHD vs. none (men);<br><br>SRR 3.5 (2.1-6.1) for probable CHD vs. none (men);<br><br>SRR 1.5 (1-2.3) for possible CHD vs. none (women);<br><br>SRR 2.2 (1.2-4.1) for probable CHD vs. none (women) |
| | | | deSantiago 2007 | CHD, Stroke, HF, Angina, PAD, ACM | RR 5.95 (2.29-15.47) for abnormal ECG |
| | | | Hadaegh 2012 | CHD, Stroke, Angina | Men: HR 2.32 (1.29-4.16) for abnl ECG w/o CHD symptoms;<br>HR 3.97 (2.27-7.02) for abnl ECG w/ CHD symptoms;<br><br>Women: HR 1.19 (0.65-2.18) for abnl ECG w/out CHD symptoms;<br>HR 1.92 (1.02-3.62) for abnl ECG w/ CHD symptoms |
| | | | Burgess 2010 | CHD | HR 4.55 (2.90-7.13) for those with silent MI, which was similar to risk for those with clinical MI;<br><br>HR 4.51 (3.08-6.61) for outcome of future CVD event. |
| | 6 | 6 | Lau 2012 | 3p MACE, Revasc, PAD | HR 27.11 (3.6-218.81) for CACS >40 |

| | | | | | |
|---|---|---|---|---|---|
| Coronary artery calcium score (CACS) | | | Elkeles 2008 | 3p MACE, Angina | HR 1.292 (1.156-1.443) (log-transformed);<br>HR 4.001 (0.867-18.465) for CACS 11-100,<br>HR 7.09 (1.604-31.33) for CACS 101-400,<br>HR 8.391 (1.843-38.209);<br>HR 13.793 (3.067-62.041) vs. CAS 0-10 (ref);<br>ROC AUC from 0.63 to 0.73, $p$ = 0.03 for CVD events vs UKPDS; ROC AUC from 0.63 to 0.73, $p$ = 0.01 for CHD vs Framingham |
| | | | vonScholten 2015 | 3p MACE, HF | HR 3.4 (1.7-6.7) (log-transformed) |
| | | | Venuraju 2019 | CHD, Revasc, ACM | HR 2.32 (1.12-4.81) |
| | | | Anand 2006 | 3p MACE, Revasc | c-statistic 0.92 (0.87-0.96) for CACS vs. 0.74 (0.65-0.83) for UKPDS $p$ <0.0001)<br>c-statistic 0.60 (0.48-0.73) for Framingham Risk Score (P=0.13);<br>RR 10.13 (1.68-61.12 for CAC 101-400 AU;<br>RR 40.65 (8.14-203.05) for CAC 401-1000 AU;<br>RR 58.05 (12.28-274.48) for >1000 AU |
| | | | Dayan 2012 | CHD, Stroke, Revasc, HF, PAD | HR 6.82 (3.42-13.58) for LnCAC score |
| SPECT scintigraphy | 6 | 5 | Petretta 2013 | CHD, CVM, Revasc | HR 2.2 (1.2-4.0) for SPECT summed difference score >6;<br>HR 3.6 (2.6-6.8);<br>C-stat 0.74-0.82, $p$ = 0.01;<br>NRI 0.39 |
| | | | LeFeuvre 2005 | CHD, CVM, Revasc | No results |
| | | | Djaileb 2021 | CHD, CVM, Revasc, Angina | HR 2.06 (1.03-4.11);<br>c-stat 0.66 vs 0.74;<br>NRI 0.63;<br>IDI 0.06 |
| | | | DeLorenzo 2002 | CHD, CVM | OR 5.04 (1.7-17.7) for abnormal SPECT;<br>OR 18.8 (5.4-63.3) for SPECT defect extent |
| | | | Cosson 2011 | 3p MACE, Revasc, HF, PAD | HR 1.76 (1-3.12);<br>C-statistic 0.705 (0.616-0.794) vs 0.788 (0.72-0.855) |
| | | | Vanzetto 1999 | CHD, CVM | OR 4.24 (1.36-13.9) *unadjusted |
| Carotid intima-media thickness (cIMT) | 5 | 5 | Roumeliotis 2019 | CHD, Stroke, PAD | HR 2.04 (1.1-3.78) for cIMT >0.86 mm |
| | | | Jeevarethinam 2018 | CHD, Stroke, Revasc, ACM | HR 5.1 (1.37-19);<br>OR 2.57 (1.34-4.92) for carotid plaque |

| | | | | | |
|---|---|---|---|---|---|
| | | | Cardoso 2019 | 3p MACE, Revasc, HF, PAD | HR 1.15 (1.02-1.31) (per 0.1 mm);<br>HR 1.83 (1.03-3.23) highest vs. lowest tertile;<br>HR 1.51 (0.97-2.33) for plaque score ≥3;<br>IDI 0.078 |
| | | | Yamasaki 2000 | CHD, Angina | OR 4.9 (1.7-14.1) |
| | | | Bernard 2005 | 3p MACE, Angina | OR=1.63 (1.01–2.63);<br>C-statistics: 0.715 |
| Ankle-brachial index (ABI) | 5 | 4 | Lin 2019 | CHD, Stroke, Revasc, PAD, ACM | HR 2.39 (1.26-4.53) for ABI <0.9, HR 0.93 (0.39-2.19) for 2nd vs. 1st quartile;<br>HR 1.02 (0.44-2.37) for 3rd vs. 1st quartile;<br>HR 3.27 (1.64, 6.49) for 4th vs. 1st quartile |
| | | | Filippella 2007 | CHD, Angina | OR 3.7 (2.2-6.2) for ABI <0.9 |
| | | | Chang 2016 | CHD, Stroke, Revasc, PAD, ACM | OR 5.35 (2.88-9.79) for ABI ≥0.9 *not adjusted |
| | | | Aboyans 2011 | CHD, Stroke, ACM | HR 2.21 (1.16-4.22) for ABI >1.40 |
| | | | Zobel 2017 | 3p MACE, HF | HR 1.44 (1.23-1.59) |
| Fibrinogen | 5 | 4 | Saito 2000 | 3p MACE, Revasc, HF | RR 1.71 (1.12-2.73) for highest vs. lowest quartile |
| | | | Koch 1997 | CHD, CVM | HR 1.0037 (1.0014-1.006) per 1 mg/dl |
| | | | Lowe 2014 | 3p MACE, Renal | C-statistics: 0.684 (0.663-0.706), $p$ = 0.24;<br>IDI: 0.000 (20.002-0.002), $p$ = 0.98;<br>NRI (cat):0.057 (20.070-0.184), $p$ = 0.35 |
| | | | Bruno 2005 | CVM | RR 1.37 (0.97-1.94) for 2nd vs. 1st quartile;<br>RR 1.58 (1.13-2.21) for 3rd vs. 1st quartile;<br>RR 1.77 (1.28-2.46) for 4th vs. 1st quartile |
| | | | Yang 2017 | 3p MACE, Revasc, Angina | HR 1.3 (1.02-1.66) per 1 SD increase |
| Blood pressure variability | 5 | 2 | Lin 2019 | CHD, Stroke, Revasc, PAD, ACM | For interarm systolic blood pressure difference:<br>HR 0.54 (0.22-1.3) for 2nd vs. 1st quartile;<br>HR 1.24 (0.61-2.53) for 3rd vs. 1st quartile;<br>HR 1.83 (0.94-3.53) for 4th vs. 1st quartile |

| | | | | | |
|---|---|---|---|---|---|
| | | | Eguchi 2009 | 3p MACE | HR 1.08 (1.01-1.16) for sleep systolic BP – 1 SD;<br>HR 1.13 (1.04-1.23) for sleep diastolic BP – 1 SD |
| | | | Hata 2013 | 3p MACE | HR 1.54 (0.99-2.39) for visit-to-visit variability in SBP (highest vs. lowest decile) |
| | | | Cardoso 2020 | 3p MACE, ACM | HR 1.25 (1.03-1.51) for visit-to-visit variability in SBP – 1 SD;<br><br>No significant change in AUC from 0.723 (0.674-0.772) in standard model to 0.724 (0.675-0.773) |
| | | | Bell 2018 | CVM | HR 1.02 (0.89-1.16) (log-transformed) |
| Adiponectin | 3 | 2 | Sharma 2020 | 3p MACE, HF | HR 1.04 (0.95-1.14) (log-transformed);<br>Adding adiponectin to clinical variable, c-statistic changed from 0.66 to 0.67; NRI 0.1455 (0.0561, 0.2311);<br>IDI 0.0091 (0.0062, 0.0129) |
| | | | Lim 2008 | 3p MACE, Revasc, Angina, Renal | RR 1.6 (0.5-5.07) for 2nd vs. 1st quartile;<br>RR 2.71 (0.92-7.95) for 3rd vs. 1st quartile;<br>RR 3.03 (1.09-8.41) for 4th vs. 1st quartile |
| | | | Van der Leeuw 2016 | 3p MACE, HF, PAD | SMART cohort: change in c-statistic 0.00 (0.00-0.01) on top of base model of 0.70 (0.67-0.74);<br><br>EPIC-NL cohort: change in c-statistic 0.01 (0.00-0.02) on top of base model of 0.69 (0.64-0.74) |
| Pulse wave velocity (PWV) | 4 | 4 | Wijkman 2016 | 3p MACE | HR 1.142 (1.003-1.301) for aortic PWV (per 1 m/s);<br>AUC 0.785 (0.714-0.857) for classical risk factors;<br>AUC 0.792 (0.720-0.863) for risk factors adding aPWV |
| | | | Katakami 2014 | 3p MACE, Revasc, HF, Angina, PAD | HR 1.33 (1.09-1.62) for brachial-ankle PWV (per 1 SD);<br>Max IMT improved AUC from 0.60 (0.54-0.67) to 0.63 (0.60-0.82) beyond Framingham risk score;<br>Adding baPWV to Framingham risk score and maxIMT further improved AUC to 0.72 (0.67-0.78) |

| | | | | | |
|---|---|---|---|---|---|
| | | | Cardoso 2013 | 3p MACE, Revasc, HF, PAD, ACM | HR 1.13 (1.03-1.23) (per 1 m/s);<br>HR 1.91 (1.16-1.38) (per 10 m/s) for carotid-femoral PWV;<br>C-statistics 0.75 (0.712-0.786) |
| | | | Nakamura 2010 | CHD, Revasc, HF, ACM | HR 1.97 (1.01-3.84) for higher brachial-ankle PWV (≥1730 cm/sec) |
| Carotid plaque | 2 | 2 | Katakami 2012 | 3p MACE, Angina | HR 0.82 (0.71-0.908);<br>C-statistic 0.76 (0.6-0.9) for carotid integrated backscatter value <17.1 dB;<br><br>HR 1.938 (1.17-3.212);<br>C-statistic 0.6 (0.45-0.79) for carotid artery plaque thickness >1.3 mm |
| | | | Irie 2013 | CHD, Stroke, Revasc, Angina, PAD | HR 4.55 (2.1-19.84);<br>C-statistic 0.82 (0.75-0.88) for gray-scale median<br>HR 1.44 (1.01-2.06), *p* = 0.005 for plaque thickness;<br>AUC 0.73 (0.63-0.82) for plaque thickeness compared to AUC (0.60, 95%CI 0.49-0.70) for FRS |
| Middle cerebral artery (MCA) stenosis | 1 | 1 | Irie 2014 | CHD, CVM | HR 1.35 (1.04-1.75) for CHD;<br>HR 1.56 (1.04-2.33) for CVM |
| Homocysteine | 3 | 1 | Stehouwer 1989 | CVM | RR 1.0 (0.88-1.13) per 1 micromol/L |
| | | | Soinio 2004 | CHD, CVM | RR 2.21 (1.38-3.54) for >15 mmol/L |
| | | | Friedman 2005 | 3p MACE, Revasc, PAD | RR 0.79 (0.55-1.13) for 2nd vs. 1st quintile;<br>RR 1.17 (0.55-1.13) for 3rd vs. 1st quintile;<br>RR 1.05 (0.72-1.53) for 4th vs. 1st quintile;<br>RR 1.2 (0.83-1.75) for 5th vs. 1st quintile |
| Copeptin | 3 | 3 | Smaradotti 2019 | 3p MACE | HR 1.75 (1.3-2.24) (log-transformed copeptin) |
| | | | Mellbin 2010 | 3p MACE | HR 1.35 (1.16-1.57 (log-transformed copeptin) |

| | | | | | |
|---|---|---|---|---|---|
| | | | Velho 2018 | CHD, CVM, Revasc, HF | DIAHYCAR cohort: HR 1.13 (0.99-1.29) (log-transformed copeptin);<br>HR 1.29 (1.04-1.549) (3rd vs. 1st tertile);<br>HR 1.25 (1.01-1.54) (2nd vs. 1st tertile);<br><br>SURDIAGENE cohort: HR 1.28 (1.12-1.46) (log-transformed copeptin);<br>HR 1.58 (1.23-2.04) (3rd vs. 1st tertile);<br>HR 1.39 (1.1-1.77 (2md vs. 1st tertile) |
| Growth differentiation factor (GDF-15) | 3 | 3 | Sharma 2020 | 3p MACE, HF | HR 1.15 (1.04.1.28) (log-transformed GDF-15);<br>Adding GDF-15 to clinical variables, c-statistic changed from 0.66 to 0.67; NRI 0.1521 (0.0631, 0.243);<br>IDI 0.0183 (0.014, 0.0235) |
| | | | Resl 2016 | CHD, Stroke, HF, PAD, ACM | HR 1.37 (1.12-1.68) (log-transformed GDF-15);<br>Adding GDF-15 and hs-TnT to clinical variables, c-statistic changed from 0.773 to 0.787;<br>NRI 0.336 (0.16-0.508) for adding both hs-TnT and GDF-15 |
| | | | Carlsson 2020 | CHD, Stroke | HR 1.34 (0.96-1.88) |
| Galectin-3 | 3 | 3 | Sharma 2020 | 3p MACE, HF | HR 1.21 (1.03-1.41) (log-transformed);<br>C-stastistic 0.67;<br>adding galectin-3 to clinical variable, c-statistic changed from 0.66 to 0.67;<br>NRI 0.1265 (0.0384, 0.2062);<br>IDI 0.0162 (0.0123, 0.0204) |
| | | | Lepojarvi 2016 | CVM, HF | HR 3.9 (1.8-8.6);<br>adding Galectin-3 to clinical variable, c-statistic changed from 0.794 to 0.830;<br>IDI 0.027 (0.006-0.049);<br>NRI 0.665 (0.382-0.949) |
| | | | | | 12.2 mg/ml |
| | | | Lorenzo-Almoros 2020 | CHD, Stroke | HR 1.83 (1.13–2.98) per 1 SD increase |
| Uric acid | 3 | 2 | Resl 2012 | CHD, HF | HR 1.331 (1.095-1.616);<br>adding uric acid to standard model, c-index changed from 0.654 to 0.681 |
| | | | Panero 2012 | CVM | HR 1.13 (0.85-1.5) |

| | | | | | |
|---|---|---|---|---|---|
| | | | Ndrepepa 2012 | CVM | HR 1.15 (1.01-1.36) per 1 SD;<br>absolute IDI 0.006;<br>relative IDI 3.7%, *p* = 0.061 |
| Triglyceride/HDL ratio | 3 | 3 | Sone 2012 | CHD, Angina | HR 1.49 (1.2-1.85) for men;<br>C-statistic 0.68 (0.615-0.746) for men;<br><br>HR 1.36 (1.01-1.85) for women;<br>C-statistic 0.683 (0.597-0.769) |
| | | | Zoppini 2010 | CVM | HR 1.11 (0.65-1.88) for 2nd vs. 1st tertile;<br>HR 1.39 (0.8-2.42) for 3rd vs. 1st tertile |
| | | | Yang 2017 | 3p MACE, Revasc, Angina | HR 2.47 (1.01-6.04) |
| Heart rate recovery (HRR) | 4 | 4 | Georgoulias 2009 | CHD, CVM, Revasc | HR 0.74 (0.67-0.82) |
| | | | Chacko 2008 | CHD, CVM, HF, PAD | HR 0.5 (0.3-0.8) for HRR 13-18 vs. <13 bpm at 1 min;<br>HR 0.6 (0.4-1.0) for HRR 19-22 (vs <13 bpm) at 1 min;<br>HR 0.5 (0.4-0.9) for HRR 23-28 (vs. <13 bpm) at 1 min;<br>HR 1.0 (0.6-1.5) (vs. <13 bpm) at 1 min. |
| | | | Zafrir 2016 | CHD, Stroke, ACM | HR 1.79 (1.12-2.84) |
| | | | Hage 2013 | CHD, CVM | HR 7.1 (1.7-30.0) for HRR <20% |
| Heart rate variability (HRV) | 2 | 2 | Eguchi 2010 | 3p MACE | HR 1.29 (1.05-1.60) for SD of sleep heart rate |
| | | | Cha 2018 | CHD, Stroke | HR 2.62 (1.30-5.31) for SD of normal RR intervals |
| Cardiovascular autonomic neuropathy (CAN) | 3 | 3 | Cha 2016 | 3p MACE, HF, PAD | HR 1.93 (0.86-4.36) for early vs. normal CAN;<br>HR 3.03 (1.39-6.6) for definite CAN vs. normal |
| | | | Chyun 2015 | CHD, CVM, Revasc, HF | HR 1.6 (1.02-2.5) for lower Valsalva heart rate ratio (Q1) |
| | | | Yun 2018 | CHD, Stroke | HR 3.32 (1.81-6.14) for progression of CAN (vs. non-progression) |
| Soluble suppressor of tumorigenicity-2 (sST2) | 2 | 1 | Lepojarvi 2016 | CVM, HF | HR 4.1 (2-8.5) for sST2;<br>adding sST2 to clinical variable, c-statistic changed from 0.794 to 0.842;<br>IDI 0.032 (0.011-0.052);<br>NRI 0.916 (0.610-1.221) |

| | | | | | |
|---|---|---|---|---|---|
| | | | Jha 2018 | CHD, Stroke, Angina | Non-significant associated with MACE *p* = 0.5 |
| Pulse pressure | 3 | 1 | Nargesi 2016 | CHD | HR 1.39 (1.0-1.94) low vs. middle pulse pressure;<br>HR 1.23 (0.86-1.75) high vs. middle pulse pressure |
| | | | Cockcroft 2005 | CHD | OR 1.69 (P=0.002) per 10 mmHg |
| | | | Theilade 2016 | 3p MACE, HF, Renal | OR 1 (0.96-1.04) |
| Urinary osmomucoid excretion rate (UOER) | 3 | 3 | Svendstrup 2013 | CVM | HR 2.08 (1.31-3.31) |
| | | | Christiansen 2002 | CVM | HR 4.94 (1.6-15.22) |
| | | | Christiansen 2005 | CVM | HR 9.81 (1.31-73.6) |
| Shorter leukocyte telomere length (LTL) | 2 | 2 | Masi 2016 | CHD | Lower LTL at baseline predicted an increased IHD risk at follow-up (age adjusted: *p* = 0.033 and *p* = 0.040) |
| | | | Cheng 2020 | CHD, Stroke, HF, PAD | HR 1.252 (1.195-1.311) |
| von Willebrand factor (vWF) | 2 | 2 | Saito 2000 | 3p MACE, Revasc, HF | RR 1.71 (1.11-2.63) for high vs. low vWF |
| | | | Standl 1996 | CVM, Stroke | HR: beta coefficient -0.003 (SE 0.002), *p* = 0.03 |
| Interleukin-6 (IL-6) | 2 | 1 | Lowe 2014 | 3p MACE, Renal | HR 1.37 (1.24-1.51);<br>adding IL-6 to base model, c-index changed from 0.682 to 0.692 (*p* = 0.012);<br>IDI 0.011 (0.007–0.015), *p* <0.001;<br>Relative IDI (%) 13.7 (8.8–19.1);<br>Continuous NRI 0.194 (0.073–0.320), *p* = 0.006;<br>Categorical NRI 0.034 (20.006–0.077), *p* = 0.09 |
| | | | Von Scholten 2016 | 3p MACE, HF | HR 1.0 (0.8-1.4) |
| Plasma thyroid-stimulating hormone (TSH) level | 1 | 0 | De Vries 2019 | 3p MACE, ACM | HR 0.93 (0.80-1.08) |
| Subclinical hypothyroidism | 1 | 1 | Chen 2007 | CHD, Stroke, HF, Angina | HR 2.93 (1.15-7.48) |

| | | | | | |
|---|---|---|---|---|---|
| 25-hydroxyvitamin D | 2 | 1 | Heidari 2015 | CHD, CVM, Angina | HR 0.43 (0.31-0.60) for highest quartileile of vitamin D compared to lowest;<br>Adding 25(OH)VitD to Framingham Risk Score, c-index changed from 0.74 to 0.75 (*p* = 0.08);<br>IDI 0.0052 (0.0016-0.0087), *p* = 0.003;<br>Relative IDI (%) 7%;<br>Continuous NRI (%) 29% (19%-40%), *p* <0.001 |
| | | | Alele 2013 | CHD, HF | HR 1.13 (0.53-2.42) in lowest quartile |
| plasma adipocyte-fatty acid binding protein lipocalin 2 (A-FABP) | 2 | 1 | Ong 2020 | 3p MACE, Revasc | HR 1.00 (0.83-1.20) |
| | | | Vavruch 2020 | 3p MACE | HR 1.60 (1.08-2.38) |
| basic fibroblast growth factor (bFGF) | 3 | 2 | Zimering 2011 | CHD | HR 1.013 (1.007-1.019) |
| | | | Zimering 2013 | CHD, CVM, Revasc | HR 1.008 (1.002-1.014) |
| | | | Van der Leeuw 2016 | 3p MACE, HF, PAD | SMART cohort: change in c-statistic 0.00 (0.00-0.00) on top of base model of 0.70 (0.67-0.74);<br><br>EPIC-NL cohort: change in c-statistic 0.01 (-0.01 to 0.02) on top of base model of 0.69 (0.64-0.74);<br><br>Continuous NRI 0.038 (-0.137 to 0.202) in the SMART cohort and 0.048 (-0.159 to 0.261) in the EPIC-NL cohort;<br><br>Risk Category-Based NRI 0.005 (-0.025 to 0.038) in the SMART cohort and -0.012 (-0.068 to 0.041) in the EPIC-NL cohort; |
| Osteoprotegerin | 2 | 0 | Reinhard 2010 | CVM | HR 1.25 (0.71-2.21) for 3rd vs. 1st tertile |
| | | | Anand 2006 | 3p MACE, Revasc | 5.45–9.51 vs. <5.44: HR 1.03 (0.14–7.34);<br>>9.52 vs. <5.44: HR 5.76 (1.28–26.0); trend test *p* = 0.01;<br><br>C-index 0.80 (0.66-0.95), *p* <0.0001 |
| Triglyceride glucose (TyG) index | 2 | 2 | Jin 2018 | 3p MACE, Revasc, Angina | HR 1.693 (1.238-2.316);<br>adding TyG index to traditional risk factors, c-statistic changed from 0.615 to 0.638 (*p* = 0.002) |

| | | | | | |
|---|---|---|---|---|---|
| | | | Zhao 2020 | CHD, Revasc, ACM | 37.7% vs. 9.6% (*p* <0.001) for higher vs. lower TyG index; adding TyG index to baseline risk model, c-statistic changed from 0.800 to 0.856 (p<0.001); category-free NRI 0.346 (0.230-0.430), *p* <0.001; IDI 0.087 (0.039-0.128), *p* <0.001 |
| Skin autofluorescence | 2 | 2 | Meerwaldt 2007 | CHD, ACM | HR 2.9 (1.3-4.4) |
| | | | Lutgers 2009 | CVM | HR 1.21 (1.05-2.22) for skin autofluorescence > median; Addition of skin autofluorescence to UKPDS risk engine resulted in re-classification of 55 of 203 patients from low-risk to high-risk; Adding skin autofluorescence to base model, c-index changed from 0.712 to 0.718 |
| Insulin resistance | 2 | 2 | Saely 2005 | 3p MACE, Revasc, HF, ACM | HR 2.57 (1.47-4.51) |
| | | | Fragoso 2015 | CHD, Stroke, HF, PAD | OR 2.847 (1.048-7.735) |
| ApoB:A-I ratio | 2 | 1 | Charlton-Menys 2009 | CHD, Stroke | HR 1.930 (1.316-2.83) for 3rd vs. 1st tertile; AUC± SE 0.5925 ± 0.0209 compared to non-HDLC:LDLC (0.5456 ± 0.0209), *p* = 0.0005 |
| | | | Moosaie 2020 | CHD, Revasc, Angina | OR 1.081 (0.322-2.002); AUC 0.52 (0.481-0.559), *p* = 0.056 |
| apolipoprotein B (Apo B) | 2 | 1 | Jiang 2004 | CHD, Stroke, Revasc | RR 2.31 (1.23-4.35) for 4th vs. 1st quartile; AUC was 0.691 for apoB, compared to AUC 0.685 for LDL-C |
| | | | Moosaie 2020 | CHD, Revasc, Angina | OR 1.006 (1.001-1.016); AUC for apoB 0.509 (0.430-0.559), *p* = 0.058 |
| Apo-A-I | 2 | 1 | Koch 1997 | CHD, CVM | RR 1.016 (SE 0.00682) for 1 mg/dl increment |
| | | | Moosaie 2020 | CHD, Revasc, Angina | OR 0.975 (0.962-1.003). AUC 0.56 (0.523-0.597), *p* = 0.098 |

| | | | | | |
|---|---|---|---|---|---|
| LDL/HDL ratio | 2 | 2 | Sone 2012 | CHD, Angina | HR 1.52 (1.29-1.79) for men;<br>HR 1.44 (1.09-1.91) for women;<br>AUC of 0.709 (0.646-0.772) for men;<br>AUC of 0.695 (0.608-0.781) for women |
| | | | Hayashi 2013 | CHD, Stroke, Revasc, Angina | HR 1.583 (1.298-1.945) per quartile |
| lipoprotein(a) - Lp(a) | 3 | 1 | Saely 2006 | 3p MACE, Revasc, HF | HR 0.812 (0.539-1.223) |
| | | | Gazzaruso 2003 | CHD, Revasc | Lp(a) and aop(a) polymorphisms do not appear to be reliable markers of restenosis |
| | | | Moosaie 2020 | CHD, Revasc, Angina | OR 1.007 (1.001-1.013);<br>AUC 0.59 (0.548-0.633) |
| non-fasting remnant cholesterol | 1 | 0 | Hong 2017 | CHD, CVM, Revasc | HR 1.05 (0.46-2.37);<br>AUC 0.64 (0.55-0.72), $p$ = 0.003 |
| Remnant-like lipoprotein particles (RPL) cholesterol | 1 | 1 | Fukushima 2004 | CHD, CVM, Revasc, Angina | OR 2.4 (1.3-4.6) for RLP >4.7 mg/dl |
| Body weight variability | 2 | 2 | Yeboah 2019 | 3p MACE | HR 1.25 (1.15-2.36) |
| | | | Nam 2020 | CHD, Stroke, ACM | HR 1.15 (1.10-1.20) |
| Waist-to-hip ratio (WHR) | 1 | 1 | Khalili 2012 | 3p MACE, Angina | HR 1.21 (1.00-1.48) for men;<br>HR 1.32 (1.06-1.65) for women;<br>C-index 0.64 (0.58-0.70) |
| resistin | 2 | 1 | Lim 2008 | 3p MACE, Revasc, Angina, Renal | NR |
| | | | Bonito 2019 | CHD | HR 1.931, $p$ = 0.031 |
| Chronotropic incompetence (<80% heart rate reserve) | 1 | 1 | Zabir 2016 | 3p MACE, ACM | HR 1.18 (1.18-3.01) |
| fourth harmonic amplitude of the radial pulse wave (C4CV) | 1 | 1 | Liao 2019 | CHD, Stroke, Revasc, Angina | HR 1.31 (1.20-1.43) |
| First Harmonic (C1) of Radial Pulse | 1 | 1 | Chang 2018 | CHD, CVM, HF | HR 2.29 (1.55-3.37) for >1.07 vs. <0.89 |

| | | | | | |
|---|---|---|---|---|---|
| Peripheral arterial endothelial dysfunction assessed using reactive hyperemia-peripheral arterial tonometry (RH-PAT) | 1 | 1 | Koo 2020 | 3p MACE | HR 3.24 (1.14-9.17) |
| Left ventricular (LV) mass and relative wall thickness (echocardiography) | 1 | 1 | Eguchi 2007 | 3p MACE, HF, Angina, Renal | RR 1.06 (1.02-1.11) |
| Left ventricular hypertrophy | 1 | 1 | Bianco 2014 | CVM | HR 3.5 (1.3-9.7) |
| Left ventricular ejection fraction (LVEF) | 1 | 1 | Shin 2020 | CVM, HF | HR 3.18 (2.03-4.98) |
| Stress-corrected midwall shortening (sc-MS) | 1 | 1 | Cioffi 2014 | CVM | HR 1.03 (1.01-1.08) |
| Depression | 2 | 1 | Lin 2010 | CHD, Stroke, ACM | HR 1.36 (1.05-1.75) |
| | | | Bruce 2005 | CVM | HR 1.21 (0.95-1.55) |
| leisure-time physical activity (LTPA) | 1 | 1 | Sone 2013 | CHD | HR 0.77 (0.43-1.38) |
| CD34+ and CD34+CD133+ stem cells | 1 | 1 | Fadini 2017 | 3p MACE | CD34+ HR: 2.21 (1.14-429);<br>CD34+CD133+ HR 2.98 (1.46-6.08);<br>addition of CD34+ improved UKPDS c-statistics, NRI, and IDI |
| Neutrophil-lymphocyte ratio (NLR) | 1 | 1 | Azab 2013 | 3p MACE | HR 2.8 (1.12-6.98) |
| leukocyte count | 1 | 1 | Saito 2000 | 3p MACE, Revasc, HF | HR 1.90 (1.16-3.13) |
| Plasma 8-OH-Deoxyguanosine (8-oxo-20 -dG) | 1 | 1 | Thomas 2018 | CVM | HR 1.10 (1.01-1.20) per 1-SD increase |
| Serum sialic acid (marker of innate immunity) | 1 | 1 | Pickup 2003 | CVM | HR 1.53 (1.12-2.10) |
| hemoglobin | 1 | 1 | Savonitto 2018 | CVM | HR 1.01 (1.00-1.02) |
| Lactoferrin | 1 | 1 | Vengen 2010 | CHD | HR 2.54 (1.00-6.45) 2nd vs. 1st tertitle; 4.06 (1.72-9.60) 3rd vs. 1st tertile |
| lipocalin 2 (LCN2) | 1 | 0 | Ong 2020 | 3p MACE, Revasc | 1.10 (0.97-1.25) |

| | | | | | |
|---|---|---|---|---|---|
| high platelet reactivity (HPR) | 1 | 0 | Angiolillo 2007 | 3p MACE | HR 3.35 (1.68-6.66) |
| Platelet distribution width (PDW) | 1 | 1 | Hu 2018 | CHD, Revasc | OR 1.289 (1.110-1.498) for 1-unit increase |
| factor VIII | 1 | 1 | Saito 2000 | 3p MACE, Revasc, HF | HR 1.58 (1.02-2.42) for highest compared to lowest grouping |
| thrombomodulin | 2 | 1 | vonScholten 2016 | 3p MACE, HF | HR 1.40 (1.0-1.9) |
| | | | Van der Leeuw 2016 | 3p MACE, HF, PAD | SMART cohort: change in c-statistic 0.01 (-0.01 to 0.02) on top of base model of 0.70 (0.67-0.74);<br><br>EPIC-NL cohort: change in c-statistic 0.00 (-0.01 to 0.01) on top of base model of 0.69 (0.64-0.74) |
| sICAM-1 | 2 | 0 | vonScholten 2016 | 3p MACE, HF | HR 1.0 (0.8-1.4) |
| | | | Van der Leeuw 2016 | 3p MACE, HF, PAD | SMART cohort: change in c-statistic 0.01 (-0.01 to 0.02) on top of base model of 0.70 (0.67-0.74);<br>NRI 0.209 (0.050-0.376) |
| | | | | #N/A | EPIC-NL cohort: change in c-statistic 0.01 (0.0 0 to 0.02) on top of base model of 0.69 (0.64-0.74);<br>NRI 0.115 (-0.124 to 0.340) |
| sICAM-3 | 2 | 0 | vonScholten 2016 | 3p MACE, HF | HR 1.4 (1.0-2.0) |
| | | | Van der Leeuw 2016 | 3p MACE, HF, PAD | SMART cohort: change in c-statistic 0.00 (-0.00 to 0.01) on top of base model of 0.70 (0.67-0.74);<br>NRI 0.043 (-0.124 to 0.203);<br><br>EPIC-NL cohort: change in c-statistic 0.00 (0.00 to 0.00) on top of base model of 0.69 (0.64-0.74);<br>NRI -0.158 (-0.377 to 0.059) |
| sVCAM-1 | 2 | 0 | vonScholten 2016 | 3p MACE, HF | HR 1.1 (0.8-1.6) |
| | | | Van der Leeuw 2016 | 3p MACE, HF, PAD | SMART cohort: change in c-statistic 0.00 (-0.01 to 0.01) on top of base model of 0.70 (0.67-0.74);<br>NRI= 0.238 (0.074–0.400);<br><br>EPIC-NL cohort: change in c-statistic 0.00 (-0.01 to 0.1) on top of base model of 0.69 (0.64-0.74);<br>NRI=0.124 (-0.098 to 0.353) |

| | | | | | |
|---|---|---|---|---|---|
| sE-selectin | 2 | 0 | vonScholten 2016 | 3p MACE, HF | HR 0.8 (0.6-1.2) |
| | | | Van der Leeuw 2016 | 3p MACE, HF, PAD | SMART cohort: change in c-statistic 0.00 (-0.01 to 0.01) on top of base model of 0.70 (0.67-0.74);<br>NRI 0.118 (-0.044 to 0.285);<br><br>EPIC-NL cohort: change in c-statistic 0.00 (0.00 to 0.00) on top of base model of 0.69 (0.64-0.74);<br>NRI -0.003 (-0.221 to 0.236) |
| sP-selectin | 2 | 0 | vonScholten 2016 | 3p MACE, HF | HR 1.1 (0.8-1.6) |
| | | | Van der Leeuw 2016 | 3p MACE, HF, PAD | SMART cohort: change in c-statistic 0.00 (0.00 to 0.01) on top of base model of 0.70 (0.67-0.74);<br>NRI -0.001 (-0.161 to 0.155);<br><br>EPIC-NL cohort: change in c-statistic 0.02 (-0.01 to 0.04) on top of base model of 0.69 (0.64-0.74);<br>NRI 0.226 (0.008–0.418) |
| IL-1-beta | 1 | 0 | vonScholten 2016 | 3p MACE, HF | HR 1.3 (1.0-1.8) |
| IL-8 | 1 | 0 | vonScholten 2016 | 3p MACE, HF | HR 1.2 (0.9-1.6) |
| TNF-alpha | 1 | 1 | vonScholten 2016 | 3p MACE, HF | HR 1.5 (1.1-2.1) |
| serum amyloid A (SAA) | 2 | 0 | vonScholten 2016 | 3p MACE, HF | HR 0.7 (0.5-1.1) |
| | | | Van der Leeuw 2016 | 3p MACE, HF, PAD | SMART cohort: change in c-statistic 0.01 (-0.00 to 0.02) on top of base model of 0.70 (0.67-0.74);<br><br>EPIC-NL cohort: change in c-statistic 0.01 (-0.01 to 0.03) on top of base model of 0.69 (0.64-0.74) |
| soluble tumor necrosis factor receptor 1 (sTNFR1) | 1 | 1 | Carlsson 2016 | 3p MACE | HR 1.66 (1.29-2.174) |
| soluble tumor necrosis factor receptor 2 (sTNFR2) | 1 | 1 | Carlsson 2016 | 3p MACE | HR 1.47 (1.13-1.91) |

| | | | | | |
|---|---|---|---|---|---|
| Endogenous testosterone concentration | 1 | 1 | Daka 2015 | CHD | HR 0.754 (0.61-0.92) |
| Glucagon-like peptide-1 (GLP-1) | 1 | 1 | Ravassa 2015 | CHD, Stroke, Revasc, HF, PAD | HR 6.9 (2.0-23.8) for lower GLP-1 levels vs. normal |
| N(epsilon)-carboxymethyllysine (CML) | 1 | 0 | Busch 2006 | 3p MACE, Revasc, HF, PAD | HR 1.23 (0.76-2.00) |
| Fibroblast growth factor 21 (FGF21) | 1 | 1 | Ong 2015 | 3p MACE, Revasc | HR 1.28 (1.10-1.50) for 3rd vs. 1st tertile; non-significant increase in c-statistic but significant improvement in IDI (0.001; 95% CI 0.000-0.003) and NRI: 10.9% with NRI >0% |
| serum PRO-C (S-PRO-C6) | 1 | 1 | Rasmussen 2018 | 3p MACE, HF | HR 3.06 (1.31-7.14) for doubling of S-PRO-C6; improved IDI by 22.5%; c-statistic 0.76 (0.70-0.82) for CV events |
| Fluorescent advanced glycation end products (AGEs) | 1 | 0 | Cournot 2018 | 3p MACE | HR 1.03 (0.77-1.37) 3rd vs. 1st tertile; C-statistic 0.748; rIDI: -0.002 (per tertile); rIDI: -0.006 per SD |
| Advanced oxidation protein products (AOPP) | 1 | 0 | Cournot 2018 | 3p MACE | HR 0.77 (0.57-1.04) (above median) |
| Carbonyls | 1 | 0 | Cournot 2018 | 3p MACE | HR 1.07 (0.82-1.40) 3rd vs. 1st tertile; c-statistic 0.748; rIDI: 0.005 per tertitle; rIDI 0.002 per SD |
| Ischemia-modified albumin (IMA) | 1 | 0 | Cournot 2018 | 3p MACE | HR 1.03 (0.78-1.36) 3rd vs. 1st tertile |
| Oxidative hemolysis inhibition assay (OxHLIA) | 1 | 0 | Cournot 2018 | 3p MACE | HR 1.06 (0.8-1.40) |
| Total reductive capacity of plasma (TRCP) | 1 | 0 | Cournot 2018 | 3p MACE | HR 1.10 (0.83-1.47) |
| Plasma total anti-oxidant status (TAOS) | 1 | 1 | Masi 2016 | CHD | HR 0.78 (0.63-0.96) 1 quintile increase |
| Plasma YKL-40 concentration | 1 | 1 | Lin 2013 | CVM, ACM | HR 2.45 (1.11-5.37) |
| Serum paraoxonase (PON1) activity | 1 | 1 | Ikeda 2009 | CHD, Stroke, HF, PAD | HR 0.988 (0.976-1.000) per 1 U/L |

| | | | | | |
|---|---|---|---|---|---|
| Connective tissue growth factor (CTGF/CCN2) | 1 | 1 | Hunt 2018 | 3p MACE, HF, PAD | HR 2.71 (1.44-5.08) for highest vs. lowest category |
| Pregnancy associated plasma protein A (PAPP-A) | 1 | 1 | Li 2017 | 3p MACE | HR 2.97 (2.11-4.18) per 1 SD increase in log(PAPP-A); C-statistic 0.79 |
| sclerostin | 1 | 1 | Novo-Rodriguez 2018 | CVM | HR 1.318 (1.090-1.595) |
| Hypomagnesemia | 1 | 0 | Peters 2013 | CHD, Revasc, Angina | HR 0.52 (0.23-1.18) per 1 mmol/L |
| Phosphorus | 1 | 1 | Silva 2013 | CHD, CVM | HR 0.235 (0.097-0.571) for 1st vs. 3rd tertile |
| hypoglycemic events (ICD codes) | 1 | 1 | Johnston 2011 | CHD, Revasc, Angina | HR 1.79 (1.69-1.89) |
| Glycated albumin (GA) | 1 | 1 | Yang 2015 | 3p MACE | HR 1.256 (1.183-1.335) |
| endogenous secretory receptor for advanced glycation endproducts (esRAGE) | 2 | 2 | Yang 2015 | 3p MACE | HR 0.733 (0.539-0.996) |
| | | | Peng 2009 | 3p MACE, Revasc | OR 23.477 (*p* <0.001) for change of esRAGE level from baseline |
| Fasting insulin level | 1 | 1 | Faghihi-Kashani 2016 | CHD, CVM, Angina | HR 1.18 (1.05-1.31) |
| Apolipoprotein CIII | 1 | 1 | Colombo 2018 | CHD, Stroke | HR 1.34 (1.12-1.60) |
| Non-HDL/HDL ratio | 1 | 1 | Eliasson 2011 | CHD | HR 0.52 (0.38-0.71) for decile 1 vs. tertile 3 (ref); C-statistic 0.70 |
| Total cholesterol/ HDL cholesterol ratio (TC/HDL) | 1 | 1 | Wan 2016 | 3p MACE, HF | Males: HR 1.04 (1.02-1.07); Females: HR 1.05 (1.02-1.09) |
| atherogenic index of plasma (AIP) | 1 | 1 | Qin 2020 | 3p MACE, Revasc | HR 1.614 (1.303-2.001) for high AIP vs. low |
| Waist-to-height ratio (WHtR) | 1 | 0 | Khalili 2012 | 3p MACE, Angina | HR 1.08 (0.89-1.31); c-statistic 0.73 (0.68-0.77) |
| Excess waist circumference (WC) | 1 | 0 | Sone 2009 | CHD, Stroke | HR 0.88 (0.51-1.50) per 1 SD (men); HR 1.26 (0.94-1.70) per 1 SD (women) |
| Neck circumference | 1 | 1 | Yang 2019 | 3p MACE, Revasc, HF, Angina | HR 2.305 (1.535-3.460) |
| sagittal abdominal diameter (SAD) | 1 | 1 | Radholm 2017 | 3p MACE | HR 2.81 (1.37-5.76) |

| | | | | | |
|---|---|---|---|---|---|
| albumin | 1 | 1 | Saito 2000 | 3p MACE, Revasc, HF | HR 0.64 (0.44-0.92) |
| leptin | 1 | 1 | Vavruch 2020 | 3p MACE | HR 1.71 (1.15-2.53) for 1 SD increase |
| Apelin | 1 | 1 | Silva 2013 | CHD, CVM | HR 0.981 (0.967-0.996) for CV mortality;<br>HR 0.548 (0.302-0.817) for CV hospitalization |
| Omentin-1 | 1 | 1 | Biscetti 2020 | 3p MACE | Log-rank P<0.001 (HR not reported);<br>C-statistic 0.804 |
| trimethylamine N-oxide (TMAO) | 1 | 0 | Cardona 2019 | 3p MACE, Revasc | TMAO levels did not predict incident MACE |
| H. pylori seropositive/cytotoxin-associated gene-A (CagA) seropositivity | 1 | 0 | Schimke 2010 | 3p MACE | OR 0.91 (0.63-1.31) |
| branched-chain amino acids (BCAAs; isoleucine, leucine, and valine) | 1 | 1 | Tobias 2018 | CHD, Stroke | HR 1.13 (1.08-1.18) per 1 SD |
| Electrochemical skin conductance (ESC) composite score | 1 | 1 | Lim 2019 | CHD, Stroke, PAD, Renal | HR 3.11 (1.27-7.62) |
| Upstroke time per cardiac cycle (UTCC) | 1 | 1 | Chang 2019 | CHD, Stroke, Revasc, PAD, ACM | HR 2.45 (1.38-4.35) |
| Thoracoabdominal calcifications in native radiograms | 1 | 1 | Juutilainen 2010 | CVM, ACM | HR 1.5 (0.8-3.0) in men;<br>HR 3.0 (1.6-5.7) in women |
| Epicardial fat volume (computed tomography coronary angiogram) | 1 | 0 | Venuraju 2021 | CHD, Revasc, ACM | Epicardial fat volume was a significant univariate (*p* = 0.01), but not multivariate, predictor of the number of coronary plaques |
| Real-time microembolization during PCI (‘‘HITS’’) detected by an intracoronary Doppler | 1 | 1 | Otto 2012 | CHD, Revasc, ACM | OR 1.07 (1.011-1.13) |
| Doppler-derived coronary flow velocity reserve (CFVR) of LAD | 1 | 1 | Cortigiani 2014 | CHD, Revasc, ACM | HR 11.20 (3.07-40.92) for CFVR |

| | | | | | |
|---|---|---|---|---|---|
| Global Longitudinal Strain (GLS) for LV Function | 1 | 1 | Backhaus 2020 | 3p MACE | HR 1.11 (1.03-1.20) |
| Biventricular volume (BiVV) | 1 | 1 | Cox 2013 | CVM, ACM | HR 4.36 (1.36-14.03);<br>C-statistic increased from 0.76 to 0.78 (*p* = 0.02) |
| Exercise capacity (peak V02) | 1 | 1 | Seyoum 2006 | 3p MACE, HF, PAD | HR 0.931 (0.879-0.987) |
| screening for silent ischemia with bicycle exercise test or dipyridamole single photon emission computed tomography | 1 | 0 | Lievre 2011 | CHD, Stroke, HF, ACM | HR 1.00 (0.59-1.71) |
| Dobutamine echocardiography (DE) | 1 | 0 | LeFeuvre 2005 | CHD, CVM, Revasc | SPECT seems more accurate than DE to detect significant coronary stenosis in high risk asymptomatic diabetic patients. |
| Exercise treadmill test (ETT) | 1 | 0 | Faglia 2002 | CHD, CVM, Angina | Incidence of cardiac events between subjects with normal ETT and subjects with abnormal ETT but with normal scintigraphy was not statistically significant (*p* = 0.715). |
| Cardiac stress test | 1 | 1 | Bates 2016 | CHD, ACM | HR 0.61 (*p* = 0.004) |
| Exercise echocardiography | 1 | 1 | Oliveira 2009 | CHD, CVM, Revasc | RR 3.63 (1.44-9.16) |
| Silent MI | 1 | 1 | Rutter 2002 | CHD, CVM, Angina | RR 21 (2-204) |
| Killip classification | 1 | 0 | Backhaus 2020 | 3p MACE | HR 1.27 (0.85-1.90) |
| Diastolic orthostatic hypertension | 1 | 1 | Wijkman 2016 | 3p MACE | HR 0.335 (0.133-0.839) |
| Cognitive dysfunction | 1 | 1 | deGalan 2009 | 3p MACE, ACM | Mild cognitive dysfunction- HR: 1.27 (1.11-1.46);<br><br>severe cognitive dysfunction 1.42 (1.10-1.99) |
| Self-rated health | 1 | 1 | Venskutonyte 2013 | CHD, Stroke | HR 0.87 (0.80-0.95) |
| WHO cardiovascular questionnaire | 1 | 1 | Azevedo 2006 | CHD, CVM, HF | RR 2.13 (1.11-4.07) |

| | | | | | |
|---|---|---|---|---|---|
| Socioeconomic status | 1 | 1 | Rawshani 2016 | CVM | HR 0.67 (0.63-0.71) for married vs. single;<br>HR 1.87 (1.72-2.05) for lowest vs. highest income;<br>HR 0.46 (0.38-0.56) for native Swedes vs. non-Western immigrants |
| Grip strength | 1 | 1 | Celis-Morales 2017 | CHD, CVM | HR 4.05 (2.72-5.80) for low vs. high grip strength |
| Total activity score (TAS) | 1 | 1 | Iijima 2012 | CHD, Stroke, HF, Angina, PAD | HR 0.54 (0.35-0.84) for highest vs. lowest quartile of TAS |
| Cardiorespiratory fitness (CRF) (quantified by percent predicted METs) | 1 | 1 | Zafrir 2015 | CHD, Stroke, ACM | HR 2.25 (1.41-3.57) for low CRF;<br>addition of CRF to model with UKPDS improved c-statistic from 0.66 to 0.71 (*p* = 0.03);<br>NRI of 0.451 (*p* = 0.002) |
| Erectile dysfunction (International Index Erectile Function-5 Questionnaire) | 1 | 1 | Gazzaruso 2008 | 3p MACE, Revasc, HF, Angina, PAD | HR 2.1 (1.6-2.6) |
| Transcutaneous oxygen tension (TcPO2) | 1 | 1 | Gazzaruso 2008 | 3p MACE, Revasc, HF, Angina, PAD | HR 1.78 (1.44-2.23) for low TcPO2 |
| acute kidney injury (AKI) | 1 | 1 | Monseu 2015 | CHD, Stroke, Revasc, HF, PAD | HR 4.81 (3.39-6.82) for CV death;<br>HR 2.97 (2.23-3.95) for MACE |
| ACSP (urinary proteomic) | 1 | 0 | Oellgaard 2018 | 3p MACE, Revasc, PAD | did not improve prediction on top of standard risk factors |
| ASCP75 (urinary proteomic) | 1 | 0 | Oellgaard 2018 | 3p MACE, Revasc, PAD | did not improve prediction on top of standard risk factors |
| CAD238 (urinary proteomic) | 1 | 0 | Oellgaard 2018 | 3p MACE, Revasc, PAD | did not improve prediction on top of standard risk factors |
| Urinary levels of kidney injury molecule 1 (u-KIM-1) | 1 | 1 | Rotbain Curovic 2018 | 3p MACE, HF, ACM | HR 2.26 (1.24-4.15);<br>IDI 0.084 (*p* = 0.21) |
| Urinary levels of neutrophil gelatinase-associated lipocalin (u-NGAL) | 1 | 0 | Rotbain Curovic 2018 | 3p MACE, HF, ACM | u-NGAL was not a predictor of any of the outcomes after adjustment |
| Urinary sodium concentration | 1 | 1 | Saulnier 2017 | CVM | HR 0.73 (0.63-0.85) per 1 SD;<br>improved prediction beyond identified risk factors IDI 4.6%, *p* = 0.02. |

| | | | | | |
|---|---|---|---|---|---|
| Urinary hepatocyte growth factor (HGF) | 1 | 1 | vonScholten 2016 | 3p MACE, HF | HR 2.0 (1.2-3.2) for higher urinary HGF |
| Urinary adiponectin | 1 | 1 | vonScholten 2016 | 3p MACE, HF | HR 1.4 (1.1-2.3) for higher urinary adiponectin |
| Urinary liver-type fatty acid-binding protein (L-FABP) | 1 | 1 | Araki 2013 | 3p MACE, Angina, PAD | HR 1.93 (1.13-3.29) for 3rd vs. 1st tertile |
| Heavy alcohol consumption | 1 | 1 | Umamahesh 2014 | CHD, Stroke, Angina, PAD | OR 8.7 (1.1-69.8) for heavy alcohol consumption (unclear if adjusted or univariate) |
| Ambulatory blood pressure monitoring (ABPM) | 1 | 1 | Salles 2013 | 3p MACE, ACM | 24 hour systolic blood pressure: HR 2.27 (1.13-4.57), which outperformed clinic-based measurement HR 1.02 (0.42-2.47) |
| Cold-pressor test (coronary artery constriction) | 1 | 1 | Nitenberg 2005 | 3p MACE, Revasc, Angina | 32.6% with coronary artery vasoconstriction vs. 7.7% without constriction (*p* <0.05) |
| Cardiac I-123 metaiodobenzylguanidine (MIBG) imaging | 1 | 0 | Nagamachi 2006 | CHD | RR 4.07 (0.81-20.4) for heart/mediastinum update <1.7 |
| Age at diabetes diagnosis | 1 | 0 | Chan 2014 | CHD, Stroke, HF, PAD | HR 0.76 (0.52-1.12) |
| Alanine aminotransferase (ALT) | 1 | 1 | Afarideh 2016 | CHD, Stroke, Revasc | HR 0.204 (0.060-0.689) for ALT ≥30;<br>Addition of ALT to Framingham risk score resulted in NRI 0.0905 (0.0801-0.1022), *p* <0.05; |
| Systolic blood pressure (SBP) <130 mmHg + proteinuria | 1 | 1 | Vepsäläinen 2012 | CVM, ACM | HR: 0.43 (0.22-0.84) for SBP 130-139 + proteinuria;<br>HR 0.61 (0.38-0.97) for SBP 140-159 + proteinuria;<br>HR 0.62 (0.38-1.02) for SBP ≥160 + proteinuria vs. reference group of SBP <130 mmHg + proteinuria. |
| Popliteal artery flow volume | 1 | 1 | Yoshimura 2006 | 3p MACE, PAD, Revasc. | HR 1.55 (0.25-9.68) for intermediate flow volume (85.3-63.3 ml/min;<br>HR 8.60 (1.61-45.97) for lower flow volume vs. reference group of higher flow volume (129.6-85.5 ml/min) |

| | | | | | |
|---|---|---|---|---|---|
| Toe-brachial index (TBI) | 1 | 1 | Zobel 2017 | 3p MACE, HF | HR 1.50 (1.27-1.65) per 1 SD decrease in TBI;<br>Addition to tradition risk factors increased c-statistic by 0.063 (0.012-0.115) from 0.743 and adding ABI did not improve c-statistic significantly;<br>rIDI 0.467 ($p \leq 0.001$) |
| Quality of Life (QOL) (EQ-5D) | 1 | 1 | Clarke 2009 | 3p MACE, Angina | HR 0.93 (per 0.10 point);<br>no c-statistic provided |
| PARP | 1 | 1 | Cui 2020 | 3p MACE, Revasc, ACM | OR 1.23 (1.14-1.32) per 1-SD increase |
| MMP-1 | 1 | 0 | Van der Leeuw 2016 | 3p MACE, HF, PAD | SMART cohort: change in c-statistic 0.00 (0.00-0.01) on top of base model of 0.70 (0.67-0.74);<br><br>EPIC-NL cohort: change in c-statistic 0.01 (-0.01 to 0.01) on top of base model of 0.69 (0.64-0.74) |
| MMP-3 | 1 | 1 | Van der Leeuw 2016 | 3p MACE, HF, PAD | SMART cohort: change in c-statistic 0.01 (0.00-0.03) on top of base model of 0.70 (0.67-0.74);<br><br>EPIC-NL cohort: change in c-statistic 0.01 (-0.01 to 0.02) on top of base model of 0.69 (0.64-0.74) |
| MMP-9 | 1 | 1 | Van der Leeuw 2016 | 3p MACE, HF, PAD | SMART cohort: change in c-statistic 0.00 (0.00-0.01) on top of base model of 0.70 (0.67-0.74);<br><br>EPIC-NL cohort: change in c-statistic 0.01 (0.00 to 0.02) on top of base model of 0.69 (0.64-0.74) |
| PLGF | 1 | 1 | Van der Leeuw 2016 | 3p MACE, HF, PAD | SMART cohort: change in c-statistic 0.00 (-0.01 to 0.00) on top of base model of 0.70 (0.67-0.74);<br><br>EPIC-NL cohort: change in c-statistic 0.01 (0.00 to 0.02) on top of base model of 0.69 (0.64-0.74) |
| sFit-1 | 1 | 1 | Van der Leeuw 2016 | 3p MACE, HF, PAD | SMART cohort: change in c-statistic 0.00 (-0.01 to 0.01) on top of base model of 0.70 (0.67-0.74);<br><br>EPIC-NL cohort: change in c-statistic 0.01 (-0.01 to 0.02) on top of base model of 0.69 (0.64-0.74) |

| | | | | | |
|---|---|---|---|---|---|
| VEGF | 1 | 0 | Van der Leeuw 2016 | 3p MACE, HF, PAD | SMART cohort: change in c-statistic 0.00 (-0.01 to 0.01) on top of base model of 0.70 (0.67-0.74);<br><br>EPIC-NL cohort: change in c-statistic 0.00 (0.00 to 0.01) on top of base model of 0.69 (0.64-0.74) |
| Osteocalcin | 1 | 1 | Van der Leeuw 2016 | 3p MACE, HF, PAD | SMART cohort: change in c-statistic 0.00 (-0.01 to 0.01) on top of base model of 0.70 (0.67-0.74);<br><br>EPIC-NL cohort: change in c-statistic 0.00 (-0.01 to 0.01) on top of base model of 0.69 (0.64-0.74) |
| Osteonectin | 1 | 1 | Van der Leeuw 2016 | 3p MACE, HF, PAD | SMART cohort: change in c-statistic 0.00 (0.00 to 0.01) on top of base model of 0.70 (0.67-0.74);<br><br>EPIC-NL cohort: change in c-statistic 0.01 (0.00 to 0.03) on top of base model of 0.69 (0.64-0.74) |
| Osteopontin | 1 | 1 | Van der Leeuw 2016 | 3p MACE, HF, PAD | SMART cohort: change in c-statistic 0.01 (0.00 to 0.03) on top of base model of 0.70 (0.67-0.74);<br><br>EPIC-NL cohort: change in c-statistic 0.01 (-0.01 to 0.02) on top of base model of 0.69 (0.64-0.74) |
| H-FABP | 1 | 0 | Van der Leeuw 2016 | 3p MACE, HF, PAD | SMART cohort: change in c-statistic 0.01 (0.00 to 0.02) on top of base model of 0.70 (0.67-0.74);<br><br>EPIC-NL cohort: change in c-statistic 0.00 (-0.01 to 0.01) on top of base model of 0.69 (0.64-0.74) |
| E-FABP | 1 | 1 | Van der Leeuw 2016 | 3p MACE, HF, PAD | SMART cohort: change in c-statistic 0.00 (0.00 to 0.01) on top of base model of 0.70 (0.67-0.74);<br><br>EPIC-NL cohort: change in c-statistic 0.00 (-0.02 to 0.01) on top of base model of 0.69 (0.64-0.74) |
| TIMP-1 | 1 | 1 | Van der Leeuw 2016 | 3p MACE, HF, PAD | SMART cohort: change in c-statistic 0.01 (0.00 to 0.02) on top of base model of 0.70 (0.67-0.74);<br><br>EPIC-NL cohort: change in c-statistic 0.02 (0.00 to 0.03) on top of base model of 0.69 (0.64-0.74) |

| | | | | | |
|---|---|---|---|---|---|
| IGF- binding protein (IGFBP)-1 | 1 | 1 | Wallander 2007 | 3p MACE | CV death: HR 1.4 (1.1-1.8), *p* = 0.003;<br>CV event: HR 1.2 (1.0-1.4), *p* = 0.096 |
| Abnormal MPI | 1 | 1 | Sultan 2006 | CHD, CVM, HF, Angina | Abnormal MPI: OR 5.6 (1.7–18.5) |
| small dense LDL (sdLDL) | 2 | 2 | Sakai 2018 | 3p MACE, Revasc, HF | HR 1.210 (1.003–1.459) |
| | | | Sultan 2006 | CHD, CVM, HF, Angina | LDL cholesterol ≥ 3.35 mmol/l: OR 7.3 (1.5–34.7) |
| An abnormal electrocardiogram | 1 | 1 | Nelson 1990 | CHD | Abnormal electrocardiogram: IRR 3.3 (1.6-6.8) |
| Medial artery calcification (MAC) | 2 | 2 | Lehto 1996 | 3p MACE, PAD | CVD mortality: OR 1.6 (1.1, 2.2);<br>CHD: OR 1.5 (1.0, 2.2) |
| | | | Nelson 1990 | CHD | Medial arterial calcification: IRR 3.5 (1.2-9.9) |
| Levels of malondialdehyde (MDA) - LDL | 1 | 1 | Lopes-Virella 2012 | 3p MACE, HF, PAD | MI: HR=2.44 (1.03, 5.77);<br>Composite end point: HR= 1.71 (1.04, 2.80) |
| MCP-1 (Monocyte chemoattractant protein-1) | 1 | 0 | Lorenzo-Almoros 2020 | CHD, Stroke | HR per SD increase 0.78 (0.44–1.38) |

NR, not reported; 3p MACE, 3-point major adverse cardiovascular events; Revasc, revascularization; HF, heart failure; CHD, coronary heart disease; PAD, peripheral artery disease; CVM, cardiovascular mortality; ACM, all-cause mortality.

#### Supplemental Table 9. Degree of variation in measurement methods used for each of these biomarkers obtained prior to CVD events in the ambulatory non-acute setting

| Study ID | Biomarker | EF used as covariate (Yes/No) | Assay used for measuring biomarker |
|---|---|---|---|
| Colombo 2018 | NT-proBNP | No | Elecsys proBNP II assay from Roche diagnostics s (Burgess Hill, UK). |
| Bruno 2014 | NT-proBNP | No | Two-site sandwich electrochemiluminescence immunoassay (Elecsys proBNP II, Roche Diagnostic, Mannheim, Germany) |
| Bruno 2013 | NT-proBNP | No | Two-site sandwich electrochemiluminescence immunoassay (Elecsys proBNP II; Roche Diagnostics |
| McMurray 2011 | NT-proBNP | No | Roche proBNP II assay |
| Resl 2016 | NT-proBNP | No | Cobas H232 system, with intra-essay and inter-essay coefficient of variations (CVs) of <15% at 60–1200 pg/mL and <20% at 1200–9000 pg/mL |
| Savonitto 2018 | NT-proBNP | No | No information (but Roche was one of the industry partner). |
| Sharma 2020 | NT-proBNP | No | Roche Diagnostics, Indianapolis, IN |
| vonScholten 2015 | NT-proBNP | Yes | Sandwich immunoassay on an Elecsys 2010 (Roche Diagnostics, Basel, Switzerland) |
| Scirica 2016 | NT-proBNP | No | Sandwich immunoassay (proBNP II; Roche Diagnostics) |
| Lorenzo-Almorós 2020 | NT-proBNP | Yes | Immunoassay (VITROS, Ortho Clinical Diagnostics Raritan, city, New York, NJ, USA). |
| Rørth 2019 | NT-proBNP | Yes | Roche Elecsys NT-proBNP (Roche Diagnostics, Indianapolis, IN, USA |
| McMurray 2011 | TnT | | Roche fourthgeneration TnT assay |
| Lepojarvi 2016 | TnT | | MODULAR ANALYTICS, Roche Diagnostics |
| Keller 2018 | TnT | | Roche Diagnostics; performed on an Elecsys 2010 system |
| Everett 2015 | TnT | | highsensitivity electrochemiluminescence assays, Roche Diagnostics |
| Scirica 2016 | TnT | | Electrochemiluminescent immunoassay assay (Roche Diagnostics) |
| Resl 2016 | TnT | | Elecsys Assay by Roche Diagnostics (Rotkreuz, Switzerland) |
| Lorenzo-Almoros 2020 | TnT | | **No information provided** |
| Rørth 2019 | TnT | | Roche Diagnostics GmbH, Mannheim, Germany |
| Lee 2017 | CCTA | | 64-slice multidetector computed tomography (MDCT) scanner (Light Speed VCT 64; GE Healthcare, Milwaukee, WI) or a dual-source computed tomography (DSCT) scanner (Somatom Definition; Siemens Healthcare, Forchheim, Germany). |
| Halon 2019 | CCTA | | 64-slice scanner (Brilliance CT, Philips Healthcare, Cleveland, Ohio) using a spiral, retrospective, electrocardiograph-gated protocol. |
| Halon 2016 | CCTA | | 64-channel scanner (Brilliance CT; Philips Healthcare, Cleveland, OH) using a spiral, retrospective, ECG-gated protocol. |
| Tian 2019 | CCTA | | Sensation 64 Slice CT scanner (Siemens AG, Munich, Germany). |
| Qi 2011 | Genetic risk score studies | | OpenArray™ SNP Genotyping System and TaqMan assays implemented on an ABI PRISM 7700 HT Sequence Detection System |
| Cox 2014 | Genetic risk score studies | | Affymetrix (SantaClara,CA) Genome Wide Human SNP Array 5.0 (GWAS) and Illumina (SanDiego,CA) Infinium Human Exome Bead Chip version 1.0 (Exome) |

| | | | |
|---|---|---|---|
| The Look AHEAD Research Group 2015 | Genetic risk score studies | | MetaboChip (Illumina,SanDiego,CA,USA) |
| Morieri 2018 | Genetic risk score studies | | Illumina Human Omni Express Exome 8v1.0 chips, Affymetrix Axiom Biobank1 chips; and Human Core Exome Bead Chip 12v1.0 and v1.1 (Illumina) |

**Supplemental Table 10. Summary of the results on the association of 79 genetic biomarkers and CVD outcomes.**

| Locus gene / closest gene | Author (Year) | Genetic marker | Risk Allele | Population ancestry | Population Characteristics | Outcome | Results | Validation | Improve Prediction (C-stat, NRI, IDI) |
|---|---|---|---|---|---|---|---|---|---|
| GRS for CAD (up to 160 loci) | Qi 2011 | GRS (5 SNP-based) | per higher GRS unit | non-Hispanic Whites | No ASCVD | CHD | OR 1.19 (1.13-1.26) | Internal validation | AUC increase 0.013 (0.008-0.018); *p* <0.001; NRI 0.28 (0.19-0.37) |
| | Cox 2014 | GRS-CAD (30 SNP-based) | per higher SD | European American | Enr. high CV risk | CVM | HR 1.46 (1.08-1.96) | none | Non-significant increase in AUC; NRI 0.06, *p* = 0.09 |
| | Mccaffery 2015 | GRS-CAD (153 SNP-based) | per higher SD | Multinational | Any CVD risk | 3p MACE, Angina | HR 1.17 (1.09-1.27) | none | NR. But similar results for primary prevention (HR 1.17 [1.06-1.29]) |
| | Morieri 2018 | GRS (204 SNP-based at 160 loci) | per higher SD | Non-Hispanic Whites | Enr. high CV risk | CHD | HR 1.17 (1.08-1.27) | external validation | AUC increase +0.007, *p* = 0.04; rIDI +8%, *p* = 0.0007; NRI 0.16, *p* <0.0001 |
| | | | | Non-Hispanic Whites | Enr. high CV risk | CHD | HR 1.32 (1.13-1.55) | external validation | |
| | | | | African-Americans | Enr. high CV risk | CHD | HR 0.95 (0.79-1.14) | external validation | |
| | | | | Hispanics | Enr. high CV risk | CHD | HR 1.20 (0.92-1.57) | external validation | |
| | | | | Asians | Enr. high CV risk | CHD | HR 1.98 (1.31-2.99) | external validation | |

| | | | | | | | | | |
|---|---|---|---|---|---|---|---|---|---|
| *APOB* | Bernard 2004 | XbA1 | X allele | White | no ASCVD | 3p MACE, Angina | X+X-vs. X−X-: OR 0.316 (0.12-0.87), p = 0.025;<br><br>X+X+ vs. X−X-: OR 0.043 (0.004-0.43), p = 0.008 | none | NR |
| *APOE* | Winkler 2010 | ε4 | ε4 | White | Dialysis patients | 3p MACE | HR 1.299 (1.045-1.615) | none | AUC increase by ε4 allele 0.005, *p* = 0.013 |
| | | | | | | CVM | HR 1.362 (1.002-1.852), *p* = 0.048 | none | NR |
| *CETP* | Porchay-Baldérelli 2007 | CETP TaqIB Polymorphism | B1 | White | CKD | CHD | HR 1.35 (1.01-1.79), *p* = 0.043 | none | NR |
| *MMP9* | Watson C 2021 | rs3918242 | T allele | Irish / Caucasian | Any CVD risk | CHD | OR 2.02, (1.13-3.63) | None | NR |
| *NOS3* | Odeberg 2008 | Asp 298 polymorphism | Asp298Asp | White | Any CVD risk | CHD | HR 3.12 (1.49-6.56), *p* = 0.003 | none | NR |
| | Kuricová 2013 | 894G>T (rs1799983) | T allele (additive) | White | DKD | 3p MACE, Revasc, PAD | HR 2.52 (1.06-5.97), *p* = 0.036 | none | NR |
| | Katakami 2014 | G894T | T allele | Japanese | No ASCVD | CHD | HR 1.05 (0.60-1.84), *p* = 0.861 | none | NR |
| *LPA* | Qi 2012 | rs10455872 | G allele | White | Any CVD risk | 3p MACE | CHD: HR 0.94 (0.69-1.28), *p* = 0.71 | none | NR |
| | | rs783147 | G allele | White | Any CVD risk | 3p MACE | CHD: HR 1.13 (0.98-1.29), *p* = 0.09 | none | NR |
| | | rs6935921 | T allele | White | Any CVD risk | 3p MACE | CHD: HR 0.91 (0.78-1.06), *p* = 0.22 | none | NR |
| | | rs2048327 | C allele | White | Any CVD risk | 3p MACE | CHD: HR 0.92 (0.80-1.07), *p* = 0.29 | none | NR |

| | | | | | | | | | |
|---|---|---|---|---|---|---|---|---|---|
| | | rs6919346 | C allele | White | Any CVD risk | 3p MACE | CHD: HR 1.01 (0.85-1.21), *p* = 0.89 | none | NR |
| | | rs12214416 | T allele | White | Any CVD risk | 3p MACE | CHD: HR 0.88 (0.62-1.25), *p* = 0.47 | none | NR |
| *LPA* genotype score | Qi 2012 | 5 SNP Genetic Score | higher score | White | Any CVD risk | 3p MACE | CHD: HR 0.98 (0.91-1.04), *p* = 0.48 | none | NR |
| *ADIPOQ* | Ho 2012 | rs1063539 | G allele | Chinese | No ASCVD | CHD | HR 0.78 (0.57-1.07) *p* = 0.12 | none | NR |
| | Ortega Moreno 2016 | rs822354 | A | White | Est. ASCVD | CVM | IRR = 1.94 (1.23-3.07), *p* = 0.005 | none | NR |
| *HNF4A* | Ho 2012 | rs1884614 | T | Chinese | No ASCVD | CHD | HR 1.09 (0.81-1.47) | none | NR |
| *FTO* | Doney 2009 | rs9939609 | A allele | white | any CVD risk | CHD | HR 2.01 (1.18-3.45) | none | NR |
| *PPARG* | Satirapoj 2019 | Pro12Ala (rs1801282) | Pro12Ala vs Pro12Pro | Thai population | Any CVD risk | CHD | HR 1.58 (0.50-5.02) | none | NR |
| | Ho 2012 | Pro12Ala (rs1801282) | Pro12Pro vs 12Ala | Chinese | No ASCVD | CHD | HR 4.38 (1.03-18.57) | none | NR |
| *TCFL7* | Satirapoj 2019 | rs7903146 | T allele | Thai population | Any CVD risk | CHD | HR 1.04 (0.98-1.11) | none | NR |
| *GLUL* | Qi 2013 | rs10911021 | C allele | White | No ASCVD (female) | CHD | OR 1.36 (1.09-1.70) | Validated in cross-sectional design. | NR |
| | | | | | No ASCVD (male) | CHD | OR 1.50 (1.20-1.89) | Validated in cross-sectional design. | NR |
| | Huggins 2016 | rs10911021 | C allele | Multinational | Any CVD risk (from Look Ahead n=3845) | 3p MACE, Angina | HR 1.11 (0.99-1.25), *p* = 0.07 for additive model; HR 1.21 (1.03-1.42) for recessive model | none | NR |

| | | | | | | | | | |
|---|---|---|---|---|---|---|---|---|---|
| | | | | | No ASCVD (subgroup from Look Ahead n=3245) | 3p MACE, Angina | HR 1.17 (1.01-1.36), *p* = 0.032 for additive model;<br><br>HR 1.32 (1.08-1.61), *p* = 0.006 for recessive model | none | NR |
| | Baeney 2016 | rs10911021 | C allelle | White / European | No ASCVD | CHD | OR 1.25 (0.94-1.67) | none | NR |
| *ACE* | Hong Huang 1998 | (I/D) polymorphism | D allele | Finland / Caucasians | Any CVD Risk | CHD | *P* = 0.028 | Internal | NR |
| | Keavney 1995 | (I/D) polymorphism | D allele (recessive) | British Caucasians | newly diagnosed T2D | CHD | OR 1.35 (0.83-2.17) | none | NR |
| | Hadjadj 2008 | (I/D) polymorphism | D allele (dominant) | Caucasian | High CVD risk | 3p MACE, ACM, Renal | OR 1.12 (0.70-1.79) | none | No |
| | Wang 2005 | (I/D) polymorphism | D allele | Chinese | Any CVD risk | 3p MACE, Revasc, PAD | HR 1.27 (0.77-2.08) | none | NR |
| *HP* | Levy 2002 | Haptoglobin phenotype | 2 allele | American Indians | Any CVD risk | CHD | 2-1 vs. 1-1 genotypes OR 1.63 (0.74-3.63), *p* = 0.228;<br><br>2-2 vs. 1-1 genotypes OR 4.96 (1.85-3.33), *p* = 0.002;<br><br>2-2 vs. 2-1 genotypes OR 3.04 (1.30-7.09), *p* = 0.010 | none | NR |
| *MTHFR* | Russo 2011 | C677T polymorphism | T allele (dominant) | White | Any CVD risk | CHD, Stroke, Revasc, PAD | OR 0.52 (0.18-1.47), compared TT genotype with other genotypes | none | NR |
| *DDAH1* | Lu 2011 | rs233112, rs1498373, rs1498374, rs587843, rs1403956, s1241321 | rs1241321 AA (recessive model) | Chinese | Suspected CAD | 3p MACE | rs1241321: HR 0.31 (0.11-0.90), *p* = 0.03 | none | NR |
| *KIF6* | Hoffman 2011 | Trp719Arg genotypes (rs20455) | Arg719 (co-dominant) | White | Dialysis patients | 3p MACE, ACM | HR 0.92 (0.77-1.09) | none | NR |

| | | | | | | | | | |
|---|---|---|---|---|---|---|---|---|---|
| *GSTM1* | Doney 2005a | 15-kb deletion | GSTM1-null | White | Any CVD risk | 3p MACE | HR 1.08 (0.81-1.46) | none | NR |
| *GSTP1* | Doney 2005a | Ile105Val | Val105 | White | Any CVD risk | 3p MACE | HR 1.04 (0.70-1.57) | none | NR |
| *GSTT1* | Doney 2005a | GSTT1 null | GSTT1 null | White | Any CVD risk | 3p MACE | HR 1.47 (1.07-2.02); *p* = 0.017 | none | NR |
| *CNDP1* | Alkhalaf 2015 | 5-leucine repeat (5L5L) | 5L-5L | 99% were Caucasian | Any CVD risk | CVM | HR 1.12 (0.79-1.58) | none | NR |
| *MTNR1B* | Tan 2020 | rs10830963 | G allele | White | Any CVD risk | CHD | HR 1.19 (1.02, 1.40), *p* = 0.03 | none | NR |
| *MGP* | Roumeliotis 2017 | MGP T-138C polymorphism | TT vs TC/CC genotype | Greek Caucasian origin | Enr. high CV risk | CVM, Renal | HR 5.07 (1.07-24.09) *p* = 0.04 | none | NR |
| *B1AR* | Poon 2010 | Arg389Gly | Gly | Chinese | T2D with DKD | 3p MACE, Revasc, Angina, PAD, Renal | RR 0.92, *p* = 0.45 | none | NR |
| *ENPP1* | Bacci 2011 | K121Q (rs1044498) | KK vs Q121 | White | Overall Est. ASCVD (T2D from GHS *n* = 330) | 3p MACE | HR 1.47 (0.80-2.70) | External validation in non-diabetic and diabetic in cross-sectional studies | NR |
| | | | | | Obese AND Est. ASCVD or ESRD (T2D from GHS, TVAS, CREED study *n* = 177) | 3p MACE | HR 5.94 (1.88-18.78), *p* = 0.002) | External validation in non-diabetic and diabetic in cross-sectional studies | NR |
| | | | | | Non-Obese AND Est. ASCVD or ESRD (T2D from GHS, TVAS, CREED study *n* = 218) | 3p MACE | HR 0.62 (0.32-1.24), *p* = 0.18 | External validation in non-diabetic and diabetic in cross-sectional studies | NR |
| *PPARA* | *Doney 2005* | C2528G variant | C2528 | Caucasian (*n* = 1810 from Go-DARTS) | Any CVD risk | CHD | HR 2.77 (1.34-5.75), *p* = 0.006 | None | NR |
| | | | | | | CHD, ACM | HR 1.52 (0.99-2.31), *p* = 0.052 | None | NR |
| | | L162V variant | V162 | | Any CVD risk | CHD | HR 0.31 (0.10-0.93), *p* = 0.037 | None | NR |

| | | | | | | | | | |
|---|---|---|---|---|---|---|---|---|---|
| | | | | Caucasian (*n* = 1810 from Go-DARTS) | | CHD, ACM | HR 0.52 (0.28-0.98), *p* = 0.044 | None | NR |
| *SCYA11* | Wang 2010 | Ala23Thr | Ala⁄Ala | Chinese | No ASCVD | CHD | HR 1.70 (1.10-2.61), *p* = 0.016 for recessive model | none | NR |
| *PON2* | Wang 2010 | Ser311Cys | Cys⁄Cys or Cys⁄ Ser | Chinese | No ASCVD | CHD | HR 1.42 (1.08-1.88), *p* = 0.013 for additive model | none | NR |
| *PON* | *Heijmans 2000* | Met-55/Leu | Leu (recessive) | NR (born in Netherlands) | Elderly | CVM | RR 1.9 (0.5-6.6) | none | NR |
| | | Gln-192/Arg | Arg (recessive) | NR (born in Netherlands) | Elderly | CVM | RR 1.8 (0.4-8.5) | none | NR |
| *ADRB3* | *Wang 2010* | Trp64Arg | Arg⁄Arg | Chinese | No ASCVD | CHD | HR 3.84 (1.18-12.50), *p* = 0.025 for recessive model | none | NR |
| | | Ala23Thr, Ser311Cys, Trp64Arg | higher no. of risk allele | Chinese | No ASCVD | CHD | 2 vs. 0-1 risk alleles: HR 1.99 (1.087-3.656), *p* = 0.026;<br><br>3 vs. 0-1 risk alleles: HR 2.74 (1.424-5.258), *p* = 0.003;<br><br>4 vs. 0-1 risk alleles: HR 4.11 (1.650-10.230), *p* = 0.002 | none | NR |
| *CFH* | Valoti 2019 | p.Glu936Asp (c.2808G>T) | Asp/Asp homozygotes, recessive model | NR (most likely white/european) | High CVD risk | 3p MACE, Revasc, HF, Angina | HR 2.68 (1.23-5.87) | none | NR |
| *SOD1* | Neves AL 2012 | rs9974610 | G allele | European and North African | DKD with micro / macroalbuminuria | 3p MACE | MI or stroke:<br>HR= 1.06 (0.80-1.39);<br><br>CVD death: | None | NR |

| | | | | | | | | | |
|---|---|---|---|---|---|---|---|---|---|
| | | | | | | | HR 0.64 (0.46-0.88) | | |
| | | rs2173962 | G allele | European and North African | DKD with micro / macroalbuminuria | 3p MACE | MI or stroke: HR 0.87 (0.51-1.38); CVD death: HR 1.80 (1.06-2.90) | None | NR |
| | | rs10432782 | G allele | European and North African | DKD with micro / macroalbuminuria | 3p MACE | MI or stroke: HR 0.99 (0.71-1.35); CVD death: HR 1.71 (1.16-2.48) | None | NR |
| | | rs2070424 | G allele | European and North African | DKD with micro / macroalbuminuria | 3p MACE | MI or stroke: HR 1.07 (0.73-1.53); CVD death: HR 1.38 (0.85-2.12) | None | NR |
| | | rs1041740 | C allele | European and North African | DKD with micro / macroalbuminuria | 3p MACE | MI or stroke: HR 0.92 (0.59-1.37); CVD death: HR 1.78 (1.10-2.78) | None | NR |
| | | rs17880135 | G allele | European and North African | DKD with micro / macroalbuminuria | 3p MACE | MI or stroke: HR 1.07 (0.69-1.58); CVD death: HR 0.85 (0.51-1.35) | None | NR |
| | | rs202449 | A allele | European and North African | DKD with micro / macroalbuminuria | 3p MACE | MI or stroke: HR 1.06 (0.80-1.40); CVD death: HR 1.25 (0.89-1.72) | None | NR |
| *SOD2* | Katakami 2014 | Val16Ala | Val allele | Japanese | No ASCVD | CHD | CHD: HR 1.01 (0.62-1.63), *p* = 0.981; CVD: HR 1.11 (0.80-1.54), *p* = 0.534 | none | NR |

| | | | | | | | | | |
|---|---|---|---|---|---|---|---|---|---|
| *SOD3* | Mohammedi 2015 | rs2284659 | G allele | French | persistent microalbuminuria or macroalbuminuria without renal failure at baseline | CHD, CVM | MI: HR 0.75 (0.59-0.94), *p* = 0.01;<br><br>CV death: HR 0.83 (0.69-0.99), *p* = 0,03 for T-allele in a dominant model | none | NR |
| | | rs17552548 | A allele | French | persistent microalbuminuria or macroalbuminuria without renal failure at baseline | CHD, CVM | MI: HR 0.59 (0.29-0.98), *p* = 0.04 for G-allele in a dominant model | none | NR |
| | | rs758946 | T allele | French | persistent microalbuminuria or macroalbuminuria without renal failure at baseline | CHD, CVM | MI: HR 0.73 (0.45-1.07), *p* = 0.11 for C-allele in dominant model | none | NR |
| *GCLM* | Katakami 2014 | C-588T | T allele | Japanese | No ASCVD | CHD | CHD: HR 1.36 (0.96-1.94), *p* = 0.086;<br><br>CVD: HR 1.09 (0.85-1.40), *p* = 0.506 | none | NR |
| *CYBA* | Katakami 2014 | C242T | C allele | Japanese | No ASCVD | CHD | CHD: HR 1.27 (0.73-2.24), *p* = 0.401;<br><br>CVD: HR 1.06 (0.75-1.49), *p* = 0.753 | none | NR |
| *MPO* | Katakami 2014 | G-463A | G allele | Japanese | No ASCVD | CHD | CHD: HR 1.15 (0.68-1.95), *p* = 0.605;<br><br>CVD: HR 0.93 (0.68-1.27), *p* = 0.644 | none | NR |

| | | | | | | | | | |
|---|---|---|---|---|---|---|---|---|---|
| Pro-oxidant gene risk score | Katakami 2014 | Val16Ala, C-588T, G894T, C242T, G-463A | ≥8 pro-oxidant alleles | Japanese | No ASCVD | CHD | CHD: HR 2.83 (1.45-5.50), *p* = 0.002;<br><br>CVD: HR 1.78 (1.05-3.02), *p* = 0.034 | none | NR |
| *ALDH2* | He 2021 | rs671 | GA/AA genotype (dominant) | NR (study from China) | no ASCVD | 3p MACE | HR 1.31 (1.00-1.73) | none | NR |
| *ALOX12* | Roumeliotis 2018 | rs14309 | G allele | Greek Caucasian | CVD risk | 3p MACE, PAD | CV mortality:<br>HR 3.533 (1.08-11.55);<br><br>CV events:<br>HR 2.226 (1.049-4.723) | None | NR |
| *HSPA1B* | PYK Poon 2014 | A+1267G | G allele | Chinese | ESRD | CHD, HF | No significant difference in the 5-year event-free survival (*p* = 0.06) | None | NR |
| *VDR* | Ferrarezi 2013 | rs1544410 | A allele | Multinational | CVD risk | CHD, Revasc | HR 1.16 (1.05-1.29) | Results were validated in an independent cross-sectional cohort | NR |
| | | rs7975232 | A allele | Multinational | CVD risk | CHD, Revasc | HR 1.08 (0.97-1.12) | Results were validated in an independent cross-sectional cohort | NR |
| | | rs731236 | C allele | Multinational | CVD risk | CHD, Revasc | HR 1.10 (0.99-1.21) | Results were validated in an independent cross-sectional cohort | NR |

| | | | | | | | | | |
|---|---|---|---|---|---|---|---|---|---|
| *ABCA1* | Porchay-Baldérelli 2009 | (+)69C>T | Recessive (TT vs C+) | Multinational | DKD | CHD | OR 0.28 (0.13-0.61) | None | NR |
| | | R219K | Dominant (K+ vs RR) | Multinational | DKD | CHD | OR 0.80 (0.65-0.98) | None | NR |
| | | R1587K | Dominant (K+ vs RR) | Multinational | DKD | CHD | OR 1.22 (1.00-1.49) | None | NR |
| *LIPC* | Zhang 2005 | −514 C to T | T allele | Caucasian | No CVD risk | CHD | OR 1.21, (0.52-2.83) | None | NR |
| *APM1* | Lu Qi 2005 | G276T | G allele | Caucasian | No CVD risk | 3p MACE, Revasc | OR 0.38 (0.18-0.79) | None | NR |
| *RANTES* | Boger 2005 | G-403A (rs2107538) | -403AA (dominant) | Caucasian | Emodialisis | CVM | HR 1.99, $p = 0.05$ | none | NR |
| | | In1.1T/C (rs2280789) | In1.1C | Caucasian | Emodialisis | CVM | HR 2.0, $p = 0.05$ | none | NR |
| *ALR2* | So 2008 | 5'-(CA) microsatellite | z+2/z+2 | Chinese | No ASCVD | 3p MACE | HR 1.06 (0.65-1.72) | none | NR |
| | | C-106T | CT/TT | Chinese | No ASCVD | 3p MACE | HR 1.44 (0.89-2.34) | none | NR |

NR, not reported; 3p MACE, 3-point major adverse cardiovascular events; Revasc, revascularization; HF, heart failure; CHD, coronary heart disease; CVD, cardiovascular disease; PAD, peripheral artery disease; CVM, cardiovascular mortality; ACM, all-cause mortality; HR, hazard ratio; OR, odds ratio.

#### Supplemental Table 11. Summary of results from risk scores studies on internal and external validation.

| Information for risk scores | | | | Internal Validation | | | | External Validation | |
|---|---|---|---|---|---|---|---|---|---|
| **Name** | **Derivation Population** | **No. of predictors in the model** | **Predictors in the model** | **Author (Year)** | **Outcome** | **Method** | **Results (c-statistic; H-L p-value; O/E ratio; Other Results)** | **Cohort** | **Results (c-statistic; H-L p-value; O/E ratio; Other Results)** |
| ACC/AHA Pooled Cohort Equation | 37,128 patients from ARIC, CHS, CARDIA, FHS (US) | 9 | age, sex, race (White, Black, or other), smoking status, SBP, hypertension treatment status, diabetes status, TC and HDL-C levels | Basu 2017 | ~5-yr fatal or non-fatal MI, stroke | N/A | N/A | LOOK AHEAD (US) | 0.66 (0.64-0.69), $p$ <0.001 |
| AD-ON | 11,140 patients from ADVANCE-ON RCT (Europe/Asia) | 13 | age, sex, SBP with and without use of antihypertensives, duration of diabetes, HbHbA1c, urinary ACR, eGFR and its square, age at completion of formal education, exercise, history of diabetic retinopathy and current or previous AF | Woodward 2016 | 10-yr fatal or non-fatal macrovascular disease | Not specified | 0.662 (0.645-0.679);<br><br>IDI 0.0168 (0.0134-0.0201);<br><br>NRI 0.0068 (-0.0311 to 0.0451) compared to original ADVANCE | N/A | N/A |
| ADVANCE | 11,140 Age >55 with history of major macro/microvascular disease or 2+ risk factors (Europe/Asia) | 10 | age at diagnosis, sex, known duration of diabetes, pulse pressure, retinopathy, AF, HbHbA1c, urinary ACR, non-HDL-C, treated hypertension | Kengne 2011 | 4-yr fatal or nonfatal MI, stroke, CVM | bootstrapping | 0.702 (0.676-0.728), $p = 0.76$ | DIABHYCAR (multinational): 1,836 patients | 0.685 (0.646-0.724), $p = 0.032$;<br><br>0.82 (0.71-0.95) |
| | | | | van der Leeuw 2015 | 5-yr CHD | N/A | N/A | EPIC-NL (Netherlands) | 0.62 (0.54-0.70) |
| | | | | | | | | EPIC-Potsdam (Germany) | 0.67 (0.59-0.75) |

| | | | | | | | | | |
|---|---|---|---|---|---|---|---|---|---|
| | | | | | | | | SMART (Netherlands) | 0.68 (0.58-0.77) |
| | | | | Woodward 2016 | 10-yr fatal, non-fatal macrovascular disease | N/A | 0.655 (0.638-0.672) | N/A | N/A |
| | | | | Read 2018 | 5-yr MI, stroke, unstable angina, TIA, PAD, and coronary, carotid, or major amputation procedures | N/A | N/A | National Scottish Diabetes Register: 181,399 people with T2D an no history of CVD (Scotland) | 0.666 (0.661-0.671) |
| | | | | Wan 2018 | 5-yr CHD, MI, CVM, sudden death, HF and fatal and non-fatal stroke | N/A | N/A | 137,935 patients with T2D aged 18-79 years without history of CVD (China) | Males: 0.688 (0.676-0.700);<br><br>Females: 0.710 (0.698-0.722) |
| AMD-QUASAR | 5,181 patients (Italy) | 7 | HbA1c, blood pressure measurement, lipid profile measurement, LDL-C, MA measurement, not treated with ACE inhibitors/ARBs despite the presence of MA, treated with ACE inhibitors/ARBs in the presence of MA or MA absent | Rossi 2011 | 2.3 yr angina, MIA stroke, TIA, coronary revasc. procedures, lower-limb com-plications, and CVM | N/A | 84% higher CVD risk in patients with a score of <15 (IRR 1.84 [1.29-2.62]) and 17% higher in those with a score between 15 and 25 (IRR 1.17 [0.93-1.49]) compared with those with a score of >25 | N/A | N/A |

| | | | | | | | | | |
|---|---|---|---|---|---|---|---|---|---|
| ARIC | 14,054 (1,500 with diabetes) | 19 | age, race, TC, HDL-C, SBP, use of antihypertensive medication, smoking status, BMI, waist-to-hip ratio, lipoprotein(a), albumin, creatinine, WBC, fibrinogen, factor VIII, sport activity, residual FEV1, Keys score, and pack-years smoking | Folsom 2003 | 10-yr MI, CHD death, unrecognized MI by ECG, coronary revasc. | Split-sample | Women: 0.771;<br><br>Men: 0.740 | N/A | N/A |
| BASCORE | 777 patients newly diagnosed with T2D older than 24 years (Spain) | 6 | age, sex, non-HDL to HDL-C, SBP, HbA1c, tobacco | Pinies 2014 | 10-yr fatal or non-fatal MI, angina, silent MI based on ECG, stroke, PAD | Not specified | 0.63 (0.57-0.69), *p* <0.01 | N/A | N/A |
| BRAVO | ACCORD: 10,251 participants aged 40-79 years with high ASCVD risk | 11 | HbA1c, LDL, age at diagnosis, severe hypoglycaemia, sex, education, smoking, race, history of MI, history of CHF, history of stroke | Shao 2018 | 3.7-yr MI | Not specified | 0.689 (0.662-0.704) | ASPEN, ADVANCE, and CARDS trials (Multinational) | Calibration slopes provided in figures |
| | | | | Shao 2020 | ~3 yr nonfatal or fatal MI, MACE, CVM | N/A | N/A | EMPA-REG, CANVAS, DECLARE-TIMI 58 trials (Multinational) | O/E results shown in figures for various outcomes but data not provided;<br><br>BRAVO risk engine was effective in predicting benefit of SGLT-2 is on CV health through improvements in commonly measured risk factors (HbA1c, SBP, and BMI) |

| | | | | | | | | | |
|---|---|---|---|---|---|---|---|---|---|
| Camridgeshire UK | 4704 patients with T2D from 18 general practices in Cambridgeshire (UK) | 9 | age, sex, HbA1c, BMI, SBP, DBP, TC, HDL-C, LDL-C | Yu 2018 | 2-yr CV hospitalization | bootstrapping | 0.7094 (0.7067-0.7205);<br><br>NR; 1.030 (0.9856-1.0747) | RCT of Peer Support in T2D trial: 1,121 patients (UK) | 0.7092 (0.7033-0.7151);<br><br>NR; 1.0001 (0.9807-1.0195) |
| CHS | 782 older adults (65+) with diabetes free of ASCVD (US) | 10 | age, former smoker, current smoker, SBP, TC, HDL-C, creatinine >110.5 mmol/L, oral hypoglycaemic agent or insulin, CRP, ABI <1, ECG LVH, carotid IMT per mm up to 3 | Mukamal 2013 | 7-yr MI, stroke, CVM | Not specified | 0.68, $p = 0.65$ | MESA: 843 adults with T2D (US) | 0.68;<br><br>NRI on internal validation 0.12;<br><br>NRI on external validation 0.09 |
| | | | | van der Leeuw 2015 | 5-yr CHD | N/A | N/A | EPIC-NL: 536 pts with T2D (Netherlands) | 0.54 (0.46-0.63) |
| | | | | | | | | EPIC-Potsdam: 1,332 patients (Germany) and 1,685 in SMART (Netherlands) | 0.61 (0.52-0.70) |
| | | | | | | | | SMART: 1,685 with T2D (Netherlands) | 0.68 (0.59-0.78) |
| | | | | Read 2018 | 5-yr MI, stroke, unstable angina, TIA, PAD, and coronary, carotid, or major amputation procedures | Not specified | N/A | National Scottish Diabetes Register: 181,399 people with T2D an no history of CVD (Scotland) | 0.674 (0.669-0.679) |

| | | | | | | | | | |
|---|---|---|---|---|---|---|---|---|---|
| Cleveland Clinic | 33,067 patients with T2D prescribed single oral hypoglycemic agent at Cleveland Clinic between 1998-2006 (United States) | 23 | age, gender, race, creatinine, LDL, HDL, TG, weight, height, HbA1c, history of heart failure, smoking status, SBP, DBP, insulin, clopidogrel, aspirin, cholesterol med, new diabetic, household income, history of heart disease, warfarin, ACE inhibitor or ARB | Wells 2013 | 5-yr CHD | 10-fold cross-validation | 0.7298 | N/A | N/A |
| CVD-DCCT | 1441 DCCT patients (T1D) (United States) | 10 | HbA1c, marital status, albumin-urine value, age, presence of neuropathy, occupation, weight, smoking status, patient attempted suicide, SBP | Lagani 2015 | 5.75-yr CVM, MI, bypass graft / angioplasty, angina, cardiac arrhythmia, major ECG abnormality, silent MI, CHF, TIA, arterial event requiring surgery. | cross-validation | 0.6757 (0.5439-0.9268), *p* = 0.003 | Chorleywood Medical Center: 344 patients (UK) | 0.7143 (0.6238-0.8056), *p* <0.001 |
| CVD-EDIC | 1394 subjects out of the original 1441 DCCT patients (T1D) (United States) | 5 | HbA1c, age, total insulin daily dosage, smoking status, creatinine clearance | Lagani 2015 | 5.75-yr CVM, MI, bypass graft / angioplasty, angina, cardiac arrhythmia, major ECG abnormality, silent MI, CHF, TIA, arterial event requiring surgery. | cross-validation | 0.6621 (0.5435-0.8387), *p* = 0.0002 | Chorleywood Medical Center: 344 patients (UK) | 0.6099 (0.5021-0.7181), *p* = 0.0165 |

| | | | | | | | | | |
|---|---|---|---|---|---|---|---|---|---|
| DART | 4,569 patients with T2D (Scotland) | 9 | duration of diabetes, age at diagnosis, TC, smoking status, sex, HbA1c, interaction of HbA1c and follow-up, SBP, treated hypertension | Donnan 2006 | 5-yr fatal and non-fatal CHD, CVM | N/A | N/A | Salford Diabetes Information System (DIS) (UK) | 0.71, *p* = 0.63; 0.79 |
| EXSCEL Risk Score | 14,752 patients with T2D with and without CVD enrolled in exenatide once-weekly RCT | 18 | age, sex, region, smoking status, previous CV event, previous MI, diabetes duration, previous revascularization, cerebrovascular disease, NYHA class, chronic respiratory disease, AF, BMI, HbA1c, eGFR, SBP, DBP, pulse pressure | Mentz 2018 | 3.2 yr MACE | N/A | HR for MACE by quintile of risk score:<br><br>1.074 (0.704-1.639) for 1st quintile;<br><br>0.868 (0.649-1.162) for 2nd quintile;<br><br>0.918 (0.723-1.166) for 3rd quintile;<br><br>0.778 (0.637-0.949) for 4th quintile;<br><br>0.942 (0.816-1.086) for 5th quintile | N/A | N/A |
| FDS | 1240 with all required risk factor data were followed from baseline (1993-1996) (Australia) | 8 | age, sex, prior CVD, urinary ACR, HbA1c, HDL-C, Southern European ethnic background, Aboriginality | Davis 2010 | 5-yr MI, stroke, CVM | N/A | N/A | Busselton Health Study: 180 patients (Australia) | 0.84, *p* = 0.85 |
| | | | | van der Leeuw 2015 | 5-yr CHD | N/A | N/A | EPIC-NL (Netherlands) | 0.58 (0.50-0.66) |
| | | | | | | | | EPIC-Potsdam (Germany) | 0.68 (0.60-0.76) |

| | | | | | | | | | |
|---|---|---|---|---|---|---|---|---|---|
| | | | | | | | | SMART (Netherlands) | 0.69 (0.59-0.79) |
| | | | | Read 2018 | 5-yr MI, stroke, unstable angina, TIA, PAD, and coronary, carotid, or major amputation procedures | N/A | N/A | National Scottish Diabetes Register: 181,399 people with T2D an no history of CVD (Scotland) | 0.670 (0.665-0.674) |
| FDS-2 | 1,551 patients with T2D | 13 | age, sex, Australian aboriginal, heart rate, diabetes duration, HbA1c, urinary ACR, eGFR <45, PAD, LVH, heart failure, CHD and/or cerebrovascular disease | Davis 2020 | 5-yr 4p MACE | Not specified | 0.82 (0.79-0.85), *p* = 0.17 | Busselton Diabetes Study: 188 adults (Australia) | 0.81 (0.74-0.89), *p* = 0.16 |
| FRS | 5,345 patients age 30-74 years in Framingham Heart Study in 1971-1974 (United States) | 6 | age, sex, TC, HDL-C, SBP, smoking status | Guzder 2005 | 10-yr CVD | N/A | N/A | Poole Diabetes Study: 428 patients age 30-74 years with newly diagnosed T2D free of CVD (UK) | 0.673, *p* <0.001 |
| | | | | Davis 2009 | 5-yr CHD | N/A | N/A | FDS: 815 patients free of CVD (Australia) | 0.59 (0.54-0.64), *p* <0.001 |
| | | | | vanderJeijden 2009 | 10-yr fatal and nonfatal CHD | N/A | N/A | Hoorn Study: 255 patients age 50-75 free of CVD (Netherlands) | 0.63 (0.50-0.76) |

| | | | | | | | | | |
|---|---|---|---|---|---|---|---|---|---|
| | | | | Elley 2010 | 5-yr fatal and nonfatal CVD | N/A | N/A | NZ DCS: 12,626 patients from the South (New Zealand) | 0.63 (0.62-0.65) |
| | | | | Kengne 2010 | 4-yr coronary revasc. and hospitalisation for unstable angina; stroke and TIA; total CVD events (including any of the above, CHF and PAD) | N/A | N/A | ADVANCE: 11,140 Age >55 with history of major macro/microvascular disease or 2+ risk factors (Europe/Asia) | 0.618, *p* <0.0001; 1.46 (1.32-1.61) |
| FRS | 5,345 patients age 30-74 years in Framingham Heart Study in 1971-1974 (United States) | 6 | age, sex, TC, HDL-C, SBP, smoking status | Kengne 2011 | 4-yr fatal or nonfatal MI, stroke, CVM | N/A | N/A | DIABHYCAR (multinational): 1,836 patients | 0.646 (0.603-0.689) |
| | | | | Wells 2013 | 10-yr CHD | N/A | N/A | Cleveland Clinic: 7,714 patients age 30-74 years with T2D prescribed single oral hypoglycemic agent at Cleveland Clinic between 1998-2006 (United States) | 0.54 |

| | | | | | | | | | |
|---|---|---|---|---|---|---|---|---|---|
| | | | | Yeboah 2014 | 8.5-yr fatal or non-fatal MI | N/A | N/A | MESA-HNR: 1343 patients without CVD (US /Germany) | 0.7;<br><br>improved risk classification versus the FRS (NRI 0.19 and IDI 0.046, *p* <0.05) and UKPDS (NRI 0.215 and IDI 0.046, *p* <0.05)<br><br>Compared with the ATP III guidelines, the MESA-HNR score has an NRI of 0.74 for the main outcome. |
| | | | | Wan 2018 | 5-yr CHD, MI, CVM and sudden death, heart failure and fatal and non-fatal stroke | N/A | N/A | 137,935 patients with T2D aged 18-79 years without history of CVD (China) | Males: 0.666 (0.654-0.677);<br><br>Females: 0.668 (0.656-0.680) |
| | | | | Simmons 2009 | 10.1-yr fatal or nonfatal MI, sudden cardiac death, CHD, fatal or nonfatal stroke, and PAD death | N/A | N/A | EPIC-Norfolk: 10,137 patients age 40-79 years from General practices from 1993-1998 (UK) | 0.73 (0.66-0.78) |
| FRS + Carotid IMT | 5,345 patients age 30-74 years in Framingham Heart Study in 1971-1974 (United States) | 7 | age, sex, TC, HDL-C, SBP, smoking status, carotid IMT | Yoshida 2012 | 5-yr CVM, nonfatal MI, unstable angina, stable angina, TIA, and ischemic stroke | N/A | N/A | Juntendo University Hospital: 783 patients with T2D (Japan) | 0.656, *p* <0.001 |

| | | | | | | | | | |
|---|---|---|---|---|---|---|---|---|---|
| FRS + combined vascular calcified plaque score | 5,345 patients age 30-74 years in Framingham Heart Study in 1971-1974 (United States) | 10 | age, sex, TC, HDL-C, smoking status, SBP, anti-hypertensive medication use, coronary artery calcified plaque, and abdominal aortic calcified plaque | Cox 2014 | 8.4-yr 3p MACE | N/A | N/A | DHS: 1,120 European Americans (US) | 0.7709;<br><br>C-statistic of 0.7314 (*p* = 0.0133) for FRS alone |
| FRS-RS | Adults age 35-74 years from Girona, Spain (population study to calibrate FRS) | 8 | age, sex, TC, HDL-C, SBP, treatment for hypertension, smoking status, diabetes status | Pinies 2014 | 5-yr fatal and non-fatal MI, angina, silent MI based on ECG, stroke, PAD. | N/A | N/A | BASCORE: 777 patients newly diagnosed with T2D older than 24 years (Spain) | 0.58 (0.51-0.65), *p* <0.0001 |
| HKDR | 7,067 patients without CHD (China) | 7 | age, sex, smoking status, diabetes duration, eGFR, urinary ACR, non-HDL-C | Yang 2008 | 5-yr MI, CHD | Split-sample | 0.704, *p* = 0.675; 0.733 | N/A | N/A |
| HKU-SG | 678,750 participants aged 20 or over from Hong Kong and 386,425 from Singapore | 16 | age, duration of diabetes, sex, smoking status, HbA1c, total and HDL-C, blood pressure, creatinine, hemoglobin, WBC count, resting heart rate, preexisting conditions (cancer, CHF, diabetes complications), medication for diabetes, hypertension and statins | Quan 2019 | 5-yr CHD | bootstrap ping | 0.700 (0.698-0.702) | N/A | N/A |

| | | | | | | | | | |
|---|---|---|---|---|---|---|---|---|---|
| Hong Kong Hospital Authority | 137,935 patients with T2D aged 18-79 years without history of CVD (China) | 13 | age, sex, BMI, waist circumference, SBP, DBP, TC, triglycerides, HDL-C, LDL-C, eGFR, HbA1c, duration of diabetes | Wan 2018 | 5-yr CHD, MI, coronary death and sudden death, CHF and fatal and non-fatal stroke | split-sample | Males: 0.705 (0.693-0.716);<br><br>Females: 0.719 (0.707-0.731) | N/A | N/A |
| Improved FRS (I-FRS) | 3,232 paients in Beijing Communities Diabetes Study (China) | 9 | age, gender, TC, HDL-C, SBP, treatment for high blood pressure, smoking status, diabetes status, history of CVD | Zhang 2020 | 10-yr CVD | | C-statistic: *p* <0.0001;<br><br>14.49% for high, 10.51% for medium 2.46% for low I-FRS, p <0.001 | N/A | N/A |
| JADE | 7534 T2D patients (Asia-Pacific) | 7 | age, diabetes duration, gender, smoking, non-HDL-C, eGFR, urinary ACR | Quan 2019 | 5-yr CHD | N/A | N/A | Hong Kong dataset: 678,750 participants (China) | 0.685 (0.683-0.688) |
| JJ Risk Engine | 1,748 patients without diabetes complications other than mild diabetic retinopathy (Japan) | 11 | age, sex, non-HDL-C, HbA1c, urinary ACR, SBP, BMI, AF, duration of diabetes, smoking, leisure time physical activity | Tanaka 2013 | 7.2-yr HD, stroke, non-CVD mortality, nephropathy | cross-validation | 0.725 (0.656-0.793);<br><br>0.14;<br><br>1.08 | N/A | N/A |
| LHDB | 4,274 adults with T2D with medical center lab data (Taiwan) | 9 | HbA1c measurement frequency, hypertension, SBP >130 or DBP >80, use of anti-hypertensives, lipid measurement frequency, LDL ≥100, eye measurement frequency, foot exam frequency, | Li 2018 | 8-yr macrovascular disease (stroke, MI, heart failure, PAD) | | Compared to score of 1, risk was 64% lower in those with quality- of-care scores ≥5 (adjusted HR 0.36 [0.28-0.45]) | N/A | N/A |

| | | | | | | | | | |
|---|---|---|---|---|---|---|---|---|---|
| | | | urine exam frequency | | | | | | |
| MESA-HNR | 1343 patients without CVD (United States/Germany) | 7 | FRS + coronary artery calcium | Yeboah 2014 | 8.5-yr outcome | cross-validation | 0.7575;<br><br>NRI of 0.190 (SE 0.076) when compared to FRS (i.e. without inclusion of CAC);<br><br>NRI of 0.215 (SE 0.074) when compared to UKPDS | N/A | N/A |
| MRFC | 35,196 age 40-79 years (UK Clinical Practice Research Data Link) (7,048 with CKD) | 4 | HbA1c <7.0%, blood pressure <140/90mm Hg, TC <5mmol/L and no smoking | Hamada 2018 | 6-yr all-cause mortality, CV mortality, fatal and non-fatal CHD, stroke | Not specified | Participants meeting four criteria also had lower relative hazards for CHD (adjusted sub-distribution HR 0.73 [0.59 to 0.91]) and stroke (0.63 [0.45 to 0.89]), considering death as a competing risk. | N/A | N/A |
| NDR | 11,646 patients aged 18 -70 years (Sweden) | 8 | onset age of diabetes, diabetes duration, sex, BMI, smoking, SBP, and | Cederholm 2008 | 8-yr fatal, non-fatal CHD | Not specified | 0.7; NR; 0.96 | N/A | N/A |
| | | | | | 5 yr CHD | N/A | N/A | EPIC-NL | 0.64 (0.56-0.72) |

| | | | | | | | | | |
|---|---|---|---|---|---|---|---|---|---|
| | | | antihypertensive and lipid-reducing drugs | van der Leeuw 2015 | 5 yr CHD | N/A | N/A | EPIC-Potsdam | 0.67 (0.59-0.74) |
| | | | | | 5 yr CHD | N/A | N/A | SMART | 0.64 (0.54-0.74) |
| | 24,288 patients age 30-74, 15% with previous CVD (Sweden) | 12 | onset-age, diabetes duration, TC:HDL-C ratio, HbA1c, SBP, BMI, sex, smoking, microalbuminuria, macroalbuminuria, AF, previous CVD | Zethelius 2011 | 11-yr fatal and non-fatal CHD | | 0.71; 0.9; 1 | UKPDS: 4906 age range 30-74 years, baseline in 2003 and separate from the derivation dataset, with follow-up for 4 years until 2007 United Kingdom) | 0.72; 0.2; 0.97 |
| | | | | Read 2018 | 5-yr MI, stroke, unstable angina, TIA, PAD, and coronary, carotid, or major amputation procedures | N/A | N/A | National Scottish Diabetes Register: 181,399 people with T2D an no history of CVD (Scotland) | 0.663 (0.658-0.668) |
| NDR-BP II | 30,179 adults age 35-74 years who were free of CHD and stroke (Sweden) | 14 | SBP, sex, age, diabetes duration, type of hypoglycaemic treatment, HbA1c, LDL-C, HDL-C, TG, BMI, smoker, albuminuria, AF, history of CVD | Wan 2018 | 5-yr CHD, MI, coronary death and sudden death, heart failure and fatal and non-fatal stroke | N/A | N/A | 137,935 patients with T2D aged 18-79 years without history of CVD (China) | Males: 0.693 (0.682-0.705);<br><br>Females: 0.708 (0.696-0.720) |
| NZ DCS | 36,127 people without CVD from North New Zealand | 9 | age at diagnosis, diabetes duration, sex, SBP, smoking status, TC-to-HDL ratio, ethnicity, HbA1c, and urine ACR | Elley 2010 | 5-yr fatal and nonfatal CVD | N/A | N/A | NZ DCS: 12,626 patients from the South of New Zealand | 0.67 (0.67-0.70) |
| | | | | van der Leeuw 2015 | 5-yr CHD | N/A | N/A | EPIC-NL | 0.63 (0.55-0.71) |
| | | | | | | | | EPIC-Potsdam | 0.66 (0.59-0.74) |

| | | | | | | | | | |
|---|---|---|---|---|---|---|---|---|---|
| | | | | | | | | SMART | 0.67 (0.57-0.77) |
| | | | | Wan 2018 | 5-yr CHD, MI, coronary death and sudden death, heart failure and fatal and non-fatal stroke | N/A | N/A | 137,935 patients with T2D aged 18-79 years without history of CVD (China) | Males: 0.687 (0.676-0.699);<br><br>Females: 0.709 (0.697-0.721) |
| | | | | Read 2018 | 5-yr MI, stroke, unstable angina, TIA, PAD, and coronary, carotid, or major amputation procedures | N/A | N/A | National Scottish Diabetes Register: 181,399 people with T2D an no history of CVD (Scotland) | 0.665 (0.660-0.670) |
| OPTUM | 181,619 patients with T2D ≥50 years in Optum Real-World Evidence Electronic Health Records and Claims Database | 16 | age, sex, race, ethnicity, study period, geographic region, insurance type, payer type, other CVD-related conditions, at least 1 diabetes-related hospitalization, adapted diabetes complications severity index, chronic pulmonary disease, cancer, fluid/electrolyte disorders, coagulopathy, time interval | Young 2018 | 1-yr MACE | Split-sample | 0.72 | N/A | N/A |

| | | | | | | | | | |
|---|---|---|---|---|---|---|---|---|---|
| QRISK2 (PMID 18573856) | 2.3 million patients age 35-74 from prospective open cohort study from general practice (England and Wales) | 14 | age, sex, diabetes status, ethnicity, BMI, TC:HDL-C ratio, SBP, AF, smoking, treated hypertension, Townsend social deprivation score, rheumatoid arthritis, family history of CVD | Read 2018 | 5-yr MI, stroke, unstable angina, TIA, PAD, and coronary, carotid, or major amputation procedures | N/A | N/A | National Scottish Diabetes Register: 181,399 people with T2D an no history of CVD (Scotland) | 0.675 (0.669-0.679) |
| Ramírez-Prado | 112 adults with T2D admitted via ED for any cause except acute MI, stroke, cancer, or palliative status (Spain) | 6 | age, sex, insulin, admitted for heart failure, history of renal failure, hypertension | Ramírez-Prado 2015 | 4-yr CVD | Not specified | 0.734 (SE 0.049) | N/A | N/A |
| RECODe | ACCORD: 10,251 participants aged 40-79 years with high ASCVD risk | 14 | age, sex, ethnicity, smoking, SBP, history of CVD, blood pressure lowering drugs, statins, anticoagulants, HbA1c, TC, HDL-C, creatinine, urine ACR | Basu 2017 | 4.7-yr fatal or non-fatal MI, stroke | Cross-validation | 0.69 (0.67-0.71), *p* = 0.14 | LOOK AHEAD | 0.73 (0.71-0.75), *p* <0.0001 |
| | | | | Basu 2018 | 8-yr fatal or non-fatal MI, stroke | N/A | N/A | MESA: 1,555 adults with T2D | 0.74, *p* = 0.005 |
| | | | | | | | | Jackson Heart Study: 1.746 adults with T2D | 0.77, *p* <0.001 |
| SCORE (PMID 12788299) | 250,000 patients aged 40-65 years (Europe) | 5 | age, sex, SBP, smoking, cholesterol | vanderJeijden 2009 | 10-yr fatal and nonfatal CHD | N/A | N/A | Hoorn Study: 255 patients age 50-75 free of CVD (Netherlands) | 0.66 (0.54-0.79) |

| | | | | | | | | | |
|---|---|---|---|---|---|---|---|---|---|
| SPRINT | 8,760 non-diabetic patients with increased CVD risk in SPRINT RCT (United States) | 7 | age, creatinine, albuminuria, anti-hypertensives, number of anti-hypertensives, history of CVD, smoking | Venäläinen 2020 | 4.7-yr MI, ACS, stroke, heart failure, CVM | split-sample | 0.7 (0.66-0.74) | ACCORD: 4,733 patients with T2D | 0.69 (0.67-0.72) |
| THIN | all T2D patients aged at least 25 years with at least 5 years of data in THIN database,contains anonymized longitudinal health care records, from 570 primary care practices, 11.7 million patients (United Kingdom) | 15 | age, female, smoking, HbA1c, SBP, TC:HDL-C ratio, MI, PAD, AF, CHF, CHD, Stroke, Blindness, Renal failure, Amputation | McEwan 2015 | 5-yr CHD | | 0.617 | N/A | N/A |
| UKPDS | 5,102 patients with newly diagnosed T2D (United Kingdom) | 8 | age, sex, ethnicity, smoking status, SBP, TC, HDL-C, presence of AF | Guzder 2005 | 10-yr CVD | N/A | N/A | Poole Diabetes Study: 428 patients age 30-74 years with newly diagnosed T2D free of CVD (UK) | 0.67, *p* = 0.029 |
| | | | | Yang 2008 | 5-yr MI, CHD | N/A | N/A | HKDR: 7,067 patients without CHD (China) | 0.61, *p* = 0.581; 0.639 |

| | | | | | | | | | |
|---|---|---|---|---|---|---|---|---|---|
| UKPDS | 5,102 patients with newly diagnosed T2D (United Kingdom) | 8 | age, sex, ethnicity, smoking status, SBP, TC, HDL-C, presence of AF | Davis 2009 | 5-yr CHD | N/A | N/A | FDS: 815 patients free of CVD (Australia) | 0.68 (0.59-0.76), *p* <0.001 |
| | | | | vanderJeijden 2009 | 10-yr fatal and nonfatal CHD | N/A | N/A | Hoorn Study: 255 patients age 50-75 free of CVD (Netherlands) | 0.66 (0.56-0.78) |
| | | | | Kengne 2010 | 4-yr coronary revasc. and hospitalisation for unstable angina; stroke and TIA; total CVD events (including any of the above, CHF and PAD) | N/A | N/A | ADVANCE: 11,140 Age >55 with history of major macro/microvascular disease or 2+ risk factors (Europe/Asia) | 0.655, *p* <0.001;<br><br>1.76 (1.60-1.94) |
| | | | | van Dieren 2011 | 5-yr MI, CHD, stroke | N/A | N/A | EPIC-NL (Netherlands) and EPIC-Potsdam (Germany) cohorts: 1,622 individuals with T2D free of ASCVD | 0.66 (0.51-0.81), *p* <0.001 |
| | | | | Tanaka 2013 | 7.2-yr HD, stroke, non-CVD mortality, nephropathy | N/A | N/A | JDCS/J-EDIT: 1,748 patients without diabetes complications other than mild diabetic retinopathy (Japan) | 0.695 (0.626-0.764), *p* <0.01;<br><br>0.3 |

| | | | | | | | | | |
|---|---|---|---|---|---|---|---|---|---|
| UKPDS | 5,102 patients with newly diagnosed T2D (United Kingdom) | 8 | age, sex, ethnicity, smoking status, SBP, TC, HDL-C, presence of AF | Yang 2013 | 3.5-yr 3p MACE | N/A | N/A | ACCORD: 10,251 age 40-79 with CVD or 55-79 with 2+ CV risk factors (North America) | NR; NR; 1.7 |
| | | | | | 4.3-yr 3p MACE | N/A | N/A | ADVANCE: 11,140 Age >55 with history of major macro / microvascular disease or 2+ risk factors (Europe/Asia) | NR; NR; 1.8 |
| | | | | | 5.6-yr 3p MACE | N/A | N/A | VADT: 1,791 age >40 with no CV events in past 6 months (U.S.) | NR; NR; 1.3 |
| | | | | | 5-yr 3p MACE | N/A | N/A | RECORD: 4,437 age 40-75 with no major CV events in last 3 months (Europe/Australia) | NR; NR; 1.5 |
| | | | | | 3-yr 3p MACE | N/A | N/A | PROactive: Age 35-75 with history of CVD (Europe) | NR; NR; 0.9 |
| | | | | | 4-yr 3p MACE | N/A | N/A | ADOPT: 4,360 age 30-75 without unstable or severe angina, CHF, or uncontrolled hypertension (North America/Europe) | NR; NR; 2 |
| | | | | | 5.3-yr 3p MACE | N/A | N/A | BARI 2D: 2,368 age >25 with diagnosis of CHD | NR; NR; 1.2 |

| | | | | | | | | | |
|---|---|---|---|---|---|---|---|---|---|
| | | | | | | | | (America/Europe) | |
| UKPDS | 5,102 patients with newly diagnosed T2D (United Kingdom) | 8 | age, sex, ethnicity, smoking status, SBP, TC, HDL-C, presence of AF | Pinies 2014 | 10-yr fatal or non-fatal MI, angina, silent MI based on ECG, stroke, PAD | N/A | 0.62 (0.57-0.68), *p* <0.001 | N/A | N/A |
| | | | | Yeboah 2014 | 8.5-yr fatal or non-fatal MI | N/A | N/A | MESA-HNR: 1343 patients without CVD (US /Germany) | 0.69 |
| UKPDS (+CVD history) | 5,102 patients with newly diagnosed T2D age 25-65 years between 1977-1991 (United Kingdom) | 9 | age, sex, ethnicity, smoking status, SBP, TC, HDL-C, presence of AF, CVD history | Yang 2013 | 3.5-yr 3p MACE | N/A | N/A | ACCORD: 10,251 age 40-79 with CVD or 55-79 with 2+ CV risk factors (North America) | NR; NR: 2.3 |
| | | | | | 4.3 yr 3p MACE | N/A | N/A | ADVANCE: 11,140 Age >55 with history of major macro / microvascular disease or 2+ risk factors (Europe/Asia) | NR; NR: 2.4 |
| | | | | | 5.6 yr 3p MACE | N/A | N/A | VADT: 1,791 age >40 with no CV events in past 6 months (U.S.) | NR; NR; 1.9 |

| | | | | | | | | | |
|---|---|---|---|---|---|---|---|---|---|
| | | | | | 5 yr 3p MACE | N/A | N/A | RECORD: 4,437 age 40-75 with no major CV events in last 3 months (Europe/Australia) | NR; NR; 1.8 |
| UKPDS (+CVD history) | 5,102 patients with newly diagnosed T2D age 25-65 years between 1977-1991 (United Kingdom) | 9 | age, sex, ethnicity, smoking status, SBP, TC, HDL-C, presence of AF, CVD history | Yang 2013 | 3 yr 3p MACE | N/A | N/A | PROactive: Age 35-75 with history of CVD (Europe) | NR; NR; 1.8 |
| | | | | | 4 yr 3p MACE | N/A | N/A | ADOPT: 4,360 age 30-75 without unstable or severe angina, CHF, or uncontrolled hypertension (North America/Europe) | NR; NR; 2 |
| | | | | | 5.3 yr 3p MACE | N/A | N/A | BARI 2D: 2,368 age >25 with diagnosis of CHD (America/Europe) | NR; NR; 2.4 |
| UKPDS (v3) | 5,102 patients with newly diagnosed T2D (United Kingdom) | 9 | age, duration of diabetes, sex, ethnicity, smoking, HbA1c, SBP, TC, HDL-C | Simmons 2009 | 10.1-yr fatal or nonfatal MI, sudden cardiac death, other incident CHD, fatal or nonfatal stroke, and PAD death | N/A | N/A | EPIC-Norfolk: 10,137 patients age 40-79 years from General practices from 1993-1998 (UK) | 0.72 (0.65-0.78) |
| UKPDS-56 | 4,540 patients with newly diagnosed T2D (United Kingdom) | 8 | age at diabetes diagnosis, sex, ethnicity, smoking, HbA1c, SBP, lipid ratio, duration of diabetes | Stevens 2001 | 10-yr fatal and non-fatal CHD | Not specified | coefficients reported only for each predictor variable but no overall model performance results provided | N/A | N/A |

| | | | | | | | | | |
|---|---|---|---|---|---|---|---|---|---|
| UKPDS-60 | 3,740 patients with newly diagnosed T2D who were recruited from 23 centers between 1977 and 1991 (UK) | 8 | age, duration, sex, smoking, BMI, SBP, TC:HDL-C ratio, AF | Quan 2019 | 5-yr CHD | N/A | N/A | Hong Kong dataset: 678,750 participants (China) | 0.676 (0.673-0.678) |
| UKPDS-OM (UKPDS-68) PMID 15517152) | 3,867 patients with newly diagnosed T2D who were recruited from 33 centers (UK) | 5 | age, sex, HbA1c, SBP, TC:HDL-C ratio | McEwan 2015 | 5-yr CHD | N/A | N/A | THIN: all T2D patients aged at least 25 years with at least 5 years of data in THIN database, which contains anonymized longitudinal health care records, from 570 primary care practices, on 11.7 million patients (United Kingdom) | 0.567 |
| | 5,102 patients with newly diagnosed T2D (United Kingdom) | 5 | age, sex, HbA1c, SBP, TC:HDL-C ratio | Clarke 2004 | 10 yr fatal and non-fatal CHD | Split sample (Yr 1 vs. Years 2-12) | Simulation results for estimated life expectancy and quality adjusted life years reported | N/A | N/A |
| UKPDS OM2 (UKPDS-82) (PMID 23793713) | 3,867 patients with newly diagnosed T2D who were recruited from 33 centers (UK) | 9 | age, sex, SBP, TC, HDL-C, eGFR, CHF, PAD, amputation | Quan 2019 | 5-yr CHD | N/A | N/A | Hong Kong dataset: 678,750 participants (China) | 0.654 (0.651-0.657) |

| | | | | | | | | | |
|---|---|---|---|---|---|---|---|---|---|
| | 5,102 UKPDS patients from 20 year trial and 4,031 survivors entering 10-year post-trial monitoring period | 12 | age, sex, ethnicity, duration of diabetes, HbA1c, SBP, TC, HDL-C, smoking status, microvascular complications, macrovascular complications, diabetes medications | Basu 2017 | 4.7-yr fatal or non-fatal MI, stroke | N/A | N/A | LOOK AHEAD (US) | 0.67 (0.64-0.69), *p* <0.001 |
| | | | | Basu 2018 | 8-yr fatal or non-fatal MI, stroke | N/A | N/A | MESA: 1,555 adults with T2D | 0.6, *p* <0.001 |
| | | | | | | | | Jackson Heart Study: 1.746 adults with T2D | 0.61, *p* <0.001 |
| UKPDS OM2 (UKPDS-82) (PMID 23793713) | 5,102 UKPDS patients from 20 years trial and 4,031 survivors entering 10-year post-trial monitoring period | 9 | age, sex, SBP, TC, HDL-C, eGFR, CHF, PAD, amputation | McEwan 2015 | 5-yr CHD | N/A | N/A | THIN: all T2D patients aged at least 25 years with at least 5 years of data in THIN database, which contains anonymized longitudinal health care records, from 570 primary care practices, on 11.7 million patients (United Kingdom) | 0.582 |
| | 5,102 patients with newly diagnosed T2D (United Kingdom) | 10 | age, sex, ethnicity, smoking status, duration of diabetes, HbA1c, SBP, TC, HDL-C, bMP | Hayes 2013 | 25-yr MI, CHD | | Simulations reported for an input dataset of 5,102 patients; predictions stabilised at around 200 Monte Carlo replications. | N/A | N/A |

| | | | | | | | | | |
|---|---|---|---|---|---|---|---|---|---|
| Zhengzhou University | 2282 patients with diabetic neuropathy (China) | 21 | age at diabetes diagnosis, gender, BMI, SBP, HbA1c, TC, HDL-C, LDL-C, triglyceride levels, smoking status, urine ACR, eGFR, use of RAS blockers, serum AST levels, serum GGT levels, serum potassium, serum sodium, serum creatinine, hemoglobin levels, albumin level | Yu 2019 | 2-yr CVM | bootstrapping | 0.736 (0.726-0.746);<br><br>calibration slope 0.993 (0.990-0.997) | Zhengzhou University:950 patients with biopsy-proven diabetic neuropathy (China) | 0.747 (0.737-0.756);<br><br>Calibration slope 1.000 (0.981-1.020);<br><br>Compared with the UKPDS risk score, which underestimated the cardiovascular disease risk, the new score is a more specific tool for patients with T2D and DN |

N/A, not applicable; SBP, systolic blood pressure; HDL-C, high-density lipoprotein cholesterol; TC, total cholesterol; ACR, albumin:creatinine ratio; AF, atrial fibrillation; IRR, incidence rate ratio; TIA, transient ischaemic attack; PAD, peripheral artery disease; CVD, cardiovascular disease; CHD, coronary heart disease; MI, myocardial infarction; CVM, cardiovascular mortality; HF, heart failure; LDL-C, low-density lipoprotein cholesterol; BMI, body mass index; T2D, type 2 diabetes; CHF, congestive heart failure; WBC, white blood cell; CRP, c-reactive protein; ABI, ankle-brachial index; ECG LVH, electrocardiogram left ventricular hypertrophy; carotid IMT, carotid intima-media thickness; DBP, diastolic blood pressure; ACE, angiotensin-converting enzyme; ARB, angiotensin receptor blockers; T1D, type 1 diabetes; NYHA, New York Heart Association; revasc., revascularization; NRI, net reclassification improvement; IDI, integrated discrimination improvement; HR, hazard ratio; SE, standard error; eGFR, estimated glomerular filtration rate; RAS, renin-angiotensin system; AST, alanine aminotransferase; GGT, gamma-glutamyl transferase; DN, diabetic nephropathy.

**Supplemental Figure 1:** Variations in the definitions of cardiovascular outcomes among the included studies.

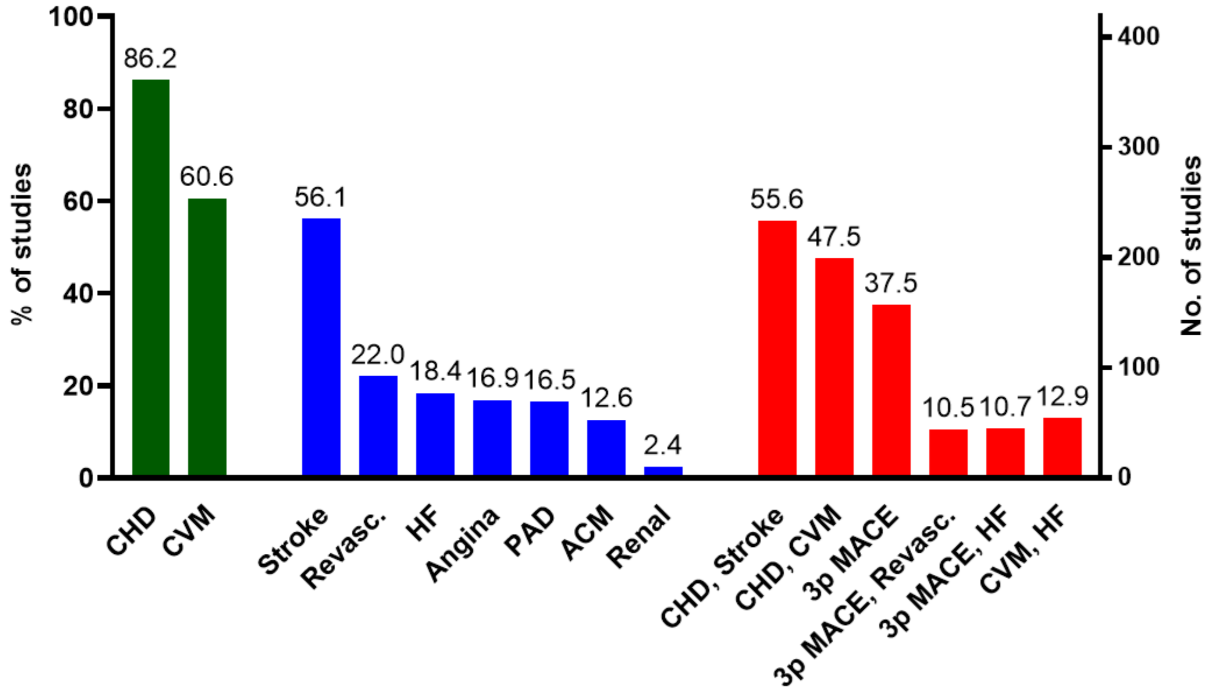

**Supplemental Figure 1** Caption: Green columns represent the "required" endpoints (Coronary Heart Disease [CHD] and/or cardiovascular mortality [CVM]). Blue columns represent additional endpoints (e.g. Stroke, Heart Failure [HF], peripheral artery disease [PAD] or all cause mortality [ACM]) being evaluated in the studies. Red columns represent the combination of specific outcomes in groups of general interest (e.g. 3-point Major Adverse Cardiovascular Events [MACE]). Note: End-points (and groups of end-points) are not mutually exclusive therefore each study can be counted multiple time in this graph (e.g. one studies can have both CHD and CVM; Or a study evaluating a composite outcome of CHD, CVM, Stroke and Revascularization can be included both in "3p MACE" and "3p MACE, Revasc." columns).

**Supplemental Figure 2.** Meta-analyses of studies N-terminal pro-B-type natriuretic peptide (NTpro-BNP) and Troponin T (TnT).

Supplemental Figure 2A. NT-proBNP (continuous- per logarithm increase)

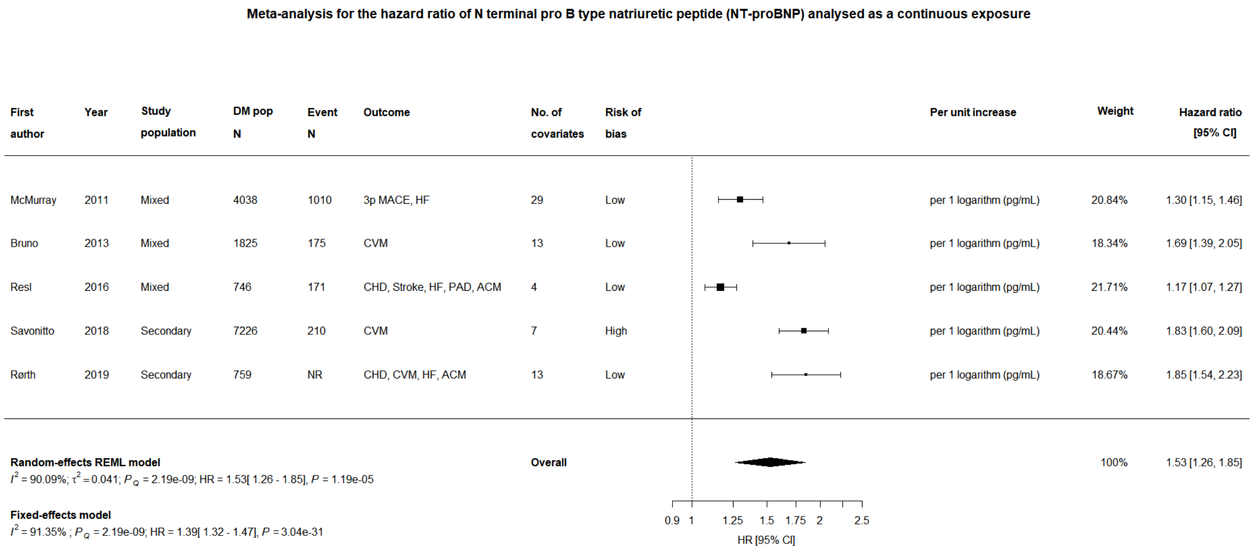

### Supplemental Figure 2B. NT-proBNP (continuous- per SD increase)

**Meta-analysis for the hazard ratio of N terminal pro B type natriuretic peptide (NT-proBNP) analysed as a continuous exposure**

| First author | Year | Study population | DM pop N | Event N | Outcome | No. of covariates | Risk of bias | Per unit increase | Weight | Hazard ratio [95% CI] |
|---|---|---|---|---|---|---|---|---|---|---|
| vonScholten | 2015 | Primary | 200 | 40 | 3p MACE, HF | 10 | Medium | per 1 SD of the logarithms (ng/L) | 14.06% | 1.70 [1.13, 2.56] |
| Colombo | 2018 | Primary | 2105 | 144 | CHD, Stroke | 16 | Low | per 1 SD (pg/mL) | 20.77% | 1.94 [1.54, 2.45] |
| Bruno | 2014 | Mixed | 1645 | 149 | CVM | 11 | Low | per 1 SD of the logarithms (pg/mL) | 22.46% | 1.84 [1.52, 2.22] |
| Scirica | 2016 | Mixed | 12310 | NR | 3p MACE, HF | 13 | Low | per 1 SD of the logarithms (pg/mL) | 26.01% | 1.62 [1.50, 1.74] |
| Lorenzo-Almoros | 2020 | Secondary | 232 | 56 | CHD, Stroke | 15 | Low | per 1 SD (pg/mL) | 16.70% | 0.95 [0.68, 1.33] |
| **Random-effects REML model** | | | | | | | **Overall** | | 100% | 1.59 [1.27, 1.99] |

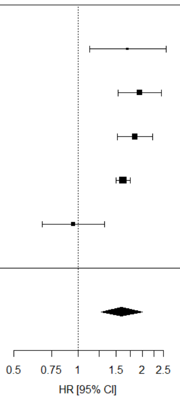

**Random-effects REML model**
$I^2$ = 82.6%; $\tau^2$ = 0.048; $P_Q$ = 0.00818; HR = 1.59[ 1.27 - 1.99], $P$ = 4.3e-05

**Fixed-effects model**
$I^2$ = 70.88% ; $P_Q$ = 0.00818; HR = 1.64[ 1.53 - 1.74], $P$ = 3.94e-51

#### Supplemental Figure 2C. TnT (continuous- per logarithm increase)

**Meta-analysis for the hazard ratio of troponin T (TnT) analysed as a continuous exposure**

| First author | Year | Study population | DM pop N | Event N | Outcome | No. of covariates | Risk of bias | Per unit increase | Weight | Hazard ratio [95% CI] |
|---|---|---|---|---|---|---|---|---|---|---|
| Resl | 2016 | Mixed | 746 | 171 | CHD, Stroke, HF, PAD, ACM | 10 | Low | per 1 logarithm (ng/L) | 57.00% | 1.43 [1.13, 1.81] |
| Rørth | 2019 | Secondary | 759 | NR | CHD, CVM, HF, ACM | 13 | Low | per 1 logarithm (ng/L) | 43.00% | 1.91 [1.45, 2.51] |
| **Random-effects REML model** | | | | | | **Overall** | | | 100% | 1.62 [1.35, 1.94] |

$I^2$ = 58.96%; $\tau^2$ = 0.025; $P_Q$ = 0.119; HR = 1.64[ 1.23 - 2.18], $P$ = 0.000627

**Fixed-effects model**

$I^2$ = 58.96% ; $P_Q$ = 0.119; HR = 1.62[ 1.35 - 1.94], $P$ = 1.5e-07

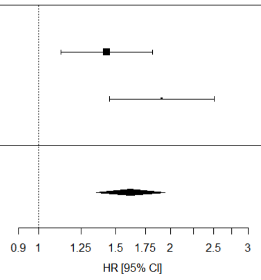

#### Supplemental Figure 2D.  TnT (binary/categorical)

**Meta-analysis for the hazard ratio of troponin T (TnT) analysed as a binary or categorical exposure**

| First author | Year | Study population | DM pop N | Event N | Outcome | No. of covariates | Risk of bias | Cutoff for biomarker | Weight | Hazard ratio [95% CI] |
|---|---|---|---|---|---|---|---|---|---|---|
| McMurray | 2011 | Mixed | 4038 | 1010 | 3p MACE, HF | 29 | Low | >0.028 ng/mL vs. ≤0.028 ng/mL | 26.53% | 1.50 [1.06, 2.13] |
| Keller | 2018 | Mixed | 1034 | 377 | 3p MACE | 17 | Medium | Q4 (>90 ng/L) vs Q1 (≤35 ng/L) | 26.57% | 2.10 [1.49, 2.95] |
| Everett | 2015 | Secondary | 1984 | 198 | 3p MACE, ACM | 18 | High | ≥14 ng/L vs. <14 ng/L | 27.03% | 1.56 [1.22, 1.99] |
| Lepojarvi | 2016 | Secondary | 1053 | 37 | CHD, CVM, HF | 11 | Low | >14 ng/L vs. ≤14 ng/L | 19.88% | 15.50 [5.12, 46.90] |
| **Overall** | | | | | | | | | 100% | 2.64 [1.03, 6.72] |

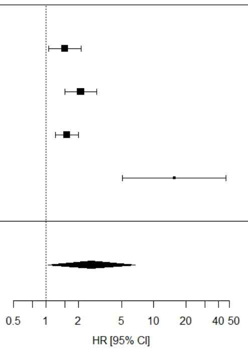

**Random-effects REML model**
$I^2$ = 95.9%; $\tau^2$ = 0.827; $P_Q$ = 0.000532; HR = 2.64[ 1.03 - 6.72], $P$ = 0.0422

**Fixed-effects model**
$I^2$ = 82.95% ; $P_Q$ = 0.000532; HR = 1.76[ 1.48 - 2.09], $P$ = 9.35e-11

#### Supplemental Figure 2E.  TnT (continuous)

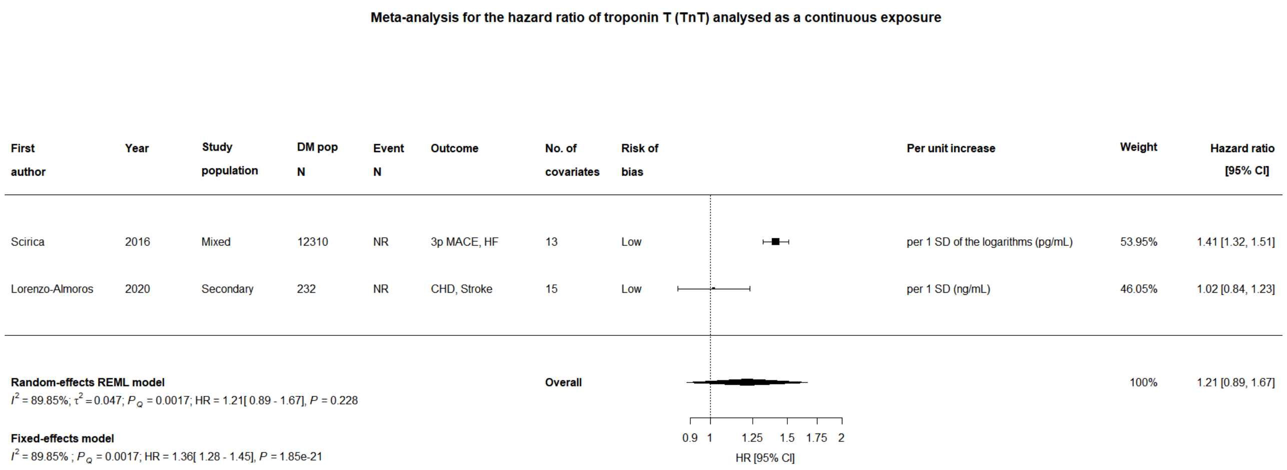

**Supplemental Figure 2** Caption: (A) NT-proBNP analyzed as a continuous exposure with a logarithm transformation, (B) NT-proBNP analyzed as a continuous exposure with a z-score transformation, (C) TnT analyzed as a binary or categorical exposure, (D) TnT analyzed as a continuous exposure with a logarithm transformation, (E) TnT analyzed as a continuous exposure with a z-score transformation.

**Supplemental Figure 3:** Forest plots for 9 biomarkers (CACS, carotid plaque, C-reactive protein (CRP), Galectin-3 (Gal-3), growth differentiation factor (GDF-15), pulse wave velocity (PWV), SPECT scintigraphy, Troponin I (TnI) and Triglyceride glucose index (TyG).

Supplemental Figure 3A. CACS

**Coronary artery calcium score (CACS)**

| First author | Year | Study population | DM pop N | Event N | Outcome | No. of covariates | Risk of bias | Per unit increase / Cutoff for biomarker | Hazard ratio [95% CI] |
|---|---|---|---|---|---|---|---|---|---|
| Lau | 2012 | Primary | 151 | 17 | 3p MACE, Revasc, PAD | 8 | High | >40 vs. ≤40 | 27.11 [ 3.48, 211.35] |
| Anand | 2006 | Primary | 510 | 20 | 3p MACE, Revasc | 7 | Low | >1000 vs. <100 | 58.05 [20.34, 165.65] |
| Elkeles | 2008 | Primary | 589 | 66 | 3p MACE, Angina | 13 | Low | per 1 logarithm to the base 2 | 1.29 [ 1.16, 1.44] |
| Dayan | 2012 | Primary | 128 | 23 | CHD, Stroke, Revasc, HF, PAD | 3 | High | per 1 logarithm | 6.82 [ 3.42, 13.59] |
| vonScholten | 2015 | Primary | 200 | 40 | 3p MACE, HF | 10 | Medium | per 1 SD of the logarithms | 3.40 [ 1.71, 6.75] |
| Lee | 2017 | Primary | 933 | 94 | CHD, Revasc, ACM | 7 | Medium | per 1 score | 1.21 [ 1.09, 1.34] |
| Venuraju | 2019 | Primary | 238 | 94 | CHD, Revasc, ACM | 5 | Low | per 1 logarithm to the base 10 (Agatston units) | 2.32 [ 1.12, 4.81] |

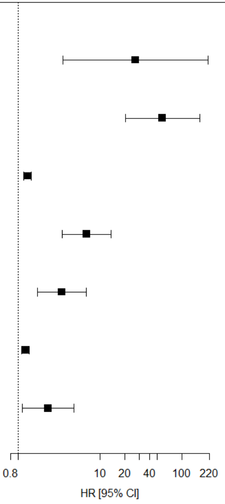

#### Supplemental Figure 3B. Carotid plaque

**Carotid plaque**

| First author | Year | Study population | DM pop N | Event N | Outcome | No. of covariates | Risk of bias | Per unit increase | Hazard ratio [95% CI] |
|---|---|---|---|---|---|---|---|---|---|
| Katakami | 2012 | Primary | 85 | 20 | 3p MACE, Angina | 7 | Medium | per 1 mm | 1.94 [1.17, 3.21] |
| Irie | 2013 | Primary | 287 | 34 | CHD, Stroke, Revasc, Angina, PAD | 6 | Medium | per 1 SD (mm) | 1.44 [1.01, 2.06] |

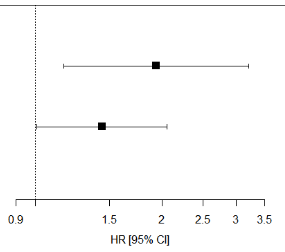

### Supplemental Figure 3C. CRP

**C-reactive protein (CRP)**

| First author | Year | Study population | DM pop N | Event N | Outcome | No. of covariates | Risk of bias | Per unit increase / Cutoff for biomarker | Hazard ratio [95% CI] |
|---|---|---|---|---|---|---|---|---|---|
| Schulze | 2004 | Primary | 746 | 103 | CHD, Stroke, Revasc | 13 | Low | Q4 vs. Q1 (mg/L) | 2.62 [1.29, 5.32] |
| vonScholten | 2016 | Primary | 200 | 40 | 3p MACE, HF | 10 | Medium | per 1 SD of the logarithms (mg/L) | 1.00 [0.71, 1.41] |
| Linnemann | 2006 | Mixed | 292 | NR | 3p MACE, PAD, ACM | 8 | Med. | Q4 (≥9.2 mg/L) vs. Q1 (≤2.7 mg/L) | 5.40 [1.45, 20.12] |
| McMurray | 2011 | Mixed | 4038 | 1010 | 3p MACE, HF | 29 | Low | ≥6.6 mg/L vs. ≤3.0 mg/L | 1.44 [1.23, 1.69] |
| Cardoso | 2016 | Mixed | 616 | 184 | 3p MACE, ACM | 17 | Low | per 1 SD of the logarithms (mg/L) | 1.30 [1.09, 1.55] |
| Scirica | 2016 | Mixed | 16492 | NR | 3p MACE, HF | 9 | Low | per 1 SD of the logarithms (mg/L) | 1.17 [1.10, 1.25] |
| Lepojarvi | 2016 | Secondary | 1053 | 37 | CHD, CVM, HF | 11 | Low | ≥1.6 mg/L vs. <1.6 mg/L | 4.60 [2.12, 9.99] |
| Lowe | 2014 | Secondary | 3865 | 709 | 3p MACE, Renal | 16 | Low | per 1 SD of the logarithms (mg/L) | 0.95 [0.85, 1.06] |
| Lorenzo-Almoros | 2020 | Secondary | 232 | 56 | CHD, Stroke | 15 | Low | per 1 mg/L | 0.97 [0.92, 1.02] |

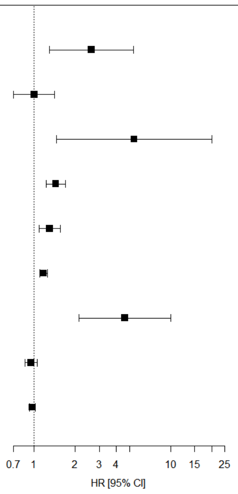

#### Supplemental Figure 3D. Gal-3

**Galectin-3**

| First author | Year | Study population | DM pop N | Event N | Outcome | No. of covariates | Risk of bias | Per unit increase / Cutoff for biomarker | Hazard ratio [95% CI] |
|---|---|---|---|---|---|---|---|---|---|
| Lepojarvi | 2016 | Secondary | 1053 | 37 | CHD, CVM, HF | 11 | Low | ≥12.2 ng/mL vs. <12.2 ng/mL | 3.90 [1.78, 8.52] |
| Lorenzo-Almoros | 2020 | Secondary | 232 | 56 | CHD, Stroke | 15 | Low | per 1 SD (ng/mL) | 1.83 [1.13, 2.97] |
| Sharma | 2020 | Secondary | 5154 | 837 | 3p MACE, HF | 10 | Medium | per 1 logarithm to the base 2 (ng/mL) | 1.21 [1.03, 1.42] |

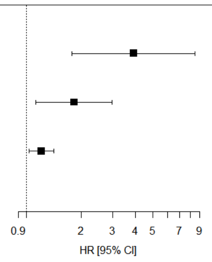

#### Supplemental Figure 3E. GDF-15

**Growth differentiation factor (GDF-15)**

| First author | Year | Study population | DM pop N | Event N | Outcome | No. of covariates | Risk of bias | Per unit increase | Hazard ratio [95% CI] |
|---|---|---|---|---|---|---|---|---|---|
| Resl | 2016 | Mixed | 746 | 171 | CHD, Stroke, HF, PAD, ACM | 10 | Low | per 1 logarithm (ng/L) | 1.50 [1.25, 1.80] |
| Carlsson | 2020 | Mixed | 813 | NR | CHD, Stroke | 9 | Low | per 1 SD of the logarithms to the base 2 | 1.34 [0.96, 1.88] |
| Sharma | 2020 | Secondary | 5154 | 837 | 3p MACE, HF | 10 | Medium | per 1 logarithms to the base 2 (pg/mL) | 1.15 [1.04, 1.28] |

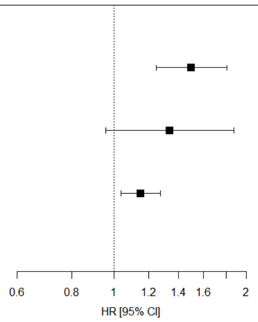

#### Supplemental Figure 3F. PWV

**Pulse wave velocity (PWV)**

| First author | Year | Study population | DM pop N | Event N | Outcome | No. of covariates | Risk of bias | Per unit increase / Cutoff for biomarker | Hazard ratio [95% CI] |
|---|---|---|---|---|---|---|---|---|---|
| Katakami | 2014 | Primary | 1040 | 113 | 3p MACE, Revasc, HF, Angina, PAD | 15 | Low | per 1 SD (cm/s) | 1.33 [1.09, 1.62] |
| Wijkman | 2016 | Primary | 627 | 45 | 3p MACE | 9 | Medium | per 1 m/s | 1.14 [1.00, 1.30] |
| Nakamura | 2010 | Mixed | 191 | 59 | CHD, Revasc, HF, ACM | 10 | Low | ≥1730 cm/sec vs. <1730 cm/sec | 1.97 [1.01, 3.84] |
| Cardoso | 2013 | Mixed | 565 | 88 | 3p MACE, Revasc, HF, PAD, ACM | 17 | Low | per 1 SD (m/s) | 1.32 [1.06, 1.64] |

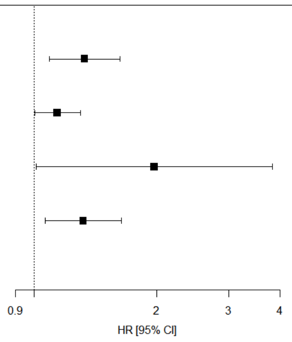

#### Supplemental Figure 3G. SPECT

**Single-photon emission computed tomography (SPECT) scintigraphy**

| First author | Year | Study population | DM pop N | Event N | Outcome | No. of covariates | Risk of bias | Cutoff for biomarker | Hazard ratio [95% CI] |
|---|---|---|---|---|---|---|---|---|---|
| Cosson | 2011 | Primary | 688 | 98 | 3p MACE, Revasc, HF, PAD | 5 | High | presence of silent myocardial ischemia vs. absence | 1.80 [1.02, 3.17] |
| Petretta | 2013 | Mixed | 672 | 63 | CHD, CVM, Revasc | 5 | Low | transient ischemic dilatation ≥1.19 vs. <1.19 | 3.60 [1.95, 6.64] |
| Djaileb | 2021 | Mixed | 166 | 44 | CHD, CVM, Revasc, Angina | 4 | High | stress perfusion defect ≥4 vs. <4 | 2.06 [1.03, 4.11] |

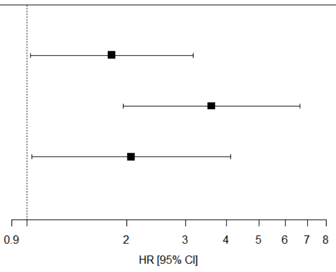

### Supplemental Figure 3H.  TnI

**Troponin I**

| First author | Year | Study population | DM pop N | Event N | Outcome | No. of covariates | Risk of bias | Per unit increase / Cutoff for biomarker | Hazard ratio [95% CI] |
|---|---|---|---|---|---|---|---|---|---|
| Yiu | 2014 | Mixed | 276 | 43 | 3p MACE, HF | 7 | Medium | per 1 ng/L | 2.85 [1.15, 7.05] |
| Wong | 2019 | Secondary | 1617 | 328 | 3p MACE, PAD | 3 | High | >14.2 pg/mL vs. ≤14.2 pg/mL | 1.69 [1.33, 2.15] |
| Sharma | 2020 | Secondary | 5154 | 837 | 3p MACE, HF | 10 | Medium | per 1 logarithm to the base 2 (ng/L) | 1.04 [1.00, 1.09] |

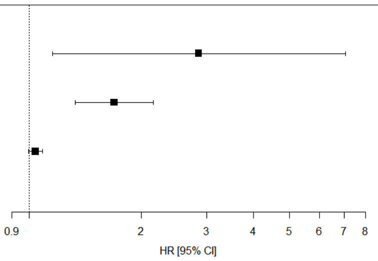

#### Supplemental Figure 3I.  TyG

**Triglyceride glucose (TyG) index**

| First author | Year | Study population | DM pop N | Event N | Outcome | No. of covariates | Risk of bias | Per unit increase | Hazard ratio [95% CI] |
|---|---|---|---|---|---|---|---|---|---|
| Jin | 2018 | Secondary | 1282 | 160 | 3p MACE, Revasc, Angina | 12 | High | per 1 unit increase of TyG index | 1.69 [1.26, 2.28] |
| Zhao | 2020 | Secondary | 798 | 180 | CHD, Revasc, ACM | 30 | Low | per 1 unit increase of TyG index | 3.21 [2.40, 4.29] |

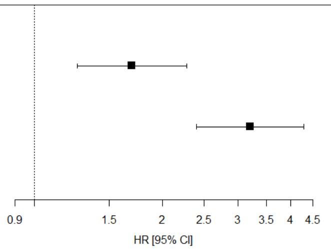

**Supplemental Figure 3** Caption: (A) Coronary artery calcium score (CACS), (B) carotid plaque, (C) c-reactive protein (CRP), (D) galectin-3, (E) growth differentiation factor (GDF-15), (F) pulse wave velocity (PWV), (G) single-photon emission computed tomography (SPECT) scintigraphy, (H) troponin I, and (I) triglyceride glucose (TyG) index]. HR, hazard ratio; CI, confidence interval; DM pop N, sample size for diabetes population; Event N, number of individuals developed CVD outcomes; 3p MACE, 3-point major adverse cardiovascular events; Revasc, revascularization; HF, heart failure; CHD, coronary heart disease; PAD, peripheral artery disease; CVM, cardiovascular mortality; ACM, all-cause mortality.

**Supplemental Figure 4:** Meta-analyses of studies on C-reactive protein (CRP), Pulse wave velocity (PWV), and Triglyceride glucose (TyG) index.

Supplemental Figure 4A. CRP (binary/categorical)

**Meta-analysis for the hazard ratio of C-reactive protein (CRP) analysed as a binary or categorical exposure**

| First author | Year | Study population | DM pop N | Event N | Outcome | No. of covariates | Risk of bias | Cutoff for biomarker | Weight | Hazard ratio [95% CI] |
|---|---|---|---|---|---|---|---|---|---|---|
| Schulze | 2004 | Primary | 746 | 103 | CHD, Stroke, Revasc | 13 | Low | Q4 vs. Q1 (mg/L) | 32.74% | 2.62 [1.29, 5.32] |
| Linnemann | 2006 | Mixed | 292 | NR | 3p MACE, PAD, ACM | 8 | Med. | Q4 (≥9.2 mg/L) vs. Q1 (≤2.7 mg/L) | 17.35% | 5.40 [1.45, 20.12] |
| McMurray | 2011 | Mixed | 4038 | 1010 | 3p MACE, HF | 29 | Low | ≥6.6 mg/L vs. ≤3.0 mg/L | 49.91% | 1.44 [1.23, 1.69] |
| **Random-effects REML model** | | | | | | | **Overall** | | 100% | 2.20 [1.12, 4.32] |

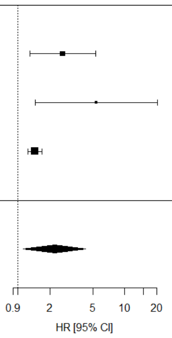

**Random-effects REML model**
$I^2$ = 68.33%; $\tau^2$ = 0.23; $P_Q$ = 0.0435; HR = 2.2[ 1.12 - 4.32], $P$ = 0.0215

**Fixed-effects model**
$I^2$ = 68.11% ; $P_Q$ = 0.0435; HR = 1.51[ 1.29 - 1.76], $P$ = 1.67e-07

#### Supplemental Figure 4B. CRP (continuous)

**Meta-analysis for the hazard ratio of C-reactive protein (CRP) analysed as a continuous exposure**

| First author | Year | Study population | DM pop N | Event N | Outcome | No. of covariates | Risk of bias | Per unit increase | Weight | Hazard ratio [95% CI] |
|---|---|---|---|---|---|---|---|---|---|---|
| vonScholten | 2016 | Primary | 200 | 40 | 3p MACE, HF | 10 | Medium | per 1 SD of the logarithms (mg/L) | 12.10% | 1.00 [0.71, 1.41] |
| Cardoso | 2016 | Mixed | 616 | 184 | 3p MACE, ACM | 17 | Low | per 1 SD of the logarithms (mg/L) | 23.83% | 1.30 [1.09, 1.55] |
| Scirica | 2016 | Mixed | 16492 | NR | 3p MACE, HF | 9 | Low | per 1 SD of the logarithms (mg/L) | 33.69% | 1.17 [1.10, 1.25] |
| Lowe | 2014 | Secondary | 3865 | 709 | 3p MACE, Renal | 16 | Low | per 1 SD of the logarithms (mg/L) | 30.38% | 0.95 [0.85, 1.06] |
| **Random-effects REML model** | | | | | | | **Overall** | | 100% | 1.10 [0.95, 1.28] |

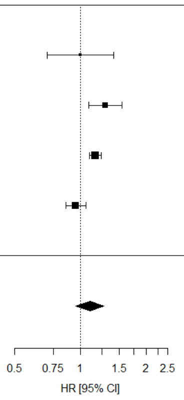

**Random-effects REML model**
$I^2$ = 79.53%; $\tau^2$ = 0.016; $P_Q$ = 0.00253; HR = 1.1[ 0.95 - 1.28], $P$ = 0.186

**Fixed-effects model**
$I^2$ = 79.01% ; $P_Q$ = 0.00253; HR = 1.12[ 1.06 - 1.18], $P$ = 1.73e-05

#### Supplemental Figure 4C.  PWV

**Meta-analysis for the hazard ratio of pulse wave velocity (PWV)**

| First author | Year | Study population | DM pop N | Event N | Outcome | No. of covariates | Risk of bias | Per unit increase | Weight | Hazard ratio [95% CI] |
|---|---|---|---|---|---|---|---|---|---|---|
| Katakami | 2014 | Primary | 1040 | 113 | 3p MACE, Revasc, HF, Angina, PAD | 15 | Low | per 1 SD (cm/s) | 54.11% | 1.33 [1.09, 1.62] |
| Cardoso | 2013 | Mixed | 565 | 88 | 3p MACE, Revasc, HF, PAD, ACM | 17 | Low | per 1 SD (m/s) | 45.89% | 1.32 [1.06, 1.64] |
| **Random-effects REML model** | | | | | | **Overall** | | | 100% | 1.33 [1.15, 1.53] |

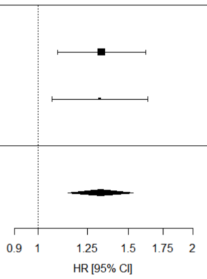

**Random-effects REML model**
$I^2$ = 0%; $\tau^2$ = 0; $P_Q$ = 0.96; HR = 1.33[ 1.15 - 1.53], $P$ = 0.000152

**Fixed-effects model**
$I^2$ = 0% ; $P_Q$ = 0.96; HR = 1.33[ 1.15 - 1.53], $P$ = 0.000152

#### Supplemental Figure 4D. TyG

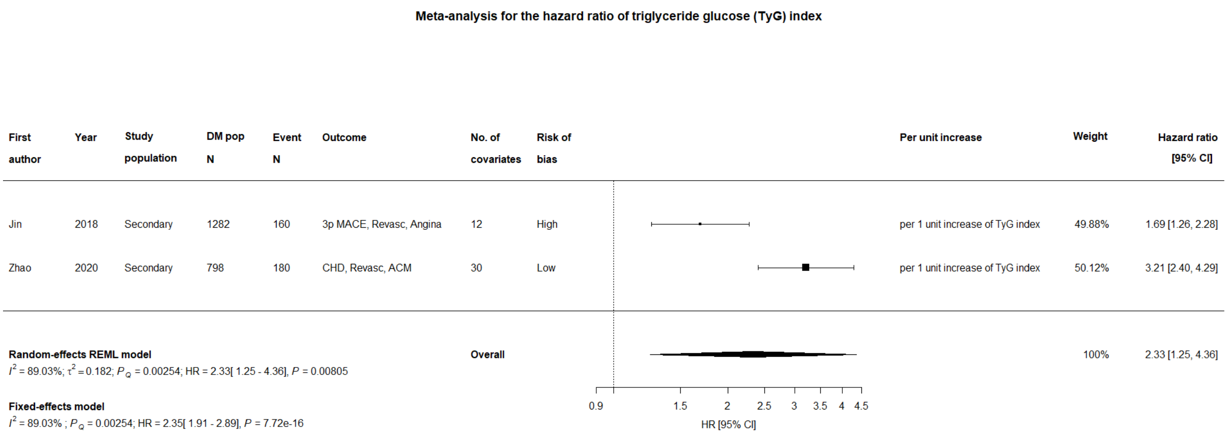

**Supplemental Figure 4** Caption: (A) CRP analyzed as a binary or categorical exposure, (B) CRP analyzed as a continuous exposure with a z-score of logarithm transformation, (C) PWV analyzed as a continuous exposure with a z-score transformation, and (D) TyG index analyzed as a continuous exposure. For the overall pooled estimate, a random-effects model was used only if the heterogeneity test was statistically significant (Cochran's Q test *p*-value <0.1 or the $I^2$ statistic > 75%). HR, hazard ratio; CI, confidence interval; DM pop N, sample size for diabetes population; Event N, number of individuals developed CVD outcomes; 3p MACE, 3-point major adverse cardiovascular events; Revasc, revascularization; HF, heart failure; CHD, coronary heart disease; PAD, peripheral artery disease; CVM, cardiovascular mortality; ACM, all-cause mortality.

#### Supplemental Figure 5: Meta-analyses of studies GRS-CHD and *GLUL*.

##### Supplemental Figure 5A. GRS-CHD

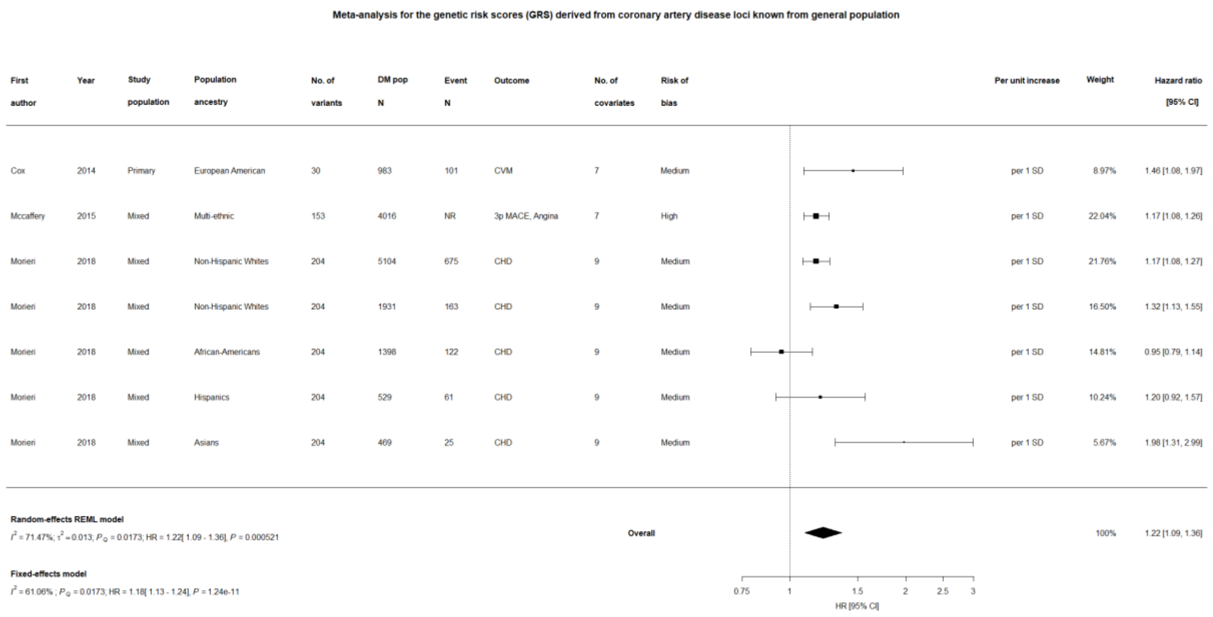

#### Supplemental Figure 5B. *GLUL*

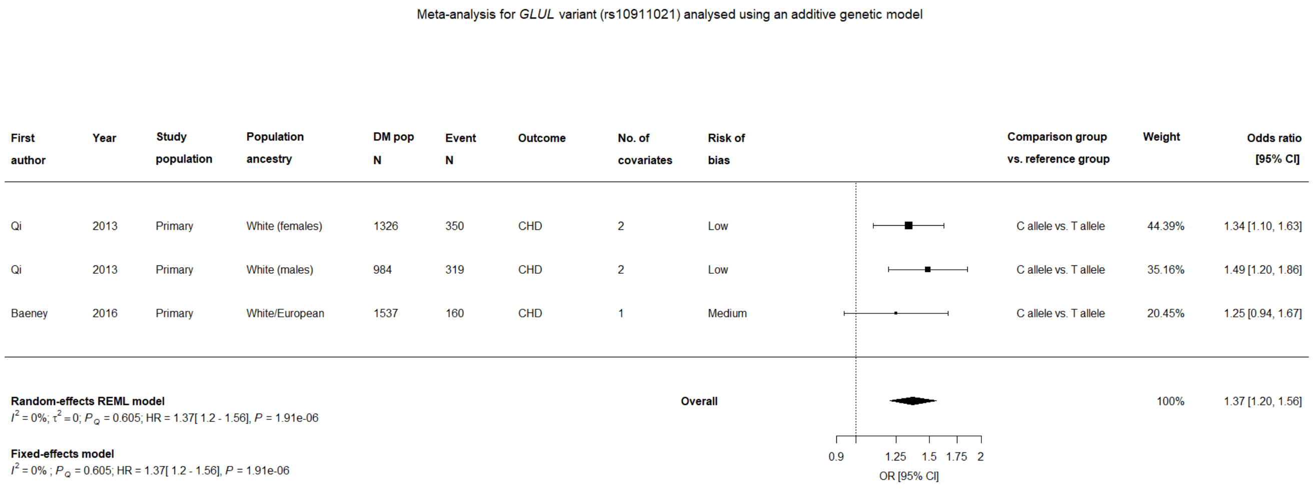

**Supplemental Figure 5** Caption: Results of meta-analysis for genetic markers [(A) z-scores of genetic risk scores, (B) *GLUL* variant rs10911021]. - For the overall pooled estimate, a random-effects model was used only if the heterogeneity test was statistically significant (Cochran's Q test *p*-value <0.1 or the $I^2$ statistic > 75%). HR, hazard ratio; CI, confidence interval; DM pop N, sample size for diabetes population; Event N, number of individuals developed CVD outcomes; 3p MACE, 3-point major adverse cardiovascular events; CHD, coronary heart disease; CVM, cardiovascular mortality.

**Supplemental Figure 6.** Concordance of c-statistics between the countries of origin (development cohort) of non-genetic risk scores versus external validation cohort.

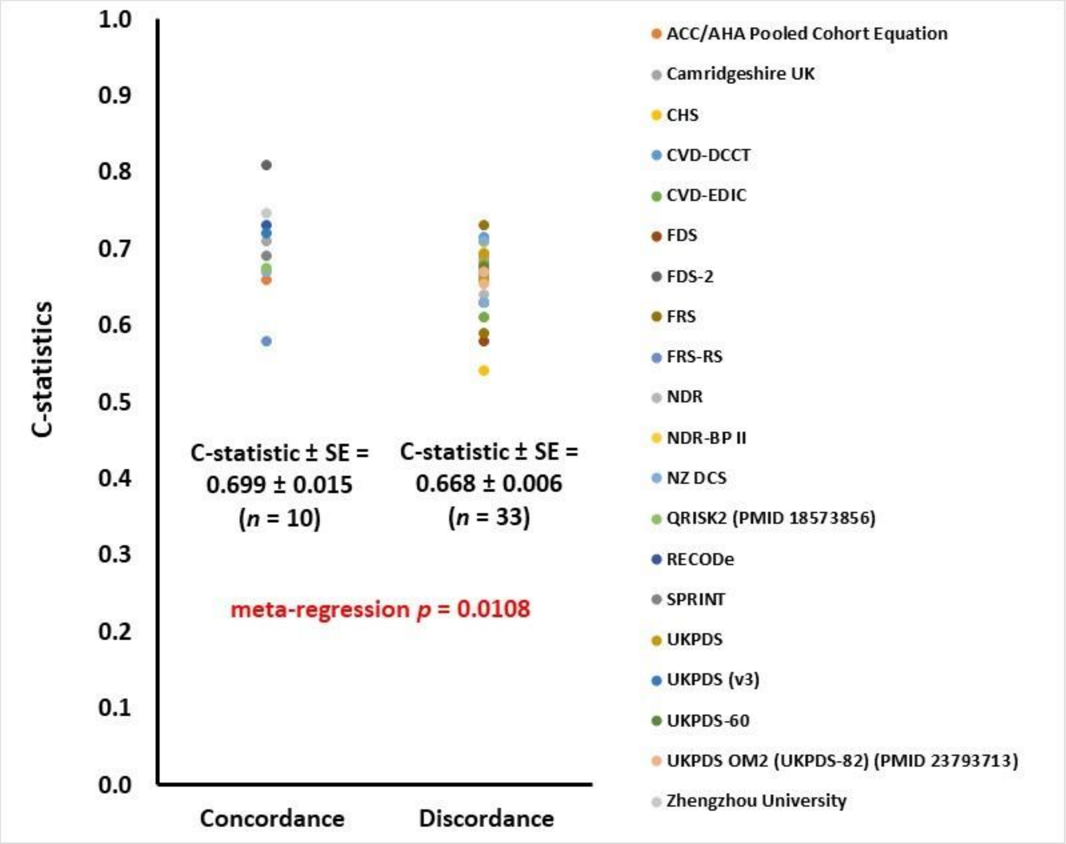

**Supplemental Figure 6** Caption: The pooled c-statistics and the corresponding standard error (SE) were computed using a random-effects model. The *p*-value was obtained from the meta-regression analysis using a mixed-effects model.

### Supplemental Figure 7. Meta-analysis for c-statistic of Risk Scores (External Validation)

#### Supplemental Figure 7A. ADVANCE

**Meta-analysis for the c-statistics of 'ADVANCE' risk scores (external validation cohorts)**

| First author | Year | Risk score | Year original risk score published | Derivation country(ies) | Validation cohort | Country(ies) of validation cohort | Risk of bias | Weight (%) | C-statistic [95% CI] |
|---|---|---|---|---|---|---|---|---|---|
| Kengne | 2011 | ADVANCE | 2011 | Europe/Asia | DIABHYCAR | Multinational | High | 12.52% | 0.68 [0.65, 0.72] |
| vanderLeeuw | 2015 | ADVANCE | 2011 | Europe/Asia | EPIC-NL | Netherlands | Medium | 4.62% | 0.62 [0.54, 0.70] |
| vanderLeeuw | 2015 | ADVANCE | 2011 | Europe/Asia | EPIC-Potsdam | Germany | Medium | 4.62% | 0.67 [0.59, 0.75] |
| vanderLeeuw | 2015 | ADVANCE | 2011 | Europe/Asia | SMART | Netherlands | Medium | 3.45% | 0.68 [0.59, 0.77] |
| Wan | 2018 | ADVANCE | 2011 | Europe/Asia | Hong Kong Hospital Authority (men) | China | Low | 24.23% | 0.69 [0.68, 0.70] |
| Wan | 2018 | ADVANCE | 2011 | Europe/Asia | Hong Kong Hospital Authority (women) | China | Low | 24.23% | 0.71 [0.70, 0.72] |
| Read | 2018 | ADVANCE | 2011 | Europe/Asia | National Scottish Diabetes Register | Scotland | Medium | 26.36% | 0.67 [0.66, 0.67] |
| Overall | | | | | | | | 100% | 0.68 [0.66, 0.70] |

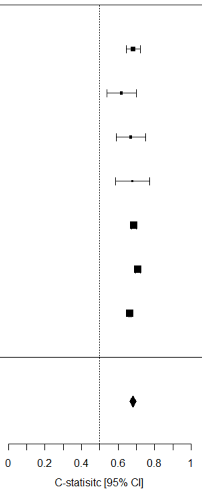

**Random-effects REML model**

$I^2$ = 84.22%; $\tau^2$ = 0; $P_Q$ = 2.17e-09; C-statistic [95% CI] = 0.68[ 0.66 - 0.7], $P$ = 0

**Fixed-effects model**

$I^2$ = 88.39% ; $P_Q$ = 2.17e-09; C-statistic [95% CI] = 0.67[ 0.67 - 0.68], $P$ = 0

#### Supplemental Figure 7B. CHS

**Meta-analysis for the c-statistics of 'CHS' risk scores (external validation cohorts)**

| First author | Year | Risk score | Year original risk score published | Derivation country(ies) | Validation cohort | Country(ies) of validation cohort | Risk of bias | Weight (%) | C-statistic [95% CI] |
|---|---|---|---|---|---|---|---|---|---|
| vanderLeeuw | 2015 | CHS | 2013 | United States | EPIC-NL | Netherlands | Medium | 22.12% | 0.54 [0.46, 0.62] |
| vanderLeeuw | 2015 | CHS | 2013 | United States | EPIC-Potsdam | Germany | Medium | 21.11% | 0.61 [0.52, 0.70] |
| vanderLeeuw | 2015 | CHS | 2013 | United States | SMART | Netherlands | Medium | 20.13% | 0.68 [0.59, 0.77] |
| vanderLeeuw | 2015 | CHS | 2013 | United States | National Scottish Diabetes Register | Scotland | Medium | 36.64% | 0.67 [0.67, 0.68] |
| **Overall** | | | | | | | | 100% | 0.63 [0.57, 0.70] |

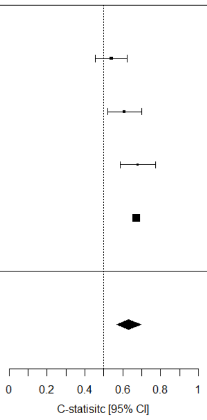

**Random-effects REML model**

$I^2$ = 72.99%; $\tau^2$ = 0.003; $P_Q$ = 0.00956; C-statistic [95% CI] = 0.63[ 0.57 - 0.7], $P$ = 5.16e-85

**Fixed-effects model**

$I^2$ = 73.78% ; $P_Q$ = 0.00956; C-statistic [95% CI] = 0.67[ 0.67 - 0.68], $P$ = 0

#### Supplemental Figure 7C. CVD-EDIC

**Meta-analysis for the c-statistics of 'CVD-EDIC' risk scores (external validation cohorts)**

| First author | Year | Risk score | Year original risk score published | Derivation country(ies) | Validation cohort | Country(ies) of validation cohort | Risk of bias | Weight (%) | C-statistic [95% CI] |
|---|---|---|---|---|---|---|---|---|---|
| Lagani | 2015 | CVD-EDIC | 2015 | United States | Chorleywood Medical Center | United Kingdom | High | 3.83% | 0.66 [0.51, 0.81] |
| vanderLeeuw | 2015 | CVD-EDIC | 2015 | United States | EPIC-NL | Netherlands | Medium | 11.46% | 0.58 [0.50, 0.66] |
| vanderLeeuw | 2015 | CVD-EDIC | 2015 | United States | EPIC-Potsdam | Germany | Medium | 11.46% | 0.68 [0.60, 0.76] |
| vanderLeeuw | 2015 | CVD-EDIC | 2015 | United States | SMART | Netherlands | Medium | 7.82% | 0.69 [0.59, 0.79] |
| Read | 2018 | CVD-EDIC | 2015 | United States | National Scottish Diabetes Register | Scotland | Medium | 65.43% | 0.67 [0.67, 0.67] |
| **Overall** | | | | | | | | 100% | 0.66 [0.63, 0.69] |

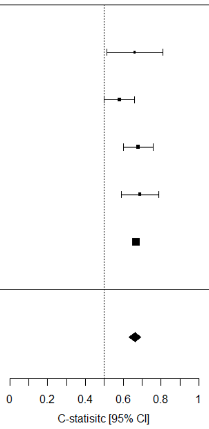

**Random-effects REML model**

$I^2$ = 23.31%; $\tau^2$ = 0; $P_Q$ = 0.279; C-statistic [95% CI] = 0.66[ 0.63 - 0.69], $P$ = 0

**Fixed-effects model**

$I^2$ = 21.22% ; $P_Q$ = 0.279; C-statistic [95% CI] = 0.67[ 0.67 - 0.67], $P$ = 0

#### Supplemental Figure 7D. NDR

**Meta-analysis for the c-statistics of 'NDR' risk scores (external validation cohorts)**

| First author | Year | Risk score | Year original risk score published | Derivation country(ies) | Validation cohort | Country(ies) of validation cohort | Risk of bias | Weight (%) | C-statistic [95% CI] |
|---|---|---|---|---|---|---|---|---|---|
| vanderLeeuw | 2015 | NDR | 2008 | Sweden | EPIC-NL | Netherlands | Medium | 6.18% | 0.64 [0.56, 0.72] |
| vanderLeeuw | 2015 | NDR | 2008 | Sweden | EPIC-Potsdam | Germany | Medium | 6.84% | 0.67 [0.60, 0.74] |
| vanderLeeuw | 2015 | NDR | 2008 | Sweden | SMART | Netherlands | Medium | 4.29% | 0.64 [0.54, 0.74] |
| Wan | 2018 | NDR | 2008 | Sweden | Hong Kong Hospital Authority (men) | China | Low | 27.07% | 0.69 [0.68, 0.70] |
| Wan | 2018 | NDR | 2008 | Sweden | Hong Kong Hospital Authority (women) | China | Low | 26.90% | 0.71 [0.70, 0.72] |
| Read | 2018 | NDR | 2008 | Sweden | National Scottish Diabetes Register | Scotland | Medium | 28.73% | 0.66 [0.66, 0.67] |
| **Overall** | | | | | | | | 100% | 0.68 [0.66, 0.70] |

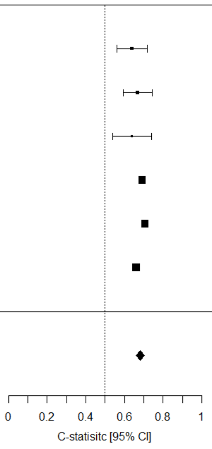

**Random-effects REML model**

$I^2$ = 89.18%; $\tau^2$ = 0; $P_Q$ = 8.59e-12; C-statistic [95% CI] = 0.68[ 0.66 - 0.7], $P$ = 0

**Fixed-effects model**

$I^2$ = 91.77% ; $P_Q$ = 8.59e-12; C-statistic [95% CI] = 0.67[ 0.67 - 0.68], $P$ = 0

#### Supplemental Figure 7E. NZ DCS

**Meta-analysis for the c-statistics of 'NZ DCS' risk scores (external validation cohorts)**

| First author | Year | Risk score | Year original risk score published | Derivation country(ies) | Validation cohort | Country(ies) of validation cohort | Risk of bias | Weight (%) | C-statistic [95% CI] |
|---|---|---|---|---|---|---|---|---|---|
| Elley | 2010 | NZ DCS | 2010 | New Zealand | New Zealand (South) | New Zealand | Medium | 27.29% | 0.67 [0.66, 0.68] |
| vanderLeeuw | 2015 | NZ DCS | 2010 | New Zealand | EPIC-NL | Netherlands | Medium | 5.28% | 0.63 [0.55, 0.71] |
| vanderLeeuw | 2015 | NZ DCS | 2010 | New Zealand | EPIC-Potsdam | Germany | Medium | 5.87% | 0.66 [0.59, 0.73] |
| vanderLeeuw | 2015 | NZ DCS | 2010 | New Zealand | SMART | Netherlands | Medium | 3.59% | 0.67 [0.57, 0.77] |
| Wan | 2018 | NZ DCS | 2010 | New Zealand | Hong Kong Hospital Authority (men) | China | Low | 29.11% | 0.69 [0.68, 0.70] |
| Wan | 2018 | NZ DCS | 2010 | New Zealand | Hong Kong Hospital Authority (women) | China | Low | 28.87% | 0.71 [0.70, 0.72] |
| **Overall** | | | | | | | | 100% | 0.68 [0.66, 0.70] |

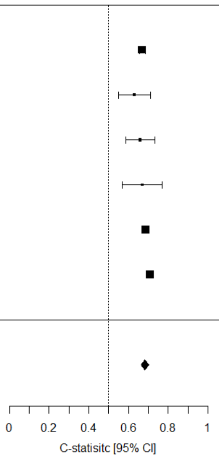

**Random-effects REML model**
$I^2$ = 76.44%; $\tau^2$ = 0; $P_Q$ = 0.00145; C-statistic [95% CI] = 0.68[ 0.66 - 0.7], $P$ = 0

**Fixed-effects model**
$I^2$ = 74.55% ; $P_Q$ = 0.00145; C-statistic [95% CI] = 0.69[ 0.68 - 0.7], $P$ = 0

### Supplemental Figure 7F. UKPDS

**Meta-analysis for the c-statistics of 'UKPDS' risk scores (external validation cohorts)**

| First author | Year | Risk score | Year original risk score published | Derivation country(ies) | Validation cohort | Country(ies) of validation cohort | Risk of bias | Weight (%) | C-statistic [95% CI] |
|---|---|---|---|---|---|---|---|---|---|
| Simmons | 2009 | UKPDS | 1998 | United Kingdom | Epic Norfolk | United Kingdom | Medium | 3.70% | 0.72 [0.66, 0.78] |
| Davis | 2009 | UKPDS | 1998 | United Kingdom | FDS | Australia | Medium | 2.28% | 0.68 [0.60, 0.76] |
| Kengne | 2010 | UKPDS | 1998 | United Kingdom | ADVANCE | Europe/Asia | Low | 8.46% | 0.68 [0.65, 0.72] |
| vanDieren | 2011 | UKPDS | 1998 | United Kingdom | Epic-NL; Epic-Potsdam | Netherlands/Germany | Low | 0.77% | 0.66 [0.51, 0.81] |
| Tanaka | 2013 | UKPDS | 1998 | United Kingdom | JDCS/J-EDIT | Japan | Medium | 3.33% | 0.69 [0.63, 0.76] |
| Piniés | 2014 | UKPDS | 1998 | United Kingdom | BASCORE | Spain | Low | 4.93% | 0.62 [0.57, 0.67] |
| Basu | 2017 | UKPDS | 1998 | United Kingdom | LOOK AHEAD | United States | High | 14.74% | 0.67 [0.65, 0.69] |
| Quan | 2019 | UKPDS | 1998 | United Kingdom | Hong Kong Dataset | China | High | 30.27% | 0.68 [0.67, 0.68] |
| Quan | 2019 | UKPDS | 1998 | United Kingdom | Hong Kong Dataset | China | High | 30.12% | 0.65 [0.65, 0.66] |
| Venäläinen | 2020 | UKPDS | 1998 | United Kingdom | Hoorn Study | Netherlands | Medium | 1.40% | 0.66 [0.55, 0.77] |
| **Overall** | | | | | | | | 100% | 0.67 [0.66, 0.68] |

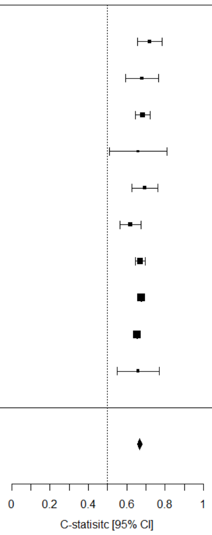

**Random-effects REML model**
$I^2$ = 89.79%; $\tau^2$ = 0; $P_Q$ = 2e-23; C-statistic [95% CI] = 0.67[ 0.66 - 0.68], $P$ = 0

**Fixed-effects model**
$I^2$ = 93.02% ; $P_Q$ = 2e-23; C-statistic [95% CI] = 0.67[ 0.67 - 0.67], $P$ = 0

**Supplemental Figure 8.** Use of covariates in biomarker studies

Supplemental Figure 8A. Total Adjusted Covariates

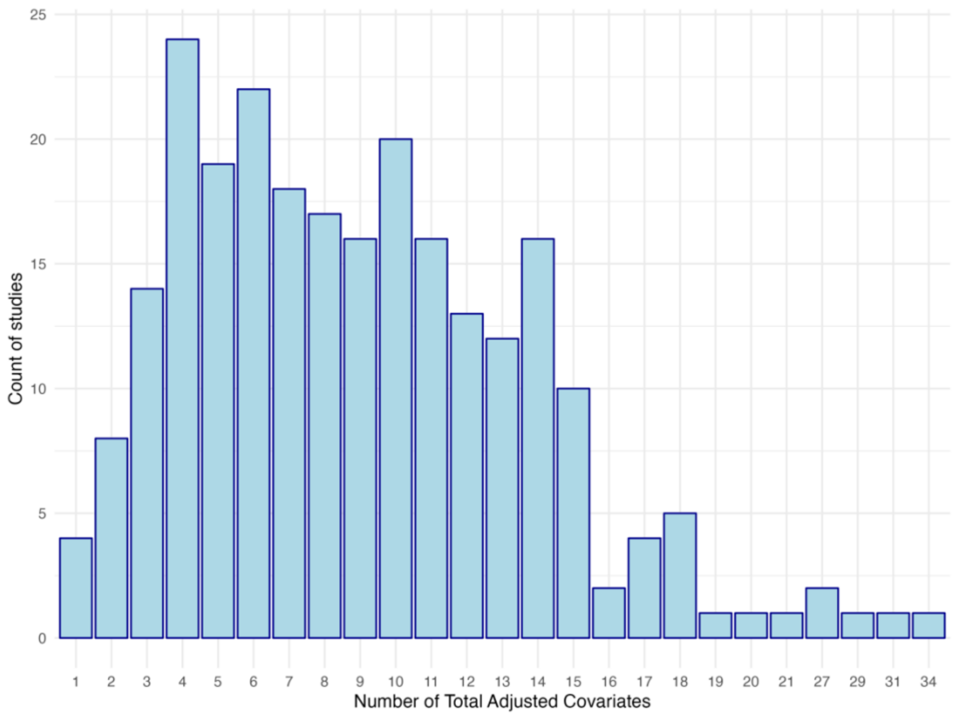

Supplemental Figure 8B. Adjusted Traditional CVD Risk Factors

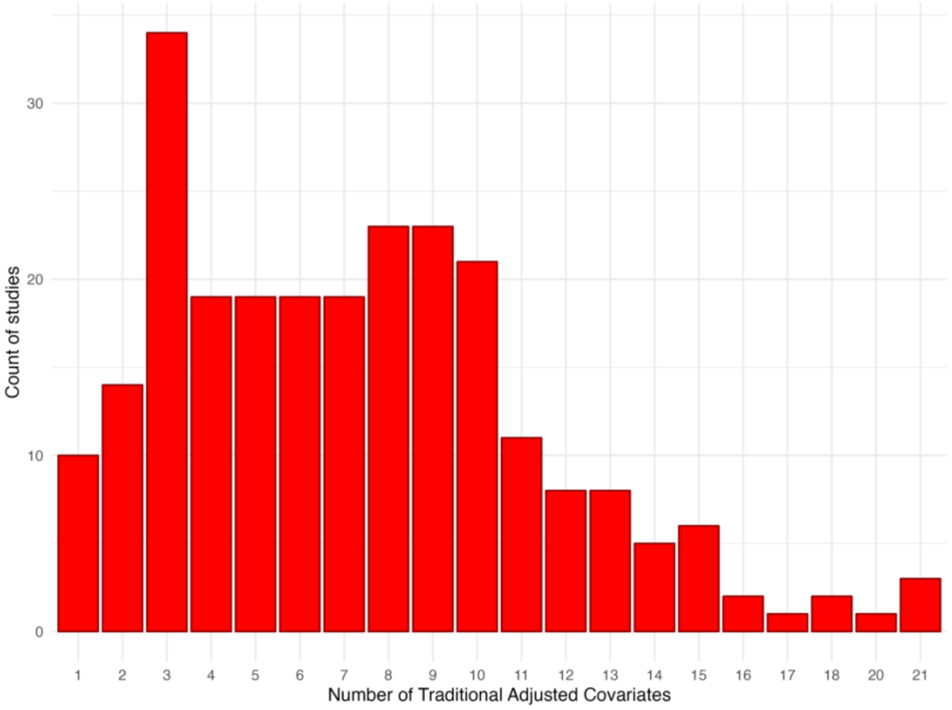

**Supplemental Figure 8** Caption: Histogram showing the count of studies per (A) number of adjusted covariates and (B) number of traditional CVD risk factors adjusted for in the analyses.

**Supplemental Figure 9.** Most commonly used covariates in biomarker studies

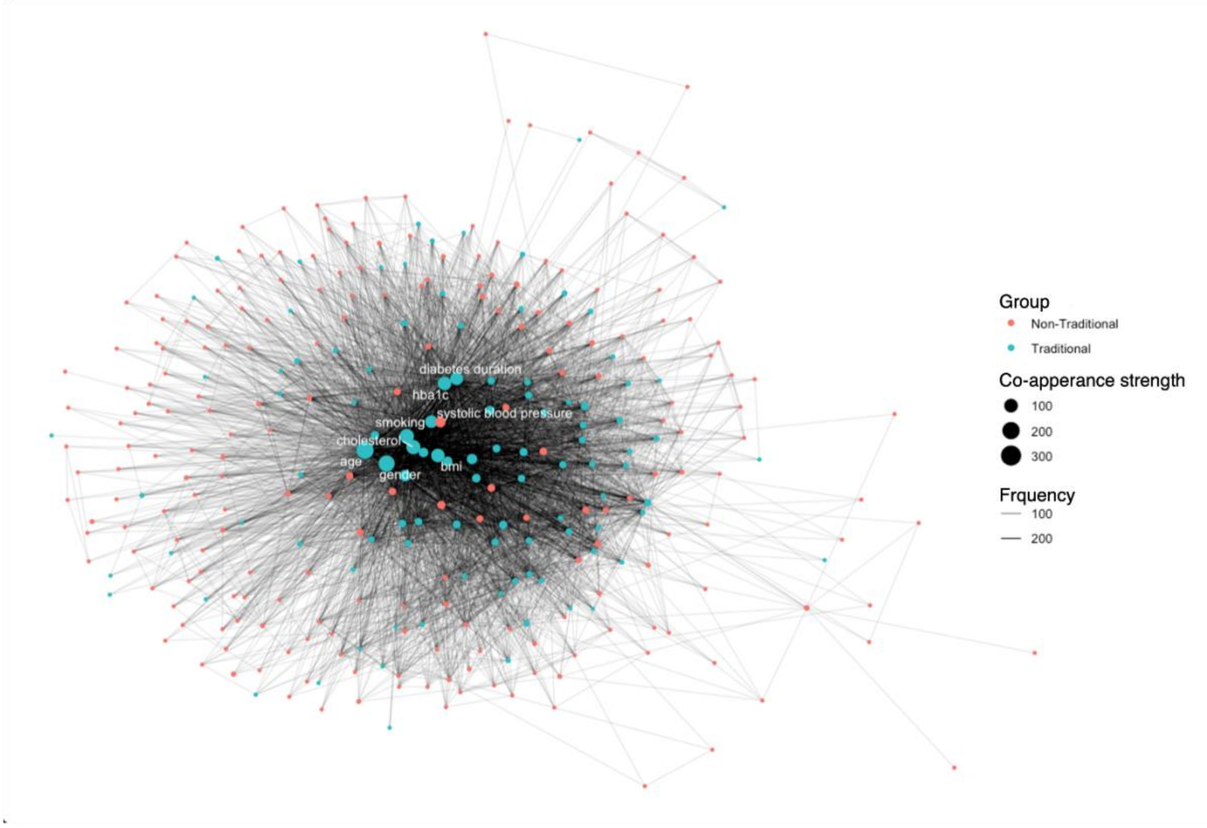

**Supplemental Figure 9** Caption: The network figure represents the connections of adjusted covariates in 416 studies. The nodes in the figure correspond to covariates, and the size of the node represents the frequency of the covariate appearing in the groups of studies. The more centrally located a node, the more important its role as a factor in the network. Nodes are color-coded based on whether they are traditional or non-traditional covariates. The edges (lines connecting co-variates) vary in clarity, indicating the frequency of connections between covariates. An interactive version of this figure can be found here: https://hugofitipaldi.shinyapps.io/T2D_prognostic/

#### Supplemental Figure 10. **Sensitivity Analyses Excluding High Risk of Bias Studies in Biomarkers**

##### Supplemental Figure 10A. NT-proBNP (continuous)

**Meta-analysis for the hazard ratio of N terminal pro B type natriuretic peptide (NT-proBNP) analysed as a continuous exposure (sensitivity analysis)**

| First author | Year | Study population | DM pop N | Event N | Outcome | No. of covariates | Risk of bias | Per unit increase | Weight | Hazard ratio [95% CI] |
|---|---|---|---|---|---|---|---|---|---|---|
| McMurray | 2011 | Mixed | 4038 | 1010 | 3p MACE, HF | 29 | Low | per 1 logarithm (pg/mL) | 26.20% | 1.30 [1.15, 1.46] |
| Bruno | 2013 | Mixed | 1825 | 175 | CVM | 13 | Low | per 1 logarithm (pg/mL) | 23.05% | 1.69 [1.39, 2.05] |
| Resl | 2016 | Mixed | 746 | 171 | CHD, Stroke, HF, PAD, ACM | 4 | Low | per 1 logarithm (pg/mL) | 27.29% | 1.17 [1.07, 1.27] |
| Rørth | 2019 | Secondary | 759 | NR | CHD, CVM, HF, ACM | 13 | Low | per 1 logarithm (pg/mL) | 23.47% | 1.85 [1.54, 2.23] |
| **Random-effects REML model** | | | | | | **Overall** | | | 100% | 1.46 [1.18, 1.80] |

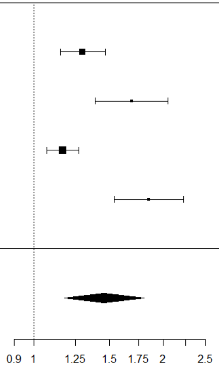

**Random-effects REML model**
$I^2$ = 89.76%; $\tau^2$ = 0.041; $P_Q$ = 6.97e-06; HR = 1.46[ 1.18 - 1.8], $P$ = 0.000504

**Fixed-effects model**
$I^2$ = 88.74% ; $P_Q$ = 6.97e-06; HR = 1.31[ 1.24 - 1.4], $P$ = 4.09e-18

#### Supplemental Figure 10B. TnT (binary/categorical)

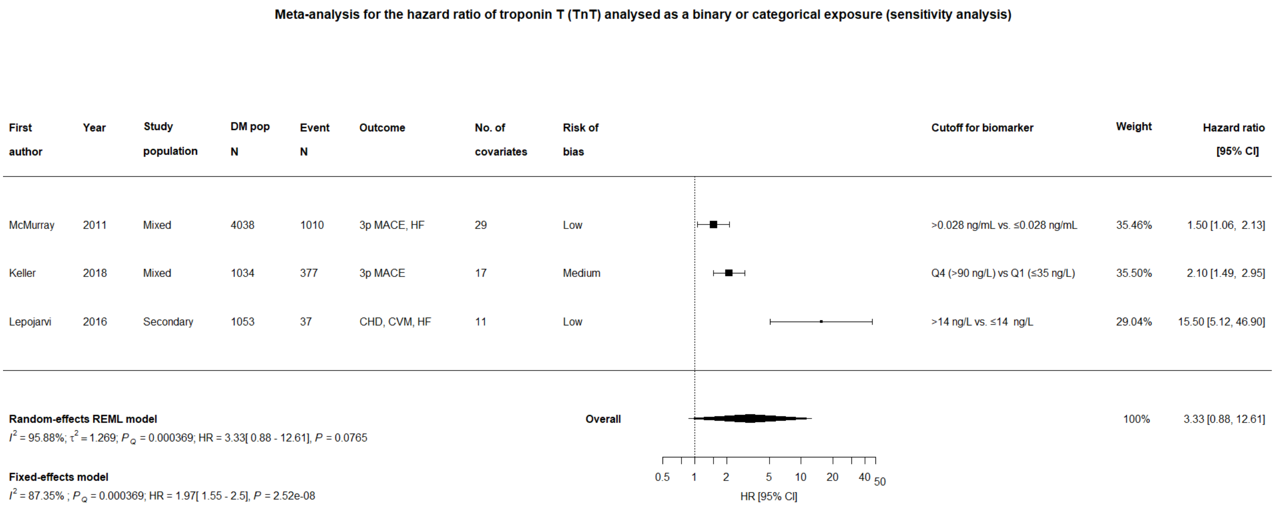

**Supplemental Figure 10** Caption: (A) NT-proBNP analyzed as a continuous exposure with a logarithm transformation, and (B) troponin T (TnT) analyzed as a binary or categorical exposure]. For the overall pooled estimate, a random-effects model was used only if the heterogeneity test was statistically significant (Cochran's Q test p-value <0.1 or the I2 statistic > 75%). HR, hazard ratio; CI, confidence interval; DM pop N, sample size for diabetes population; Event N, number of individuals developed CVD outcomes; 3p MACE, 3-point major adverse cardiovascular events; HF, heart failure; CHD, coronary heart disease; PAD, peripheral artery disease; CVM, cardiovascular mortality; ACM, all-cause mortality.

#### **Supplemental Figure 11.** Sensitivity Analyses Excluding High Risk of Bias Studies in Genetic Studies.

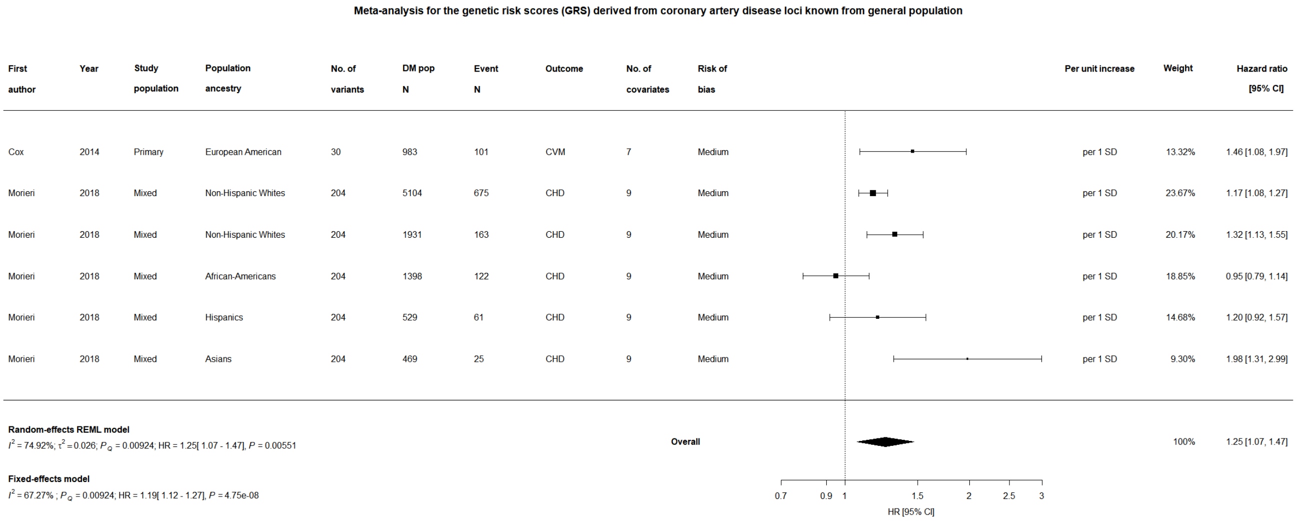

**Supplemental Figure 11** Caption: For the overall pooled estimate, a random-effects model was used only if the heterogeneity test was statistically significant (Cochran's Q test *p*-value <0.1 or the $I^2$ statistic > 75%). HR, hazard ratio; CI, confidence interval; DM pop N, sample size for diabetes population; Event N, number of individuals developed CVD outcomes; CHD, coronary heart disease.

#### Supplemental Figure 12. Sensitivity Analyses Excluding High Risk of Bias Studies in Risk Scores (External Validation)

##### Supplemental Figure 12A. ADVANCE Risk Score

**Meta-analysis for the c-statistics of 'ADVANCE' risk scores (external validation cohorts)**

| First author | Year | Risk score | Year original risk score published | Derivation country(ies) | Validation cohort | Country(ies) of validation cohort | Risk of bias | Weight (%) | C-statistic [95% CI] |
|---|---|---|---|---|---|---|---|---|---|
| vanderLeeuw | 2015 | ADVANCE | 2011 | Europe/Asia | EPIC-NL | Netherlands | Medium | 6.07% | 0.62 [0.54, 0.70] |
| vanderLeeuw | 2015 | ADVANCE | 2011 | Europe/Asia | EPIC-Potsdam | Germany | Medium | 6.07% | 0.67 [0.59, 0.75] |
| vanderLeeuw | 2015 | ADVANCE | 2011 | Europe/Asia | SMART | Netherlands | Medium | 4.57% | 0.68 [0.59, 0.77] |
| Wan | 2018 | ADVANCE | 2011 | Europe/Asia | Hong Kong Hospital Authority (men) | China | Low | 27.13% | 0.69 [0.68, 0.70] |
| Wan | 2018 | ADVANCE | 2011 | Europe/Asia | Hong Kong Hospital Authority (women) | China | Low | 27.13% | 0.71 [0.70, 0.72] |
| Read | 2018 | ADVANCE | 2011 | Europe/Asia | National Scottish Diabetes Register | Scotland | Medium | 29.04% | 0.67 [0.66, 0.67] |
| **Overall** | | | | | | | | 100% | 0.68 [0.66, 0.70] |

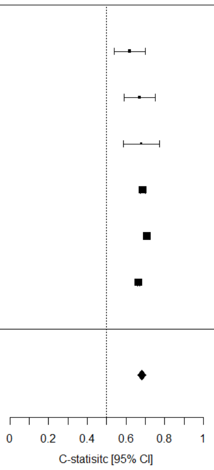

**Random-effects REML model**

$I^2$ = 88.44%; $\tau^2$ = 0; $P_Q$ = 7.2e-10; C-statistic [95% CI] = 0.68[ 0.66 - 0.7], $P$ = 0

**Fixed-effects model**

$I^2$ = 90.27% ; $P_Q$ = 7.2e-10; C-statistic [95% CI] = 0.67[ 0.67 - 0.68], $P$ = 0

#### Supplemental Figure 12B. CVD-EDIC

**Meta-analysis for the c-statistics of 'CVD-EDIC' risk scores (external validation cohorts)**

| First author | Year | Risk score | Year original risk score published | Derivation country(ies) | Validation cohort | Country(ies) of validation cohort | Risk of bias | Weight (%) | C-statistic [95% CI] |
|---|---|---|---|---|---|---|---|---|---|
| vanderLeeuw | 2015 | CVD-EDIC | 2015 | United States | EPIC-NL | Netherlands | Medium | 15.17% | 0.58 [0.50, 0.66] |
| vanderLeeuw | 2015 | CVD-EDIC | 2015 | United States | EPIC-Potsdam | Germany | Medium | 15.17% | 0.68 [0.60, 0.76] |
| vanderLeeuw | 2015 | CVD-EDIC | 2015 | United States | SMART | Netherlands | Medium | 10.69% | 0.69 [0.59, 0.79] |
| Read | 2018 | CVD-EDIC | 2015 | United States | National Scottish Diabetes Register | Scotland | Medium | 58.96% | 0.67 [0.67, 0.67] |
| **Overall** | | | | | | | | 100% | 0.66 [0.62, 0.70] |

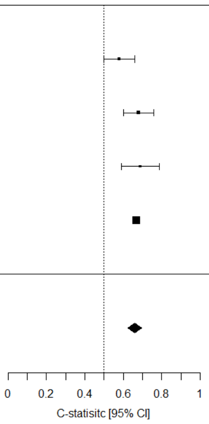

**Random-effects REML model**

$I^2$ = 37.46%; $\tau^2$ = 0.001; $P_Q$ = 0.167; C-statistic [95% CI] = 0.66[ 0.62 - 0.7], $P$ = 3.61e-281

**Fixed-effects model**

$I^2$ = 40.8% ; $P_Q$ = 0.167; C-statistic [95% CI] = 0.67[ 0.67 - 0.67], $P$ = 0

#### Supplemental Figure 12C. UKPDS

**Meta-analysis for the c-statistics of 'UKPDS' risk scores (external validation cohorts)**

| First author | Year | Risk score | Year original risk score published | Derivation country(ies) | Validation cohort | Country(ies) of validation cohort | Risk of bias | Weight (%) | C-statistic [95% CI] |
|---|---|---|---|---|---|---|---|---|---|
| Simmons | 2009 | UKPDS | 1998 | United Kingdom | Epic Norfolk | United Kingdom | Medium | 15.61% | 0.72 [0.66, 0.78] |
| Davis | 2009 | UKPDS | 1998 | United Kingdom | FDS | Australia | Medium | 10.21% | 0.68 [0.60, 0.76] |
| Kengne | 2010 | UKPDS | 1998 | United Kingdom | ADVANCE | Europe/Asia | Low | 29.88% | 0.68 [0.65, 0.72] |
| vanDieren | 2011 | UKPDS | 1998 | United Kingdom | Epic-NL; Epic-Potsdam | Netherlands/Germany | Low | 3.69% | 0.66 [0.51, 0.81] |
| Tanaka | 2013 | UKPDS | 1998 | United Kingdom | JDCS/J-EDIT | Japan | Medium | 14.26% | 0.69 [0.63, 0.76] |
| Piniés | 2014 | UKPDS | 1998 | United Kingdom | BASCORE | Spain | Low | 19.81% | 0.62 [0.57, 0.67] |
| Venäläinen | 2020 | UKPDS | 1998 | United Kingdom | Hoorn Study | Netherlands | Medium | 6.53% | 0.66 [0.55, 0.77] |
| **Overall** | | | | | | | | 100% | 0.68 [0.65, 0.71] |

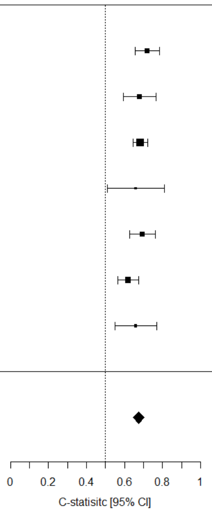

**Random-effects REML model**

$I^2$ = 23.84%; $\tau^2$ = 0; $P_Q$ = 0.383; C-statistic [95% CI] = 0.68[ 0.65 - 0.71], $P$ = 0

**Fixed-effects model**

$I^2$ = 5.84% ; $P_Q$ = 0.383; C-statistic [95% CI] = 0.68[ 0.65 - 0.7], $P$ = 0

**Supplemental Figure 13.** Quality assessment of included studies using modified Newcastle-Ottawa Scale

Supplemental Figure 13A. Biomarker Studies

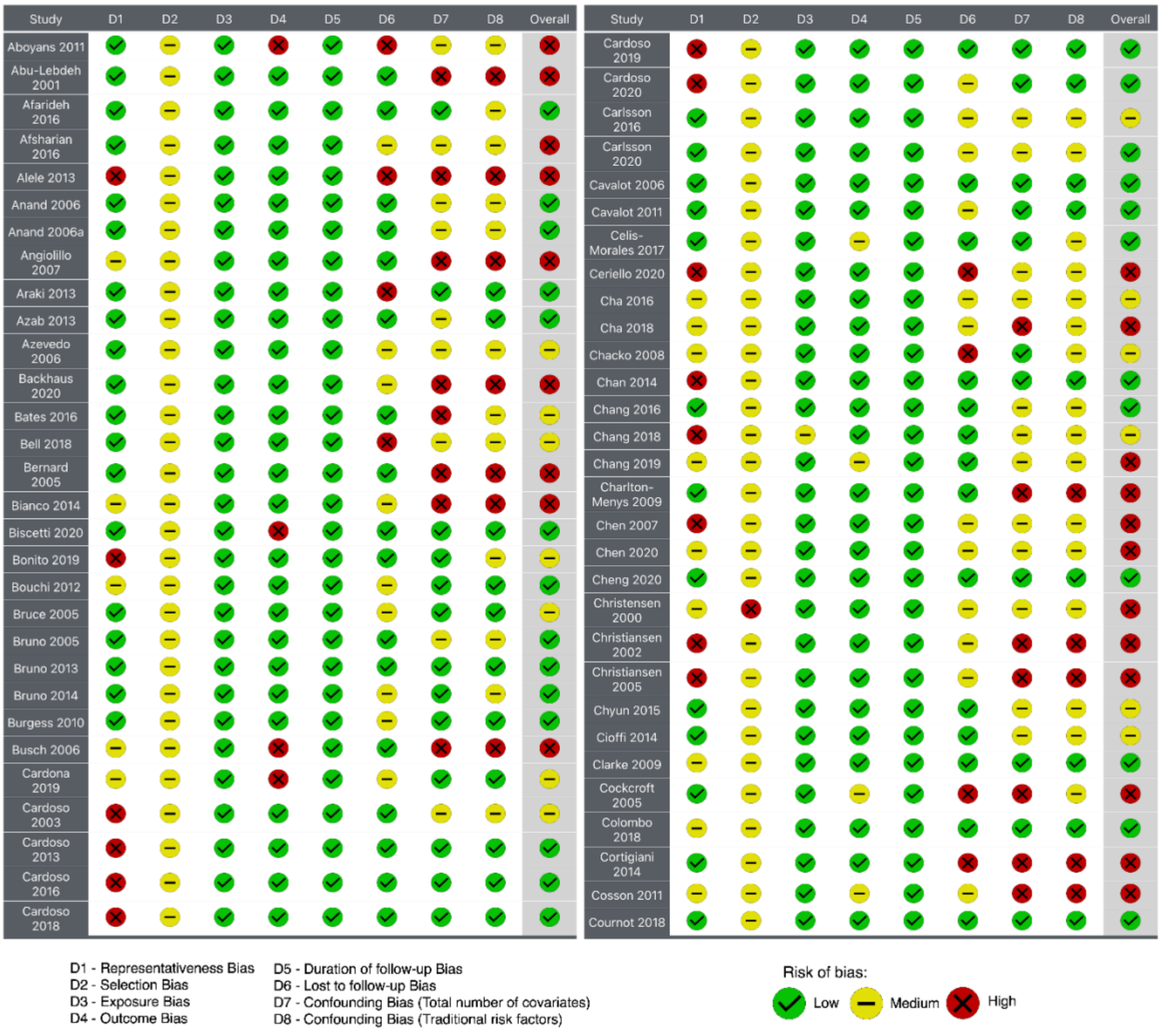

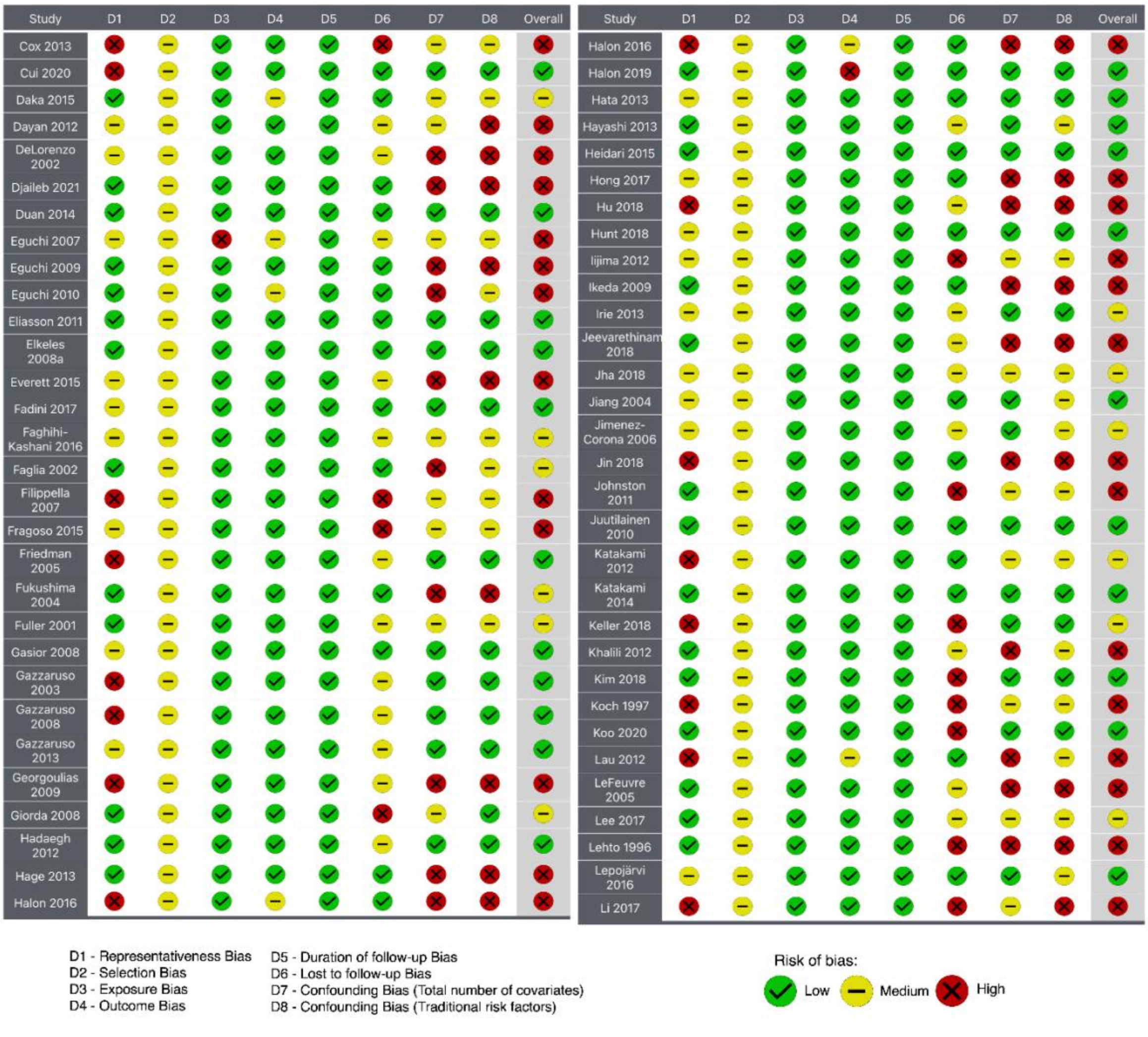
Study
D1
D2
D3
D4
D5
D6
D7
D8
Overall
Cox 2013
Cui 2020
Daka 2015
Dayan 2012
DeLorenzo 2002
Djaileb 2021
Duan 2014
Eguchi 2007
Eguchi 2009
Eguchi 2010
Eliasson 2011
Elkeles 2008a
Everett 2015
Fadini 2017
Faghihi-Kashani 2016
Faglia 2002
Filippella 2007
Fragoso 2015
Friedman 2005
Fukushima 2004
Fuller 2001
Gasior 2008
Gazzaruso 2003
Gazzaruso 2008
Gazzaruso 2013
Georgoulias 2009
Giorda 2008
Hadaegh 2012
Hage 2013
Halon 2016
Halon 2016
Halon 2019
Hata 2013
Hayashi 2013
Heidari 2015
Hong 2017
Hu 2018
Hunt 2018
Iijima 2012
Ikeda 2009
Irie 2013
Jeevarethinam 2018
Jha 2018
Jiang 2004
Jimenez-Corona 2006
Jin 2018
Johnston 2011
Juutilainen 2010
Katakami 2012
Katakami 2014
Keller 2018
Khalili 2012
Kim 2018
Koch 1997
Koo 2020
Lau 2012
LeFeuvre 2005
Lee 2017
Lehto 1996
Lepojärvi 2016
Li 2017
D1 - Representativeness Bias
D2 - Selection Bias
D3 - Exposure Bias
D4 - Outcome Bias
D5 - Duration of follow-up Bias
D6 - Lost to follow-up Bias
D7 - Confounding Bias (Total number of covariates)
D8 - Confounding Bias (Traditional risk factors)
Risk of bias:
Low
Medium
High

| Study | D1 | D2 | D3 | D4 | D5 | D6 | D7 | D8 | Overall |
|---|---|---|---|---|---|---|---|---|---|
| Liao 2019 | Low | Medium | Low | Low | Low | High | Medium | Low | Low |
| Lievre 2011 | Low | Medium | Low | Low | Low | Medium | High | High | High |
| Lim 2008 | High | Medium | Low | Medium | Low | High | High | Medium | High |
| Lim 2019 | Low | Medium | Low | Low | Low | High | Low | Low | Low |
| Lin 2010 | Low | Medium | Low | Low | Low | Medium | Medium | Medium | Medium |
| Lin 2013 | High | Medium | Low | Low | Low | Low | Low | Low | Low |
| Lin 2019 | Low | Medium | Low | Low | Low | Low | Medium | Medium | Medium |
| Linnemann 2003 | Low | Medium | Low | Low | Low | Medium | Medium | Medium | Medium |
| Linnemann 2006 | Medium | Medium | Low | Low | Low | Low | Medium | Medium | Medium |
| Lopes-Virella 2012 | Medium | Medium | Low | Low | Low | Medium | High | High | High |
| Lorenzo-Almorós 2020 | High | Medium | Low | Low | Low | Low | Low | Low | Low |
| Lowe 2014 | Low | Medium | Low | Low | Low | Low | Low | Low | Low |
| Lutgers 2009 | Low | Medium | Low | Low | Low | Medium | Medium | Medium | Medium |
| Masi 2016 | Medium | Medium | Low | Medium | Low | Low | High | High | High |
| Massardo 2020 | High | Medium | Low | Medium | Low | Low | Medium | Medium | High |
| McMurray 2011 | Medium | Medium | Low | Low | Low | Medium | Low | Medium | Low |
| Meerwaldt 2007 | High | Medium | Low | Low | Low | Low | High | Medium | High |
| Mellbin 2010 | High | Medium | Low | Low | Low | Low | High | High | High |
| Monseu 2015 | Low | Medium | Low | Low | Low | Medium | High | High | High |
| Moosaie 2020 | Medium | Medium | Low | Low | Low | Medium | Low | Low | Low |
| Nagamachi 2006 | Low | Medium | Low | Low | Low | Low | Medium | Medium | Medium |
| Nakamura 2010 | Low | Medium | Low | Low | Low | High | Low | Low | Low |
| Nam 2020 | Low | Medium | Low | Low | Low | Low | Low | Medium | Low |
| Nargesi 2016 | Medium | Medium | Low | Low | Low | Low | High | High | High |
| Ndrepepa 2013 | High | Medium | Low | Low | Low | Low | Low | Low | Low |
| Nelson 1990 | High | Medium | Low | Low | Low | High | High | High | High |
| Niskanen 1998 | Low | Medium | Low | Low | Low | High | Medium | Medium | High |
| Nitenberg 2005 | Low | Medium | Low | Low | Low | Medium | High | High | High |
| Novo-Rodríguez 2018 | Medium | Medium | Low | Low | Low | Low | Medium | Medium | Medium |
| Oellgaard 2018 | Medium | Medium | Low | Low | Low | Low | Medium | Medium | Medium |
| Oliveira 2009 | Low | Medium | Low | Low | Low | High | High | High | High |
| Ong 2015 | Medium | Medium | Low | Low | Low | Low | Low | Low | Low |
| Ong 2020 | Low | Medium | Low | Low | Low | High | Low | Medium | High |
| Otto 2012 | Medium | Medium | Low | Low | Low | Low | High | High | High |
| Panero 2012 | Low | Medium | Low | Low | Low | Low | Low | Low | Low |
| Park 2014 | High | Medium | Low | Low | Low | Medium | High | High | High |
| Peng 2009 | Low | Medium | Low | Low | Low | Medium | Low | Low | Low |
| Peters 2013 | Low | Medium | Low | Low | Low | Low | Medium | High | High |
| Petretta 2013 | Low | Medium | Low | Low | Low | Low | Low | Low | Low |
| Pfister 2011 | High | Medium | Low | Low | Low | Medium | Low | Low | Low |
| Pickup 2003 | High | Medium | Low | Low | Low | Low | High | High | High |
| Pintó 2007 | Low | Medium | Low | Low | Low | Medium | High | High | High |
| Prentice 2016 | Low | Medium | Low | Low | Low | Low | Low | Medium | Low |
| Qin 2020 | Low | Medium | Low | Low | Low | High | Low | Low | Low |
| Radholm 2017 | Low | Medium | Low | Low | Low | Medium | Medium | Medium | Medium |
| Rana 2005 | Low | Medium | Low | Low | Low | Low | High | High | High |
| Rasmussen 2018 | Low | Medium | Low | Low | Low | High | Medium | Low | Medium |
| Ravassa 2015 | Medium | Medium | Low | High | Low | Low | Medium | Medium | High |
| Rawshani 2016 | Low | Medium | Low | Low | Low | High | Low | Low | Low |
| Reinhard 2010 | Medium | Medium | Low | Low | Low | Medium | Medium | Medium | Medium |
| Resl 2012 | Medium | Medium | Low | Low | Low | Low | Medium | Medium | Medium |
| Resl 2016 | Low | Medium | Low | Low | Low | Low | Low | Low | Low |
| RotbainCurovi 2018 | Low | Medium | Low | Low | Low | Low | Medium | Medium | Low |
| Roumeliotis 2019 | Low | Medium | Low | Low | Low | Low | Low | Low | Low |
| Rozing 2019 | Low | Medium | Low | Low | Low | Low | Low | Low | Low |
| Rutter 2002 | Low | Medium | Low | Low | Low | Low | Low | Low | Low |
| Rørth 2019 | Low | Medium | Low | Low | Low | Low | Low | Medium | Low |
| Saely 2005 | High | Medium | Low | Low | Low | Medium | High | Medium | High |
| Saely 2006 | High | Medium | Low | Low | Low | Medium | Low | Low | Low |
| Saito 2000 | Low | Medium | Low | Low | Low | High | Low | Low | Medium |

D1 - Representativeness Bias
D2 - Selection Bias
D3 - Exposure Bias
D4 - Outcome Bias
D5 - Duration of follow-up Bias
D6 - Lost to follow-up Bias
D7 - Confounding Bias (Total number of covariates)
D8 - Confounding Bias (Traditional risk factors)

Risk of bias: Low, Medium, High

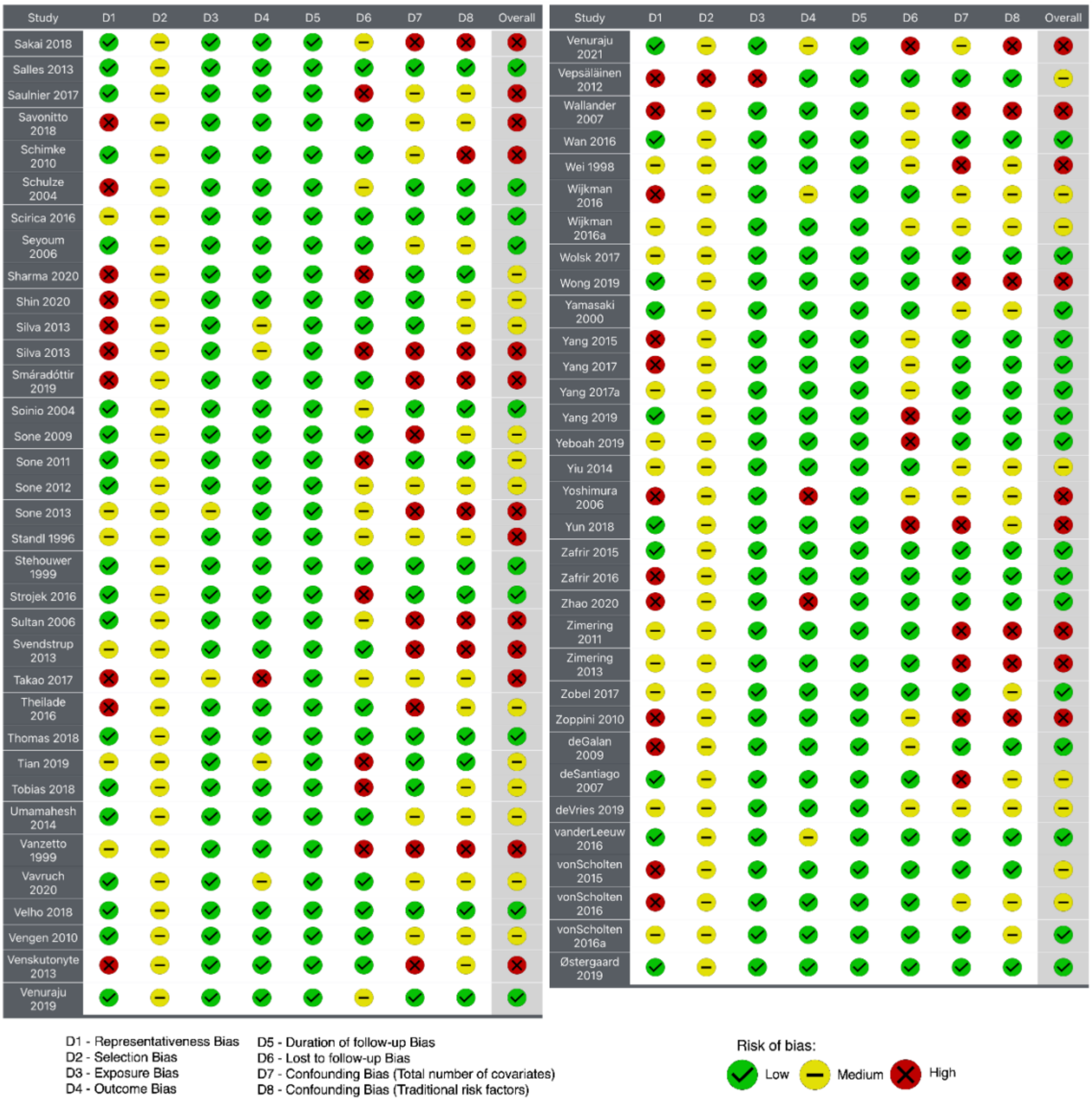
Study
D1
D2
D3
D4
D5
D6
D7
D8
Overall
Sakai 2018
Salles 2013
Saulnier 2017
Savonitto 2018
Schimke 2010
Schulze 2004
Scirica 2016
Seyoum 2006
Sharma 2020
Shin 2020
Silva 2013
Silva 2013
Smáradóttir 2019
Soinio 2004
Sone 2009
Sone 2011
Sone 2012
Sone 2013
Standl 1996
Stehouwer 1999
Strojek 2016
Sultan 2006
Svendstrup 2013
Takao 2017
Theilade 2016
Thomas 2018
Tian 2019
Tobias 2018
Umamahesh 2014
Vanzetto 1999
Vavruch 2020
Velho 2018
Vengen 2010
Venskutonyte 2013
Venuraju 2019
Venuraju 2021
Vepsäläinen 2012
Wallander 2007
Wan 2016
Wei 1998
Wijkman 2016
Wijkman 2016a
Wolsk 2017
Wong 2019
Yamasaki 2000
Yang 2015
Yang 2017
Yang 2017a
Yang 2019
Yeboah 2019
Yiu 2014
Yoshimura 2006
Yun 2018
Zafrir 2015
Zafrir 2016
Zhao 2020
Zimering 2011
Zimering 2013
Zobel 2017
Zoppini 2010
deGalan 2009
deSantiago 2007
deVries 2019
vanderLeeuw 2016
vonScholten 2015
vonScholten 2016
vonScholten 2016a
Østergaard 2019
D1 - Representativeness Bias
D2 - Selection Bias
D3 - Exposure Bias
D4 - Outcome Bias
D5 - Duration of follow-up Bias
D6 - Lost to follow-up Bias
D7 - Confounding Bias (Total number of covariates)
D8 - Confounding Bias (Traditional risk factors)
Risk of bias:
Low
Medium
High

#### Supplemental Figure 13B. Genetics Studies

| Study | D1 | D2 | D3 | D4 | D5 | D6 | Overall |
|---|---|---|---|---|---|---|---|
| Alkhalaf 2015 | High | Medium | Low | Low | Low | Medium | High |
| Bacci 2011 | High | Medium | Low | Medium | Low | Medium | High |
| Baeney 2016 | Low | Medium | Low | Low | Low | Medium | Medium |
| Bernard 2004 | High | Medium | Low | Low | Low | Low | Medium |
| Boger 2005 | High | Medium | Low | High | Low | Medium | High |
| Cox 2014 | High | Medium | Low | Low | Low | Low | Medium |
| Doney 2005 | Low | Medium | Low | Medium | Low | Low | Medium |
| Doney 2005a | Low | Medium | Low | Medium | Low | Low | Medium |
| Doney 2009 | Low | Medium | Low | High | Low | Medium | High |
| Ferrarezi 2013 | Low | Medium | Low | Low | Low | Low | Low |
| Hadjadj 2008 | High | Medium | Low | Low | Low | Low | Medium |
| He 2021 | Low | Medium | Low | Low | Low | Medium | Low |
| Heijmans 2000 | High | Medium | Low | Low | Low | Medium | High |
| Ho 2012 | High | Medium | Low | Low | Low | Low | Medium |
| Hoffman 2011 | Medium | Medium | Low | Low | Low | Low | Medium |
| Hong Huang 1998 | Low | Medium | Low | Low | Low | Low | Low |
| Huggins 2016 | Medium | Medium | Low | Low | Low | Low | Medium |
| Katakami 2014 | Low | Medium | Low | Low | Low | Low | Low |
| Keavney 1995 | Low | Medium | Low | Low | Low | Medium | Medium |
| Kuricová 2013 | Low | Medium | Low | High | Low | Low | High |
| Levy 2002 | Low | Medium | Low | Low | Low | Low | Low |
| Lu 2011 | High | Medium | Low | Low | Low | Low | Low |
| Lu Qi 2005 | High | Medium | Low | Low | Low | Low | Medium |
| Mccaffery 2015 | High | Medium | Low | Low | Low | Medium | High |
| Mohammedi 2015 | Medium | Medium | Low | Low | Low | Low | Medium |
| Morieri 2018 | Low | Medium | Low | Low | Low | Medium | Medium |
| Neves 2012 | Low | Medium | Low | Low | Low | Low | Low |
| Odeberg 2008 | Low | Medium | Low | Medium | Low | Low | Medium |
| Ortega Moreno 2016 | Medium | Medium | Low | Low | Low | Medium | Medium |
| Poon 2010 | High | Medium | Low | Low | Low | Low | Medium |
| Poon 2014 | High | Medium | Low | Medium | Low | Low | High |
| Porchay-Baldérelli 2007 | Medium | Medium | Low | Low | Low | Low | Medium |
| Porchay-Baldérelli 2009 | High | Medium | Low | Low | Low | Low | Medium |
| Qi 2011 | Low | Medium | Low | Low | Medium | High | High |
| Qi 2012 | Low | Medium | Low | Low | Low | Low | Low |
| Qi 2013 | Low | Medium | Low | Low | Low | Low | Low |
| Roumeliotis 2017 | High | Medium | Low | Low | Low | Medium | High |
| Roumeliotis 2018 | High | Medium | Low | Low | Low | Low | Medium |
| Russo 2011 | Low | Medium | Low | Low | Low | Medium | Medium |
| Satirapoj 2019 | High | Medium | Low | Low | Low | Low | Medium |
| So 2008 | Medium | Medium | Low | High | Low | Medium | High |
| Tan 2020 | Low | Medium | Low | Low | Low | Medium | Medium |
| Valoti 2019 | Low | Medium | Low | Low | Low | Low | Low |
| Wang 2005 | Low | Medium | Medium | High | Low | High | High |
| Wang 2010 | High | Medium | Low | Low | Low | Medium | High |
| Watson C 2021 | Low | Medium | Low | Low | Low | Low | Low |
| Winkler 2010 | Low | Medium | Low | Low | Low | Low | Low |
| Zhang 2005 | High | Medium | Low | Low | Low | Low | Medium |

D1 - Representativeness Bias
D2 - Selection Bias
D3 - Exposure Bias
D4 - Outcome Bias
D5 - Duration of follow-up Bias
D6 - Lost to follow-up Bias

Risk of bias: Low, Medium, High

Supplemental Figure 13C.  Risk Score Studies

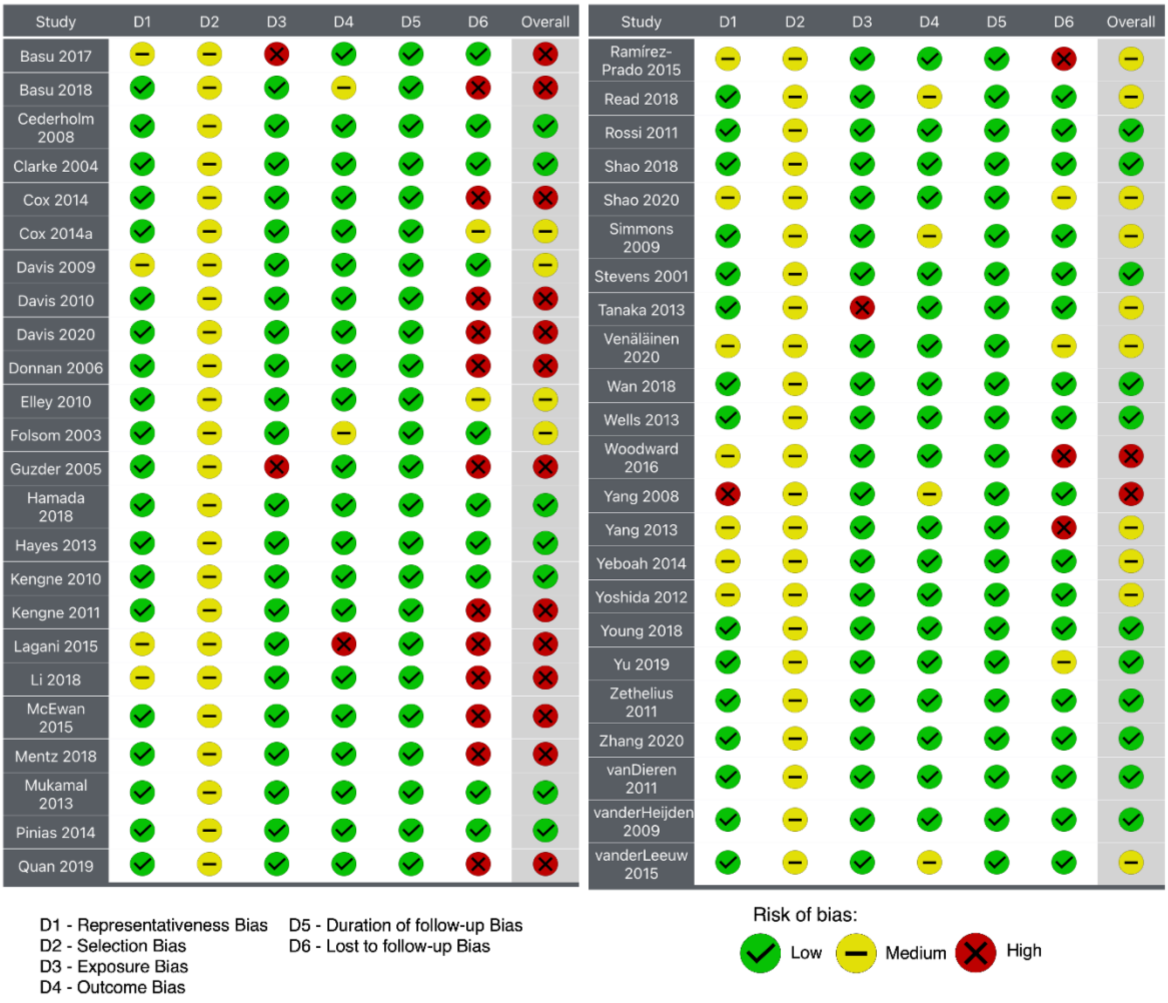

**Supplemental Figure 13** Caption:   Quality assessment (risk of bias traffic light plot) for (A) biomarkers, (B) genetic markers, and (C) risk scores.
